# Rational Empiric Antibiotic Escalation Applied to Specific Patient Groups

**DOI:** 10.1101/2023.11.03.23298025

**Authors:** Ranjeet Bamber, Brian Sullivan, Léo Gorman, Winnie WY Lee, Matthew B Avison, Andrew W Dowsey, Philip Williams

**Affiliations:** Department of Population Health Sciences, Bristol Medical School, Faculty of Health Sciences, University of Bristol, Bristol. UK; Jean Golding Institute, University of Bristol, Bristol. UK; School of Cellular and Molecular Medicine, Faculty of Life Sciences, University of Bristol, Bristol. UK; Department of Population Health Sciences, Bristol Medical School, Faculty of Health Sciences, University of Bristol; University Hospitals Bristol and Weston NHS Foundation Trust, Bristol Royal Infirmary, Bristol. UK

## Abstract

**Background:** Clinicians commonly escalate empiric antibiotic therapy due to poor clinical progress, without microbiology guidance. When escalating, they should take account of how resistance to an initial antibiotic affects the probability of resistance to subsequent options. The term Escalation Antibiogram (EA) has been coined to describe this concept. One difficulty when applying the EA concept to clinical practice is understanding the uncertainty in results and how this changes for specific patient subgroups.

**Methods:** A Bayesian model was developed to estimate antibiotic resistance rates in Gram-negative bloodstream infections based on phenotypic resistance data. It provides an expected value (posterior mean) with 95% credible interval to illustrate uncertainty, based on the size of the patient subgroup, and estimates probability of inferiority between two antibiotics. This model can be applied to specific patient groups where resistance rates and underlying microbiology may differ from the whole hospital population.

**Results:** Rates of resistance to empiric first choice and potential escalation antibiotics were calculated for the whole hospitalised population based on 10,486 individual bloodstream infections, and for a range of specific patient groups, including ICU, haematology-oncology, and paediatric patients. Differences in optimal escalation antibiotic options between specific patient groups were noted.

**Conclusions:** EA analysis informed by our Bayesian model is a useful tool to support empiric antibiotic switches, providing an estimate of local resistances rates, and a comparison of antibiotic options with a measure of the uncertainty in the data. We demonstrate that EAs calculated for the whole population cannot be assumed to apply to specific patient groups.

## Key Summary Points

Clinicians commonly escalate empiric antibiotic therapy due to poor clinical progress, without microbiology guidance.
When escalating, they should take account of how resistance to an initial antibiotic affects the probability of resistance to subsequent options.
We describe a Bayesian model to guide empiric antibiotic escalation, based on local resistance data, that can be applied to small patient groups to predict rates of resistance with credible intervals.
We demonstrate that the optimal antibiotic escalation for the whole population cannot be assumed to apply to specific patient groups, such as ICU or haematology patients.

## Introduction

In many countries, initial empiric antibiotic (AB) choices for the management of bacterial infections are informed by local guidelines that in turn rely on an understanding of local phenotypic AB resistance (ABR) patterns from blood culture isolates and other sample types. This initial AB selection usually takes place within an hour of diagnosis of infection, and well before any microbiological results are available. Furthermore, microbiological culture is a relatively insensitive technique. Even in patients with sepsis, only 30–50% will have a positive blood culture (1, 2). Hence, it is also common for clinicians to empirically escalate AB therapy due to poor clinical progress in the absence of positive microbiology as a guide. While a poor clinical response may be due to a variety of factors, for example inadequate source control, it is the possibility of resistance to the first-choice treatment that usually drives escalated AB therapy. In this context, therefore, it is not the local rate of resistance to the possible second-choice AB options that should be considered, but the rate of co-resistance to these agents in isolates resistant to the first AB choice. Though widely available, local phenotypic resistance data are an underused resource, and could be used to gain a better understanding of circulating co-resistance patterns and improve initial and escalated empiric AB choice.

Some prior studies have explored this approach. As part of a medical decision support system tool Zalounina *et al* (3) included prior AB treatments and local co-resistance data to guide subsequent AB choice. Wong *et al* (4) investigated 3,280 Gram-negative bacillus (GNB) blood stream infections (BSI) looking at the correlation coefficients between pairs of ABs, and the ABR profiles of subsets of isolates resistant to a specified AB. They discussed examples of how these data could be used to guide empiric AB escalation. They also suggested that local, unit based, ABR profiles and co-resistance patterns may be more useful than nationally collected data. Recently, Teitelbaum *et al* (5) took a similar approach looking at GNB BSIs. They coined the term “escalation antibiogram (EA)” to describe a profile of resistance to a set of ABs given resistance to an initial set of 12 ABs. Their local resistance patterns were stable over the period studied, including 6577 GNB BSI episodes, from 6 hospitals in their area allowing the data to be combined and averaged over time. They noted local EAs were easy to generate, however they did not have the data available to subgroup by presumed BSI source, and noted that the data may not generalise to specific patient groups.

One difficulty faced when applying the local EA concept to specific patient groups is the small numbers involved. Moreover, rates of ABR may vary over time and can vary significantly from country to country (6) and between regions within a country (7,8). Hence when attempting to produce an EA applicable to certain patient groups, taking an average over several years will not always be appropriate, nor is it possible to get extra power by combining data from other regions without diluting the desired effects of providing an EA based on local resistance data. Hence, our objective was to develop and validate a model that allows tracking of variation in local ABR and co-resistance over time, and so maximises our ability to define an EA for specific patient groups. Our focus here was on ICU patients, haemato-oncology patients, patients with specific sources of BSI, and adults over 80 years. The Bayesian model scripts we developed for local use can be applied to train the model for any region.

## Methods

### Data collection and cleaning

Data were collected from 3 NHS Trusts covering 4 hospitals in the Southwest of England serving a population of approximately 1.5 million people (Royal United Hospital Bath NHS Foundation Trust, University Hospitals Bristol & Weston NHS Foundation Trust, and North Bristol NHS Trust) which share a single laboratory information management system (Winpath Enterprise 7.23, Clinisys). Positive blood cultures from all 4 hospitals where GNBs were isolated over a 6-year period, from 2017 to 2022, were included.

For each isolate, the ABR profile was determined by the European Committee on Antimicrobial Susceptibility Testing (EUCAST) disc testing or by the Biomerieux Vitek 2 automated system. Direct disc sensitivities were obtained using EUCAST criteria following the Gram stain. In most cases (85%) isolates from a purity plate were tested using the Vitek 2 system the following day giving ABR profiles for a wider range of ABs. Vitek results, when available, were used in preference to the initial disc testing. Results were expressed as “Sensitive”, “Intermediate”, or “Resistant”. Following usual clinical practices “Intermediate” and “Resistant” isolates were grouped together as non-susceptible, which from now on we will refer to as “Resistant”. Healthcare systems that have adopted EUCAST V14 break points may elect to combine “Sensitive” and “Increase dose” as one group. When using a time series of categorical data, changes to the definition of a category e.g. a change in break points should be delt with by re-categorising the whole data set for affected organisms using MIC data and new break points. Producing an R script to do this would be a trivial task.

Repeat samples from the same patient with an indistinguishable isolate (Species ID and ABR profile) which occurred within 1 year were removed. Following de-duplication, we had 10,486 GNB BSIs over the 6-year period. For *Pseudomonas* spp. and *Stenotrophomonas* spp. isolates, ABs that are not tested due to assumed intrinsic resistance were automatically set to Resistant.

From 2020 onwards, the presumed source of BSI was recorded in a two-level hierarchical system, with the highest level consisting of the following groups: central nervous system, cardiac, gastro-intestinal, urine/renal, bone/joint, skin/soft tissue, respiratory, reproductive tract, mouth/head and neck, line infection, contaminant, unknown, or other. Data were extracted every 3 months and missing data filled in by case note review.

### Technical details of the Bayesian model

A Bayesian model was developed that estimates ABR rate in GNB BSI and provides an expected value (posterior mean) and 95% credible interval of ABR rate for a given AB at any chosen time point within the 6 years of data collected.

Formally, we use a generalised additive model with a Bernoulli likelihood and logit link function for the binary outcome (Sensitive/Resistant) and a time varying covariate modelled by a penalised thin-plate regression spline. The model generates a series of “credible” curves to fit the resistance data, each with the same probability of representing the true rate given the inherent uncertainty. To avoid overfitting, an integrated penalisation term adaptively smooths the curves given the level of evidence. The model is implemented with the R package ‘brms’ (9), which adopts the Stan platform for Bayesian inference with Hamiltonian Monte-Carlo sampling (10). Four chains are run for 4,000 iterations, with the first 50% discarded as warm-up, leading to 8,000 plausible curves fitting the data.

Statistics such as mean and credible intervals can be directly computed from samples of the curves at the required times.

Resistance rates between two groups or time points are compared by subtracting one set of curves from another, allowing us to calculate the posterior probability of an increase (PPI) or decrease (PPD) in resistance over time or the posterior probability of inferiority (PPInf), or superiority (PPSup) between two AB options. In keeping with the philosophy of a Bayesian approach we do not attempt to define what probability of difference or superiority is significant, but to simply provide that probability of a difference with the 95% credible interval.

## Results & Discussion

### Changes in ABR over time

Over a 6-year period between 2017 to 2022, ABR rates were calculated for deduplicated GNB BSI in patients across our local hospitals (n=10,486). An example of this output, piperacillin/tazobactam resistance in haemato-oncology patients is reported (Fig. 1).

**Figure 1.**
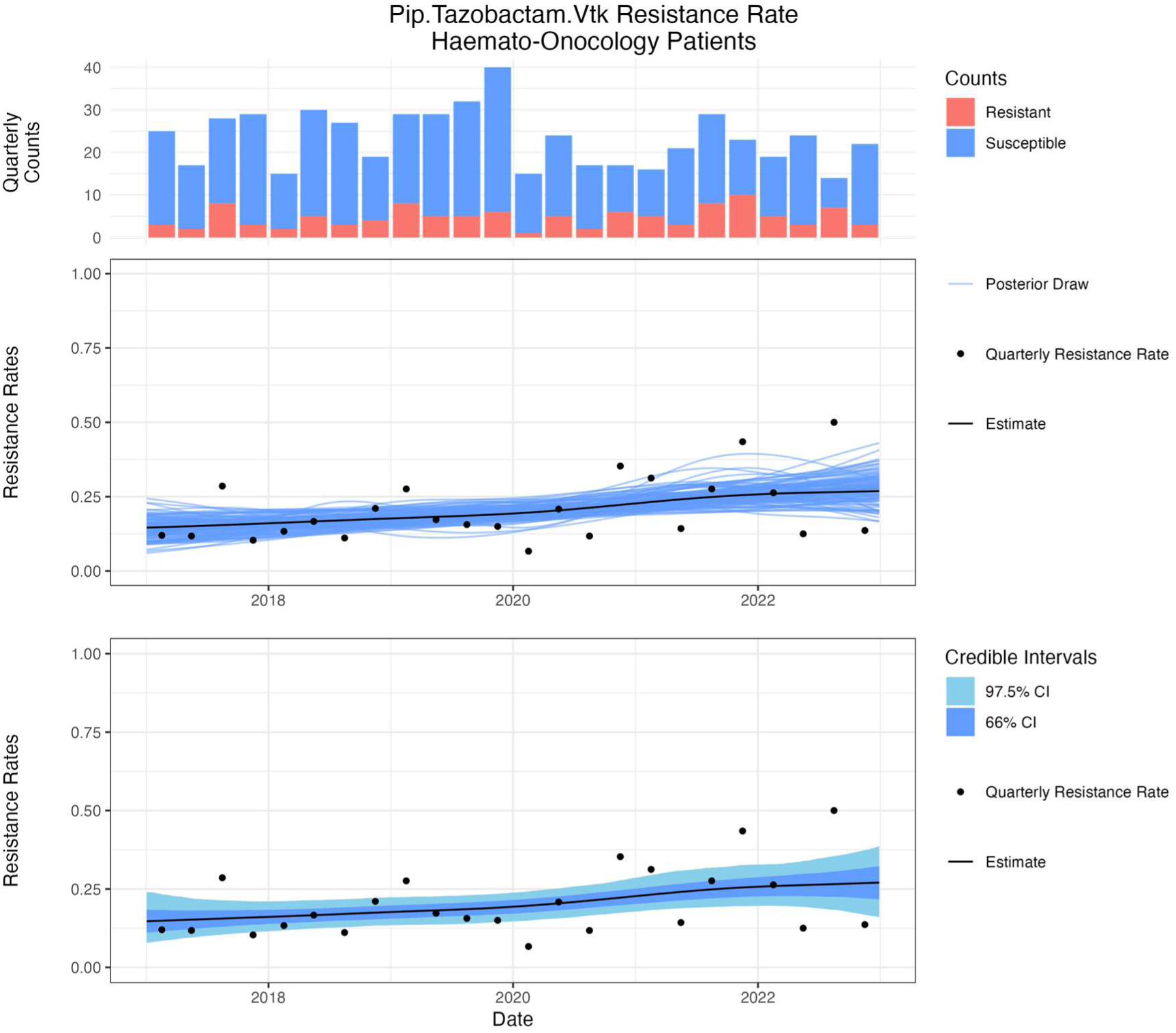
Piperacillin/tazobactam resistance in haemato-oncology patients. (Top) A stacked barplot representing the number of samples in the data. (Middle) A ‘spaghetti plot’ of a random selection of splines fitted to the data that make up the model, and (Bottom) a ‘ribbon’ plot showing the inferred posterior mean (black line) with the 66% and 95% credible interval (dark and light blue shaded areas) with the quarterly rates shown as dots.

The resistance rate increased for meropenem (1.5% to 2.2%, PPI = 88.7%), but reduced slightly (See supplementary material) in all other ABs studied (piperacillin/tazobactam (Tazocin), gentamicin, amikacin, cefotaxime, ceftazidime, ciprofloxacin, trimethoprim/sulfamethoxazole (Cotrimoxazole) and amoxicillin/clavulanate (Coamoxiclav). The reductions ranged from 2.3% in Ceftazidime (Fig. 2a) (PPD = 94.3%), to 6.9% in trimethoprim/sulfamethoxazole resistance rate (Fig. 2b) (PPD = 98.5%). This larger reduction in resistance may be related to the removal of trimethoprim as first-line treatment for lower urinary tract infection in local community guidelines from April 2017, which resulted in a reduction in trimethoprim resistance in *E. coli* from community urine samples (11).

**Figure 2.**
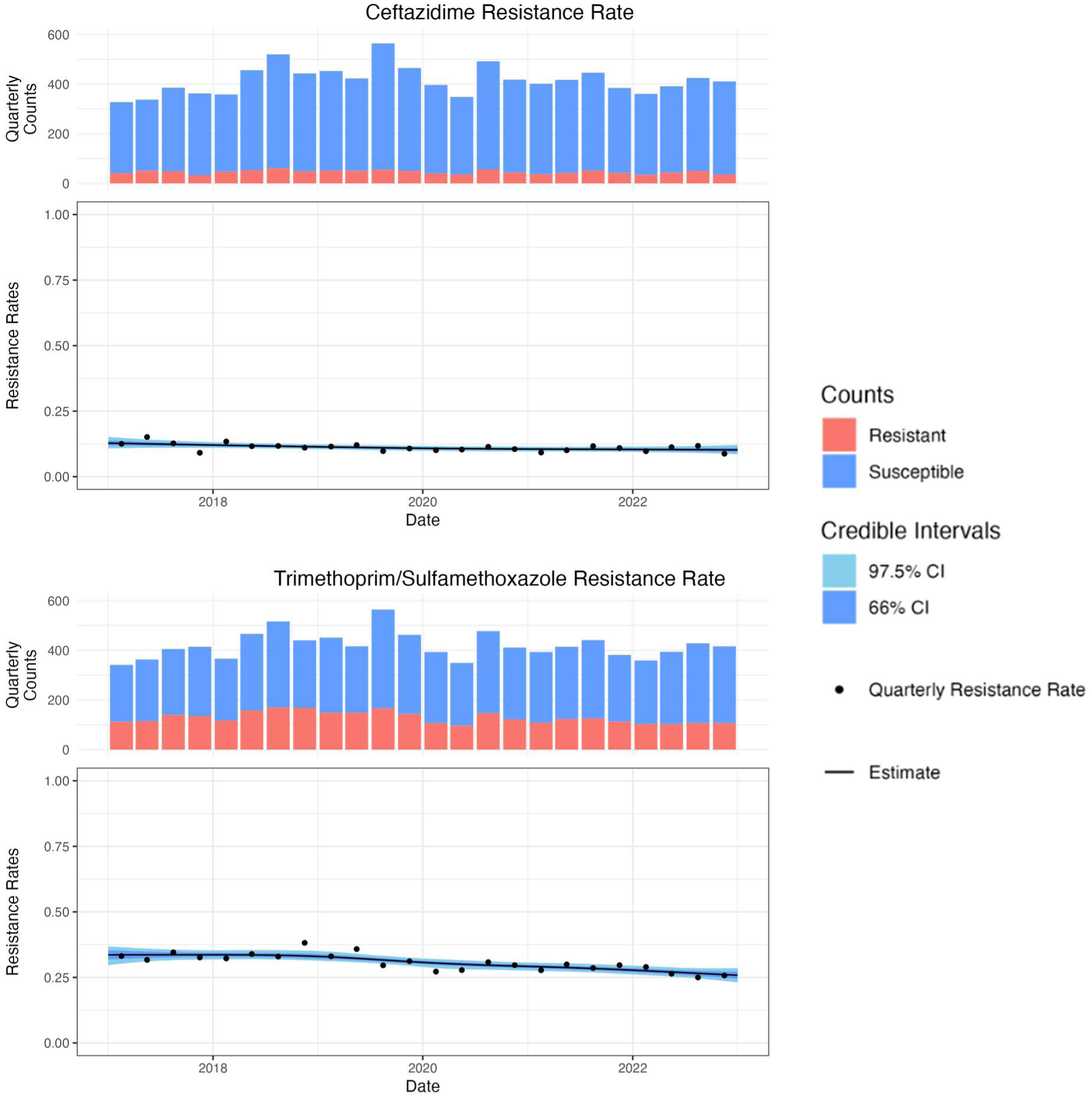
All deduplicated GNB BSI isolates (n=10486) (a) Percent Resistance to Ceftazidime reduces from 12.9% (95% CI 10.9 to 15.3) in 2017 to 10.6% (95% CI 8.8 to 12.9) in 2022. (b) % Resistance to trimethoprim/sulfamethoxazole reduces from 32.9% (95% CI 28.3 to 36.4) in 2017 to 26.0% (95% CI 22.7 to 29.4) in 2022

### Informing empiric AB choice in specific patient groups

When examining resistance to specific ABs in specific patient populations, the 95% CI tends to widen due to the reduced sample size. For example, piperacillin/tazobactam resistance in GNB BSIs in ICU patients has a much wider 95% CI compared to the whole hospital data, as the former is based on 695 positive blood cultures compared to 10,486 for the latter (Fig. 3a, 3b). Over time, piperacillin/tazobactam resistance rates have increased for ICU patients and haemato-oncology patients (fig. 3b, 3c), while reducing over the whole hospital population (fig. 3a). Clearly, therefore, using the mean resistance rate for piperacillin/tazobactam over the 6-year period to inform empiric AB choice would underestimate the current resistance rate in the ICU and haemato-oncology populations, while using only the last year or few months data would be excessively influenced by random month-to-month variation in small patient groups. Our model overcomes this issue as the time series data allows us to estimate current ABR rates, and the uncertainty, while the model borrows information from the earlier data.

**Figure 3.**
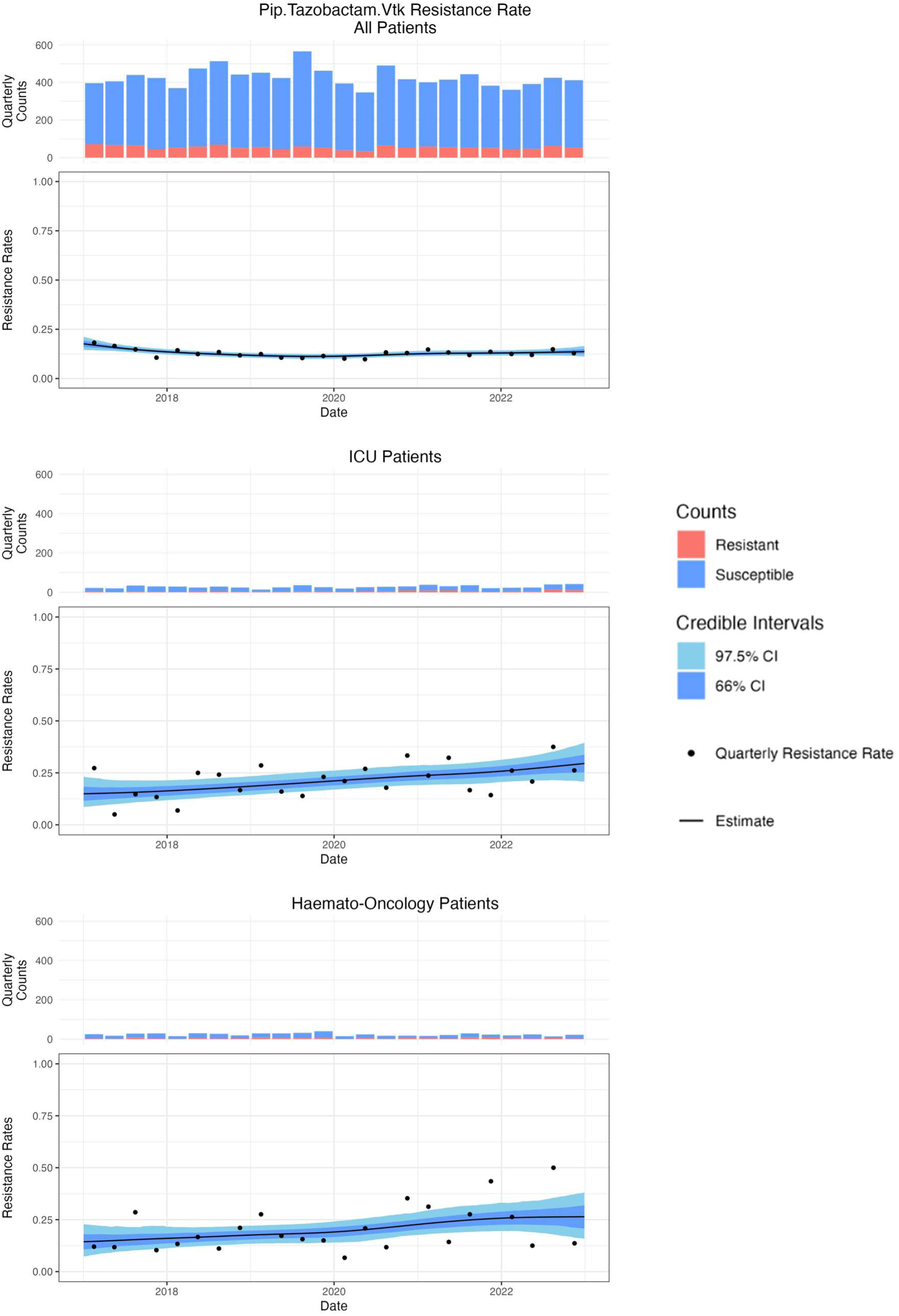
Piperacillin/tazobactam resistance for all patients (Top) compared to ICU patients (Middle) and haemato-oncology patients (bottom)

Following this example, the posterior means between piperacillin/tazobactam resistance rates in GNB BSI isolates from the ICU population and the whole hospital population are different (27.4% vs 13.4%, respectively), and the 95% CIs do not overlap, suggesting this difference is reliable. The probability that ICU patients have higher resistance rates can be computed simply as the proportion of curves that are higher in models of the ICU population than the whole hospital population at this timepoint, in this case, PPI = 99.9%.

In contrast, the difference between the posterior mean piperacillin/tazobactam resistance rates for BSI GNBs from ICU and haemato-oncology patients is small (27.4% vs 30.7%) and there is a considerable overlap in the 95% CIs. This is reflected in the calculated posterior probability of 67.5% that piperacillin/tazobactam resistance rate is lower in the ICU population compared with the haemato-oncology population (e.g., PPD = 67.5%) or conversely there is a 32.5% probability that the resistance rate is greater in ICU patients than in haemato-oncology patients (e.g., PPI = 32.5%).

The situation for ceftazidime is similar, with a higher (and increasing) rate of resistance in both ICU and haemato-oncology patients compared to the whole hospital population. Both ICU and haemato-oncology patients have a higher proportion of potentially AmpC hyper-producing “SPACE” (*Serratia*, *Pseudomonas*, *Acinetobacter*, *Citrobacter* & *Enterobacter*) isolates compared to the whole population (ICU 38% vs 25%, PPD = 99.3%) and haemato-oncology (30% vs 25%, PPD = 87.0%) which explains a proportion of the higher resistance rates.

The increase in ceftazidime resistance over time is likely greater in haemato-oncology patients (15.2% to 24.5%, PPI = 89.4%) than ICU (17.6% TO 20%, PPI = 66.3%), and is due to both increased resistance in non-*E. coli* isolates include in SPACE organisms, (Figure 4) and in an increase in the proportion of non-*E. coli* isolates (*E. coli* reduced from 44% to 35%, PPD = 82%, in haemato-oncology, while in ICU the proportion of *E. coli* remained at 27%)

**Figure 4.**
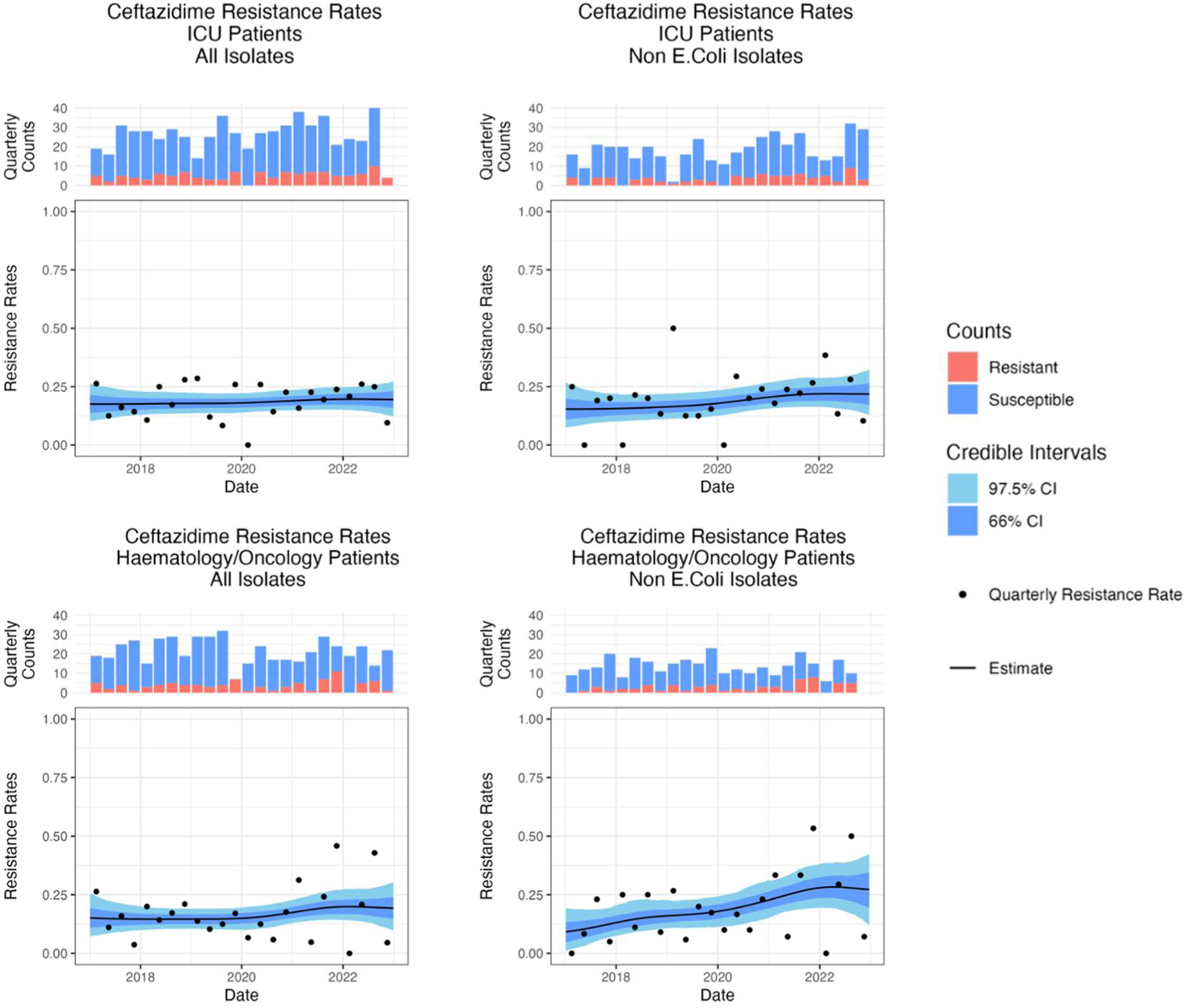
Percent resistance to ceftazidime in all isolates (left column) and non-E coli isolates (right column) located in either (Top Row) ICU (Bottom row) or Haematology/Oncology.

### Calculating an escalation antibiogram (EA) for specific patient groups

To calculate an EA, our Bayesian model is applied for any given second-choice AB to all isolates resistant to the first-choice AB. Table 1 reports these data for the whole hospital population express as a most likely rate of resistance with 95% credible intervals. In this population if moving away from piperacillin/tazobactam we would predict 27% (95% CI 18 to 24) if moving to gentamicin and 31% (95% CI 21 to 40) if switching to ciprofloxacin.

**Table 1.**
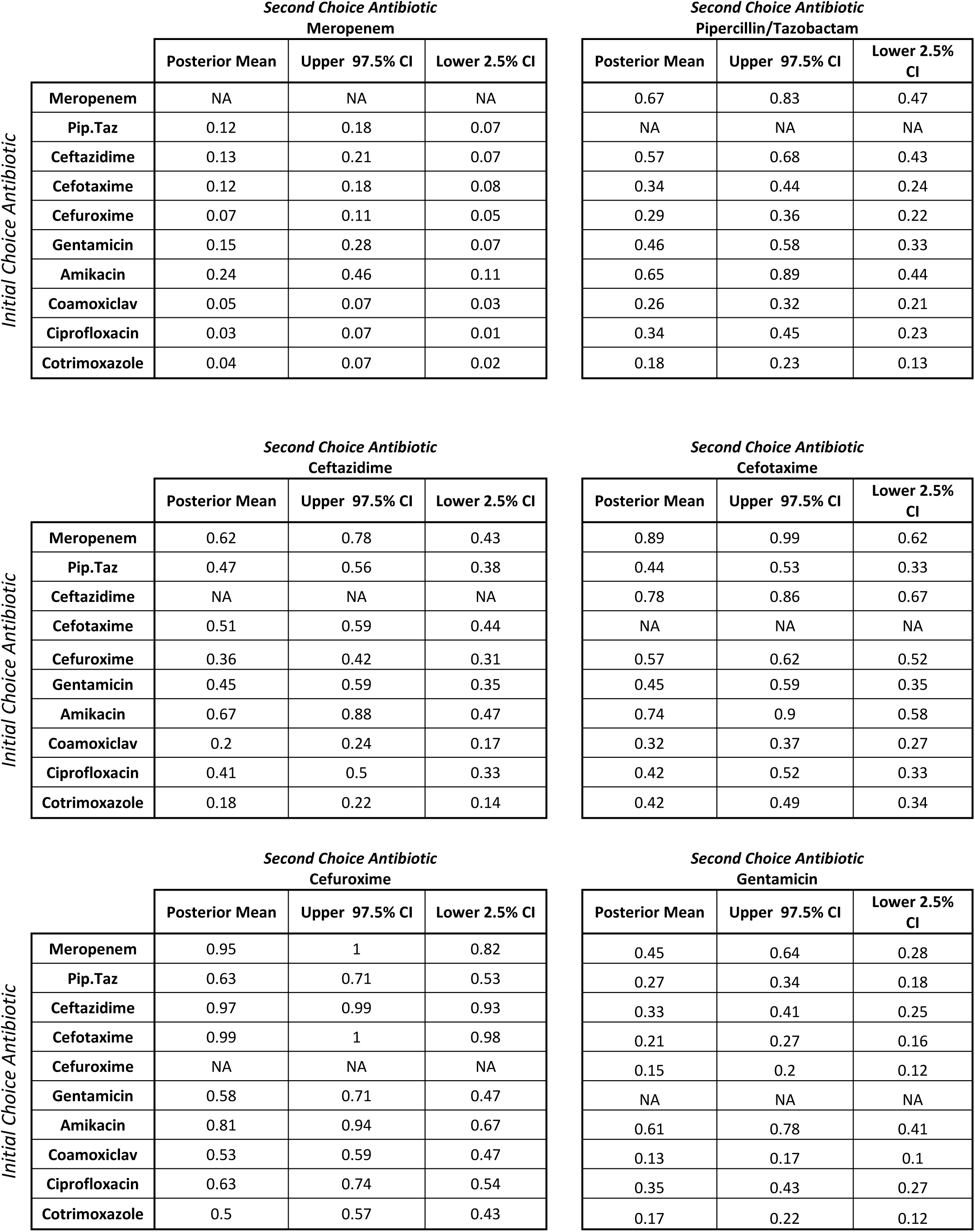
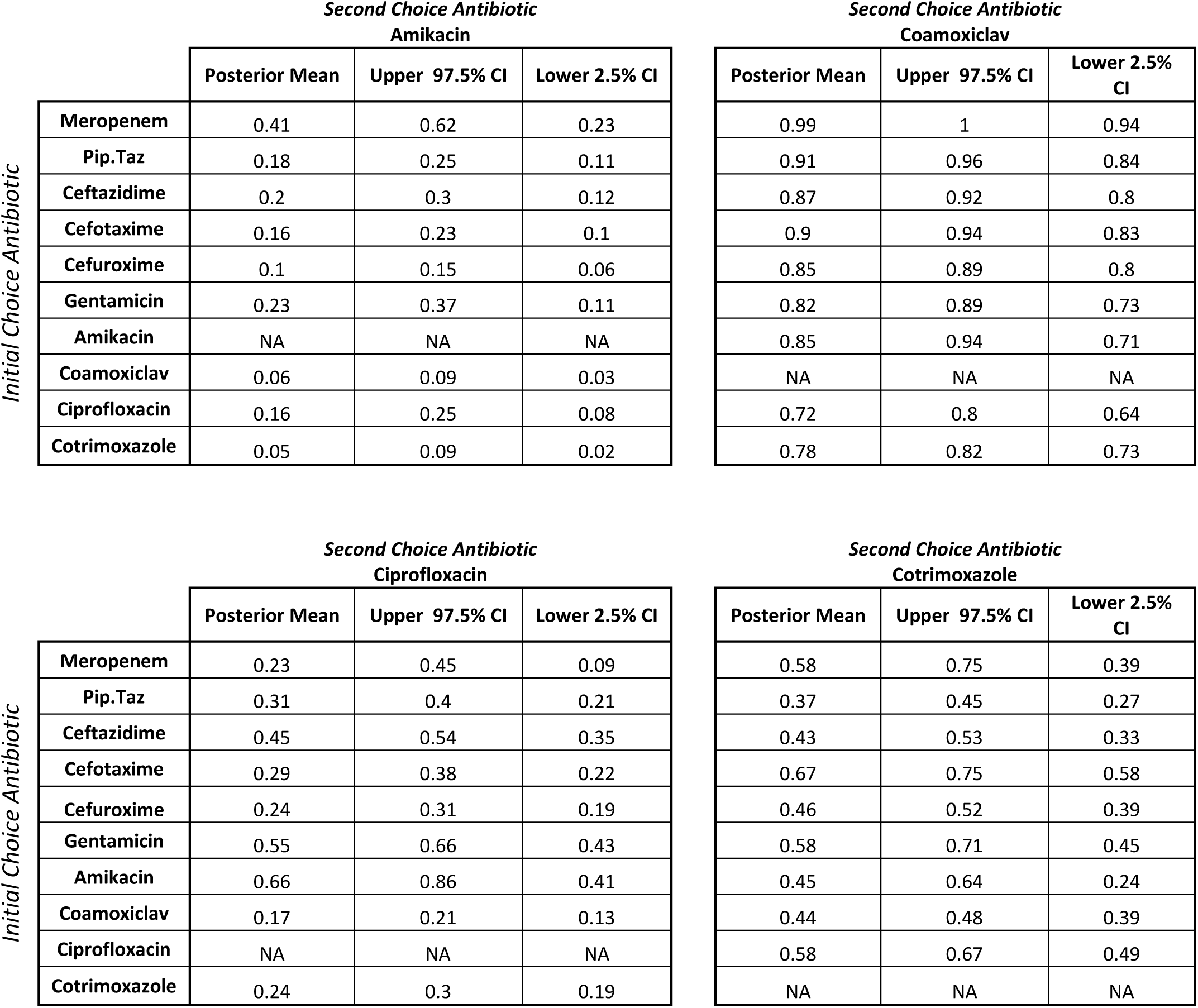
Posterior mean resistance estimates with 95% credible intervals for the whole hospital population. Tables for haemato-oncology, paediatrics, GI or urinary source infections are in the supplementary material.

We can also calculate the probability of inferiority (PPInf) between pairs of alternative antibiotics. This is shown in figure 5 using data for ICU patients assuming piperacillin/tazobactam resistance. When making a clinical decision on antibiotic selection, the probability of inferiority is only useful when both alternatives have a suitably low rate of resistance.

**Figure 5.**
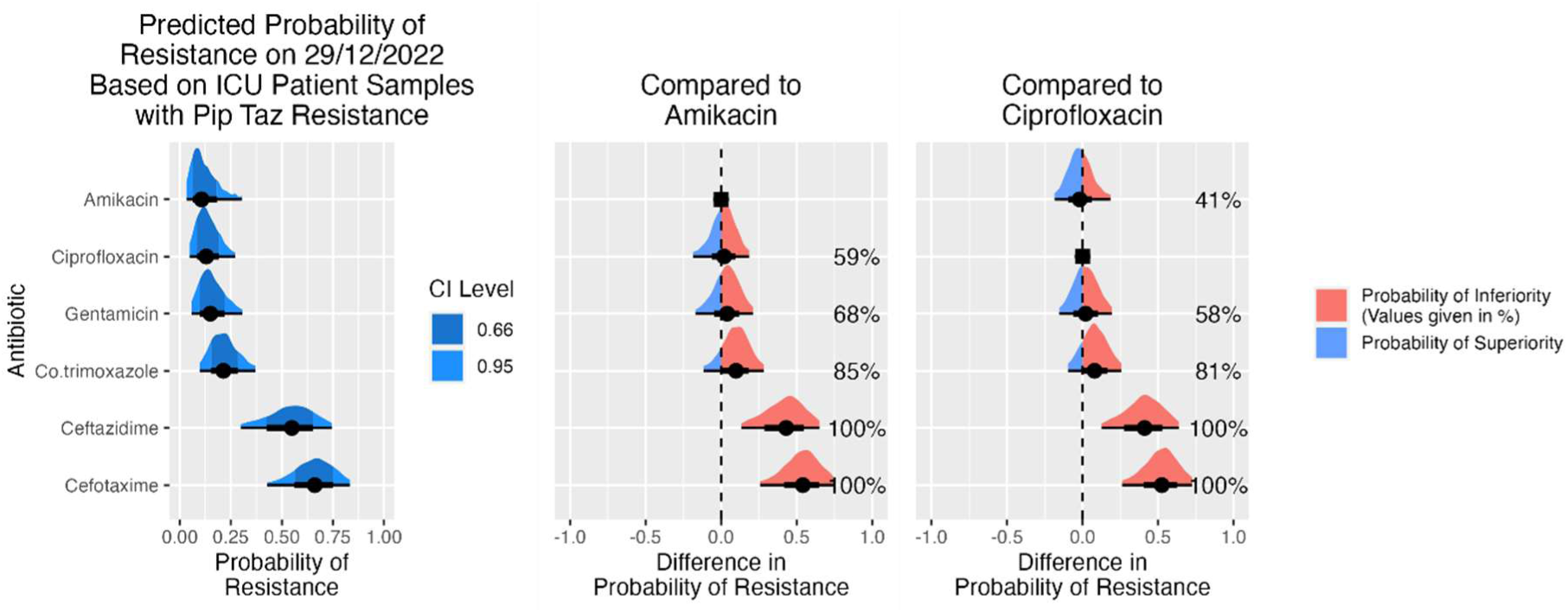
For ICU patients with assumed resistance to piperacillin/tazobactam. The estimated resistance rate to 6 antibiotic options with the 66% and 95% credible interval is shown on the left. The centre and right plots each show a head to head comparison between one antibiotic (Amikacin or Ciprofloxacin) and the other five antibiotics. This gives the estimated difference in probability of resistance, for example, there is 41% probability of higher resistance in amikacin compared to ciprofloxacin (right), only marginally favouring amikacin, while there is an 81% probability of higher resistance in co-trimoxazole compared to ciprofloxacin (right) favouring the use of ciprofloxacin.

It is worth noting that GNB BSI resistance rates to meropenem are low in this region (∼2%), where meropenem is invariably superior as an empiric choice. Our analysis has therefore focused on other ABs as meropenem-sparing alternatives. Similarly within our region, resistance rates to newer antibiotics such as Cetazidime-Avibactam or Meropenem-Vaborbactam are currently too low to allow inclusion in our analysis.

#### Escalation Antibiogram Example 1

In severe Gram-negative infections, two of the most commonly used β-lactams are piperacillin/tazobactam (first-line AB for neutropenic sepsis and widely used in ICU) and ceftazidime (a useful alternative to piperacillin/tazobactam especially in non-severe penicillin allergy). In our region, piperacillin/tazobactam and ceftazidime resistance rates among GNB BSIs are similar: 13.4% (95% CI 10.8 to 16.1) and 10.6% (95% CI 8.9 to 12.7) respectively. The merits of the addition of an aminoglycoside to a β-lactam has been much debated (12,13) but will inevitably depend on local resistance and co-resistance rates. We will look at the effect of resistance to piperacillin/tazobactam and ceftazidime on the probability of resistance to gentamicin (our mostly widely used aminoglycoside) and amikacin (which is rarely used locally) in two specific patient groups, ICU patients and haemato-oncology and compare these results with those generated from the whole hospital population.

When comparing a wide group of second line options when adding to, or switching from piperacillin/tazobactam or ceftazidime (Tables 2 and 3), ABs are ordered in preference for whole hospital populations, showing the percent resistant with 95% CI, and with the PPInf to that in the column on its left in the table. Values under 50% indicate the AB is superior to that on its left.

**Table 2.**
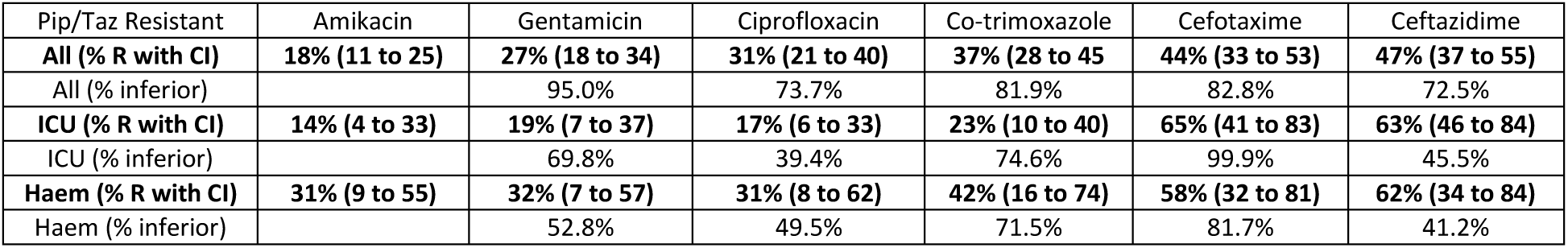
Escalation antibiogram for piperacillin/tazobactam (pip/taz) resistant isolates showing resistance rate and the posterior probability of inferiority (PPInf) to the antibiotic choice to the choice to the immediate left (e.g., Gentamicin v. Amikacin, Ciprofloxacin v. Gentamicin, etc.) across the whole hospital and in subgroups from ICU and Haematology/Oncology. Numbers in brackets indicate upper and lower bounds for the 95% credible interval.

**Table 3.** Escalation antibiogram for Ceftazidime resistant isolates showing resistance rate and the PPInf to the antibiotic choice to the choice to the immediate left (e.g., Gentamicin v. Amikacin, Ciprofloxacin v. Gentamicin, etc.) across the whole hospital and in subgroups from ICU and Haematology/Oncology. Numbers in brackets indicate upper and lower bounds for the 95% credible interval.

| Ceftazidime Resistant | Amikacin | Gentamicin | Co-trimoxazole | Ciprofloxacin | Pip/Taz | Cefotaxime |
| --- | --- | --- | --- | --- | --- | --- |
| **All (% R or I)** | **21% (12 to 30)** | **33% (24 to 41)** | **43% (33 to 52)** | **45% (35 to 54)** | **57% (43 to 68)** | **78% (67 to 86)** |
| All (% inferior) |  | 97.2% | 94.4% | 62.4% | 92.4% | 99.7% |
| **ICU (% R or I)** | **17% (4 to 42)** | **24% (7 to 44)** | **27% (10 to 49)** | **28% (11 to 52)** | **86% (66 to 98)** | **88% (70 to 98)** |
| ICU (% inferior) |  | 71.0% | 58.3% | 53.5% | 99.9% | 59.7% |
| **Haem (% R or I)** | **38% (18 to 62)** | **39% (19 to 64)** | **39% (16 to 64)** | **35% (13 to 61)** | **77% (54 to 92)** | **88% (69 to 98)** |
| Haem (% inferior) |  | 52.3% | 50.1% | 40.1% | 99.2% | 84.6% |

Resistance rates to both aminoglycosides are much higher in isolates resistant to either ceftazidime or piperacillin/tazobactam, with resistance rates vary from 14% to 39% depending on the patient group and antibiotic combinations compared to 3% to 13% aminoglycoside resistance in all isolates.

Within the whole hospital population, amikacin remains the best option for patients switching from either ceftazidime or piperacillin/tazobactam, with a 97% & 95% probability of superiority compared to gentamicin, although there is a greater relative increase in amikacin resistance compared to gentamicin (see Tables 2 and 3).

Within the ceftazidime and piperacillin/tazobactam resistant isolates, both the ICU and haemato-oncology populations differ from the whole hospital population with lower rates of aminoglycoside resistance in ICU, and higher rates of resistance in haemato-oncology patients (Fig 6). Within ICU patients amikacin is likely to be superior, PPSup = 69.8% if switching from piperacillin/tazobactam and PPSup = 71% if switching from ceftazidime. For the haemato-oncology patients there is little difference between amikacin and gentamicin, (PPSup = 52%) although there is a noticeably higher rate of resistance for both when switching from ceftazidime compared to piperacillin/tazobactam (also see plots in supplementary material).

**Figure 6.**
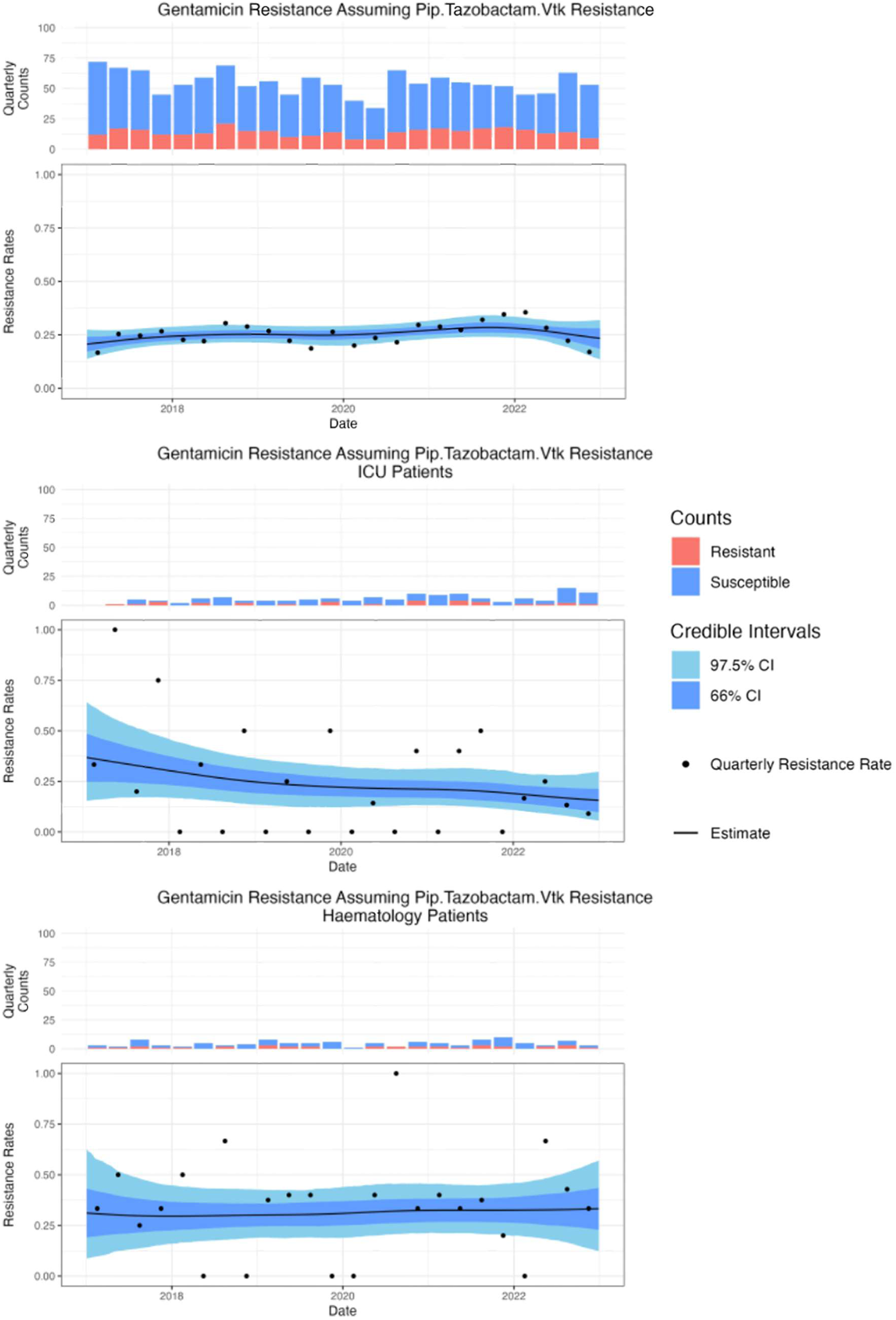
Percent Resistance to gentamicin assuming piperacillin/tazobactam resistance in (Top) all isolates 27.0% (95% CI 18.5 to 34.5), (Middle) ICU 19.2% (95% CI 7.1 to 37.0), (Bottom) haemato-oncology 33.5% (95% CI 14.6 to 57.5)

A difference between whole population and ICU or haemato-oncology populations are seen in a range of other antibiotics. Within the ICU patients, ciprofloxacin, trimethoprim/sulphamethoxazole and gentamicin have a similar probability of sensitivity, which is noticeably lower than in the whole hospital population. While within the haemato-oncology cohort, resistance rates to amikacin and gentamicin are higher than in the general population, and resistance rates to ciprofloxacin and trimethoprim/sulphamethoxazole are similar, despite both being used in some prophylaxis regimes. The resistance rates for all options are high (30-40%), so in a neutropenic patient, meropenem would be a more suitable alternative.

It is clear therefore, that sub-population analysis will be required by those intending to apply the EA in clinical practice.

#### Escalation Antibiogram Example 2

Having noted how changes in resistance over time differ between species, for example cefotaxime resistance in *E. coli* and non-*E. coli* (See supplementary material), we confirmed that this effect was also present in piperacillin/tazobactam resistant isolates. (Fig 7). We then looked at two clinical groups with a high proportion of *E. coli* infections (patients over 80 years old and patients with a urinary source of infection) to determine if they differed from the whole hospital population (Tables 4 and 5).

**Figure 7.**
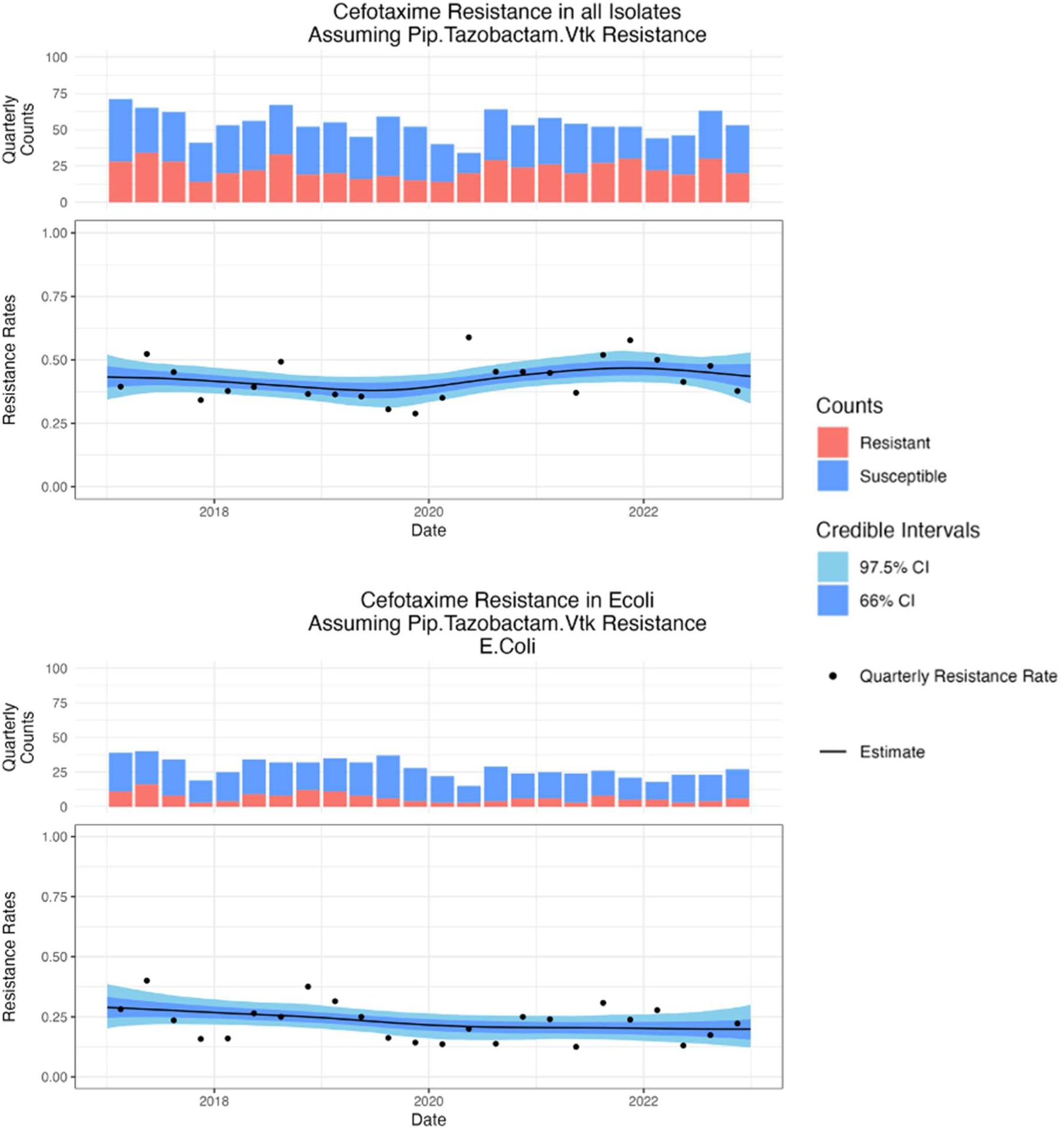
(Top) Cefotaxime resistance in all isolates assuming piperacillin/tazobactam resistance was stable over time 43.7% (95% CI 33 to 53) PPD = 46.1%. (Bottom) Cefotaxime resistance in E. coli assuming piperacillin/tazobactam resistance reduced from 29.2% (95%CI 20.4 to 38.6) to 18.3% (95%CI 11 to 28) PPD = 95.5%

**Table 4.** Escalation antibiogram for piperacillin/tazobactam resistant isolates showing resistance rate and the posterior probability of inferiority (PPInf) to the antibiotic choice to the the choice to the immediate left (e.g., Gentamicin v. Amikacin, Ciprofloxacin v. Gentamicin, etc.). Data for whole hospital, and subgroups of patients over 80 years, and infections with urine/renal source. Numbers in brackets indicate upper and lower bounds for the 95% credible interval.

| Pip/Taz Resistant | Amikacin | Gentamicin | Ciprofloxacin | Co-trimoxazole | Cefotaxime | Ceftazidime |
| --- | --- | --- | --- | --- | --- | --- |
| **All (% R or I)** | **18% (11 to 25)** | **27% (18 to 34)** | **31% (21 to 40)** | **37% (28 to 45)** | **44% (33 to 53)** | **47% (37 to 55)** |
| All (% inferior) |  | 95.0% | 73.7% | 81.9% | 82.8% | 72.5% |
| **Over 80 (% R or I)** | **16% (5 to 29)** | **19% (7 to 34)** | **39% (22 to 54)** | **39% (25 to 55)** | **37% (20 to 53)** | **43% (28 to 56)** |
| Over 80 (% inferior) |  | 60.0% | 94.6% | 48.2% | 43.6% | 67.3% |
| **U/R Source (% R or I)** | **13% (3 to 27)** | **29% (13 to 46)** | **33% (16 to 53)** | **39% (22 to 59)** | **29% (15 to 47)** | **36% (19 to 59)** |
| U/R Src (% inferior) |  | 93.2% | 63.5% | 68.3% | 20.9% | 68.4% |

**Table 5.** Escalation antibiogram for ceftazidime resistant isolates showing resistance rate and the posterior probability of inferiority (PPInf) to the antibiotic choice to the the choice to the immediate left (e.g., Gentamicin v. Amikacin, Ciprofloxacin v. Gentamicin, etc.). Data for whole hospital, andsubgroups of patients over 80 years, and infections with urine/renal source. Numbers in brackets indicate upper and lower bounds for the 95% credible interval.

| Ceftazidime Resistant | Amikacin | Gentamicin | Co-trimoxazole | Ciprofloxacin | Pip/Taz | Cefotaxime |
| --- | --- | --- | --- | --- | --- | --- |
| **All (% R or I)** | **21% (12 to 30)** | **33% (24 to 41)** | **43% (33 to 52)** | **45% (35 to 54)** | **57% (43 to 68)** | **78% (67 to 86)** |
| All (% inferior) |  | 97.2% | 94.4% | 62.4% | 92.4% | 99.7% |
| **Over 80 (% R or I)** | **21% (9 to 35)** | **25% (11 to 40)** | **44% (27 to 60)** | **44% (24 to 63)** | **49% (33 to 66)** | **77% (62 to 88)** |
| Over 80 (% inferior) |  | 68% | 94.3% | 52.7% | 60.9% | 99.3% |
| **U/R Source R or I)** | **13% (2 to 33)** | **34% (15 to 54)** | **48% (24 to 72)** | **57% (35 to 76)** | **49% (29 to 72)** | **80% (55 to 96)** |
| U/R Src (% inferior) |  | 93.7% | 80.8% | 72.2% | 28.3% | 96.6% |

Overall, the proportion of BSI caused by *E. coli* has very likely decreased from 59.1% (95%CI 54.9 to 63.8) at the start of 2017 to 51.4% (95%CI 47.1 to 55.5, PPD = 99.6%) at the end of 2022. Within patients over 80 years old, and patients with a BSI of urine or renal tract source we have seen a similar decrease in the proportion of *E. coli* BSI, to a current rate of 59.7% (95%CI 54.3 to 64.4) from 68% (95%CI 63.5 to 72.9, PPD = 99.3%) and 57.8 (95%CI 48.5 to 64.4) from 65.6% (95%CI 58.2 to 71.9, PPD = 94.5%) respectively.

When switching from piperacillin/tazobactam (Table 4) it appears that amikacin is the best option, but in circumstances where an aminoglycoside could not be used (e.g., poor renal function), cefotaxime is comparable to ciprofloxacin and superior to co-trimoxazole for urinary source infections.

We would not usually think of empirically changing from piperacillin/tazobactam to cefotaxime, as the former is usually considered to be broader spectrum, and it is often assumed that resistance to piperacillin/tazobactam would also confer resistance to 3^rd^ generation cephalosporins. While AmpC enzymes can confer resistance to both classes, when ESBL-producing isolates are reported as piperacillin/tazobactam resistant, this can be due to a range of β-lactamases which are not routinely identified in clinical isolates. β-Lactamases including OXA-1, inhibitor resistant TEM, or the high levels of TEM-1 can result in piperacillin/tazobactam resistance, independently of 3^rd^ generation cephalosporins resistance (14).

Assuming ceftazidime resistance (Table 5) showed that amikacin is clearly superior in the urinary source group, whereas for over 80s, amikacin or gentamicin are broadly comparable. Due to the high resistance rates of co-trimoxazole and ciprofloxacin, meropenem would be suitable if an aminoglycoside could not be used.

## Conclusions

Failure of empiric AB therapy is a regularly encountered clinical problem for infection speicialist. EA analysis informed by our Bayesian model is a useful tool to support clinician decision making in escalation of empiric AB therapy for the deteriorating patient when microbiology results are not available. The model produces a mean resistance rate, with 95% credible intervals for any AB option, and allows the calculation of a posterior probability that one AB choice is superior to another based on local ABR patterns.

The clinical application of this model requires the use of data appropriate to the patient group being treated and an understanding of the uncertainty in the data – this uncertainty will increase when applying the concept to relatively small patient groups. We conclude that within our region, the application of whole hospital data to groups with different underlying presentations and AB exposures such as haemato-oncology patients or paediatric patients is not appropriate. To overcome this shortcoming of simple EA analysis, we have focused on patient groups that can be determined before blood culture results are known, based on age, presumed source of infection, ICU patients or haemato-oncology patients. We have avoided the use of species-specific antibiograms, preferring to use those based on the mix of species found in the patient group under consideration. We note the differences in species proportions between groups appears to explain a significant amount of the resistance difference.

We anticipate that the results of our EA analysis in a given institution would be utilised by infection specialists, rather than general clinicians, to avoid potentially inappropriate AB choices, such as the use of aminoglycosides as a single agent where this contraindicated, and to ensure that alternative reasons for poor response to empiric AB therapy (e.g. lack of source control) are considered.

Our model can be easily introduced in any institution using the freely available software package, R. It requires only appropriately cleaned and de-duplicated ABR data which, in many cases, is already generated regularly to inform antimicrobial guideline development. This process of gathering data can also be automated, as in our centre, to further reduce the associated workload, and the model can be run as often as indicated by local demands. We suggest that this model is suitable for widespread adoption across institutions where infection specialists are making antimicrobial escalation decisions in the absence of helpful microbiological results.

## Data Availability

This was a retrospective study using anonymised data already collected as part of routine clinical care. This project was approved by Public Health England’s Research Ethics and Governance Group (REGG). Data produced in the present study are available upon reasonable request to the BNSSG ICB. To express interest contact the authors and BNSSG via and visit https://bnssg.icb.nhs.uk/about-us/research-and-evidence/

## Ethics

This was a retrospective study using anonymised data already collected as part of routine clinical care. This project was approved by Public Health England’s Research Ethics and Governance Group (REGG).

## Transparency declarations

All authors confirm that they have no conflicts of interest to declare.

## Author Contributions

Conceived the Study and Obtained Funding: P.W., A.W.D., M.B.A.

Cleaning and Analysis of Data: R.B., B.S., L.G., W.W.Y.L., A.W.D., P.W.

Initial Drafting of Manuscript: P.W., M.B.A.

Corrected and Approved Manuscript: All authors

