## Supplementary - AMR Percentages for "Rational Empiric Antibiotic Escalation Applied to Specific Patient Groups" (from another version of this preprint: version 2, 2023-11-07)

Supplementary Material (AMR Percentages)

Antibiotic resistance over time, with the probability of decreasing resistance between the start and end points (All patients)

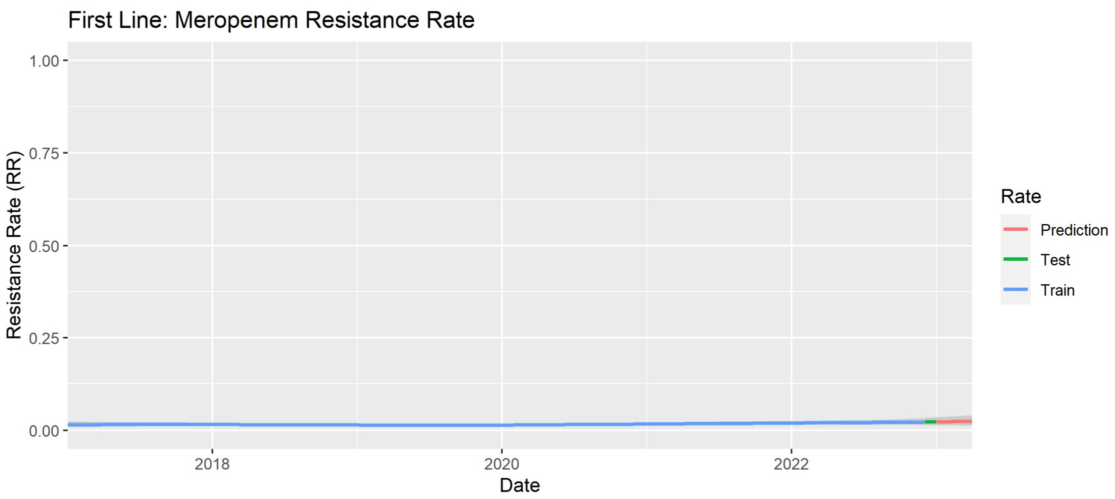


[1] "2017-01-01 Mid 0.015 (95%CI 0.008 to 0.024) Estimated Error 0.0039"

[1] "2022-12-02 Mid 0.022 (95%CI 0.014 to 0.033) Estimated Error 0.0050"

[1] "2022-12-31 Mid 0.022 (95%CI 0.013 to 0.035) Estimated Error 0.0055"

11.3% posterior probability of decrease/superiority

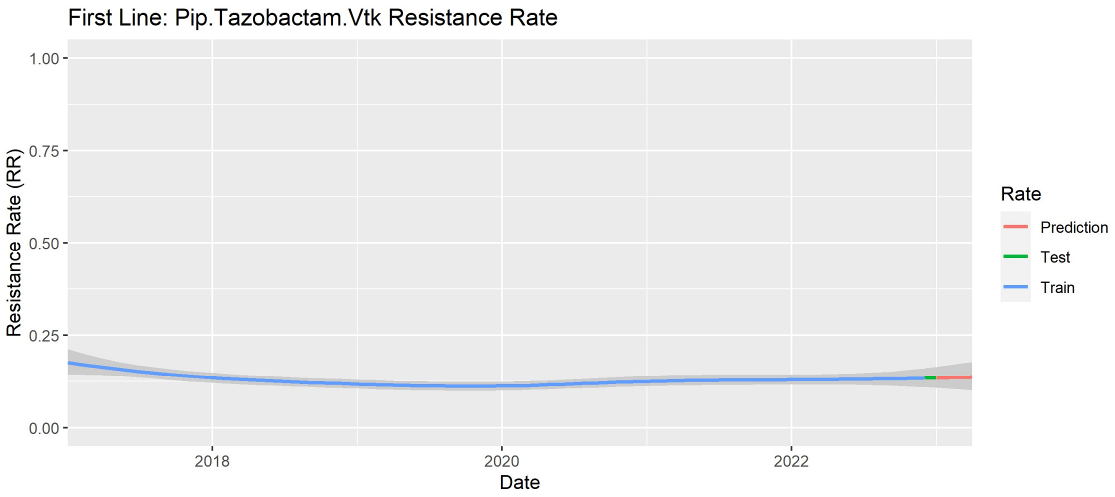


[1] "2017-01-01 Mid 0.176 (95%CI 0.145 to 0.213) Estimated Error 0.0171"

[1] "2022-12-02 Mid 0.134 (95%CI 0.110 to 0.161) Estimated Error 0.0130"

[1] "2022-12-31 Mid 0.135 (95%CI 0.108 to 0.164) Estimated Error 0.0144"

97.9% posterior probability of decrease/superiority

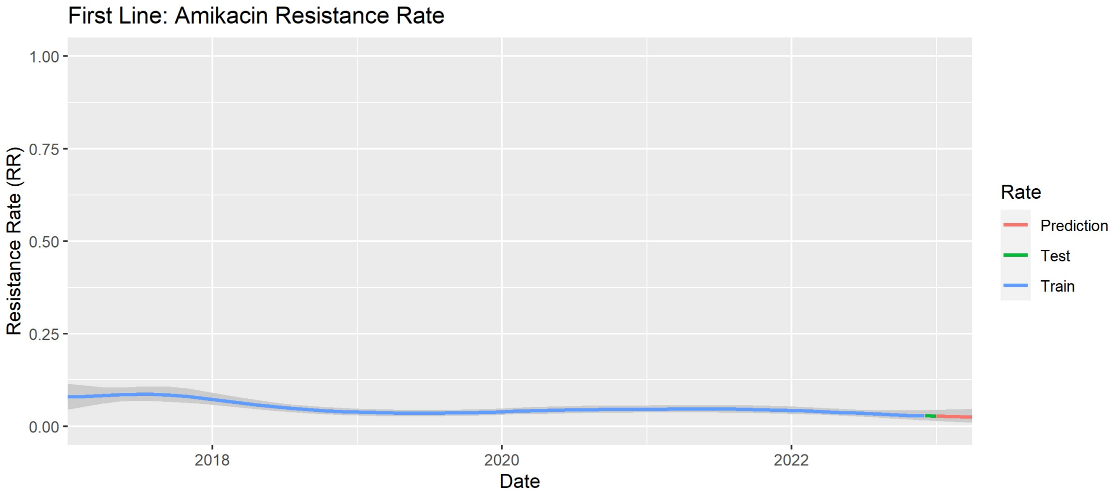


[1] "2017-01-01 Mid 0.079 (95%CI 0.045 to 0.115) Estimated Error 0.0179"

[1] "2022-12-03 Mid 0.028 (95%CI 0.016 to 0.043) Estimated Error 0.0072"

[1] "2022-12-31 Mid 0.027 (95%CI 0.014 to 0.044) Estimated Error 0.0077"

99.7% posterior probability of decrease/superiority

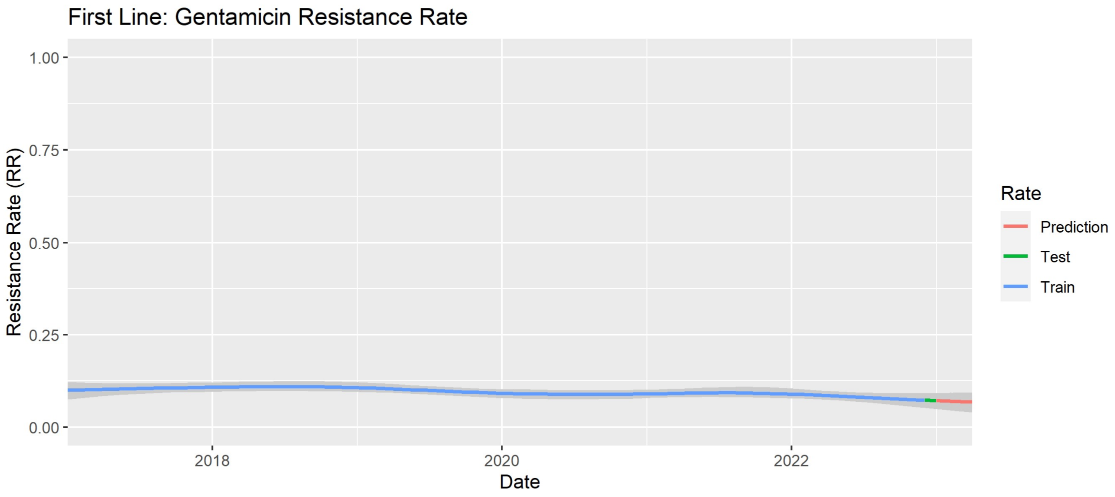


[1] "2017-01-01 Mid 0.099 (95%CI 0.075 to 0.122) Estimated Error 0.0123"

[1] "2022-12-02 Mid 0.072 (95%CI 0.052 to 0.091) Estimated Error 0.0103"

[1] "2022-12-31 Mid 0.071 (95%CI 0.048 to 0.091) Estimated Error 0.0112"

97.2% posterior probability of decrease/superiority

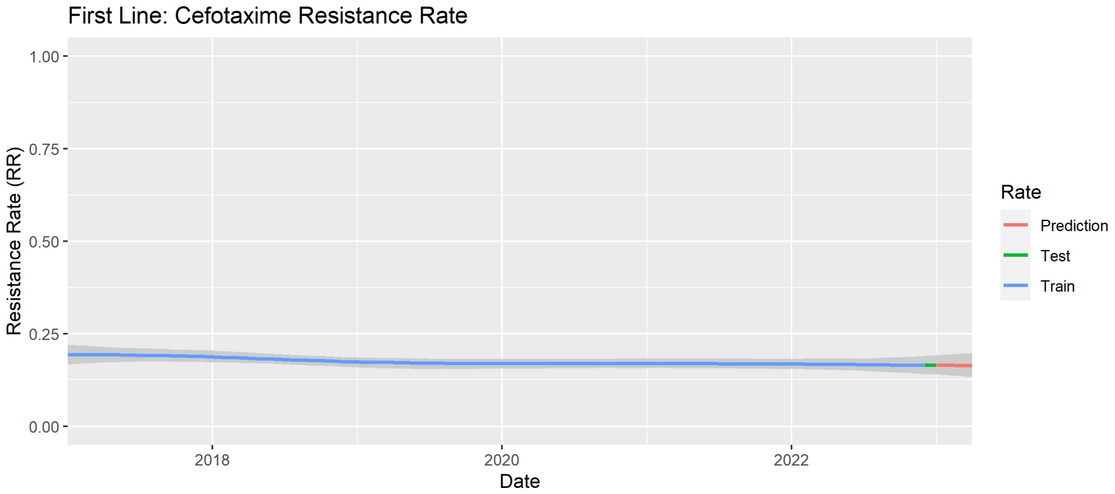


[1] "2017-01-01 Mid 0.194 (95%CI 0.168 to 0.221) Estimated Error 0.0135"

[1] "2022-12-02 Mid 0.165 (95%CI 0.142 to 0.190) Estimated Error 0.0121"

[1] "2022-12-31 Mid 0.165 (95%CI 0.139 to 0.192) Estimated Error 0.0130"

94.6% posterior probability of decrease/superiority

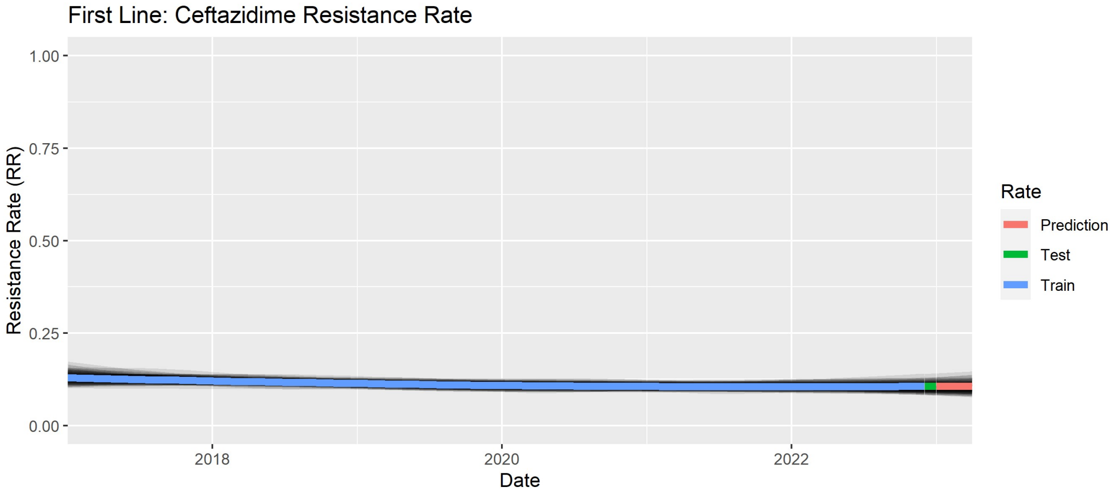


[1] "2017-01-01 Mid 0.129 (95%CI 0.109 to 0.153) Estimated Error 0.0112"

[1] "2022-12-02 Mid 0.106 (95%CI 0.089 to 0.128) Estimated Error 0.0097"

[1] "2022-12-31 Mid 0.106 (95%CI 0.088 to 0.129) Estimated Error 0.0103"

94.3% posterior probability of decrease/superiority

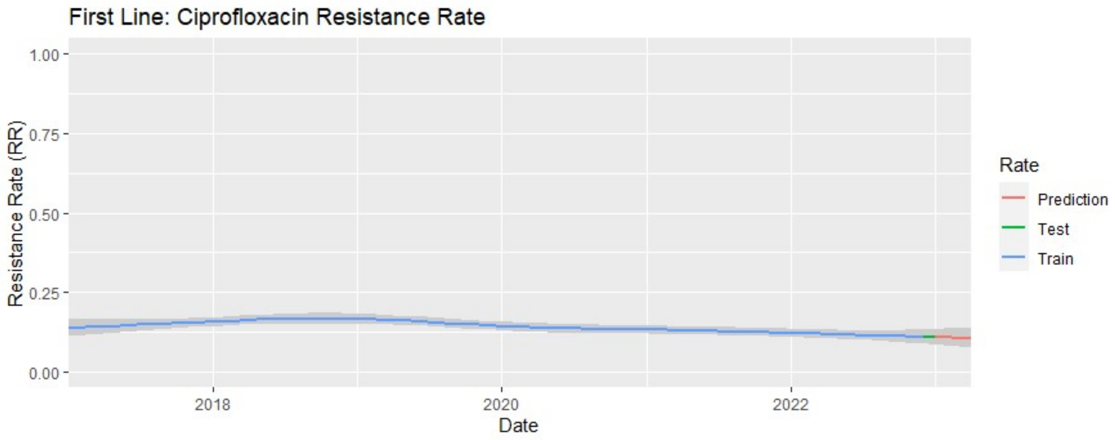


[1] "2017-01-01 Mid 0.139 (95%CI 0.111 to 0.168) Estimated Error 0.0144"

[1] "2022-12-02 Mid 0.109 (95%CI 0.086 to 0.132) Estimated Error 0.0115"

[1] "2022-12-31 Mid 0.108 (95%CI 0.083 to 0.133) Estimated Error 0.0125"

95.1% posterior probability of decrease/superiority

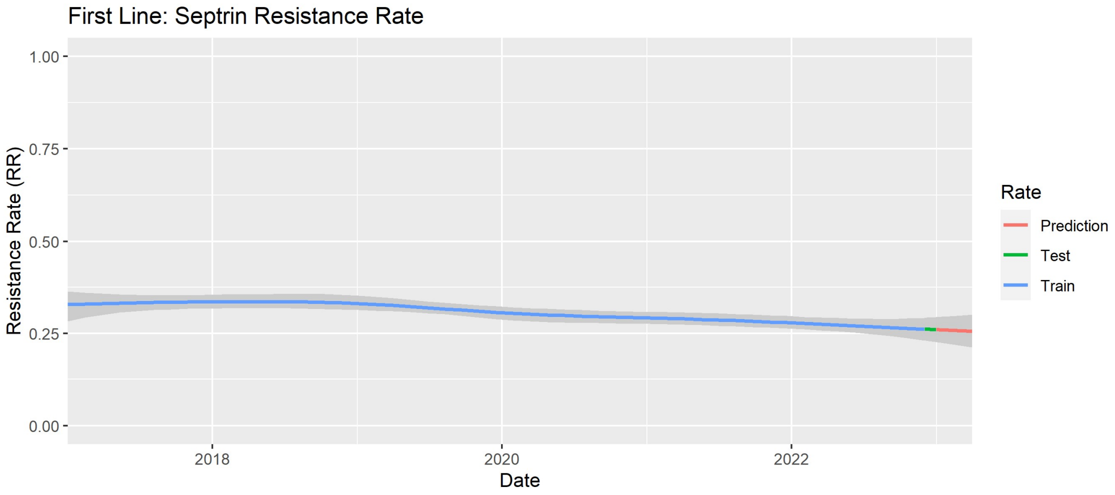


[1] "2017-01-01 Mid 0.329 (95%CI 0.283 to 0.364) Estimated Error 0.0203"

[1] "2022-12-02 Mid 0.262 (95%CI 0.231 to 0.292) Estimated Error 0.0153"

[1] "2022-12-31 Mid 0.260 (95%CI 0.227 to 0.294) Estimated Error 0.0167"

98.5% posterior probability of decrease/superiority

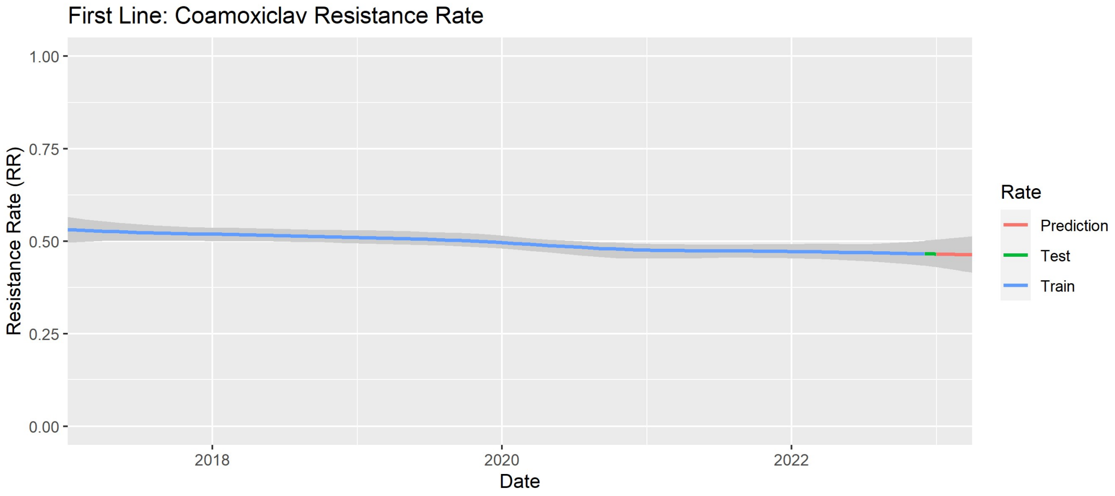


[1] "2017-01-01 Mid 0.531 (95%CI 0.497 to 0.566) Estimated Error 0.0169"

[1] "2022-12-02 Mid 0.466 (95%CI 0.434 to 0.502) Estimated Error 0.0169"

[1] "2022-12-31 Mid 0.465 (95%CI 0.430 to 0.504) Estimated Error 0.0183"

99.1% posterior probability of decrease/superiority

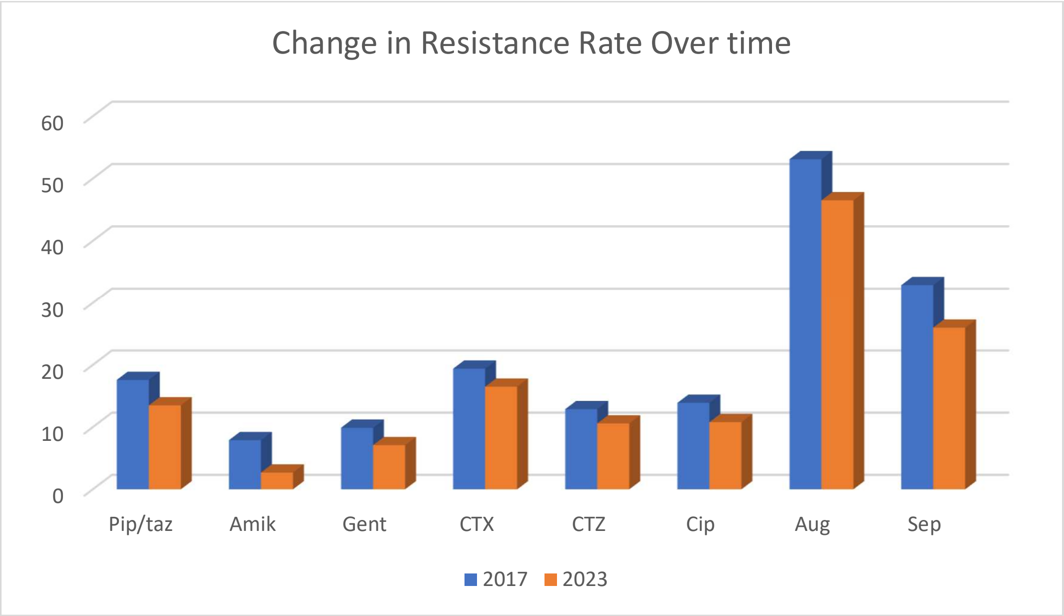

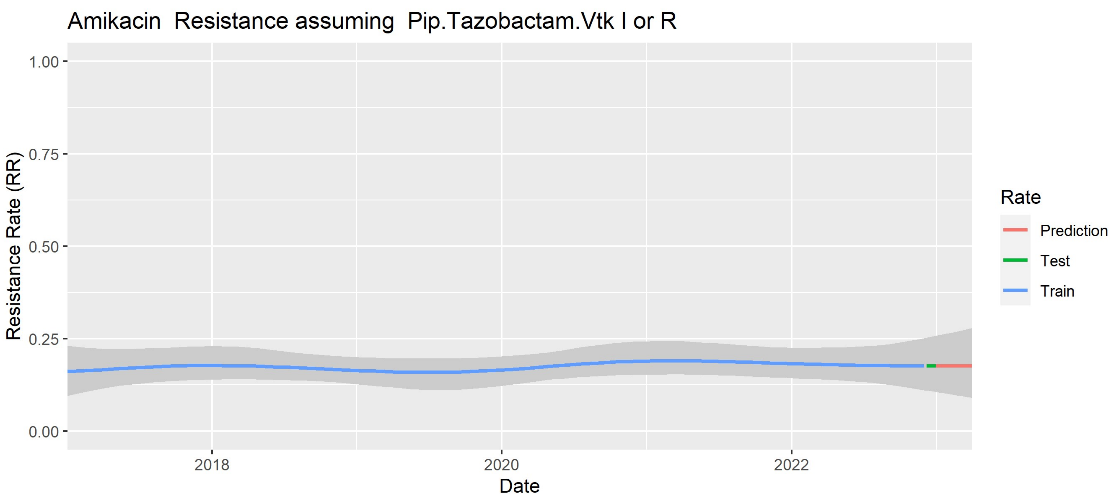


[1] "2017-01-02 Mid 0.161 (95%CI 0.095 to 0.231) Estimated Error 0.0341"

[1] "2022-12-06 Mid 0.177 (95%CI 0.109 to 0.253) Estimated Error 0.0358"

[1] "2022-12-30 Mid 0.177 (95%CI 0.105 to 0.258) Estimated Error 0.0380"

"37.0% posterior probability of decrease "

All patients

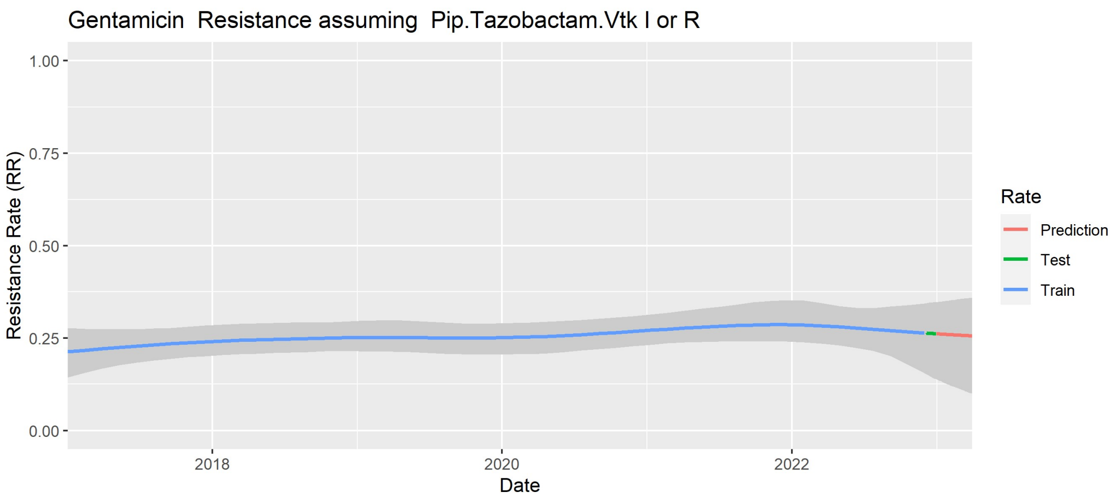


[1] "2017-01-01 Mid 0.214 (95%CI 0.145 to 0.278) Estimated Error 0.0333"

[1] "2022-12-06 Mid 0.263 (95%CI 0.153 to 0.344) Estimated Error 0.0466"

[1] "2022-12-30 Mid 0.262 (95%CI 0.139 to 0.347) Estimated Error 0.0500"

"16.8% posterior probability of decrease "

All patients

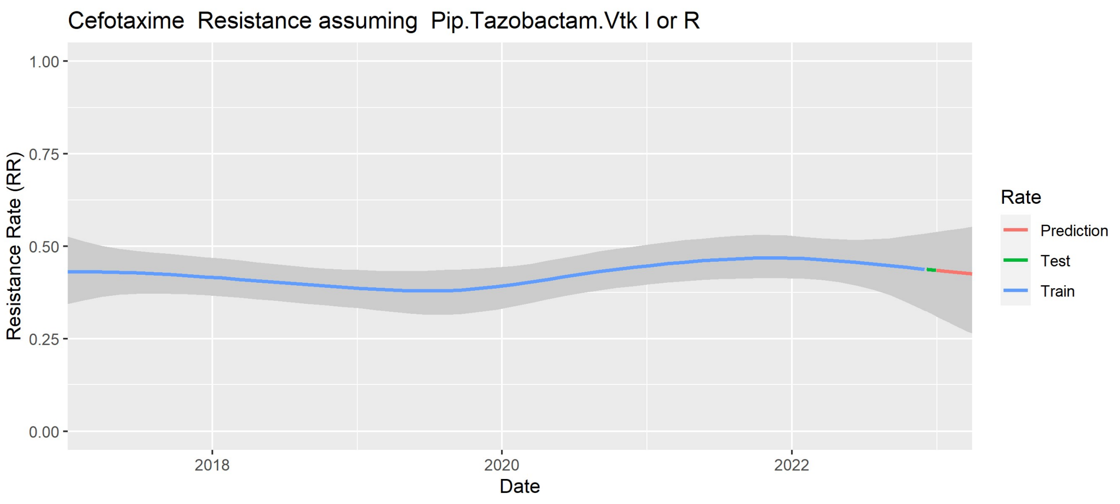


[1] "2017-01-01 Mid 0.432 (95%CI 0.344 to 0.526) Estimated Error 0.0461"

[1] "2022-12-06 Mid 0.437 (95%CI 0.324 to 0.534) Estimated Error 0.0535"

[1] "2022-12-30 Mid 0.435 (95%CI 0.311 to 0.538) Estimated Error 0.0577"

"44.4% posterior probability of decrease "

All patients

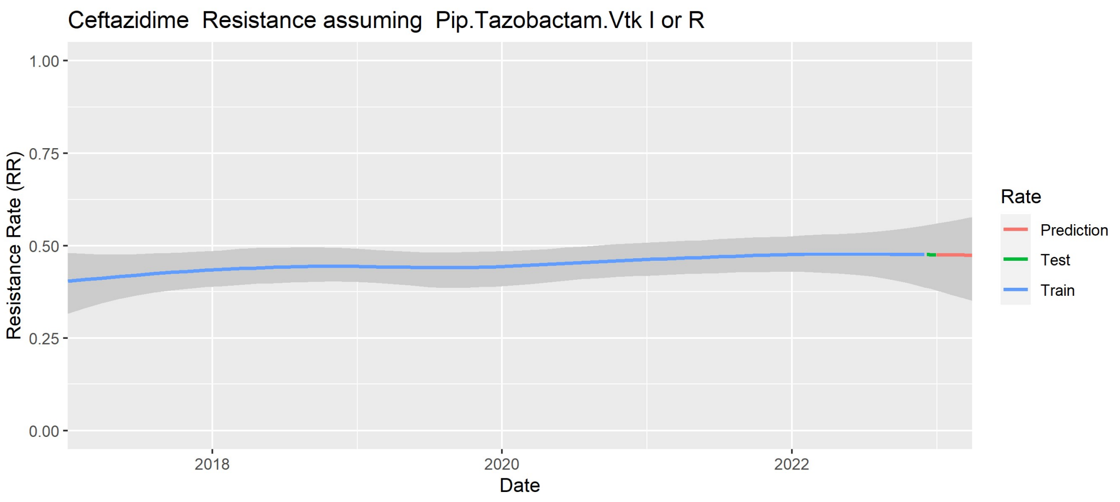


[1] "2017-01-01 Mid 0.404 (95%CI 0.316 to 0.479) Estimated Error 0.0407"

[1] "2022-12-06 Mid 0.475 (95%CI 0.385 to 0.555) Estimated Error 0.0423"

[1] "2022-12-30 Mid 0.474 (95%CI 0.378 to 0.559) Estimated Error 0.0448"

"10.6% posterior probability of decrease "

All patients

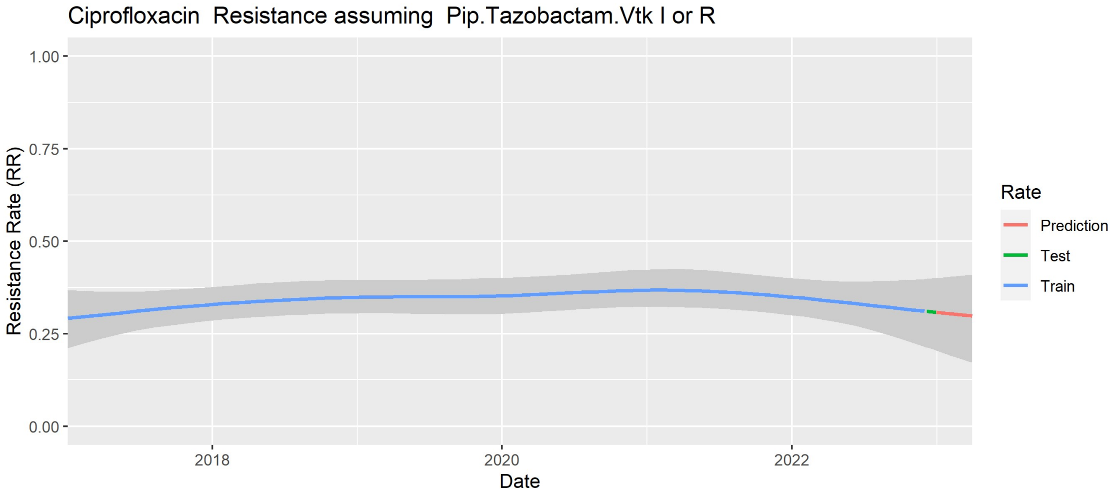


[1] "2017-01-01 Mid 0.292 (95%CI 0.212 to 0.368) Estimated Error 0.0398"

[1] "2022-12-06 Mid 0.311 (95%CI 0.213 to 0.399) Estimated Error 0.0484"

[1] "2022-12-30 Mid 0.308 (95%CI 0.205 to 0.400) Estimated Error 0.0512"

[1] "36.5% posterior probability of decrease "

All patients

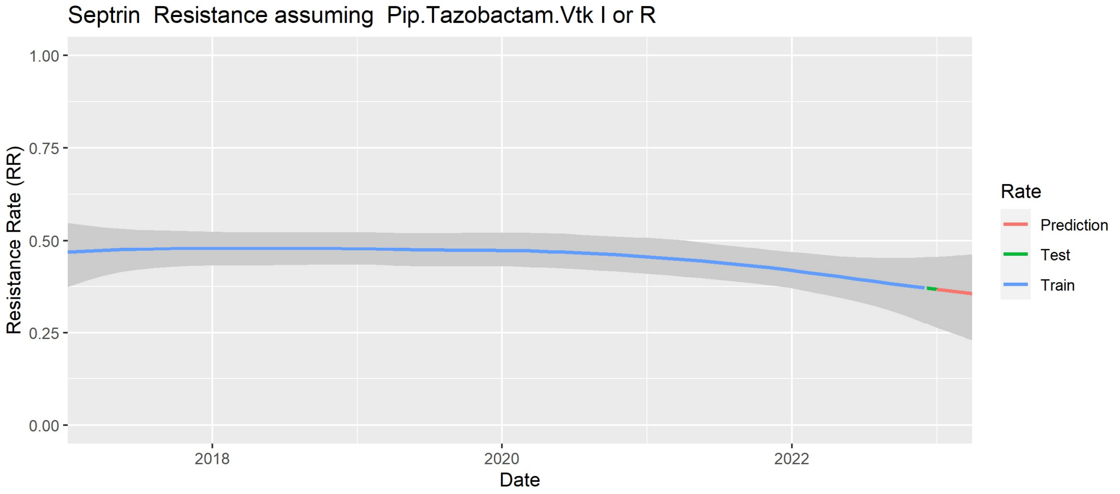


[1] "2017-01-01 Mid 0.468 (95%CI 0.374 to 0.547) Estimated Error 0.0438"

[1] "2022-12-06 Mid 0.371 (95%CI 0.275 to 0.454) Estimated Error 0.0459"

[1] "2022-12-30 Mid 0.368 (95%CI 0.265 to 0.455) Estimated Error 0.0486"

[1] "94.6% posterior probability of decrease "

All patients

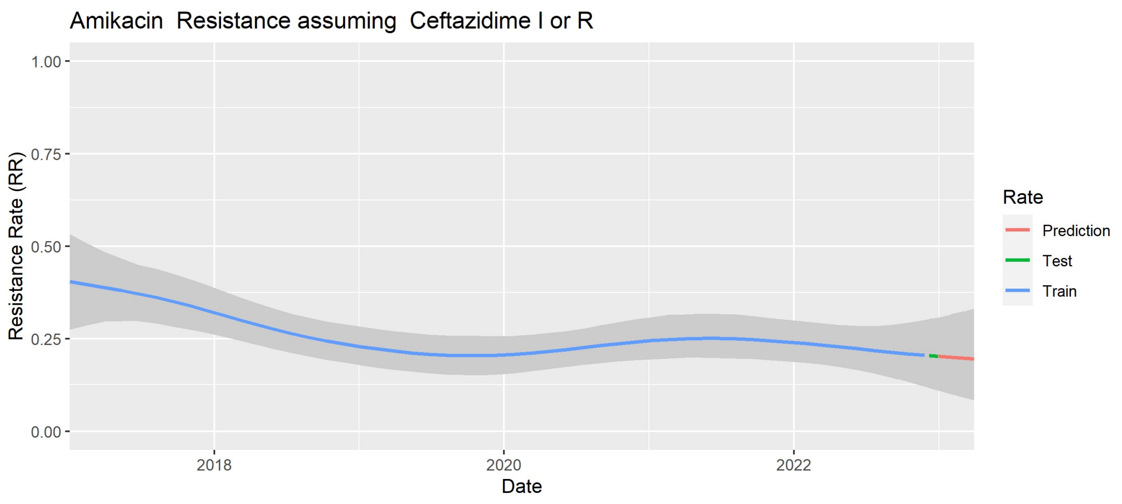


[1] "2017-01-02 Mid 0.404 (95%CI 0.275 to 0.532) Estimated Error 0.0649"

[1] "2022-12-07 Mid 0.205 (95%CI 0.116 to 0.304) Estimated Error 0.0476"

[1] "2022-12-30 Mid 0.203 (95%CI 0.109 to 0.307) Estimated Error 0.0506"

[1] "99.0% posterior probability of decrease "

All patients

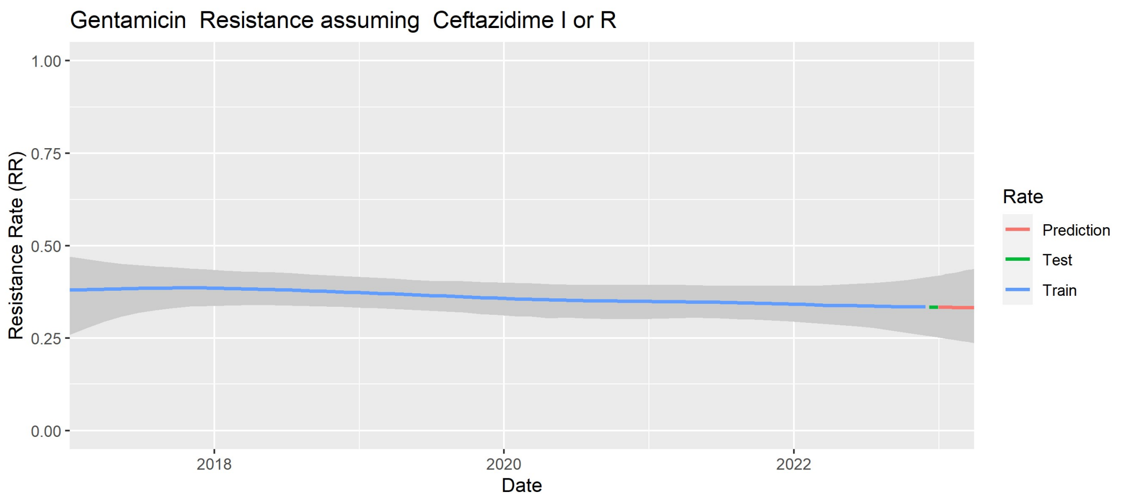


[1] "2017-01-01 Mid 0.380 (95%CI 0.259 to 0.469) Estimated Error 0.0494"

[1] "2022-12-07 Mid 0.334 (95%CI 0.256 to 0.415) Estimated Error 0.0401"

[1] "2022-12-30 Mid 0.333 (95%CI 0.252 to 0.418) Estimated Error 0.0419"

"77.9% posterior probability of decrease "

All patients

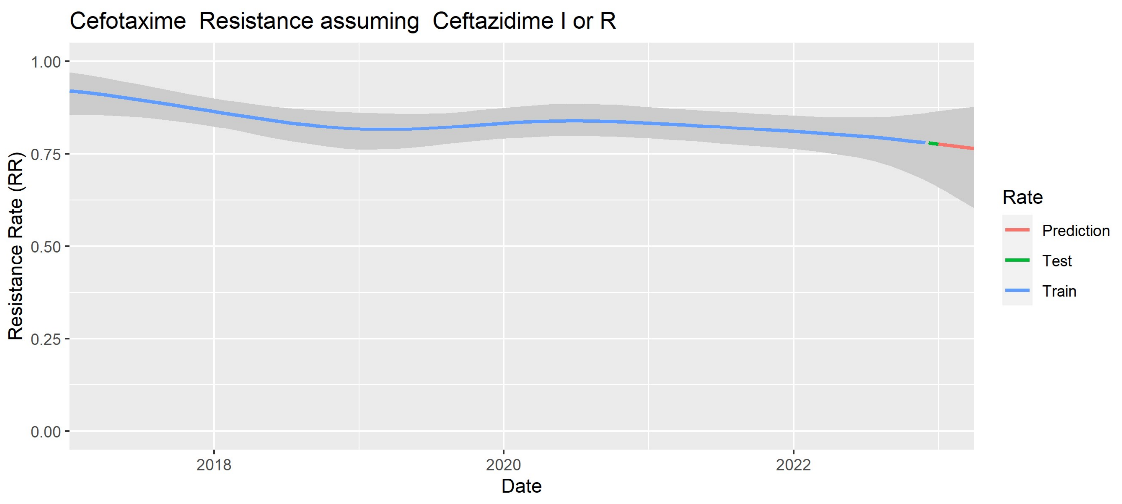


[1] "2017-01-01 Mid 0.921 (95%CI 0.855 to 0.971) Estimated Error 0.0303"

[1] "2022-12-07 Mid 0.779 (95%CI 0.673 to 0.862) Estimated Error 0.0480"

[1] "2022-12-30 Mid 0.776 (95%CI 0.660 to 0.866) Estimated Error 0.0520"

[1] "99.5% posterior probability of decrease "

All patients

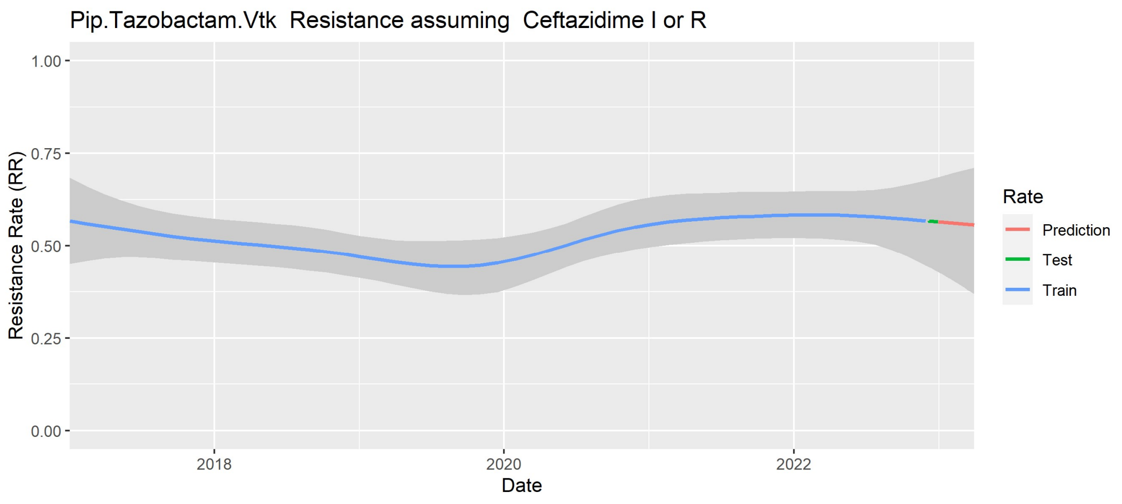


[1] "2017-01-01 Mid 0.567 (95%CI 0.450 to 0.683) Estimated Error 0.0600"

[1] "2022-12-07 Mid 0.566 (95%CI 0.441 to 0.678) Estimated Error 0.0603"

[1] "2022-12-30 Mid 0.564 (95%CI 0.428 to 0.684) Estimated Error 0.0654"

[1] "49.2% posterior probability of decrease "

All patients

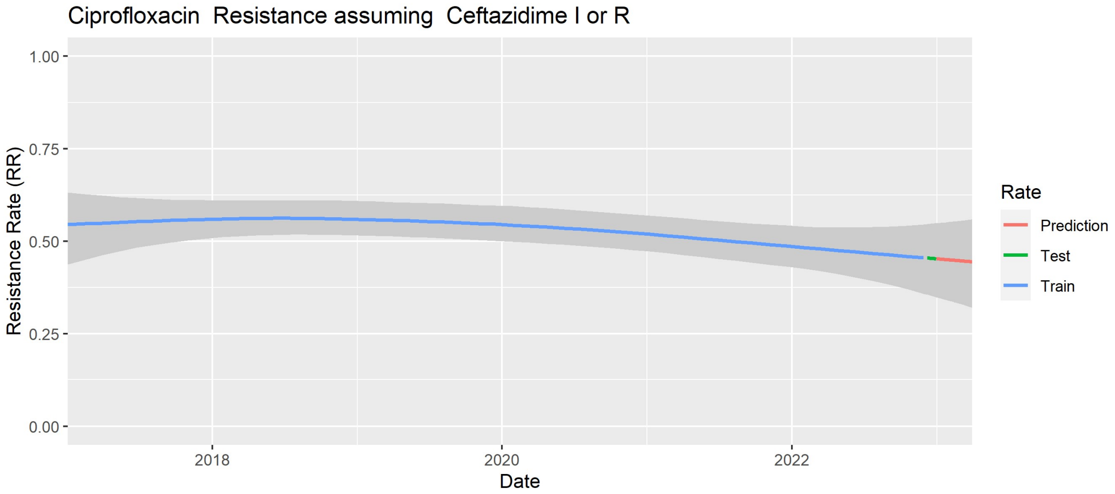


[1] "2017-01-01 Mid 0.545 (95%CI 0.437 to 0.632) Estimated Error 0.0502"

[1] "2022-12-07 Mid 0.455 (95%CI 0.356 to 0.546) Estimated Error 0.0476"

[1] "2022-12-30 Mid 0.453 (95%CI 0.349 to 0.549) Estimated Error 0.0498"

[1] "90.7% posterior probability of decrease "

All patients

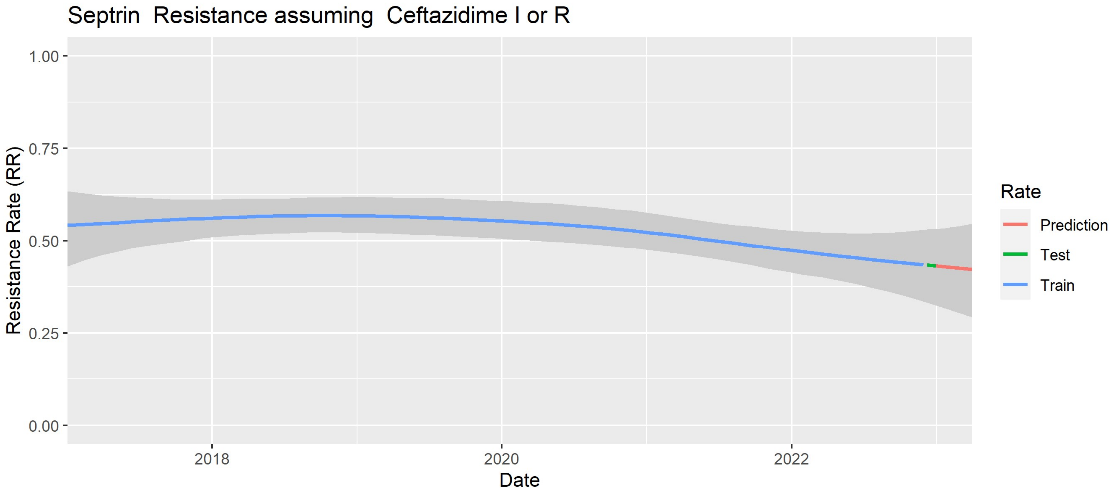


[1] "2017-01-01 Mid 0.542 (95%CI 0.430 to 0.634) Estimated Error 0.0519"

[1] "2022-12-07 Mid 0.433 (95%CI 0.332 to 0.531) Estimated Error 0.0496"

[1] "2022-12-30 Mid 0.431 (95%CI 0.324 to 0.532) Estimated Error 0.0522"

[1] "94.2% posterior probability of decrease "

All patients
