## Supplementary - Isolate Percentages for "Rational Empiric Antibiotic Escalation Applied to Specific Patient Groups" (from another version of this preprint: version 2, 2023-11-07)

Supplementary Material (Isolate Percentages)

Isolate Percentages Over time.

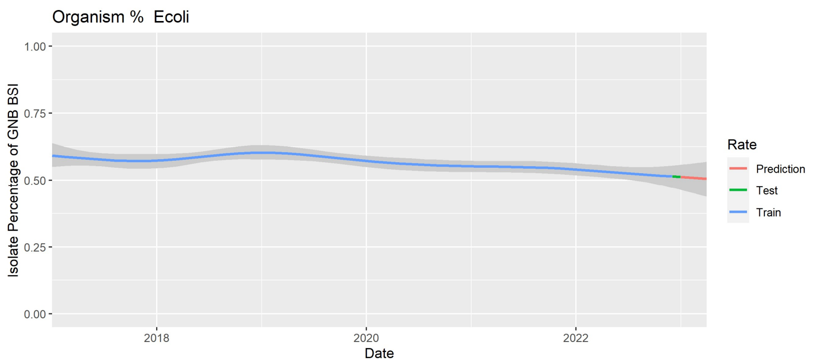


[1] "2017-01-01 Mid 0.592 (95%CI 0.550 to 0.638) Estimated Error 0.0224"

[1] "2022-12-02 Mid 0.514 (95%CI 0.471 to 0.554) Estimated Error 0.0210"

[1] "2022-12-31 Mid 0.512 (95%CI 0.463 to 0.557) Estimated Error 0.0236"

99.5% Posterior Probability of decrease from 2017 to 2023

E. coli   All patients

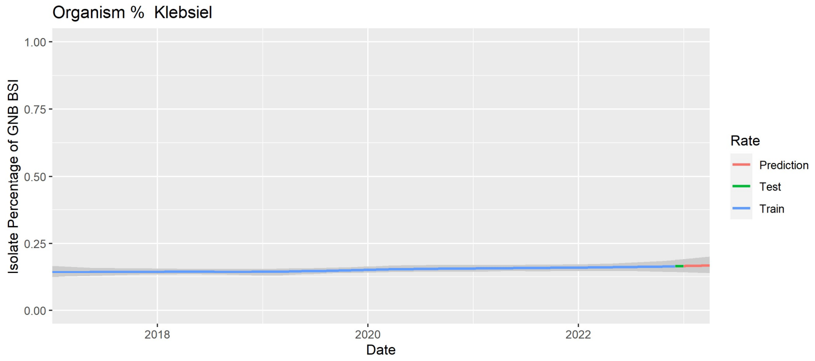


[1] "2017-01-01 Mid 0.144 (95%CI 0.125 to 0.167) Estimated Error 0.0108"

[1] "2022-12-02 Mid 0.166 (95%CI 0.144 to 0.190) Estimated Error 0.0113"

[1] "2022-12-31 Mid 0.167 (95%CI 0.144 to 0.192) Estimated Error 0.0122"

8.2% x greater than y

Klebsiells sp.  All patients

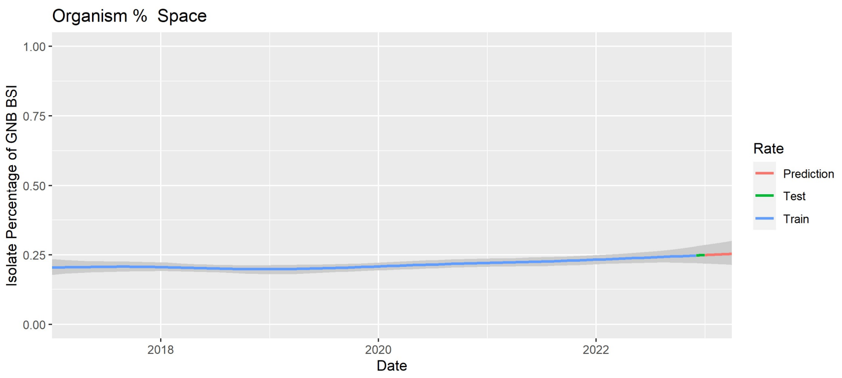


[1] "2017-01-01 Mid 0.205 (95%CI 0.179 to 0.235) Estimated Error 0.0143"

[1] "2022-12-02 Mid 0.248 (95%CI 0.220 to 0.281) Estimated Error 0.0151"

[1] "2022-12-31 Mid 0.250 (95%CI 0.219 to 0.286) Estimated Error 0.0165"

1.8% Posterior Probability of decrease from 2017 to 2023

Space Organisms   All patients

#####################################################################

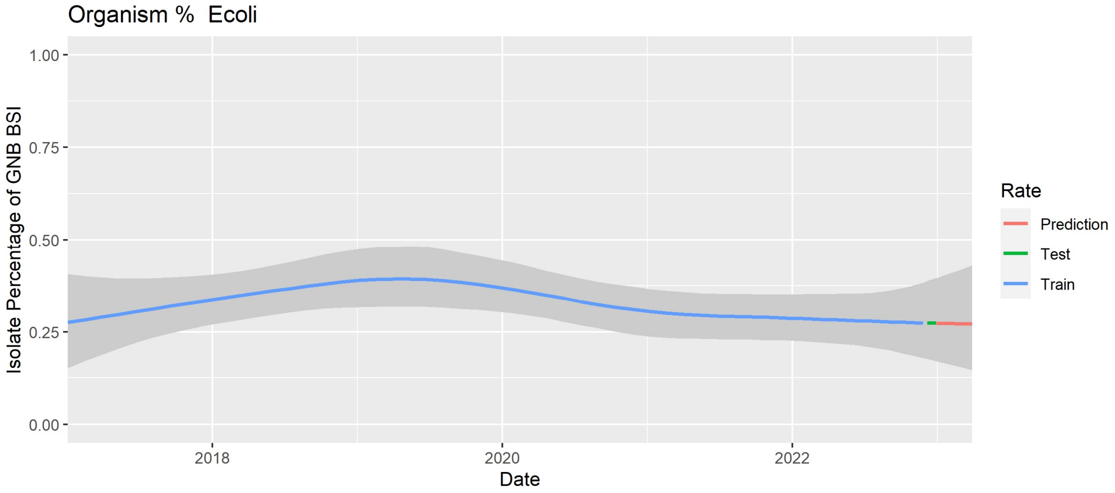


[1] "2017-01-01 Mid 0.276 (95%CI 0.153 to 0.407) Estimated Error 0.0661"

[1] "2022-12-06 Mid 0.274 (95%CI 0.177 to 0.388) Estimated Error 0.0533"

[1] "2022-12-29 Mid 0.274 (95%CI 0.171 to 0.395) Estimated Error 0.0567"

52.0% Posterior Probability of decrease from 2017 to 2023

E. coli  ICU

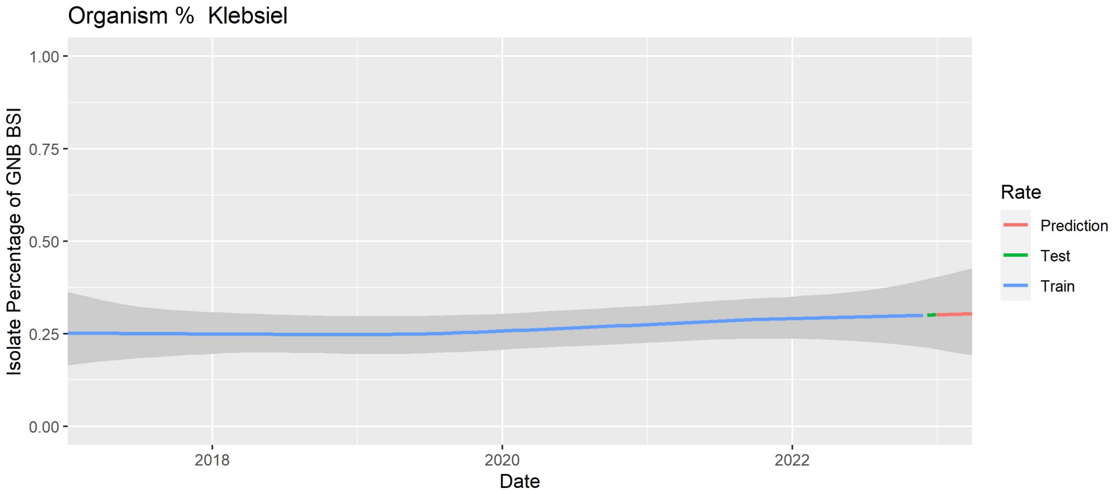


[1] "2017-01-01 Mid 0.252 (95%CI 0.165 to 0.363) Estimated Error 0.0498"

[1] "2022-12-06 Mid 0.300 (95%CI 0.213 to 0.398) Estimated Error 0.0464"

[1] "2022-12-29 Mid 0.301 (95%CI 0.209 to 0.403) Estimated Error 0.0486"

24.7% Posterior Probability of decrease from 2017 to 2023

Klebsiells sp. ICU

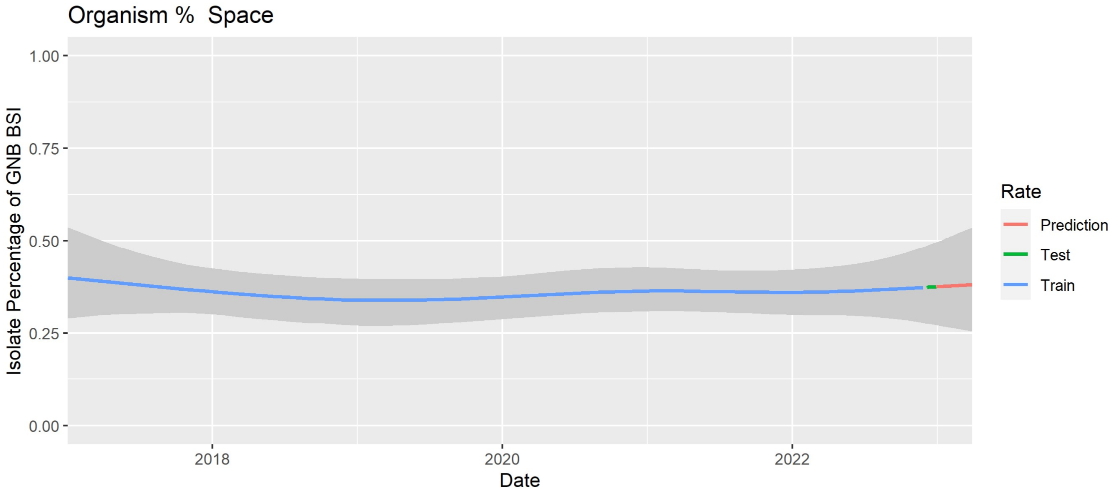


[1] "2017-01-01 Mid 0.399 (95%CI 0.291 to 0.537) Estimated Error 0.0637"

[1] "2022-12-06 Mid 0.373 (95%CI 0.276 to 0.486) Estimated Error 0.0525"

[1] "2022-12-29 Mid 0.375 (95%CI 0.272 to 0.495) Estimated Error 0.0556"

61.5% Posterior Probability of decrease from 2017 to 2023

SPACE ICU

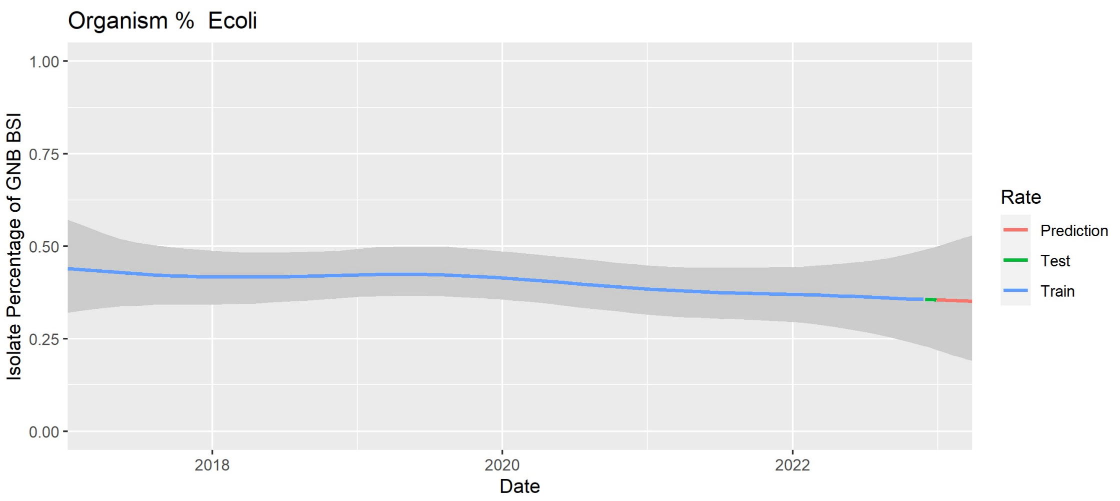


[1] "2017-01-02 Mid 0.440 (95%CI 0.321 to 0.572) Estimated Error 0.0630"

[1] "2022-11-30 Mid 0.356 (95%CI 0.230 to 0.491) Estimated Error 0.0659"

[1] "2022-12-28 Mid 0.355 (95%CI 0.220 to 0.499) Estimated Error 0.0698"

81.9% Posterior Probability of decrease from 2017 to 2023

E. coli Haemato-Oncology

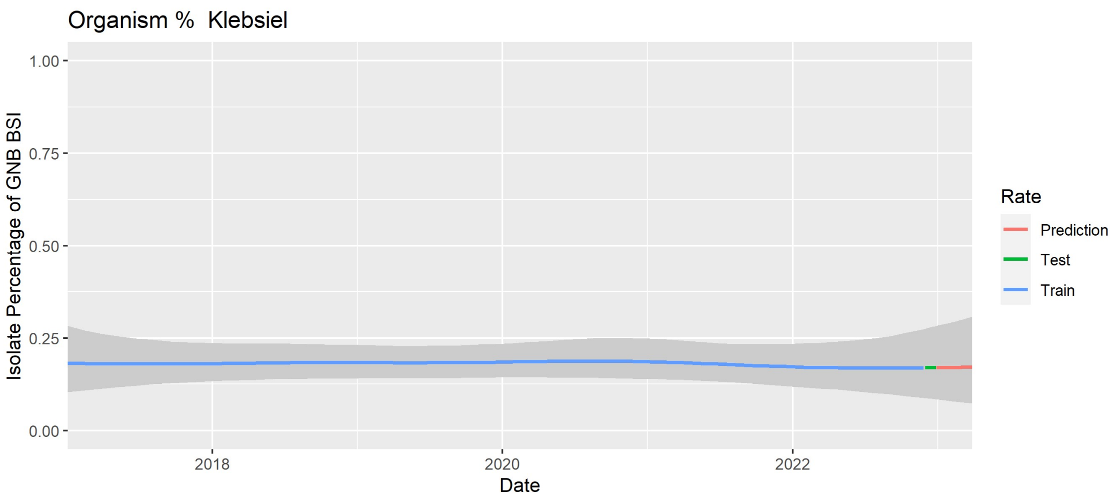


[1] "2017-01-02 Mid 0.183 (95%CI 0.104 to 0.283) Estimated Error 0.0453"

[1] "2022-11-30 Mid 0.170 (95%CI 0.087 to 0.275) Estimated Error 0.0473"

[1] "2022-12-28 Mid 0.171 (95%CI 0.084 to 0.282) Estimated Error 0.0499"

57.6% Posterior Probability of decrease from 2017 to 2023

Klebsiella sp. Haemato-Oncology

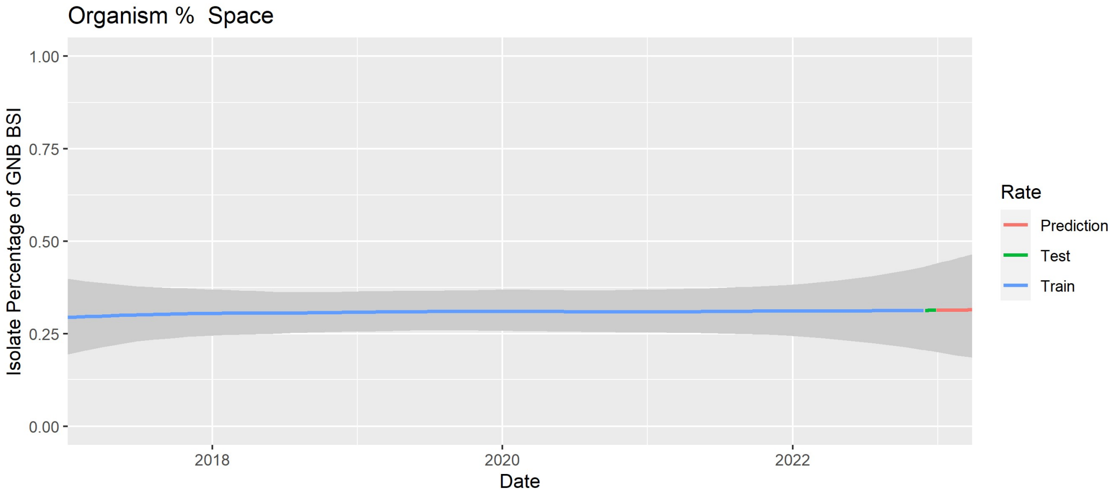


[1] "2017-01-02 Mid 0.295 (95%CI 0.194 to 0.399) Estimated Error 0.0519"

[1] "2022-11-30 Mid 0.313 (95%CI 0.206 to 0.432) Estimated Error 0.0573"

[1] "2022-12-28 Mid 0.314 (95%CI 0.201 to 0.439) Estimated Error 0.0602"

40.9% Posterior Probability of decrease from 2017 to 2023

SPACE organisms Haemato-Oncology

######################################################################

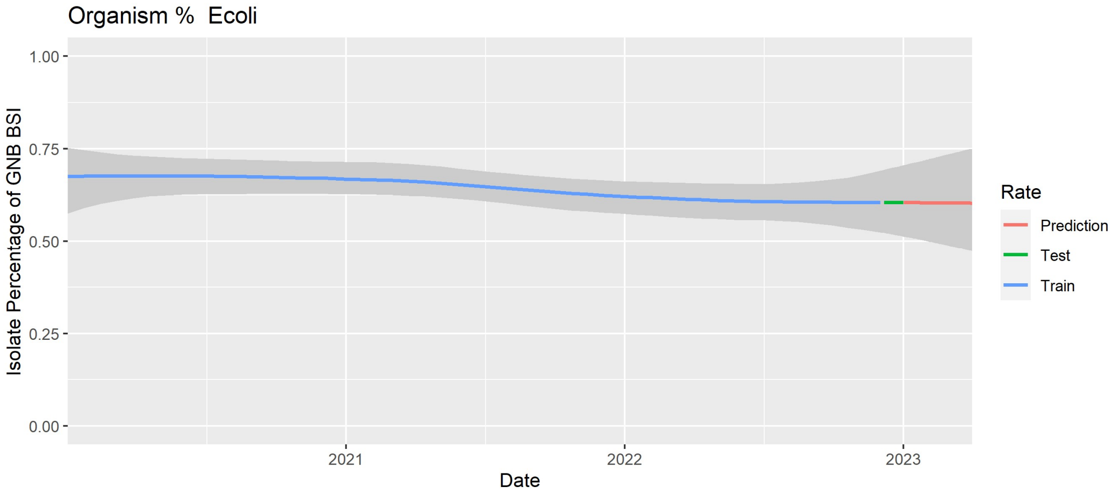


[1] "2020-01-02 Mid 0.675 (95%CI 0.575 to 0.751) Estimated Error 0.0452"

[1] "2022-12-06 Mid 0.604 (95%CI 0.522 to 0.693) Estimated Error 0.0426"

[1] "2022-12-31 Mid 0.604 (95%CI 0.512 to 0.705) Estimated Error 0.0478"

86.7% Posterior Probability of decrease from 2017 to 2023

GI source E. coli

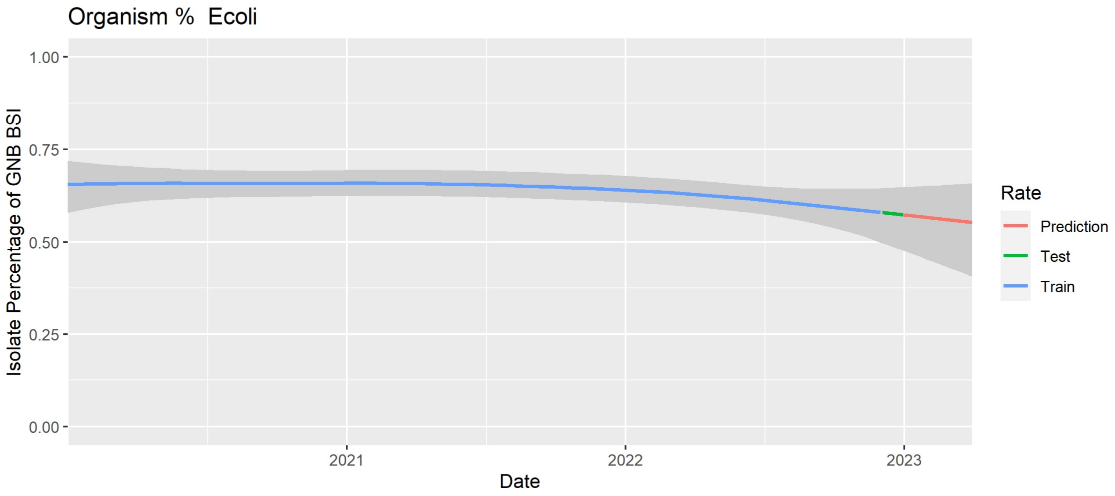


1] "2020-01-01 Mid 0.656 (95%CI 0.579 to 0.719) Estimated Error 0.0354"

[1] "2022-12-03 Mid 0.580 (95%CI 0.497 to 0.646) Estimated Error 0.0379"

[1] "2022-12-30 Mid 0.573 (95%CI 0.476 to 0.648) Estimated Error 0.0434"

"94.3% x greater than y

Urine source E. coli

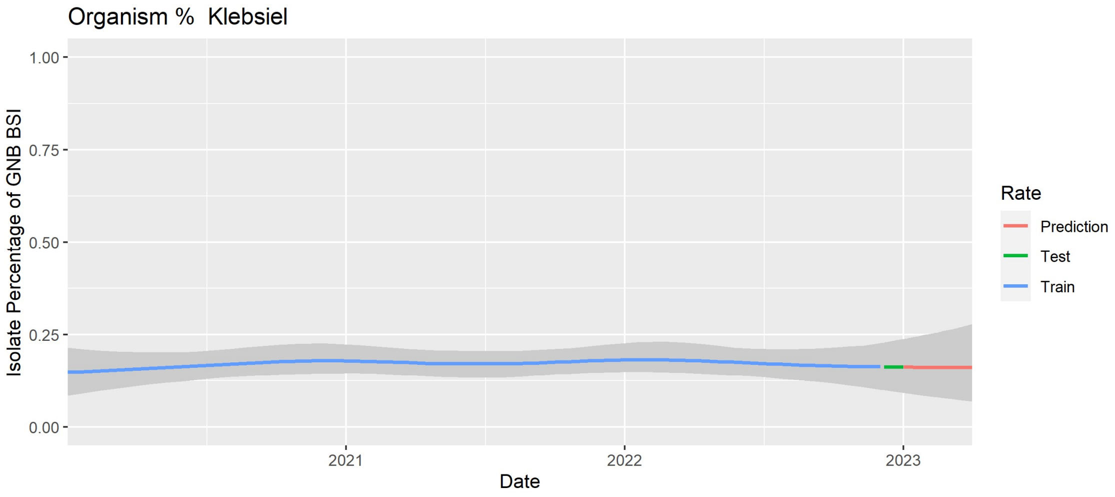


[1] "2020-01-02 Mid 0.149 (95%CI 0.085 to 0.215) Estimated Error 0.0335"

[1] "2022-12-06 Mid 0.163 (95%CI 0.099 to 0.229) Estimated Error 0.0327"

[1] "2022-12-31 Mid 0.163 (95%CI 0.092 to 0.238) Estimated Error 0.0365"

37.2% Posterior Probability of decrease from 2017 to 2023

GI Klebsiells sp.

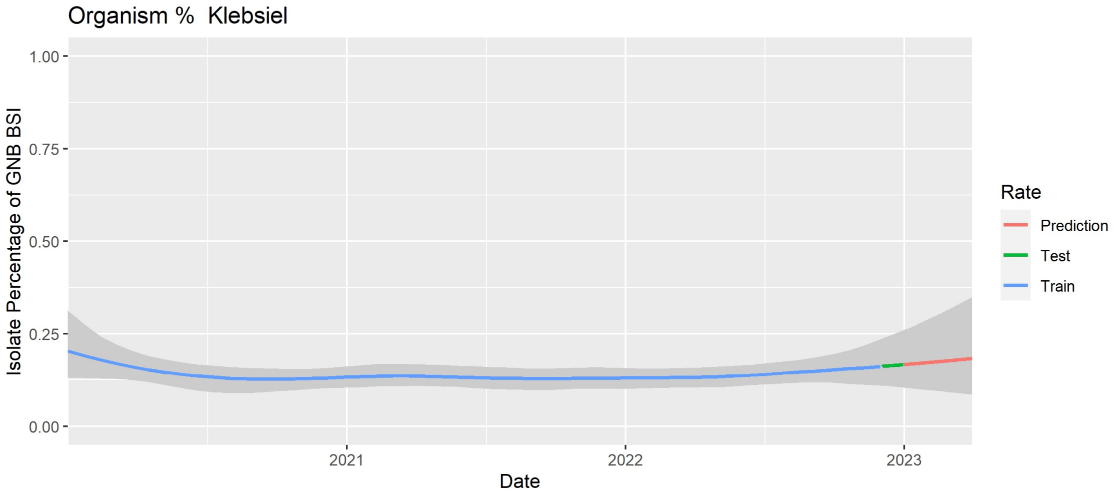


[1] "2020-01-01 Mid 0.204 (95%CI 0.130 to 0.313) Estimated Error 0.0486"

[1] "2022-12-03 Mid 0.162 (95%CI 0.109 to 0.238) Estimated Error 0.0325"

[1] "2022-12-30 Mid 0.167 (95%CI 0.104 to 0.260) Estimated Error 0.0395"

77.6% Posterior Probability of decrease from 2017 to 2023

Urine Source  Klebsiells sp.

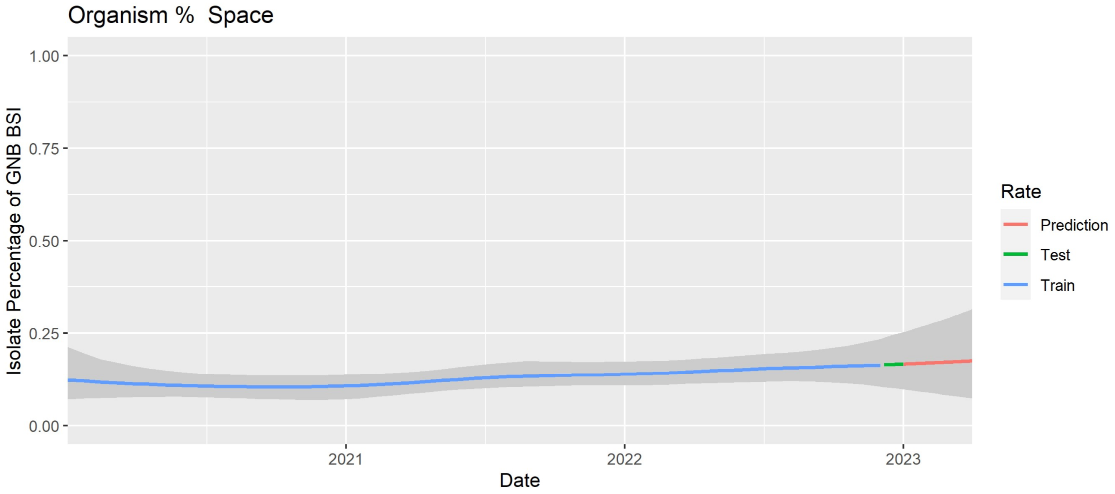


[1] "2020-01-02 Mid 0.124 (95%CI 0.071 to 0.213) Estimated Error 0.0362"

[1] "2022-12-06 Mid 0.165 (95%CI 0.103 to 0.238) Estimated Error 0.0338"

[1] "2022-12-31 Mid 0.167 (95%CI 0.097 to 0.253) Estimated Error 0.0388"

19.3% Posterior Probability of decrease from 2017 to 2023

GI Source  SPACE Organisms

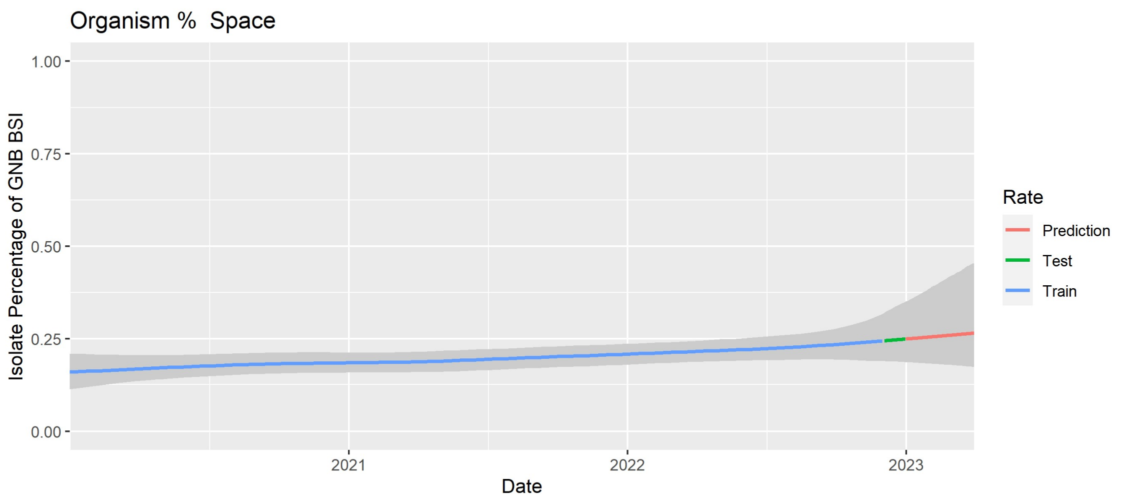


[1] "2020-01-01 Mid 0.161 (95%CI 0.113 to 0.211) Estimated Error 0.0245"

[1] "2022-12-03 Mid 0.245 (95%CI 0.190 to 0.321) Estimated Error 0.0322"

[1] "2022-12-30 Mid 0.249 (95%CI 0.187 to 0.350) Estimated Error 0.0385"

1.6% Posterior Probability of decrease from 2017 to 2023

Urine Source  SPACE Organisms

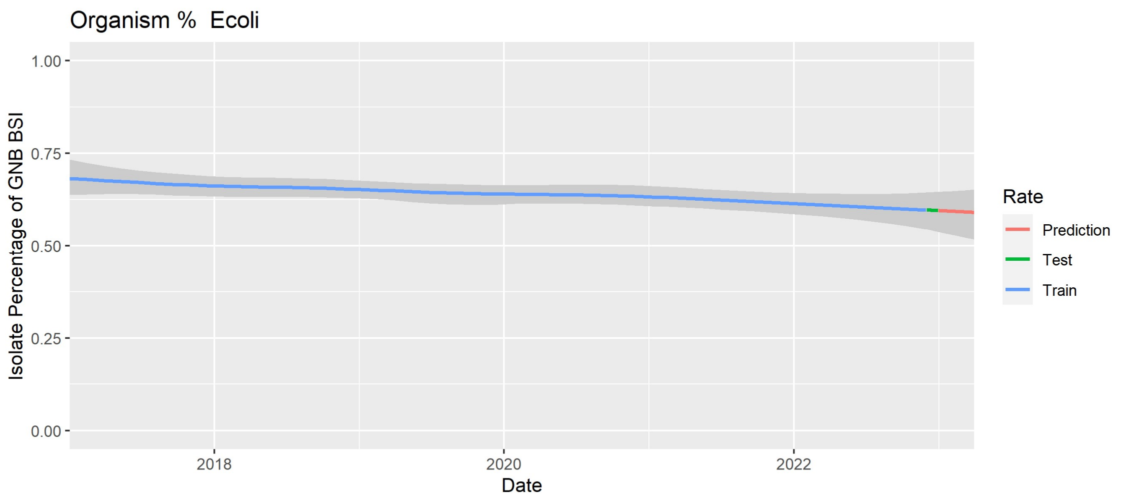


[1] "2017-01-01 Mid 0.682 (95%CI 0.637 to 0.732) Estimated Error 0.0236"

[1] "2022-12-01 Mid 0.596 (95%CI 0.544 to 0.644) Estimated Error 0.0254"

[1] "2022-12-30 Mid 0.595 (95%CI 0.537 to 0.645) Estimated Error 0.0272"

99.4% Posterior Probability of decrease from 2017 to 2023

Patients Over 80 years  E. coli

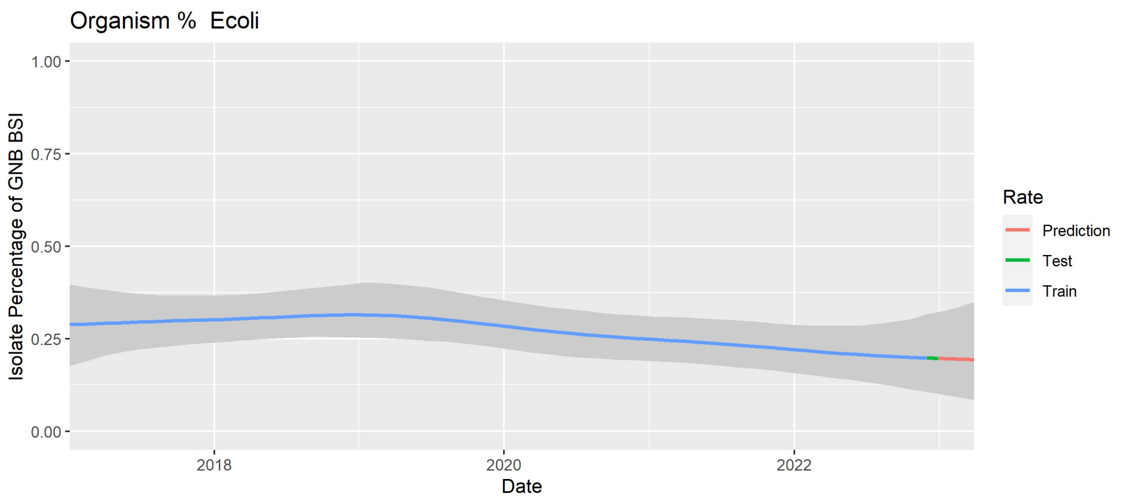


[1] "2017-01-02 Mid 0.289 (95%CI 0.178 to 0.396) Estimated Error 0.0560"

[1] "2022-11-30 Mid 0.198 (95%CI 0.105 to 0.316) Estimated Error 0.0525"

[1] "2022-12-29 Mid 0.197 (95%CI 0.101 to 0.322) Estimated Error 0.0558"

88.5% x greater than y

Patients Under 18 years   E. coli

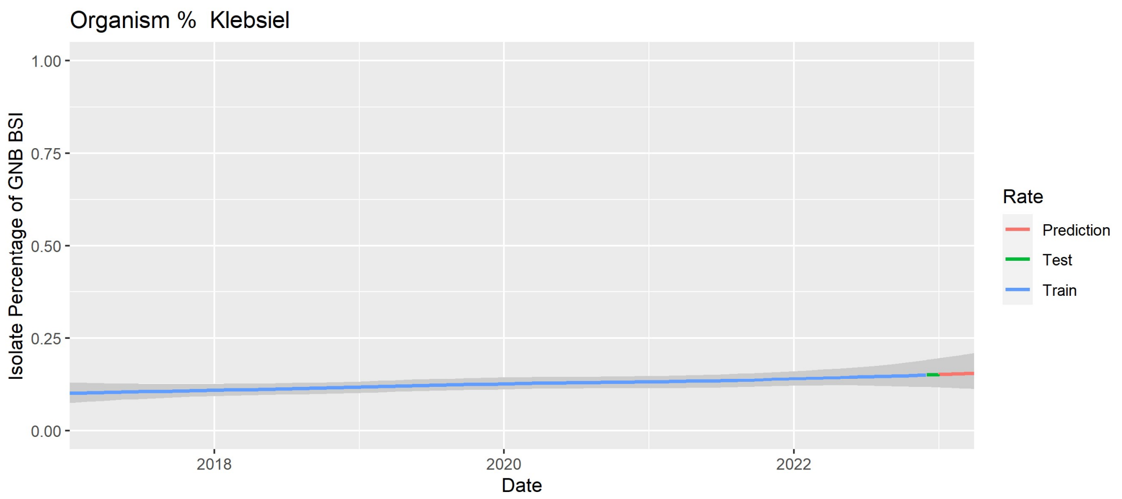


[1] "2017-01-01 Mid 0.101 (95%CI 0.075 to 0.130) Estimated Error 0.0139"

[1] "2022-12-01 Mid 0.151 (95%CI 0.117 to 0.192) Estimated Error 0.0185"

[1] "2022-12-30 Mid 0.152 (95%CI 0.116 to 0.196) Estimated Error 0.0198"

1.5% Posterior Probability of decrease from 2017 to 2023

Patients Over 80 years  Klebsiells sp.

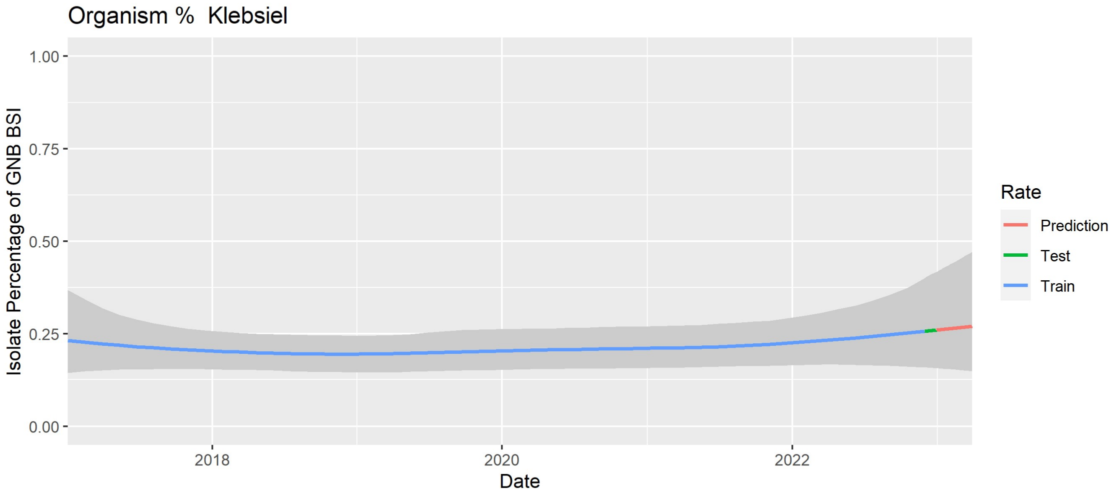


[1] "2017-01-02 Mid 0.232 (95%CI 0.145 to 0.369) Estimated Error 0.0561"

[1] "2022-11-30 Mid 0.257 (95%CI 0.159 to 0.400) Estimated Error 0.0606"

[1] "2022-12-29 Mid 0.260 (95%CI 0.157 to 0.416) Estimated Error 0.0652"

37.0% Posterior Probability of decrease from 2017 to 2023

Patients Under 18 years    Klebsiells sp.

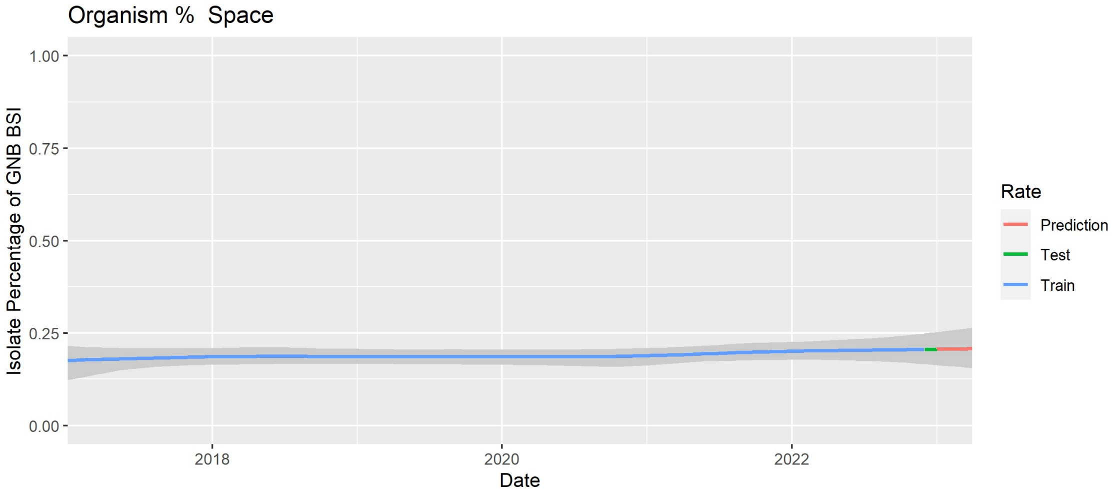


[1] "2017-01-01 Mid 0.177 (95%CI 0.123 to 0.217) Estimated Error 0.0217"

[1] "2022-12-01 Mid 0.206 (95%CI 0.167 to 0.250) Estimated Error 0.0207"

[1] "2022-12-30 Mid 0.207 (95%CI 0.164 to 0.252) Estimated Error 0.0221"

16.3% Posterior Probability of decrease from 2017 to 2023

Patients Over 80 years  SPACE Organisms

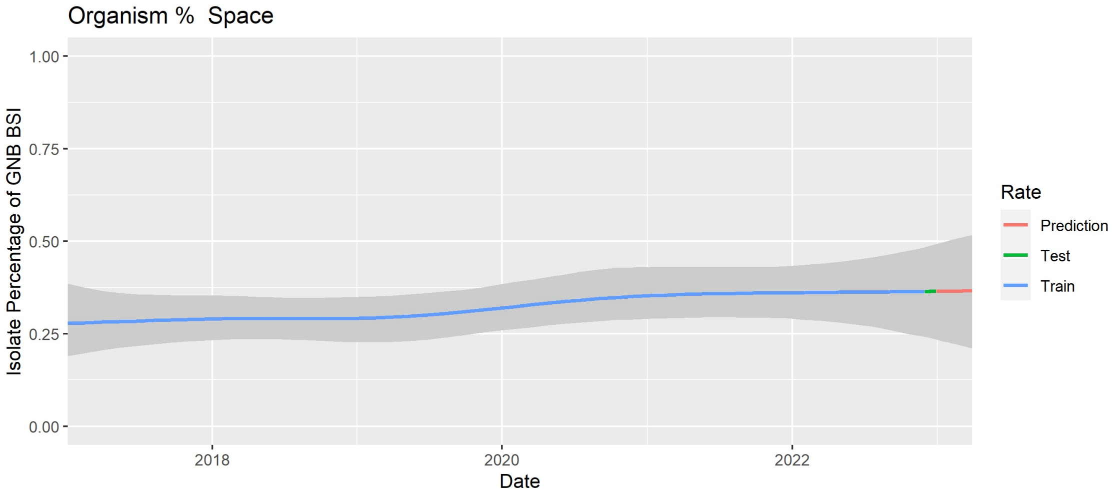


[1] "2017-01-02 Mid 0.279 (95%CI 0.191 to 0.385) Estimated Error 0.0492"

[1] "2022-11-30 Mid 0.364 (95%CI 0.242 to 0.484) Estimated Error 0.0618"

[1] "2022-12-29 Mid 0.365 (95%CI 0.235 to 0.492) Estimated Error 0.0654"

14.4% Posterior Probability of decrease from 2017 to 2023

Patients Under 18 years   SPACE Organisms
