## Supplementary - AMR Ecoli and Non E coli for "Rational Empiric Antibiotic Escalation Applied to Specific Patient Groups" (from another version of this preprint: version 2, 2023-11-07)

Supplementary Material (AMR Percentages)

Antibiotic resistance over time, with the probability of decreasing resistance between the start and end points (All patients, for E. coli and Non-E. coli isolates)

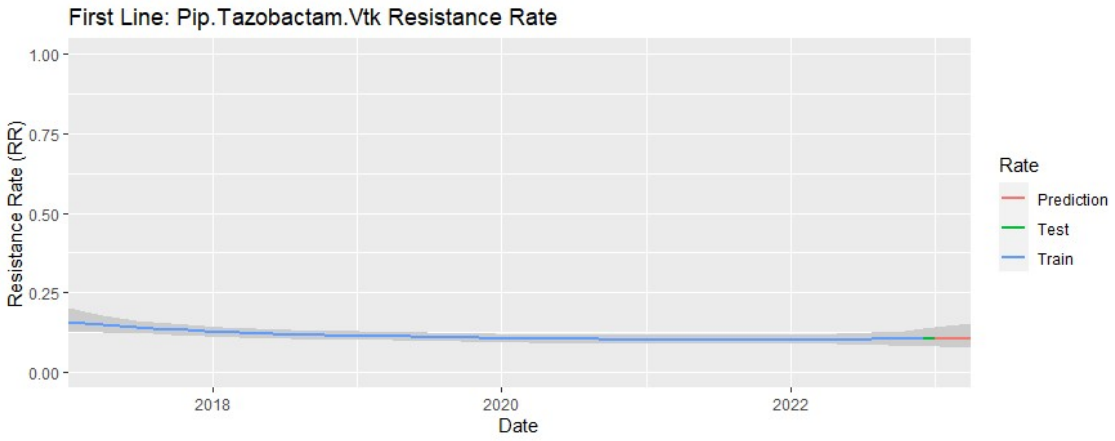


[1] "2017-01-01 Mid 0.157 (95%CI 0.125 to 0.200) Estimated Error 0.0196"

[1] "2022-12-03 Mid 0.104 (95%CI 0.080 to 0.135) Estimated Error 0.0142"

[1] "2022-12-31 Mid 0.104 (95%CI 0.078 to 0.139) Estimated Error 0.0154"

"99.4% posterior probability of decrease "

E. coli

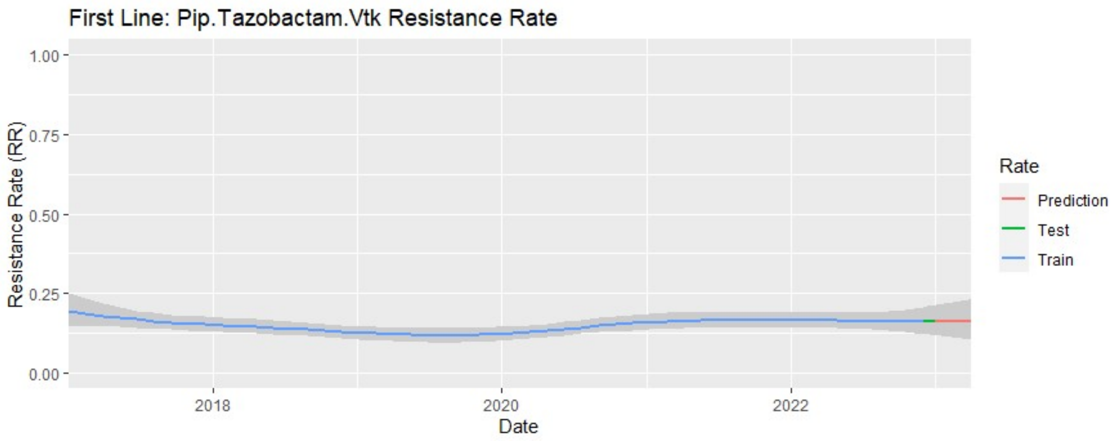


[1] "2017-01-01 Mid 0.193 (95%CI 0.143 to 0.251) Estimated Error 0.0271"

[1] "2022-12-02 Mid 0.162 (95%CI 0.121 to 0.206) Estimated Error 0.0218"

[1] "2022-12-31 Mid 0.162 (95%CI 0.117 to 0.211) Estimated Error 0.0241"

"80.4% posterior probability of decrease "

Non-Ecoli

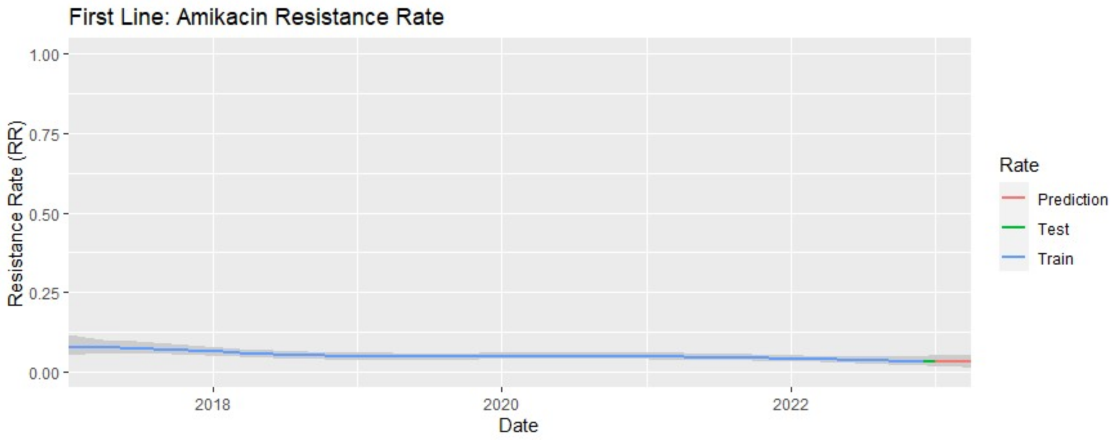


[1] "2017-01-02 Mid 0.079 (95%CI 0.052 to 0.115) Estimated Error 0.0160"

[1] "2022-12-03 Mid 0.032 (95%CI 0.017 to 0.049) Estimated Error 0.0081"

[1] "2022-12-31 Mid 0.031 (95%CI 0.016 to 0.050) Estimated Error 0.0086"

"99.9% posterior probability of decrease "

E. coli

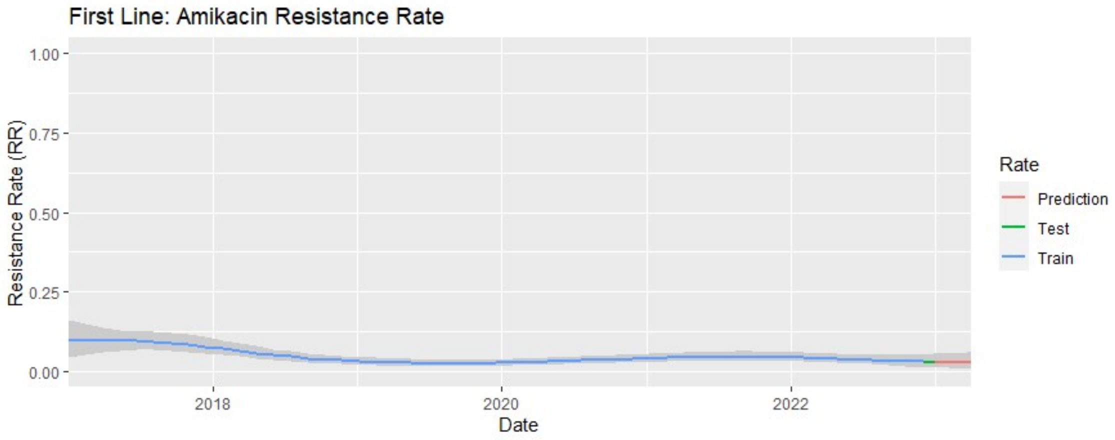


[1] "2017-01-01 Mid 0.096 (95%CI 0.045 to 0.160) Estimated Error 0.0294"

[1] "2022-12-03 Mid 0.029 (95%CI 0.012 to 0.052) Estimated Error 0.0103"

[1] "2022-12-31 Mid 0.028 (95%CI 0.011 to 0.053) Estimated Error 0.0111"

"99.3% posterior probability of decrease "

Non-Ecoli

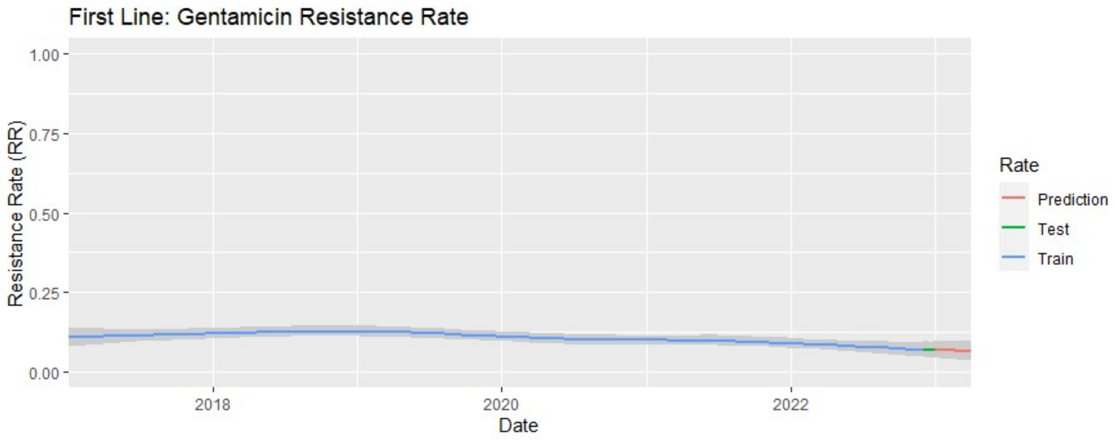


[1] "2017-01-01 Mid 0.107 (95%CI 0.079 to 0.138) Estimated Error 0.0156"

[1] "2022-12-03 Mid 0.069 (95%CI 0.046 to 0.094) Estimated Error 0.0123"

[1] "2022-12-31 Mid 0.068 (95%CI 0.043 to 0.094) Estimated Error 0.0131"

"97.9% posterior probability of decrease "

E. coli

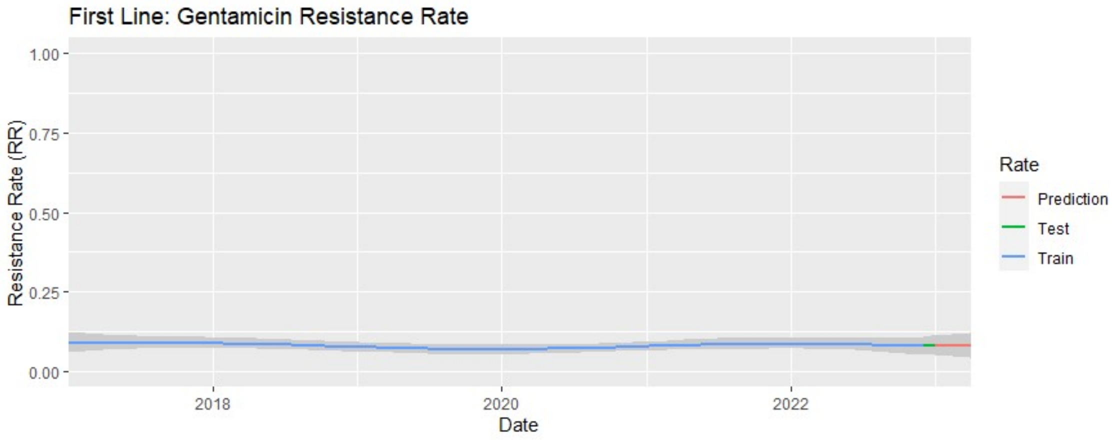


[1] "2017-01-01 Mid 0.090 (95%CI 0.060 to 0.126) Estimated Error 0.0164"

[1] "2022-12-02 Mid 0.080 (95%CI 0.052 to 0.109) Estimated Error 0.0143"

[1] "2022-12-31 Mid 0.079 (95%CI 0.050 to 0.111) Estimated Error 0.0155"

"69.5% posterior probability of decrease "

Non-Ecoli

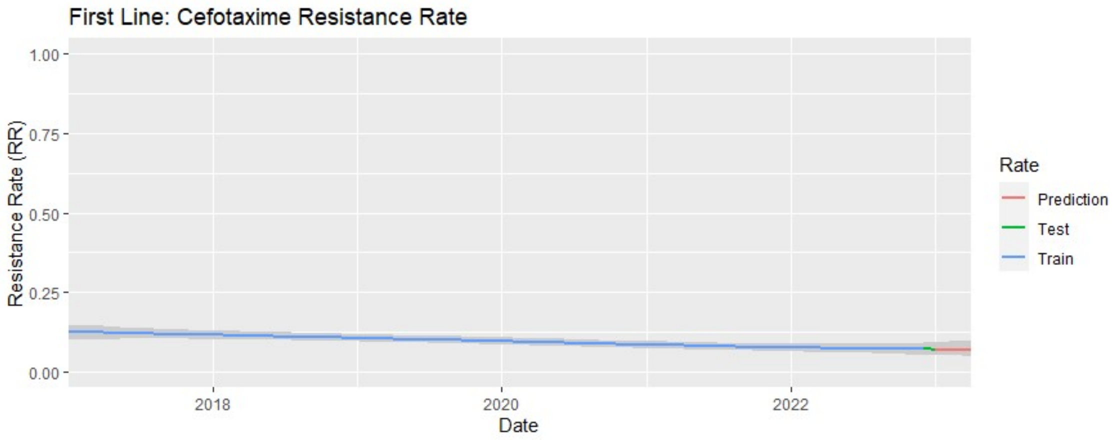


[1] "2017-01-01 Mid 0.125 (95%CI 0.099 to 0.151) Estimated Error 0.0130"

[1] "2022-12-03 Mid 0.070 (95%CI 0.053 to 0.091) Estimated Error 0.0095"

[1] "2022-12-31 Mid 0.070 (95%CI 0.052 to 0.092) Estimated Error 0.0100"

"99.9% posterior probability of decrease "

E. coli

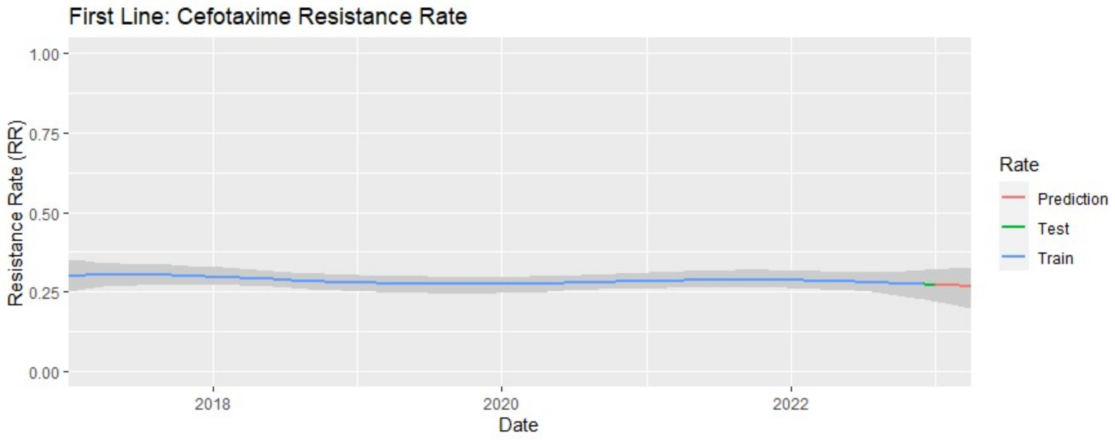


[1] "2017-01-01 Mid 0.301 (95%CI 0.249 to 0.352) Estimated Error 0.0257"

[1] "2022-12-02 Mid 0.273 (95%CI 0.225 to 0.317) Estimated Error 0.0231"

[1] "2022-12-31 Mid 0.272 (95%CI 0.218 to 0.319) Estimated Error 0.0252"

"80.2% posterior probability of decrease "

Non-Ecoli

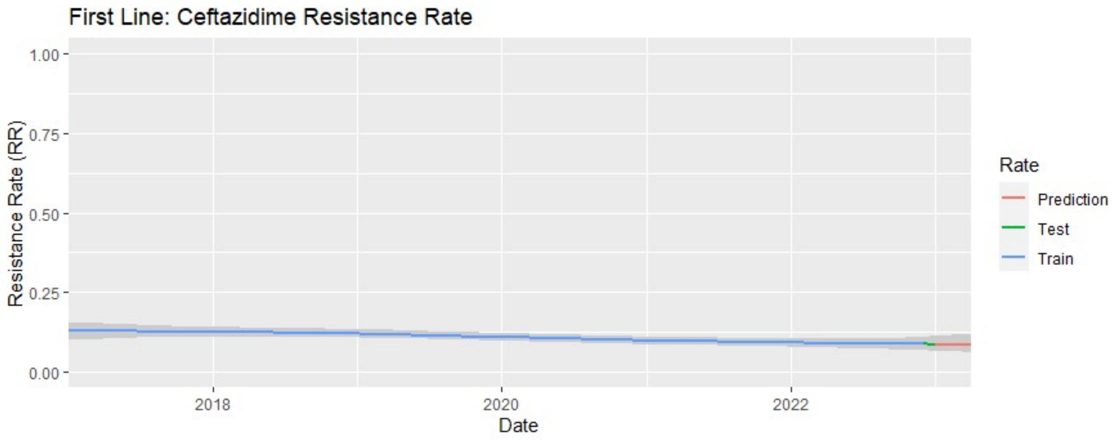


[1] "2017-01-01 Mid 0.129 (95%CI 0.099 to 0.158) Estimated Error 0.0147"

[1] "2022-12-03 Mid 0.086 (95%CI 0.066 to 0.109) Estimated Error 0.0108"

[1] "2022-12-31 Mid 0.086 (95%CI 0.065 to 0.111) Estimated Error 0.0115"

"98.5% posterior probability of decrease "

E. coli

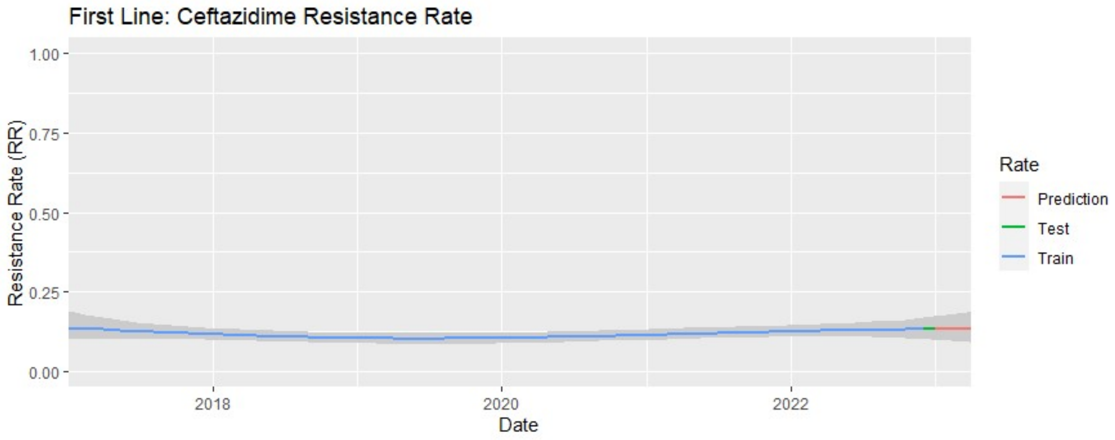


[1] "2017-01-01 Mid 0.137 (95%CI 0.100 to 0.188) Estimated Error 0.0228"

[1] "2022-12-02 Mid 0.132 (95%CI 0.100 to 0.168) Estimated Error 0.0174"

[1] "2022-12-31 Mid 0.133 (95%CI 0.098 to 0.172) Estimated Error 0.0189"

"54.2% posterior probability of decrease "

Non-Ecoli

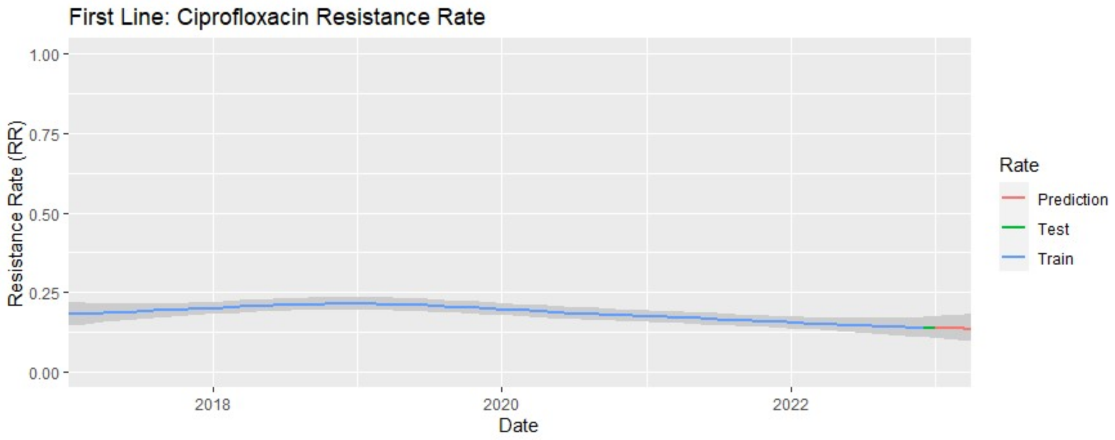


[1] "2017-01-01 Mid 0.180 (95%CI 0.143 to 0.218) Estimated Error 0.0192"

[1] "2022-12-03 Mid 0.139 (95%CI 0.108 to 0.172) Estimated Error 0.0163"

[1] "2022-12-31 Mid 0.137 (95%CI 0.105 to 0.174) Estimated Error 0.0175"

"95.0% posterior probability of decrease "

E. coli

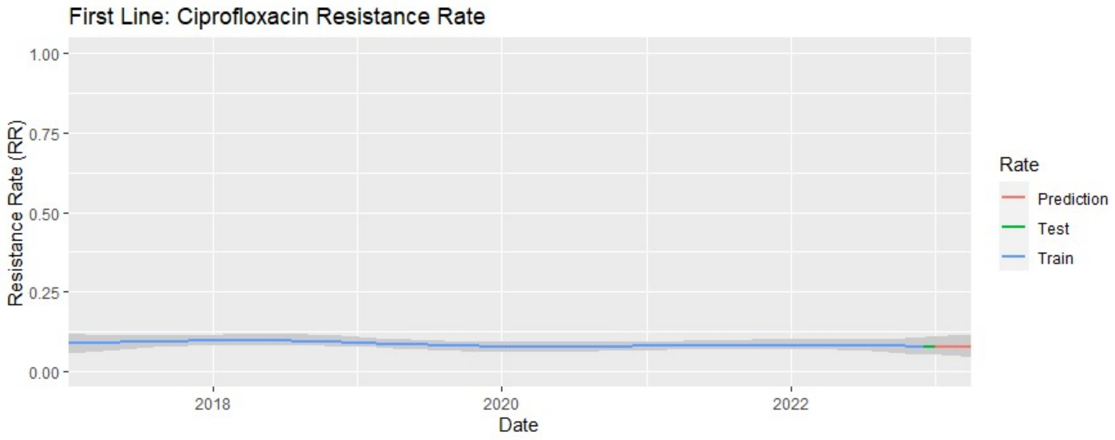


[1] "2017-01-01 Mid 0.088 (95%CI 0.055 to 0.118) Estimated Error 0.0159"

[1] "2022-12-02 Mid 0.078 (95%CI 0.053 to 0.106) Estimated Error 0.0129"

[1] "2022-12-31 Mid 0.077 (95%CI 0.051 to 0.108) Estimated Error 0.0140"

"70.5% posterior probability of decrease "

Non-Ecoli

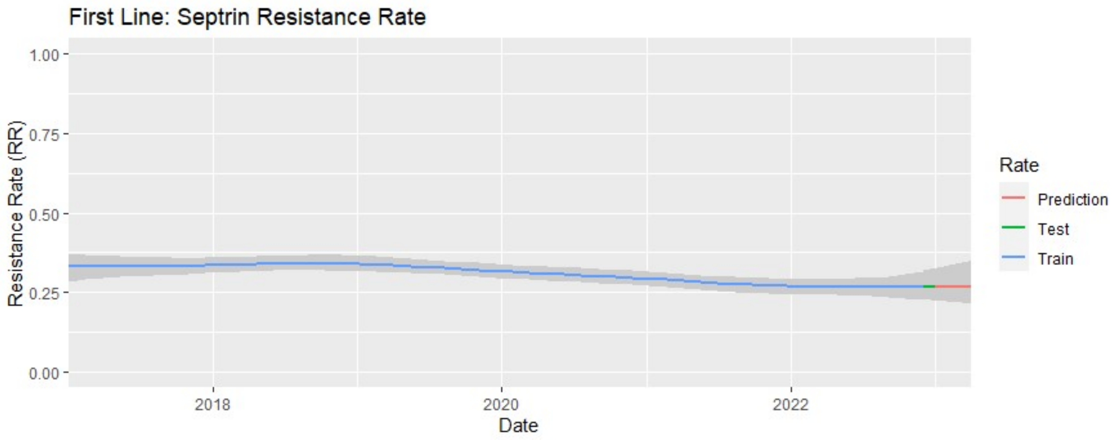


[1] "2017-01-01 Mid 0.330 (95%CI 0.283 to 0.373) Estimated Error 0.0229"

[1] "2022-12-03 Mid 0.267 (95%CI 0.226 to 0.318) Estimated Error 0.0227"

[1] "2022-12-31 Mid 0.268 (95%CI 0.223 to 0.324) Estimated Error 0.0249"

"96.0% posterior probability of decrease "

E. coli

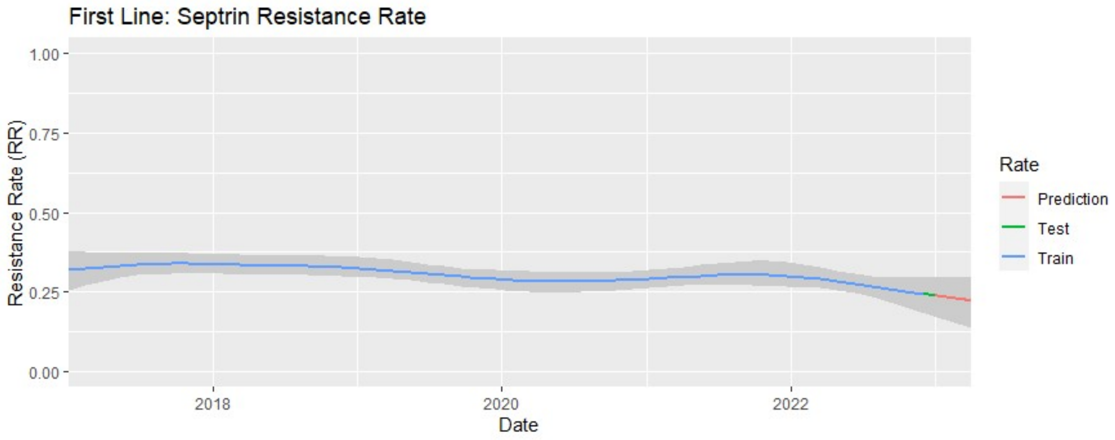


[1] "2017-01-01 Mid 0.321 (95%CI 0.254 to 0.380) Estimated Error 0.0323"

[1] "2022-12-02 Mid 0.242 (95%CI 0.182 to 0.295) Estimated Error 0.0297"

[1] "2022-12-31 Mid 0.237 (95%CI 0.170 to 0.294) Estimated Error 0.0331"

"98.4% posterior probability of decrease "
Non-Ecoli
