## Supplementary - Under18 and Over 80 for "Rational Empiric Antibiotic Escalation Applied to Specific Patient Groups" (from another version of this preprint: version 2, 2023-11-07)

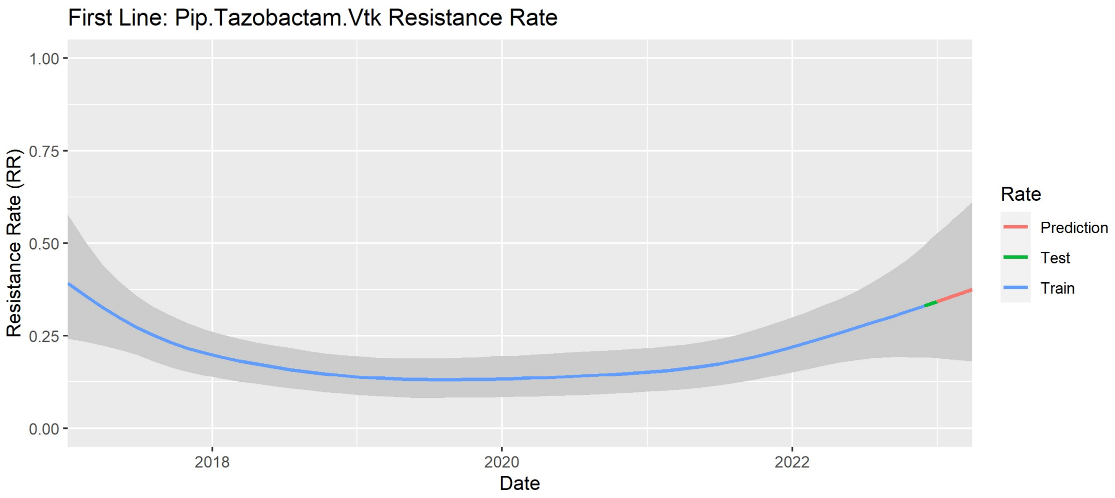


[1] "2017-01-02 Mid 0.392 (95%CI 0.243 to 0.577) Estimated Error 0.0853"

[1] "2022-11-30 Mid 0.331 (95%CI 0.192 to 0.495) Estimated Error 0.0786"

[1] "2022-12-29 Mid 0.342 (95%CI 0.190 to 0.524) Estimated Error 0.0862"

"70.6% posterior probability of decrease "

Under 18

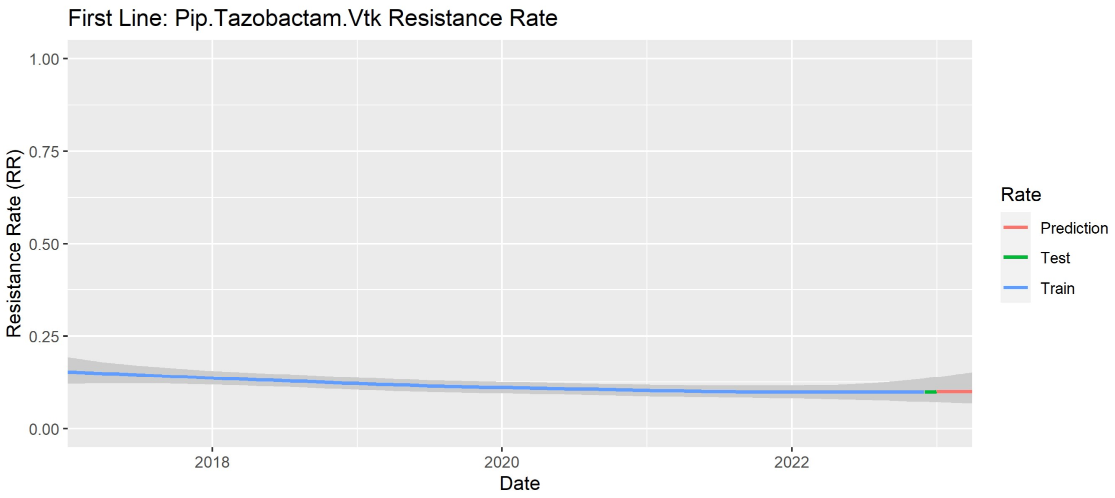


[1] "2017-01-01 Mid 0.154 (95%CI 0.121 to 0.194) Estimated Error 0.0183"

[1] "2022-12-01 Mid 0.099 (95%CI 0.072 to 0.136) Estimated Error 0.0162"

[1] "2022-12-30 Mid 0.099 (95%CI 0.071 to 0.139) Estimated Error 0.0174"

"98.5% posterior probability of decrease "

Over 80

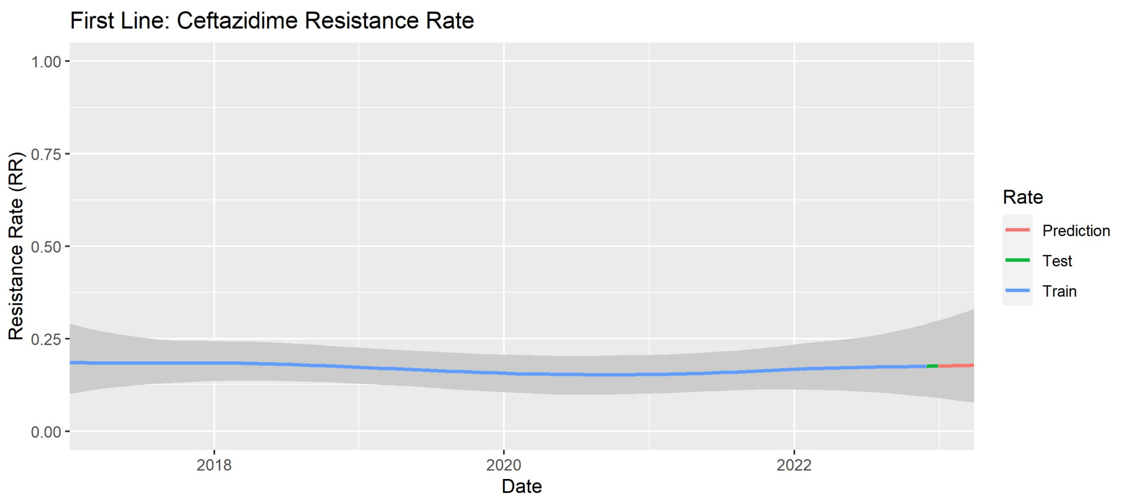


[1] "2017-01-02 Mid 0.186 (95%CI 0.101 to 0.291) Estimated Error 0.0476"

[1] "2022-11-30 Mid 0.176 (95%CI 0.093 to 0.291) Estimated Error 0.0500"

[1] "2022-12-29 Mid 0.176 (95%CI 0.089 to 0.299) Estimated Error 0.0530"

"56.6% posterior probability of decrease "

Under 18

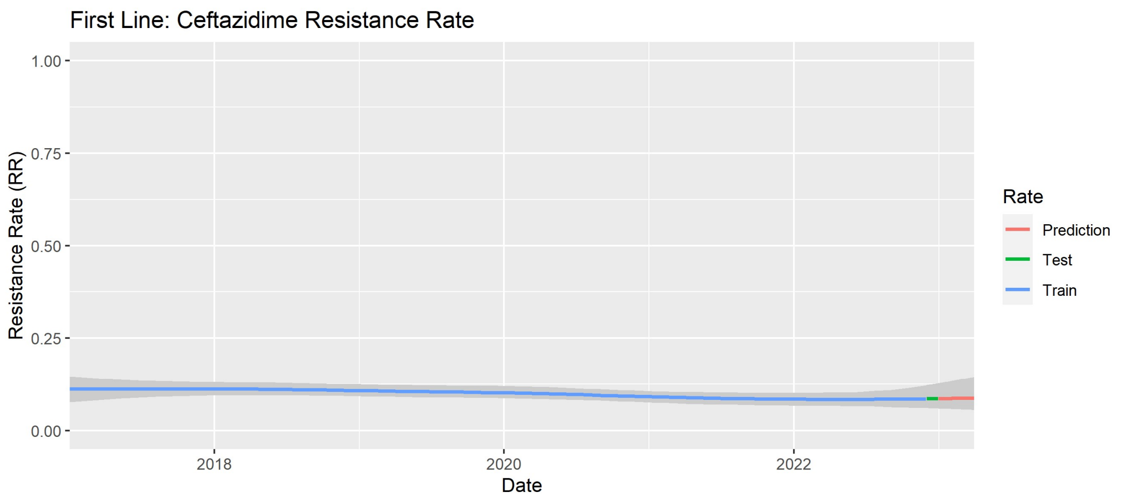


[1] "2017-01-01 Mid 0.112 (95%CI 0.076 to 0.147) Estimated Error 0.0180"

[1] "2022-12-01 Mid 0.086 (95%CI 0.060 to 0.123) Estimated Error 0.0155"

[1] "2022-12-30 Mid 0.086 (95%CI 0.059 to 0.128) Estimated Error 0.0170"

"86.1% posterior probability of decrease "

Over 80

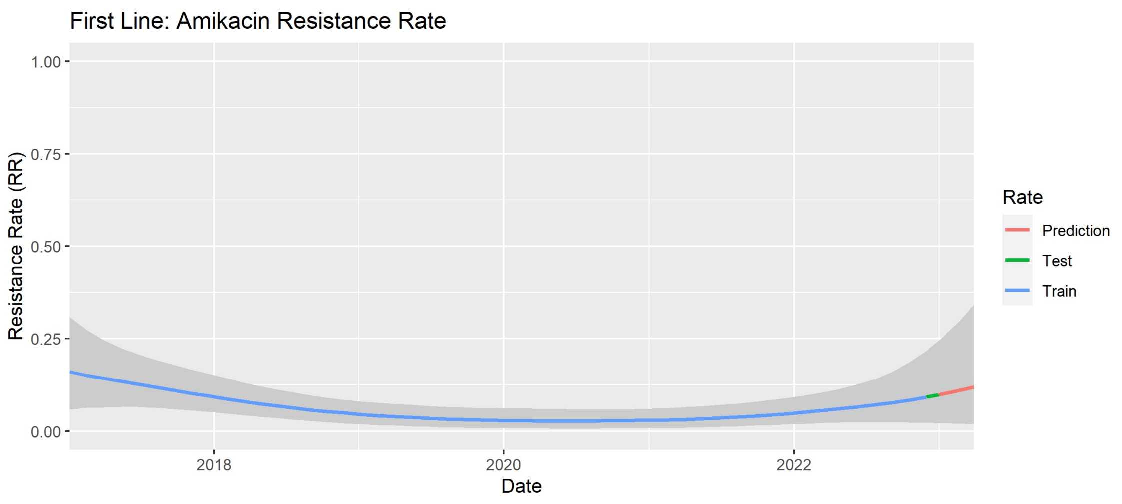


[1] "2017-01-02 Mid 0.160 (95%CI 0.059 to 0.308) Estimated Error 0.0647"

[1] "2022-11-30 Mid 0.092 (95%CI 0.022 to 0.217) Estimated Error 0.0515"

[1] "2022-12-29 Mid 0.098 (95%CI 0.021 to 0.242) Estimated Error 0.0585"

"81.7% posterior probability of decrease "

Under 18

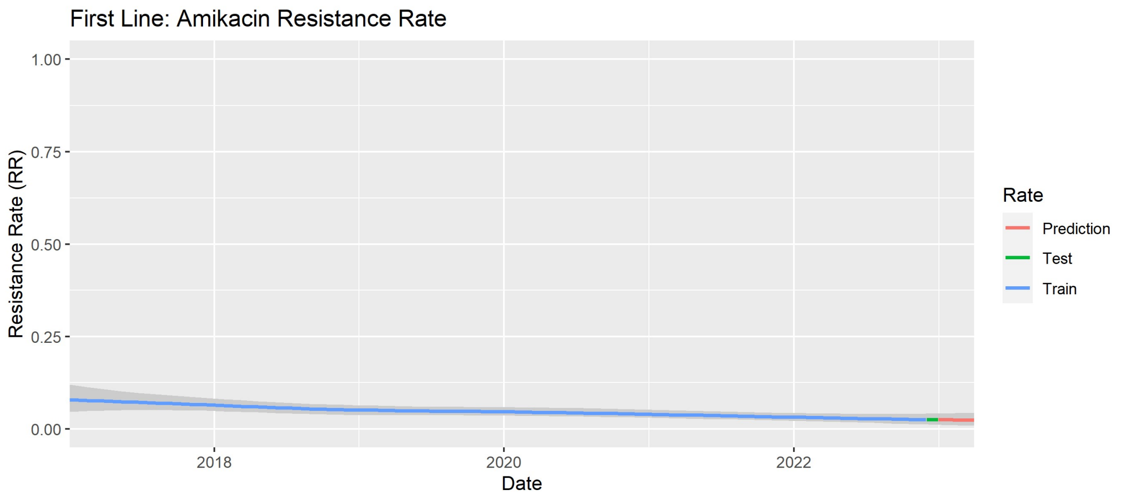


[1] "2017-01-02 Mid 0.079 (95%CI 0.046 to 0.120) Estimated Error 0.0185"

[1] "2022-12-01 Mid 0.025 (95%CI 0.012 to 0.041) Estimated Error 0.0074"

[1] "2022-12-30 Mid 0.025 (95%CI 0.011 to 0.041) Estimated Error 0.0077"

"99.9% posterior probability of decrease "

Over 80

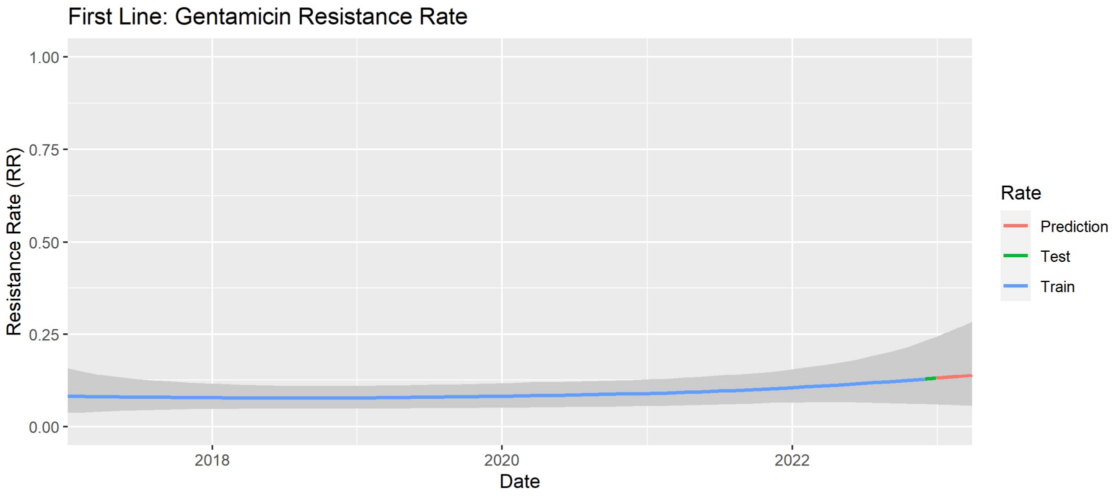


[1] "2017-01-02 Mid 0.082 (95%CI 0.037 to 0.158) Estimated Error 0.0307"

[1] "2022-11-30 Mid 0.128 (95%CI 0.060 to 0.232) Estimated Error 0.0441"

[1] "2022-12-29 Mid 0.130 (95%CI 0.059 to 0.243) Estimated Error 0.0473"

"18.2% posterior probability of decrease "

Under 18

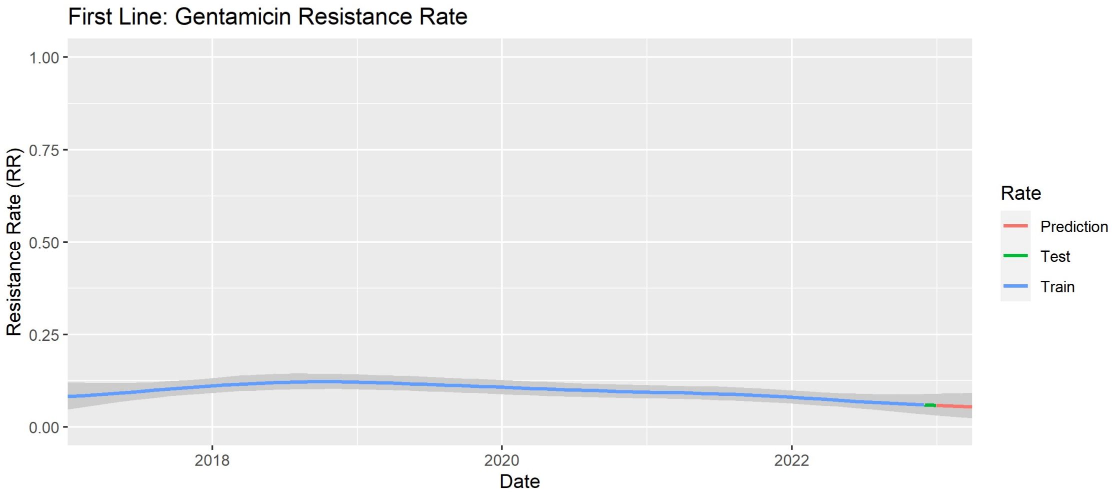


[1] "2017-01-01 Mid 0.082 (95%CI 0.047 to 0.120) Estimated Error 0.0183"

[1] "2022-12-01 Mid 0.059 (95%CI 0.033 to 0.089) Estimated Error 0.0142"

[1] "2022-12-30 Mid 0.058 (95%CI 0.031 to 0.089) Estimated Error 0.0150"

"86.1% posterior probability of decrease "

Over 80

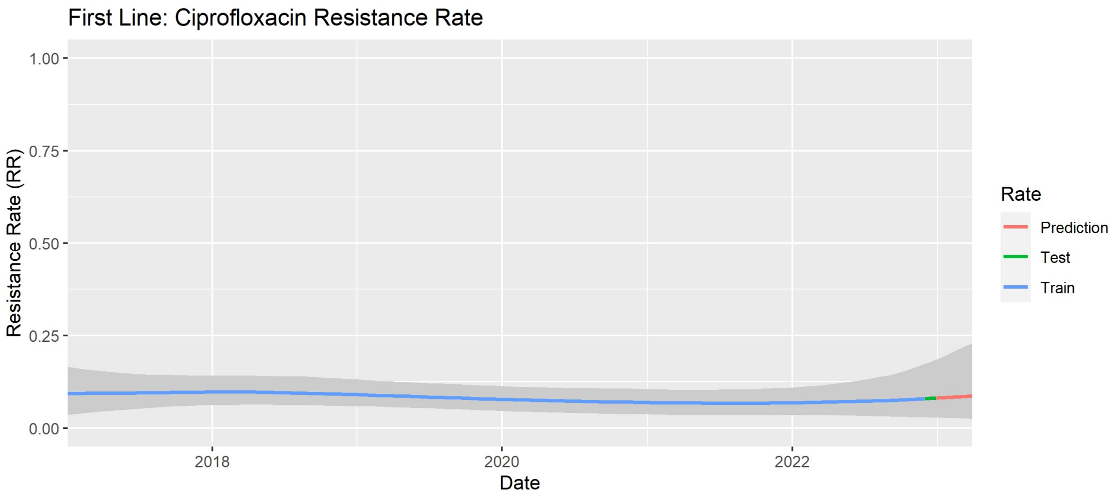


[1] "2017-01-02 Mid 0.093 (95%CI 0.036 to 0.166) Estimated Error 0.0330"

[1] "2022-11-30 Mid 0.079 (95%CI 0.029 to 0.174) Estimated Error 0.0368"

[1] "2022-12-29 Mid 0.081 (95%CI 0.028 to 0.185) Estimated Error 0.0400"

"64.2% posterior probability of decrease "

Under 18

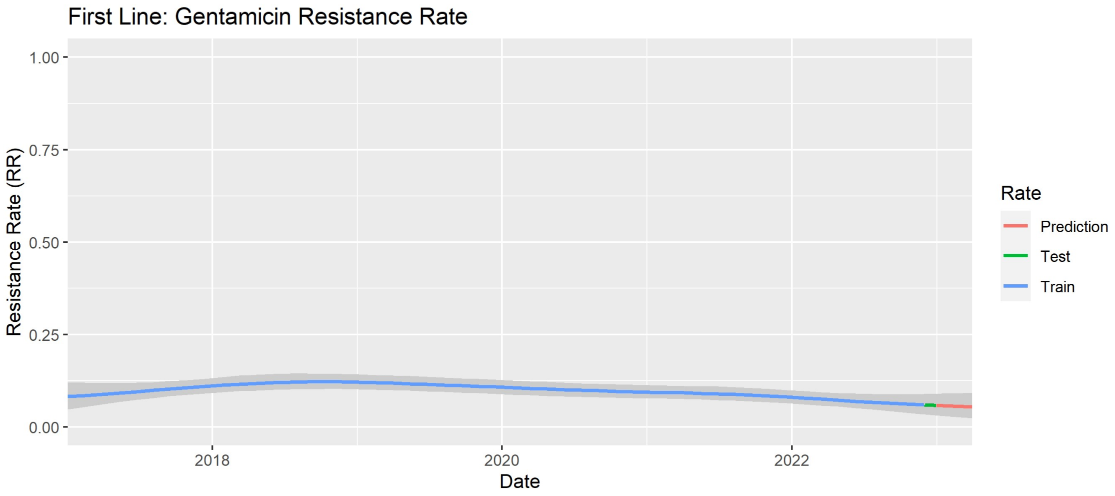


[1] "2017-01-01 Mid 0.141 (95%CI 0.098 to 0.186) Estimated Error 0.0230"

[1] "2022-12-01 Mid 0.111 (95%CI 0.077 to 0.149) Estimated Error 0.0182"

[1] "2022-12-30 Mid 0.110 (95%CI 0.074 to 0.150) Estimated Error 0.0194"

"85.5% posterior probability of decrease "

Over 80

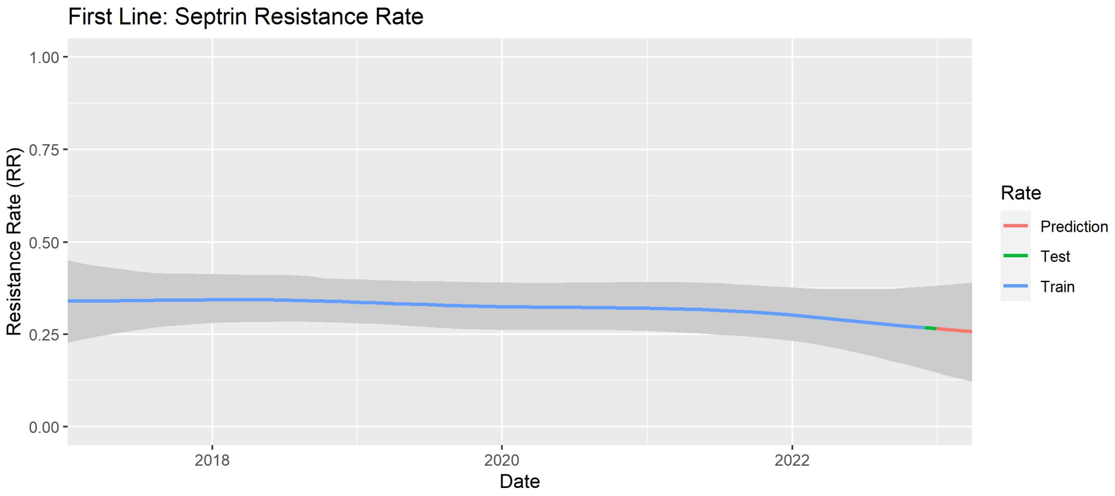


[1] "2017-01-01 Mid 0.336 (95%CI 0.273 to 0.395) Estimated Error 0.0318"

[1] "2022-12-01 Mid 0.293 (95%CI 0.239 to 0.359) Estimated Error 0.0310"

[1] "2022-12-30 Mid 0.294 (95%CI 0.235 to 0.366) Estimated Error 0.0340"

[1] "82.0% posterior probability of decrease "

Under 18

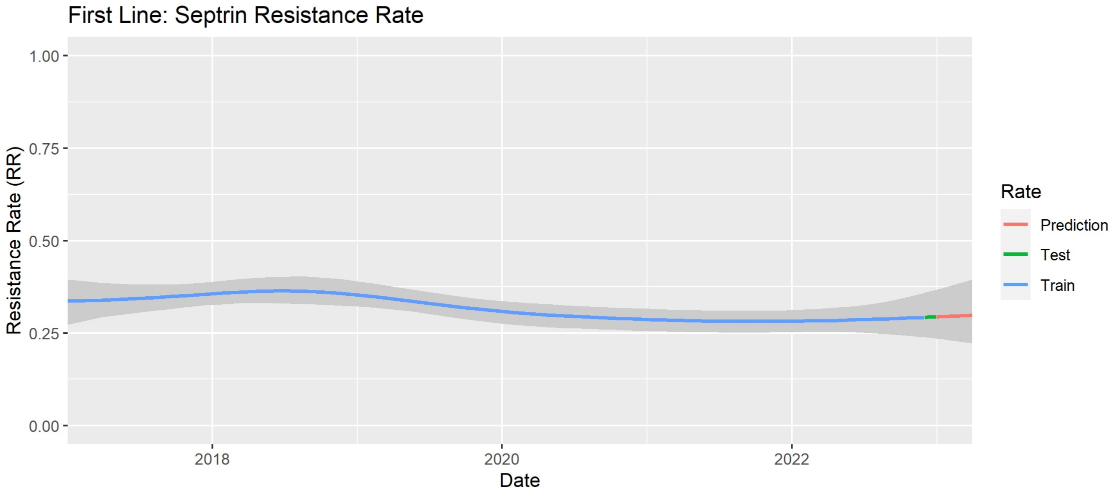


[1] "2017-01-02 Mid 0.339 (95%CI 0.228 to 0.450) Estimated Error 0.0560"

[1] "2022-11-30 Mid 0.268 (95%CI 0.155 to 0.379) Estimated Error 0.0565"

[1] "2022-12-29 Mid 0.265 (95%CI 0.147 to 0.381) Estimated Error 0.0592"

[1] "82.4% posterior probability of decrease "

Over 80

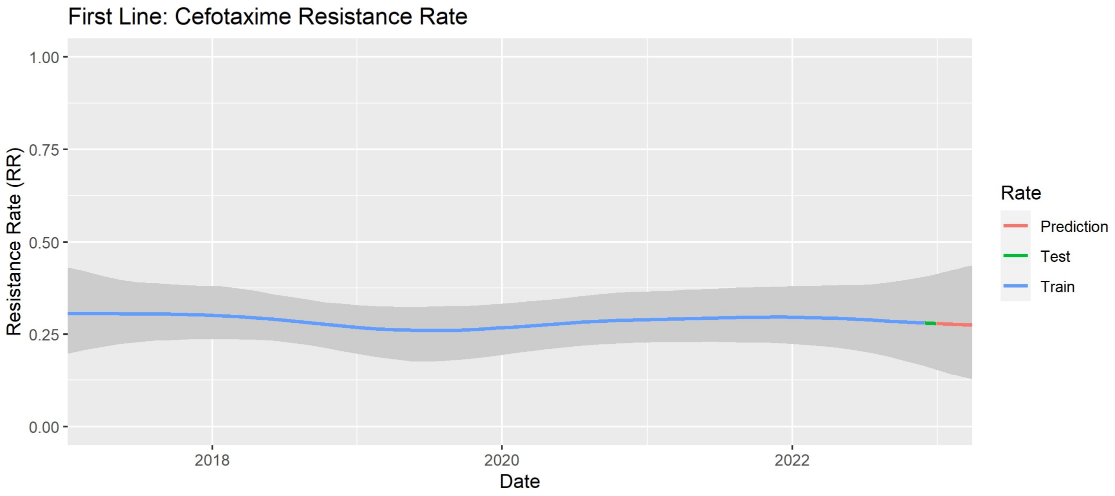


[1] "2017-01-02 Mid 0.307 (95%CI 0.198 to 0.431) Estimated Error 0.0585"

[1] "2022-11-30 Mid 0.281 (95%CI 0.164 to 0.406) Estimated Error 0.0610"

[1] "2022-12-29 Mid 0.279 (95%CI 0.154 to 0.412) Estimated Error 0.0648"

"62.3% posterior probability of decrease "

Under 18

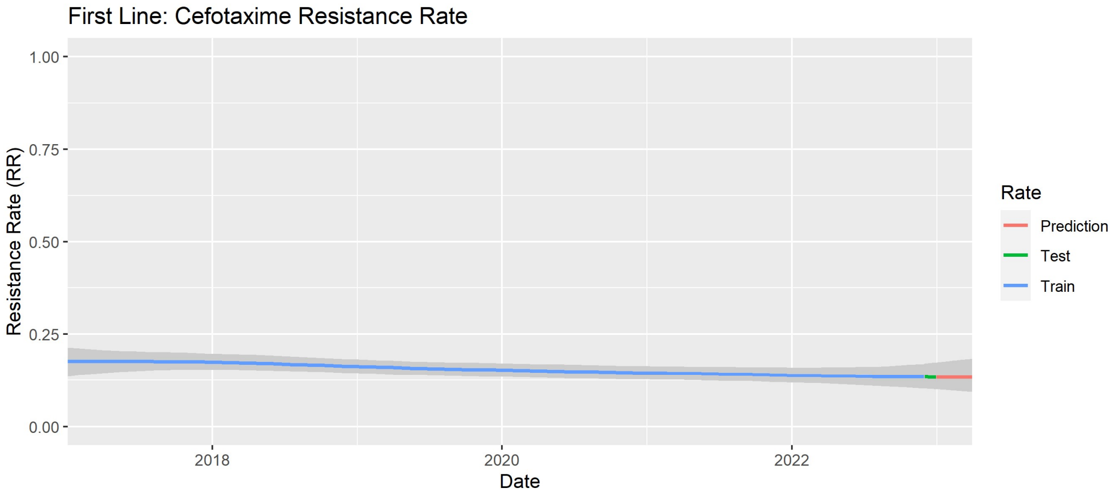


[1] "2017-01-01 Mid 0.176 (95%CI 0.136 to 0.214) Estimated Error 0.0198"

[1] "2022-12-01 Mid 0.134 (95%CI 0.103 to 0.171) Estimated Error 0.0170"

[1] "2022-12-30 Mid 0.134 (95%CI 0.101 to 0.174) Estimated Error 0.0181"

"93.8% posterior probability of decrease "

Over 80

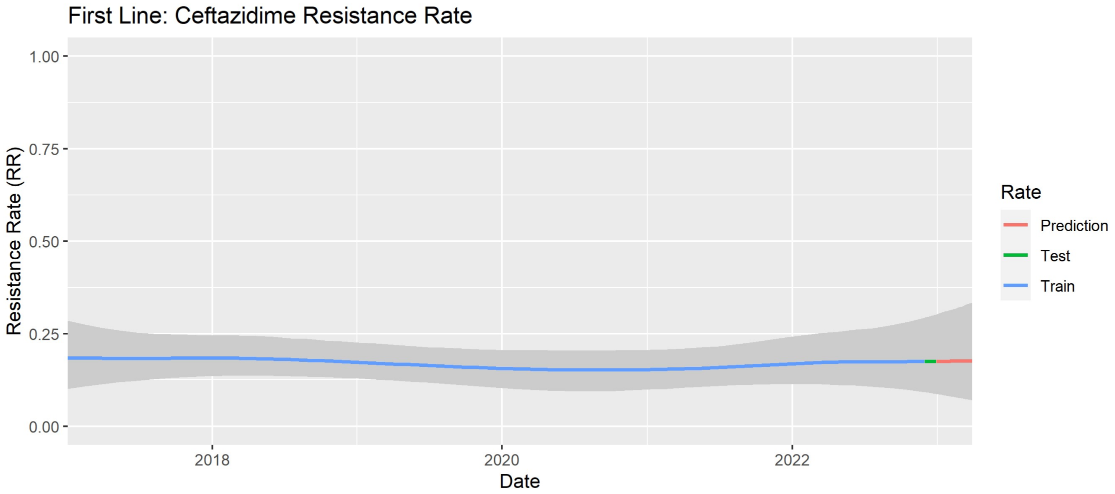


[1] "2017-01-02 Mid 0.185 (95%CI 0.100 to 0.286) Estimated Error 0.0470"

[1] "2022-11-30 Mid 0.175 (95%CI 0.091 to 0.293) Estimated Error 0.0509"

[1] "2022-12-29 Mid 0.176 (95%CI 0.086 to 0.302) Estimated Error 0.0541"

[1] "57.1% posterior probability of decrease "Under 18

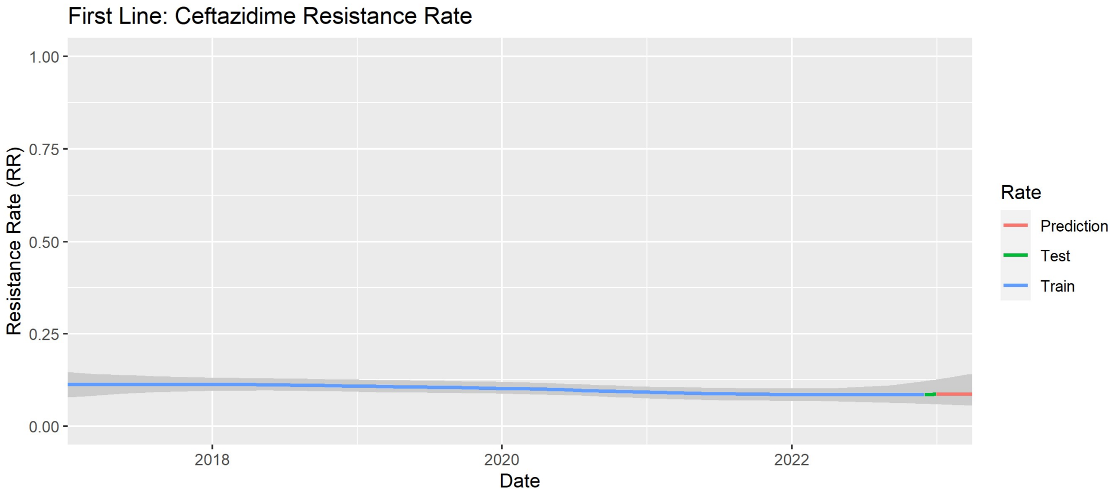


[1] "2017-01-01 Mid 0.113 (95%CI 0.077 to 0.147) Estimated Error 0.0177"

[1] "2022-12-01 Mid 0.085 (95%CI 0.060 to 0.121) Estimated Error 0.0152"

[1] "2022-12-30 Mid 0.085 (95%CI 0.059 to 0.125) Estimated Error 0.0165"

"87.2% posterior probability of decrease "

Over 80

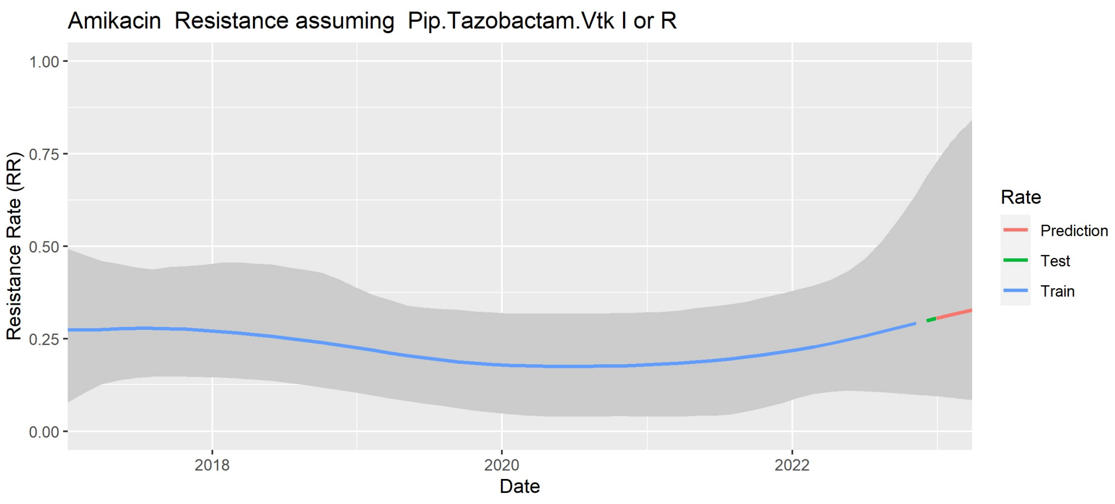


[1] "2017-01-02 Mid 0.274 (95%CI 0.078 to 0.493) Estimated Error 0.1026"

[1] "2022-12-06 Mid 0.299 (95%CI 0.096 to 0.690) Estimated Error 0.1466"

[1] "2022-12-29 Mid 0.305 (95%CI 0.094 to 0.726) Estimated Error 0.1553"

"49.6% posterior probability of decrease "

Under 18

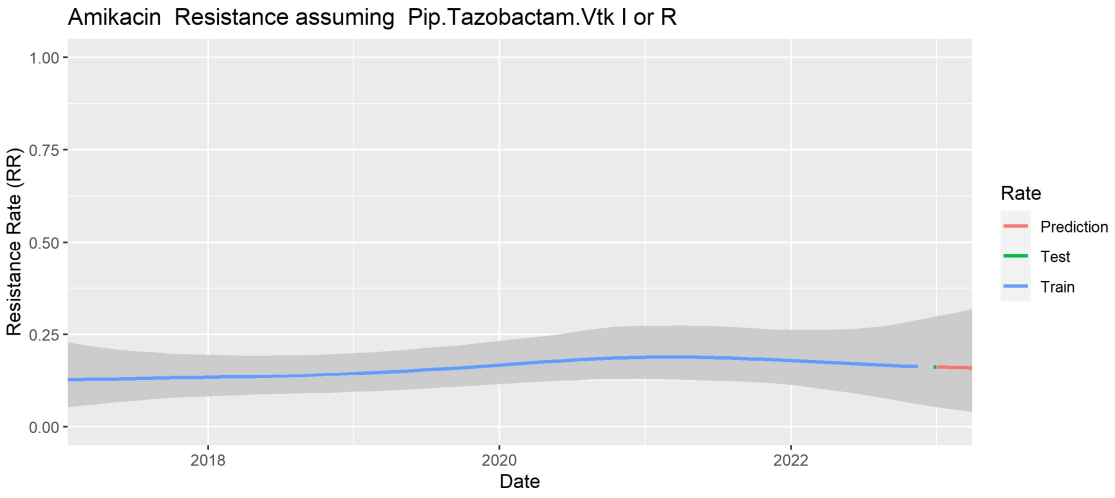


[1] "2017-01-14 Mid 0.127 (95%CI 0.052 to 0.230) Estimated Error 0.0450"

[1] "2022-12-24 Mid 0.162 (95%CI 0.054 to 0.298) Estimated Error 0.0629"

[1] "2022-12-30 Mid 0.162 (95%CI 0.053 to 0.299) Estimated Error 0.0635"

"32.5% posterior probability of decrease "

Over 80

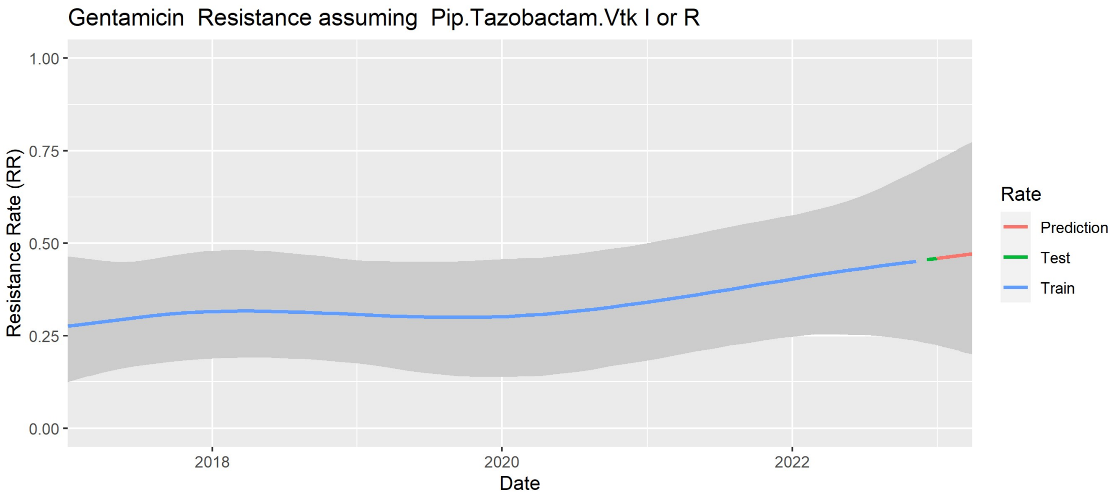


[1] "2017-01-02 Mid 0.277 (95%CI 0.124 to 0.465) Estimated Error 0.0875"

[1] "2022-12-06 Mid 0.455 (95%CI 0.230 to 0.711) Estimated Error 0.1235"

[1] "2022-12-29 Mid 0.459 (95%CI 0.225 to 0.723) Estimated Error 0.1283"

"12.6% posterior probability of decrease "

Under 18

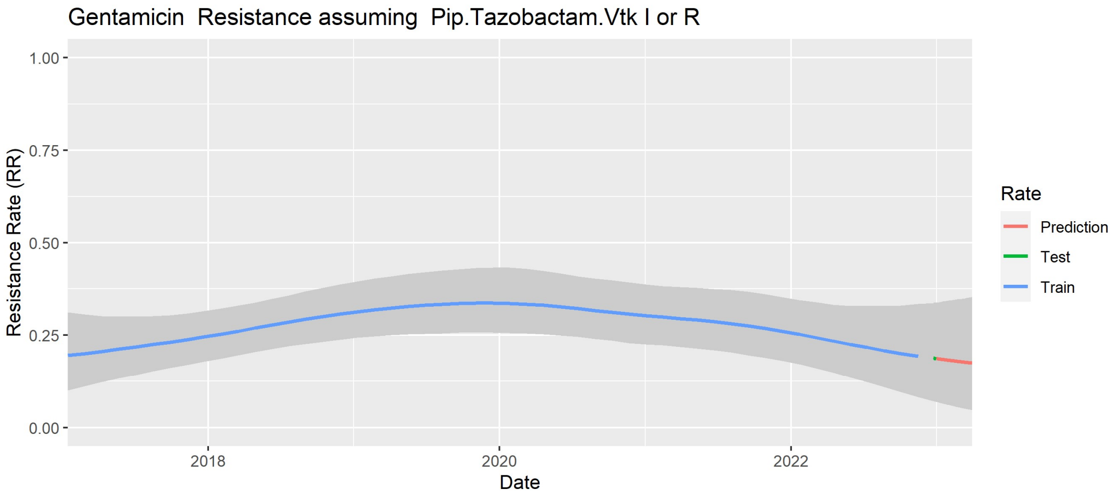


[1] "2017-01-14 Mid 0.196 (95%CI 0.101 to 0.311) Estimated Error 0.0544"

[1] "2022-12-24 Mid 0.188 (95%CI 0.071 to 0.337) Estimated Error 0.0700"

[1] "2022-12-30 Mid 0.187 (95%CI 0.069 to 0.338) Estimated Error 0.0706"

"53.8% posterior probability of decrease "

Over 80

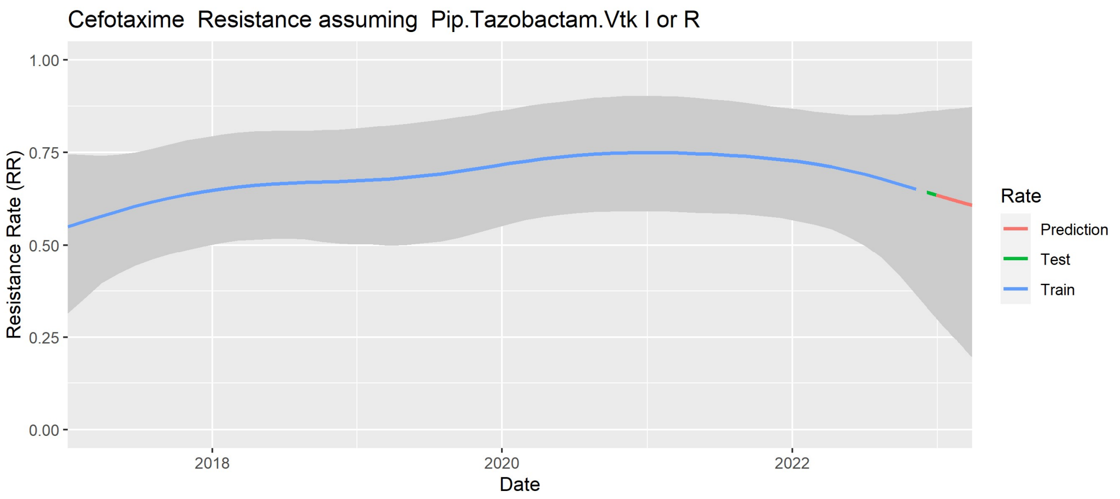


[1] "2017-01-02 Mid 0.550 (95%CI 0.315 to 0.745) Estimated Error 0.1097"

[1] "2022-12-06 Mid 0.642 (95%CI 0.330 to 0.861) Estimated Error 0.1364"

[1] "2022-12-29 Mid 0.635 (95%CI 0.302 to 0.863) Estimated Error 0.1449"

"25.8% posterior probability of decrease "

Under 18

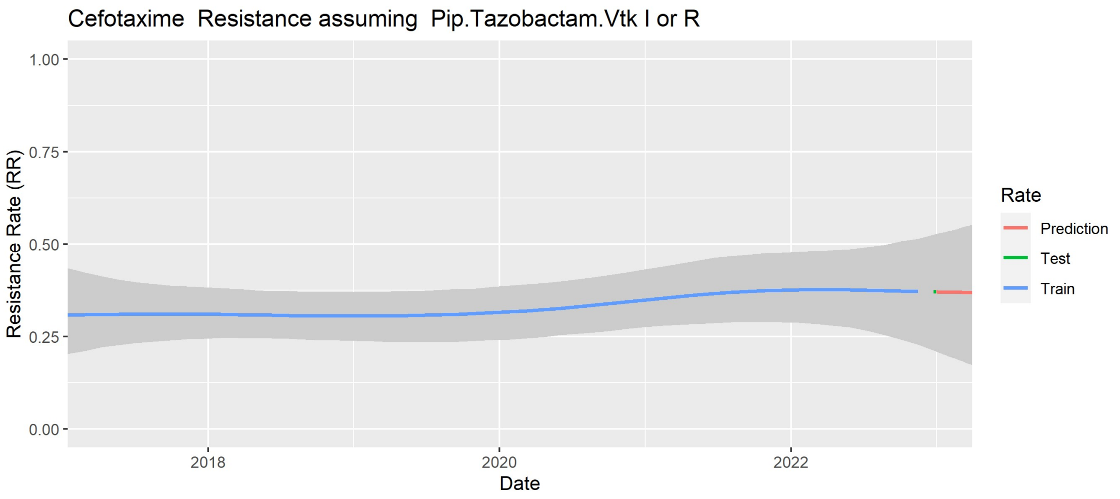


[1] "2017-01-14 Mid 0.309 (95%CI 0.204 to 0.434) Estimated Error 0.0583"

[1] "2022-12-24 Mid 0.370 (95%CI 0.213 to 0.526) Estimated Error 0.0782"

[1] "2022-12-30 Mid 0.370 (95%CI 0.210 to 0.527) Estimated Error 0.0791"

"28.0% posterior probability of decrease "

Over 80

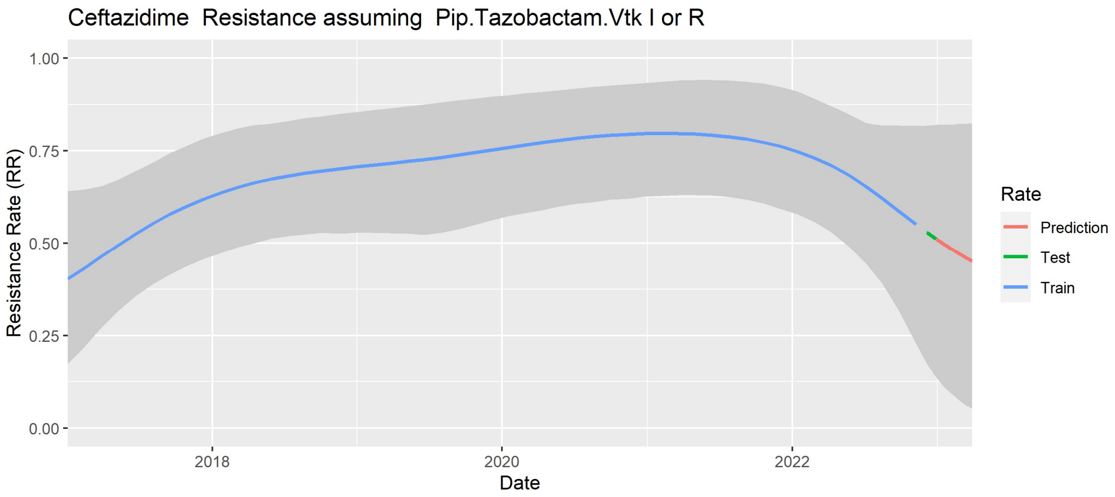


[1] "2017-01-02 Mid 0.404 (95%CI 0.175 to 0.641) Estimated Error 0.1219"

[1] "2022-12-06 Mid 0.529 (95%CI 0.174 to 0.817) Estimated Error 0.1687"

[1] "2022-12-29 Mid 0.511 (95%CI 0.138 to 0.819) Estimated Error 0.1795"

[1] "24.7% posterior probability of decrease "

Under 18

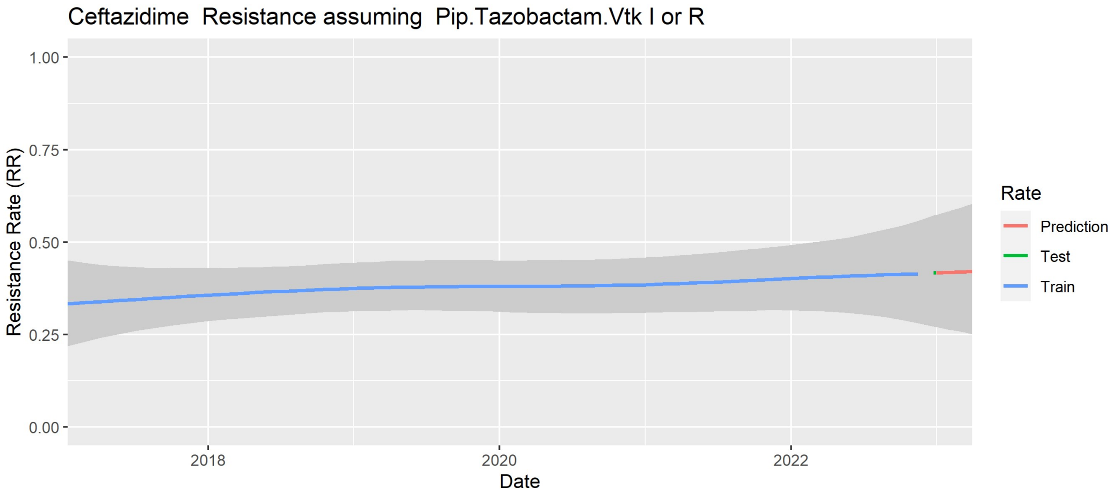


[1] "2017-01-14 Mid 0.333 (95%CI 0.219 to 0.450) Estimated Error 0.0585"

[1] "2022-12-24 Mid 0.416 (95%CI 0.271 to 0.572) Estimated Error 0.0749"

[1] "2022-12-30 Mid 0.416 (95%CI 0.270 to 0.574) Estimated Error 0.0757"

[1] "20.4% posterior probability of decrease "

Over 80

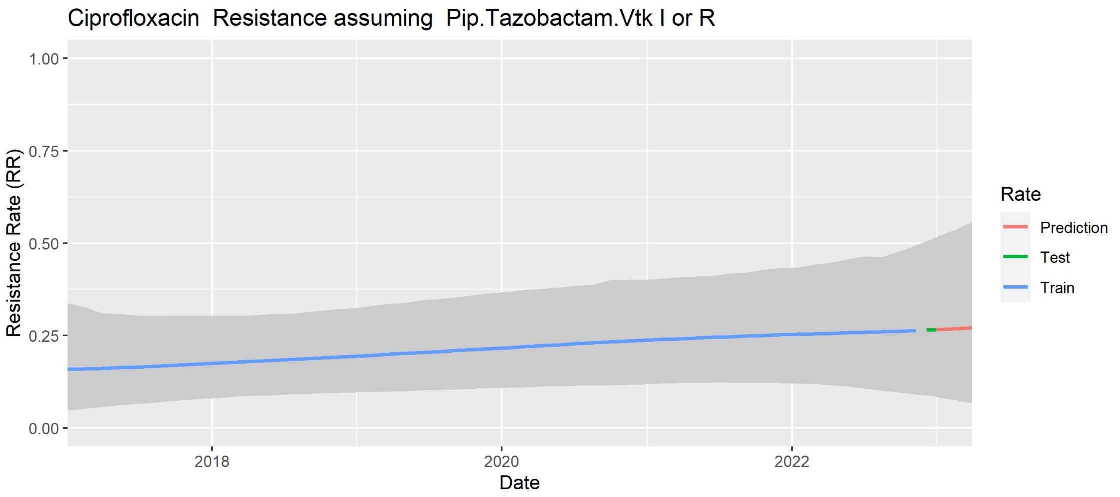


[1] "2017-01-02 Mid 0.160 (95%CI 0.048 to 0.338) Estimated Error 0.0750"

[1] "2022-12-06 Mid 0.265 (95%CI 0.087 to 0.505) Estimated Error 0.1091"

[1] "2022-12-29 Mid 0.266 (95%CI 0.084 to 0.515) Estimated Error 0.1126"

"22.4% posterior probability of decrease "

Under 18

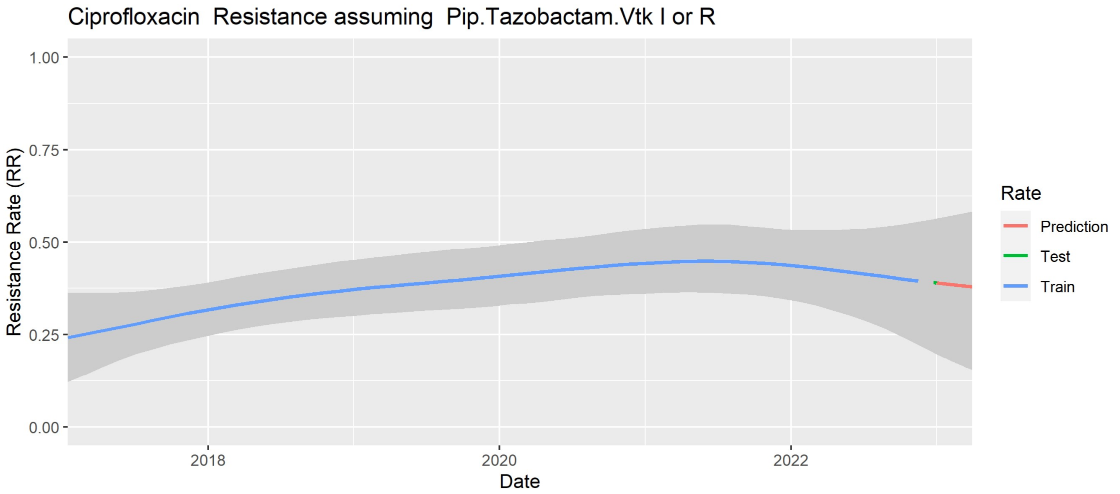


[1] "2017-01-14 Mid 0.242 (95%CI 0.122 to 0.362) Estimated Error 0.0612"

[1] "2022-12-24 Mid 0.390 (95%CI 0.201 to 0.562) Estimated Error 0.0938"

[1] "2022-12-30 Mid 0.389 (95%CI 0.198 to 0.564) Estimated Error 0.0950"

"7.4% posterior probability of decrease "

Over 80

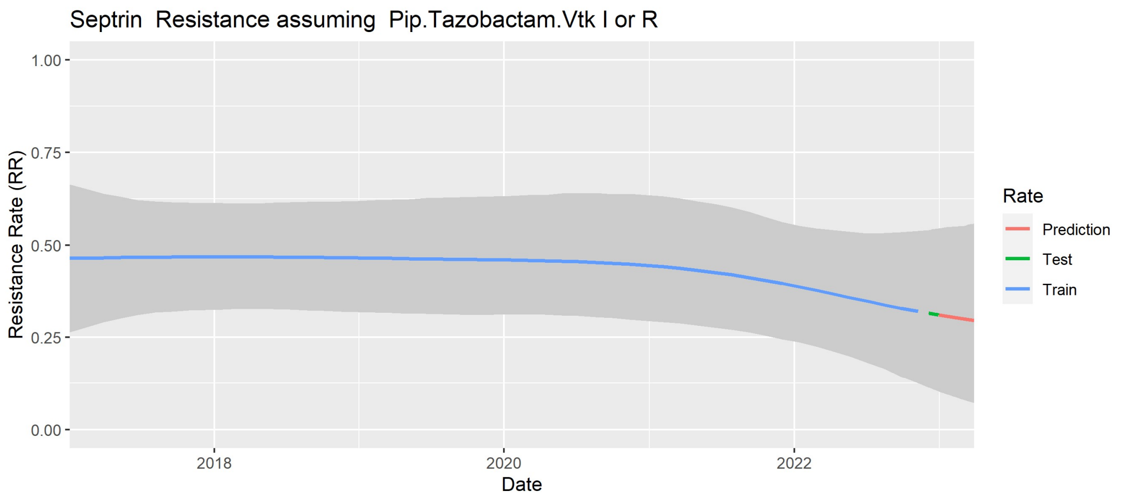


[1] "2017-01-02 Mid 0.463 (95%CI 0.264 to 0.664) Estimated Error 0.1016"

[1] "2022-12-06 Mid 0.315 (95%CI 0.113 to 0.541) Estimated Error 0.1111"

[1] "2022-12-29 Mid 0.311 (95%CI 0.103 to 0.545) Estimated Error 0.1144"

"83.2% posterior probability of decrease "

Under 18

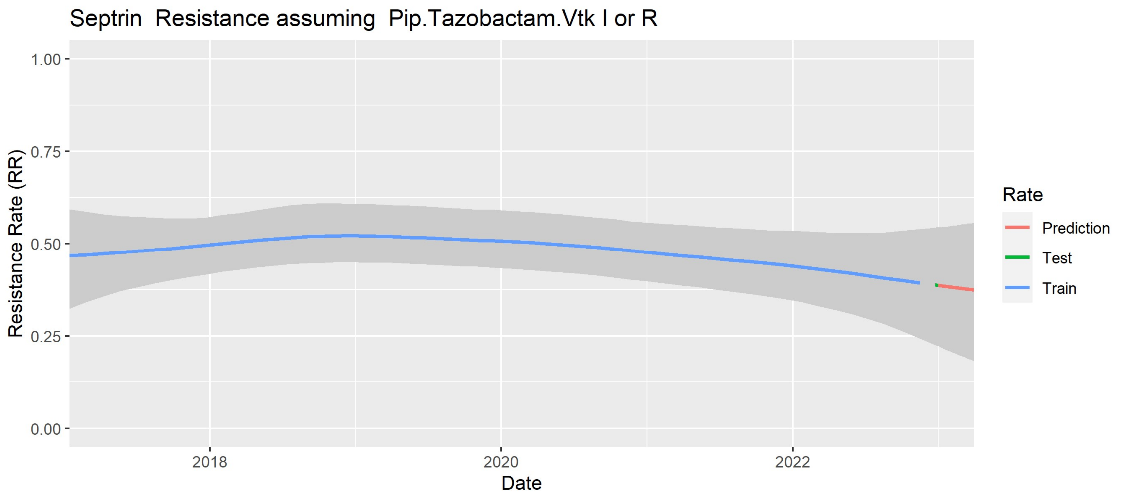


[1] "2017-01-14 Mid 0.467 (95%CI 0.324 to 0.594) Estimated Error 0.0682"

[1] "2022-12-24 Mid 0.387 (95%CI 0.225 to 0.543) Estimated Error 0.0804"

[1] "2022-12-30 Mid 0.387 (95%CI 0.223 to 0.544) Estimated Error 0.0813"

[1] "78.2% posterior probability of decrease "

Over 80

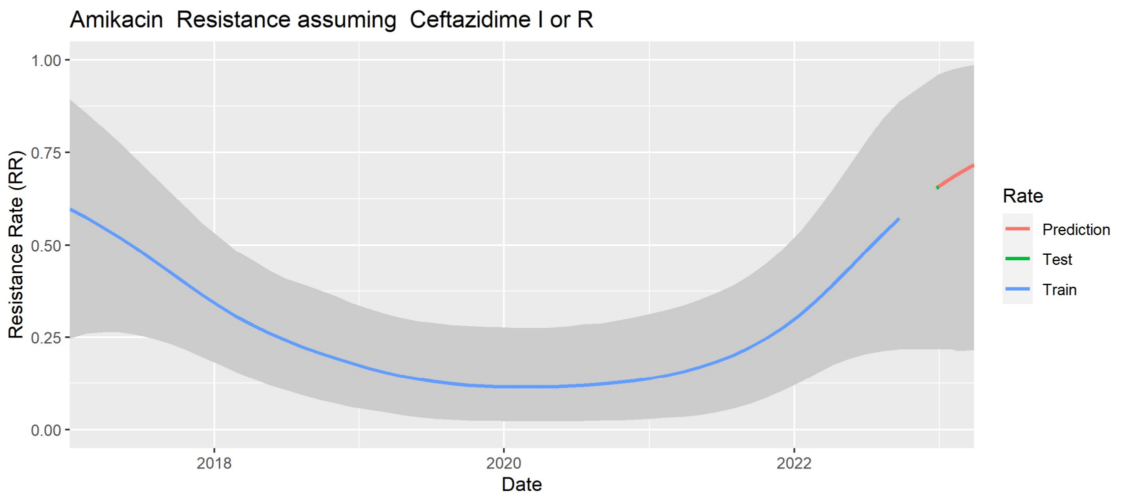


1] "2017-01-02 Mid 0.597 (95%CI 0.247 to 0.893) Estimated Error 0.1729"

[1] "2022-12-26 Mid 0.655 (95%CI 0.218 to 0.958) Estimated Error 0.2024"

[1] "2022-12-29 Mid 0.657 (95%CI 0.218 to 0.960) Estimated Error 0.2030"

"40.5% posterior probability of decrease "

Under 18

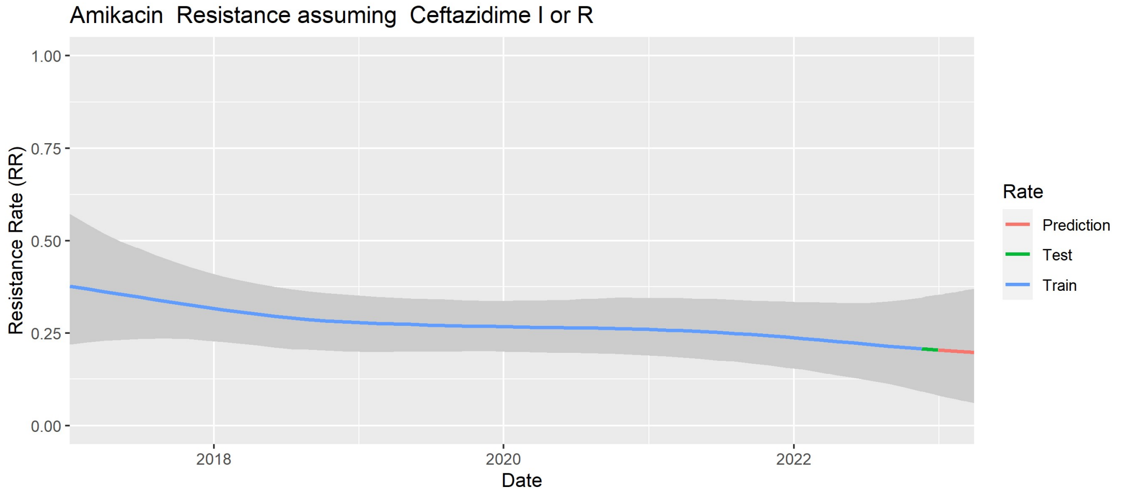


[1] "2017-01-03 Mid 0.376 (95%CI 0.219 to 0.573) Estimated Error 0.0889"

[1] "2022-11-19 Mid 0.208 (95%CI 0.092 to 0.346) Estimated Error 0.0642"

[1] "2022-12-30 Mid 0.205 (95%CI 0.081 to 0.353) Estimated Error 0.0681"

"93.5% posterior probability of decrease "

Over 80

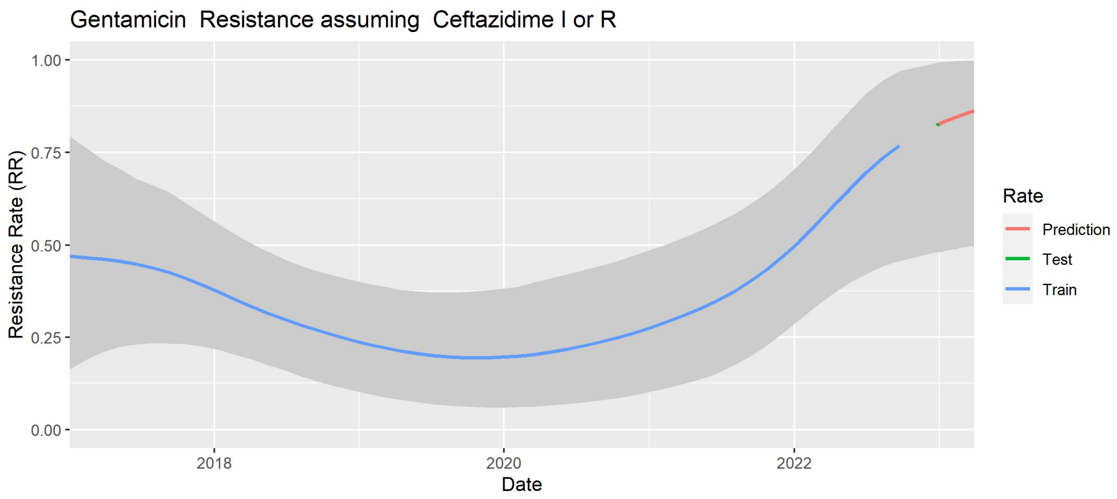


[1] "2017-01-02 Mid 0.469 (95%CI 0.164 to 0.792) Estimated Error 0.1662"

[1] "2022-12-26 Mid 0.825 (95%CI 0.480 to 0.992) Estimated Error 0.1392"

[1] "2022-12-29 Mid 0.826 (95%CI 0.480 to 0.993) Estimated Error 0.1392"

"4.9% posterior probability of decrease "

Under 18

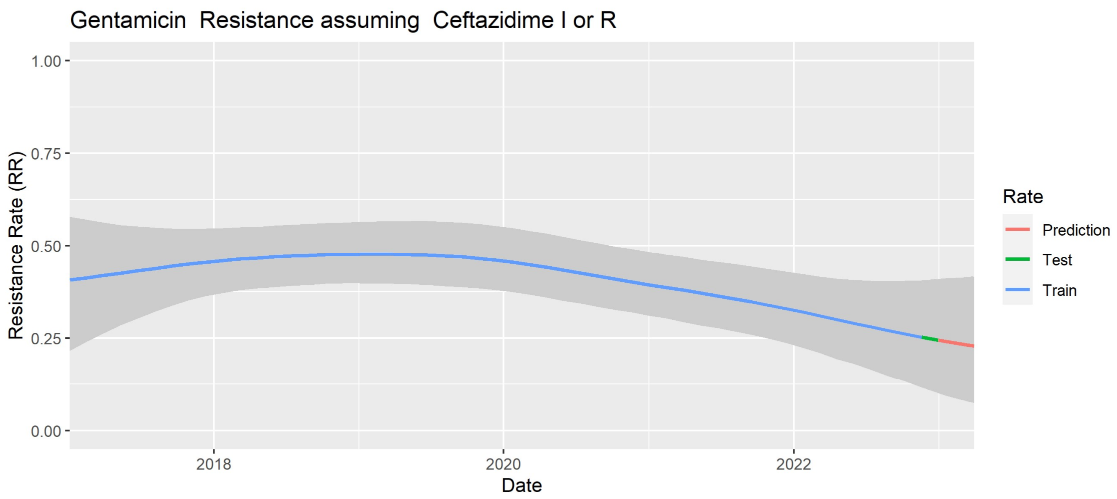


[1] "2017-01-03 Mid 0.407 (95%CI 0.217 to 0.578) Estimated Error 0.0940"

[1] "2022-11-19 Mid 0.253 (95%CI 0.116 to 0.405) Estimated Error 0.0743"

[1] "2022-12-30 Mid 0.245 (95%CI 0.101 to 0.408) Estimated Error 0.0789"

"91.6% posterior probability of decrease "

Over 80

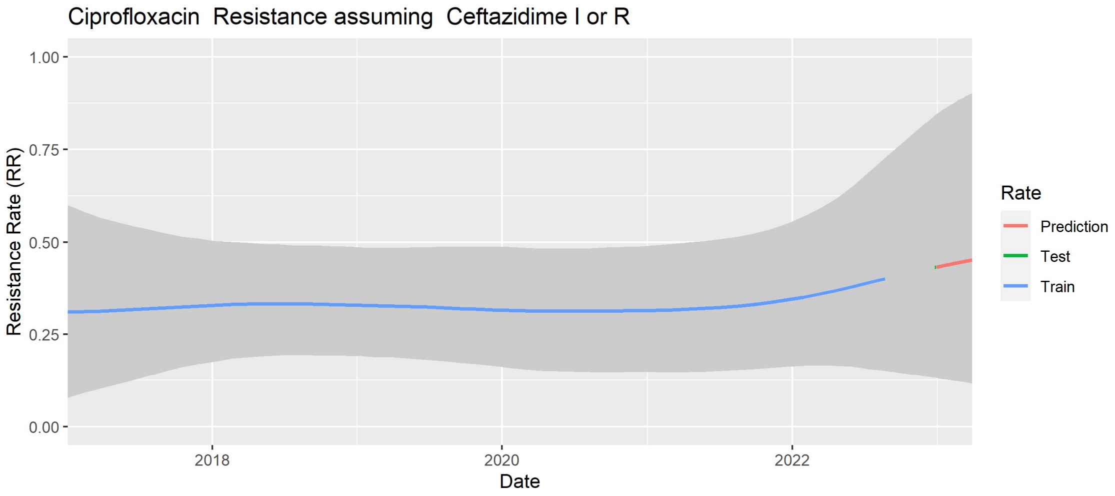


[1] "2017-01-02 Mid 0.310 (95%CI 0.078 to 0.600) Estimated Error 0.1370"

[1] "2022-12-26 Mid 0.431 (95%CI 0.133 to 0.843) Estimated Error 0.1860"

[1] "2022-12-29 Mid 0.431 (95%CI 0.132 to 0.845) Estimated Error 0.1868"

[1] "32.0% posterior probability of decrease "

Under 18

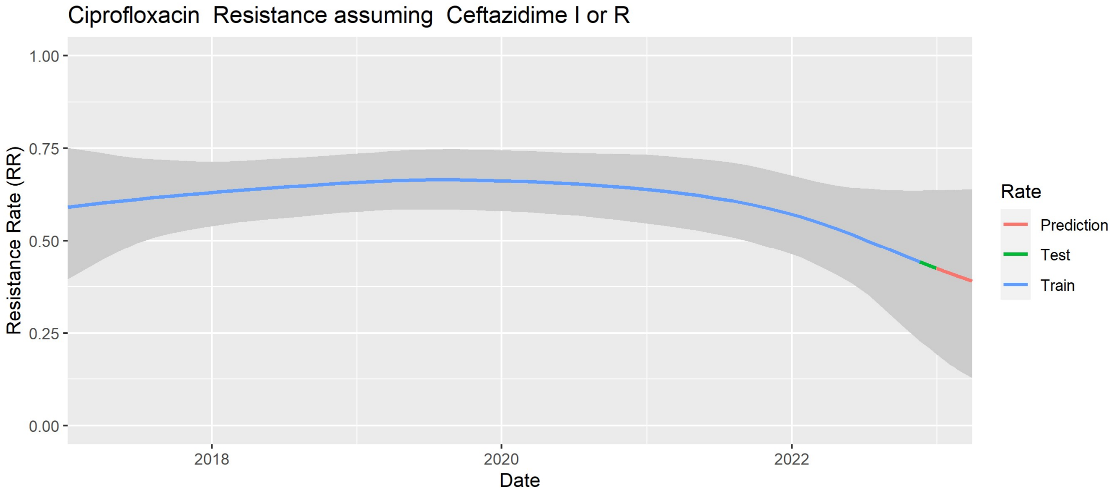


[1] "2017-01-03 Mid 0.591 (95%CI 0.396 to 0.751) Estimated Error 0.0904"

[1] "2022-11-19 Mid 0.442 (95%CI 0.228 to 0.635) Estimated Error 0.1048"

[1] "2022-12-30 Mid 0.425 (95%CI 0.194 to 0.636) Estimated Error 0.1147"

[1] "88.7% posterior probability of decrease "

Over 80

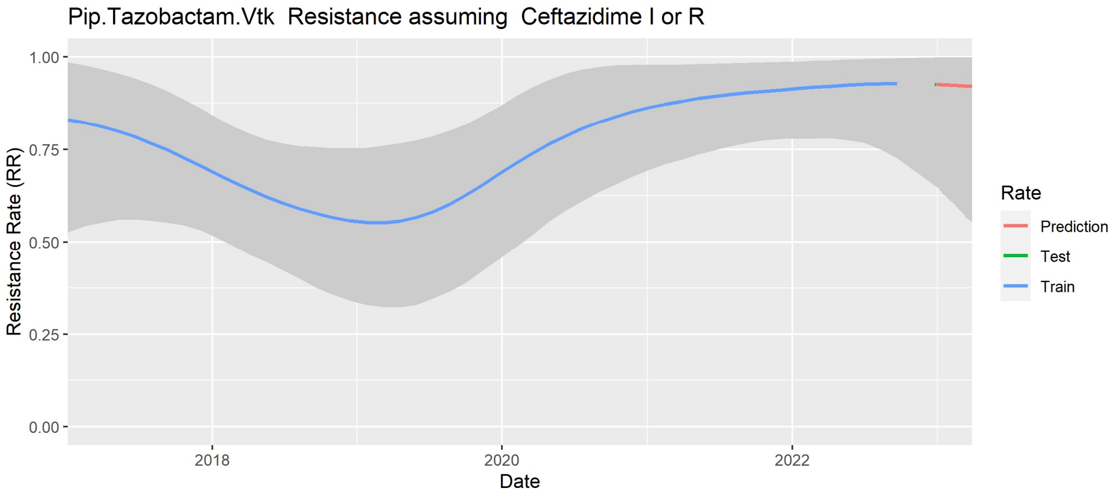


[1] "2017-01-02 Mid 0.830 (95%CI 0.526 to 0.985) Estimated Error 0.1224"

[1] "2022-12-26 Mid 0.926 (95%CI 0.652 to 0.999) Estimated Error 0.0948"

[1] "2022-12-29 Mid 0.925 (95%CI 0.651 to 0.999) Estimated Error 0.0956"

"21.0% posterior probability of decrease "

Under 18

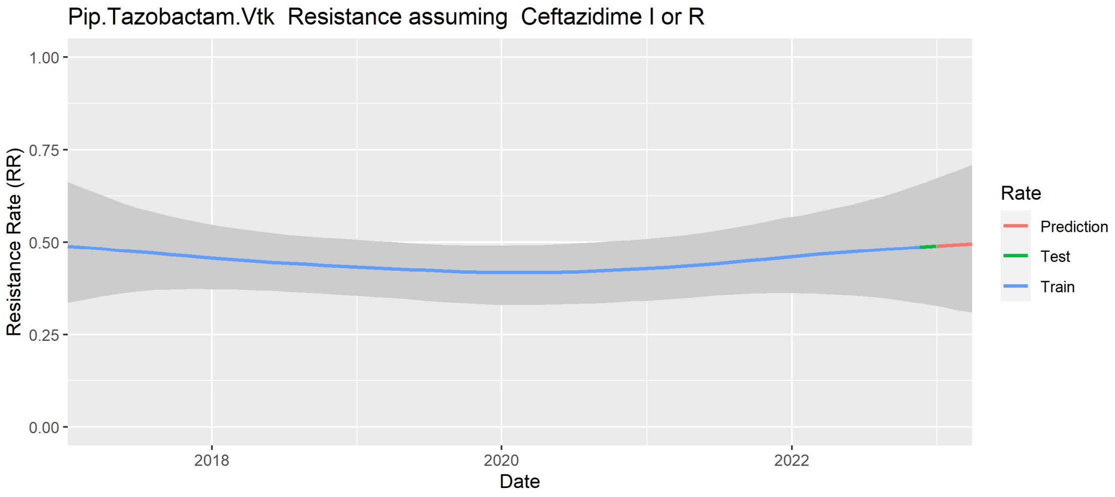


[1] "2017-01-03 Mid 0.489 (95%CI 0.337 to 0.662) Estimated Error 0.0833"

[1] "2022-11-19 Mid 0.486 (95%CI 0.335 to 0.656) Estimated Error 0.0822"

[1] "2022-12-30 Mid 0.489 (95%CI 0.327 to 0.672) Estimated Error 0.0883"

"50.8% posterior probability of decrease "

Over 80

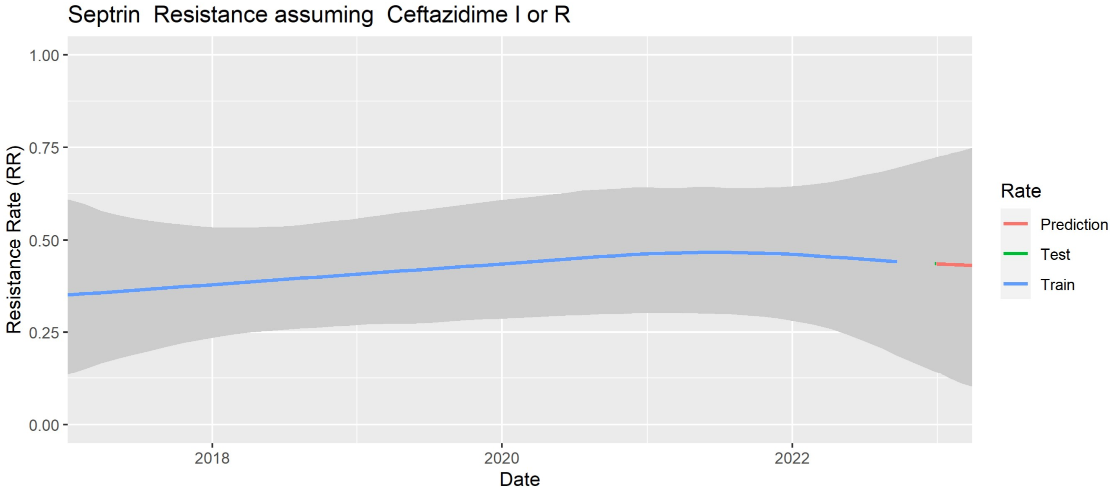


[1] "2017-01-02 Mid 0.352 (95%CI 0.136 to 0.610) Estimated Error 0.1221"

[1] "2022-12-26 Mid 0.435 (95%CI 0.144 to 0.723) Estimated Error 0.1481"

[1] "2022-12-29 Mid 0.435 (95%CI 0.142 to 0.723) Estimated Error 0.1487"

[1] "33.3% posterior probability of decrease "

Under 18

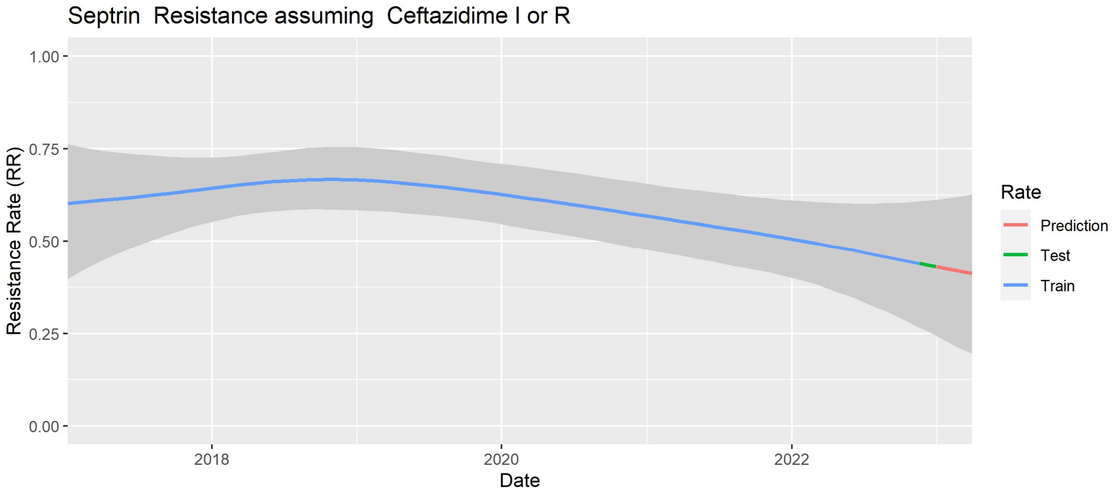


[1] "2017-01-03 Mid 0.602 (95%CI 0.398 to 0.762) Estimated Error 0.0940"

[1] "2022-11-19 Mid 0.439 (95%CI 0.265 to 0.608) Estimated Error 0.0874"

[1] "2022-12-30 Mid 0.430 (95%CI 0.245 to 0.612) Estimated Error 0.0944"

"90.5% posterior probability of decrease "

Over 80
