## Supplementary - EA tables for "Rational Empiric Antibiotic Escalation Applied to Specific Patient Groups" (from another version of this preprint: version 2, 2023-11-07)

Supplementary Material (EA Tables)

Posterior probability of resistance (Mid) with upper and lower 95% credible intervals

| | | Meropenem | | | Pip.Taz. | | | Ceftazidime | | | Cefotaxime. | | | Cefuroxime | |
|---|---|---|---|---|---|---|---|---|---|---|---|---|---|---|---|
| **Initial.Antibiotic** | **Mid** | **Upper** | **Lower** | **Mid** | **Upper** | **Lower** | **Mid** | **Upper** | **Lower** | **Mid** | **Upper** | **Lower** | **Mid** | **Upper** | **Lower** |
| **Meropenem** | NA | NA | NA | 0.67 | 0.83 | 0.47 | 0.62 | 0.78 | 0.43 | 0.89 | 0.99 | 0.62 | 0.95 | 1.00 | 0.82 |
| **Pip.Taz** | 0.12 | 0.18 | 0.07 | NA | NA | NA | 0.47 | 0.56 | 0.38 | 0.44 | 0.53 | 0.33 | 0.63 | 0.71 | 0.53 |
| **Ceftazidime** | 0.13 | 0.21 | 0.07 | 0.57 | 0.68 | 0.43 | NA | NA | NA | 0.78 | 0.86 | 0.67 | 0.97 | 0.99 | 0.93 |
| **Cefotaxime** | 0.12 | 0.18 | 0.08 | 0.34 | 0.44 | 0.24 | 0.51 | 0.59 | 0.44 | NA | NA | NA | 0.99 | 1.00 | 0.98 |
| **Cefuroxime** | 0.07 | 0.11 | 0.05 | 0.29 | 0.36 | 0.22 | 0.36 | 0.42 | 0.31 | 0.57 | 0.62 | 0.52 | NA | NA | NA |
| **Gentamicin** | 0.15 | 0.28 | 0.07 | 0.46 | 0.58 | 0.33 | 0.45 | 0.59 | 0.35 | 0.45 | 0.59 | 0.35 | 0.58 | 0.71 | 0.47 |
| **Amikacin** | 0.24 | 0.46 | 0.11 | 0.65 | 0.89 | 0.44 | 0.67 | 0.88 | 0.47 | 0.74 | 0.90 | 0.58 | 0.81 | 0.94 | 0.67 |
| **Coamoxiclav** | 0.05 | 0.07 | 0.03 | 0.26 | 0.32 | 0.21 | 0.20 | 0.24 | 0.17 | 0.32 | 0.37 | 0.27 | 0.53 | 0.59 | 0.47 |
| **Ciprofloxacin** | 0.03 | 0.07 | 0.01 | 0.34 | 0.45 | 0.23 | 0.41 | 0.50 | 0.33 | 0.42 | 0.52 | 0.33 | 0.63 | 0.74 | 0.54 |
| **Septrin** | 0.04 | 0.07 | 0.02 | 0.18 | 0.23 | 0.13 | 0.18 | 0.22 | 0.14 | 0.42 | 0.49 | 0.34 | 0.50 | 0.57 | 0.43 |

| | | Gentamicin | | | Amikacin | | | Coamoxiclav | | | Ciprofloxacin | | | Septrin. | |
|---|---|---|---|---|---|---|---|---|---|---|---|---|---|---|---|
| **Initial.Antibiotic** | **Mid** | **Upper** | **Lower** | **Mid** | **Upper** | **Lower** | **Mid** | **Upper** | **Lower** | **Mid** | **Upper** | **Lower** | **Mid** | **Upper** | **Lower** |
| **Meropenem** | 0.45 | 0.64 | 0.28 | 0.41 | 0.62 | 0.23 | 0.99 | 1.00 | 0.94 | 0.23 | 0.45 | 0.09 | 0.58 | 0.75 | 0.39 |
| **Pip.Taz** | 0.27 | 0.34 | 0.18 | 0.18 | 0.25 | 0.11 | 0.91 | 0.96 | 0.84 | 0.31 | 0.40 | 0.21 | 0.37 | 0.45 | 0.27 |
| **Ceftazidime** | 0.33 | 0.41 | 0.25 | 0.20 | 0.30 | 0.12 | 0.87 | 0.92 | 0.80 | 0.45 | 0.54 | 0.35 | 0.43 | 0.53 | 0.33 |
| **Cefotaxime** | 0.21 | 0.27 | 0.16 | 0.16 | 0.23 | 0.10 | 0.90 | 0.94 | 0.83 | 0.29 | 0.38 | 0.22 | 0.67 | 0.75 | 0.58 |
| **Cefuroxime** | 0.15 | 0.20 | 0.12 | 0.10 | 0.15 | 0.06 | 0.85 | 0.89 | 0.80 | 0.24 | 0.31 | 0.19 | 0.46 | 0.52 | 0.39 |
| **Gentamicin** | NA | NA | NA | 0.23 | 0.37 | 0.11 | 0.82 | 0.89 | 0.73 | 0.55 | 0.66 | 0.43 | 0.58 | 0.71 | 0.45 |
| **Amikacin** | 0.61 | 0.78 | 0.41 | NA | NA | NA | 0.85 | 0.94 | 0.71 | 0.66 | 0.86 | 0.41 | 0.45 | 0.64 | 0.24 |
| **Coamoxiclav** | 0.13 | 0.17 | 0.10 | 0.06 | 0.09 | 0.03 | NA | NA | NA | 0.17 | 0.21 | 0.13 | 0.44 | 0.48 | 0.39 |
| **Ciprofloxacin** | 0.35 | 0.43 | 0.27 | 0.16 | 0.25 | 0.08 | 0.72 | 0.80 | 0.64 | NA | NA | NA | 0.58 | 0.67 | 0.49 |
| **Septrin** | 0.17 | 0.22 | 0.12 | 0.05 | 0.09 | 0.02 | 0.78 | 0.82 | 0.73 | 0.24 | 0.30 | 0.19 | NA | NA | NA |

All Patients

| | | Meropenem | | | Pip.Taz. | | | Ceftazidime | | | Cefotaxime. | | | Cefuroxime | |
|---|---|---|---|---|---|---|---|---|---|---|---|---|---|---|---|
| **Initial.Antibioti** | **Mid** | **Upper** | **Lower** | **Mid** | **Upper** | **Lower** | **Mid** | **Upper** | **Lower** | **Mid** | **Upper** | **Lower** | **Mid** | **Upper** | **Lower** |
| **Meropenem** | NA | NA | NA | 0.86 | 1.00 | 0.43 | 0.84 | 1.00 | 0.40 | 0.95 | 1.00 | 0.51 | 0.95 | 1.00 | 0.52 |
| **Pip.Taz** | 0.10 | 0.23 | 0.02 | NA | NA | NA | 0.62 | 0.83 | 0.36 | 0.65 | 0.83 | 0.41 | 0.82 | 0.94 | 0.63 |
| **Ceftazidime** | 0.13 | 0.32 | 0.03 | 0.86 | 0.98 | 0.65 | NA | NA | NA | 0.89 | 0.98 | 0.72 | 0.97 | 1.00 | 0.88 |
| **Cefotaxime** | 0.13 | 0.27 | 0.04 | 0.65 | 0.80 | 0.44 | 0.66 | 0.82 | 0.43 | NA | NA | NA | 1.00 | 1.00 | 0.97 |
| **Cefuroxime** | 0.09 | 0.20 | 0.03 | 0.55 | 0.69 | 0.38 | 0.49 | 0.64 | 0.33 | 0.68 | 0.82 | 0.53 | NA | NA | NA |
| **Gentamicin** | 0.05 | 0.22 | 0.00 | 0.42 | 0.74 | 0.13 | 0.36 | 0.68 | 0.06 | 0.34 | 0.64 | 0.06 | 0.45 | 0.75 | 0.13 |
| **Amikacin** | 0.19 | 0.64 | 0.01 | 0.64 | 0.96 | 0.20 | 0.46 | 0.90 | 0.05 | 0.66 | 0.97 | 0.19 | 0.62 | 0.97 | 0.12 |
| **Coamoxiclav** | 0.06 | 0.13 | 0.02 | 0.41 | 0.53 | 0.29 | 0.30 | 0.42 | 0.18 | 0.43 | 0.57 | 0.29 | 0.63 | 0.76 | 0.50 |
| **Ciprofloxacin** | 0.02 | 0.10 | 0.00 | 0.43 | 0.75 | 0.16 | 0.55 | 0.85 | 0.25 | 0.43 | 0.74 | 0.14 | 0.53 | 0.82 | 0.22 |
| **Septrin** | 0.09 | 0.23 | 0.02 | 0.23 | 0.42 | 0.08 | 0.20 | 0.37 | 0.07 | 0.56 | 0.76 | 0.37 | 0.59 | 0.78 | 0.41 |

| | | Gentamicin | | | Amikacin | | | Coamoxiclav | | | Ciprofloxacin | | | Septrin. | |
|---|---|---|---|---|---|---|---|---|---|---|---|---|---|---|---|
| **Initial.Antibioti** | **Mid** | **Upper** | **Lower** | **Mid** | **Upper** | **Lower** | **Mid** | **Upper** | **Lower** | **Mid** | **Upper** | **Lower** | **Mid** | **Upper** | **Lower** |
| **Meropenem** | 0.26 | 0.76 | 0.01 | 0.24 | 0.86 | 0.00 | 0.95 | 1.00 | 0.53 | 0.04 | 0.38 | 0.00 | 0.18 | 0.68 | 0.00 |
| **Pip.Taz** | 0.19 | 0.36 | 0.07 | 0.14 | 0.31 | 0.04 | 0.94 | 1.00 | 0.79 | 0.17 | 0.34 | 0.06 | 0.23 | 0.42 | 0.10 |
| **Ceftazidime** | 0.24 | 0.45 | 0.06 | 0.17 | 0.40 | 0.04 | 0.98 | 1.00 | 0.88 | 0.28 | 0.53 | 0.11 | 0.30 | 0.70 | 0.11 |
| **Cefotaxime** | 0.17 | 0.32 | 0.05 | 0.14 | 0.31 | 0.04 | 0.98 | 1.00 | 0.91 | 0.18 | 0.33 | 0.06 | 0.50 | 0.70 | 0.32 |
| **Cefuroxime** | 0.14 | 0.26 | 0.04 | 0.10 | 0.22 | 0.03 | 0.99 | 1.00 | 0.94 | 0.14 | 0.26 | 0.05 | 0.37 | 0.55 | 0.24 |
| **Gentamicin** | NA | NA | NA | 0.24 | 0.59 | 0.03 | 0.80 | 0.97 | 0.46 | 0.43 | 0.72 | 0.16 | 0.50 | 0.81 | 0.18 |
| **Amikacin** | 0.93 | 1.00 | 0.60 | NA | NA | NA | 0.62 | 0.97 | 0.13 | 0.23 | 0.69 | 0.01 | 0.25 | 0.72 | 0.02 |
| **Coamoxiclav** | 0.15 | 0.24 | 0.07 | 0.07 | 0.15 | 0.02 | NA | NA | NA | 0.11 | 0.20 | 0.04 | 0.37 | 0.52 | 0.25 |
| **Ciprofloxacin** | 0.57 | 0.85 | 0.26 | 0.18 | 0.52 | 0.02 | 0.74 | 0.97 | 0.40 | NA | NA | NA | 0.37 | 0.70 | 0.07 |
| **Septrin** | 0.27 | 0.45 | 0.12 | 0.07 | 0.21 | 0.01 | 0.86 | 0.96 | 0.71 | 0.13 | 0.30 | 0.03 | NA | NA | NA |

ICU Patients

| | | Meropenem | | | Pip.Taz. | | | Ceftazidime | | | Cefotaxime. | | | Cefuroxime | |
|---|---|---|---|---|---|---|---|---|---|---|---|---|---|---|---|
| **Initial.Antibiotic** | **Mid** | **Upper** | **Lower** | **Mid** | **Upper** | **Lower** | **Mid** | **Upper** | **Lower** | **Mid** | **Upper** | **Lower** | **Mid** | **Upper** | **Lower** |
| **Meropenem** | NA | NA | NA | 0.93 | 1.00 | 0.71 | 0.93 | 1.00 | 0.72 | 0.97 | 1.00 | 0.71 | 0.97 | 1.00 | 0.72 |
| **Pip.Taz** | 0.37 | 0.66 | 0.16 | NA | NA | NA | 0.62 | 0.82 | 0.38 | 0.58 | 0.81 | 0.33 | 0.72 | 0.91 | 0.45 |
| **Ceftazidime** | 0.42 | 0.66 | 0.21 | 0.77 | 0.92 | 0.54 | NA | NA | NA | 0.88 | 0.98 | 0.70 | 0.99 | 1.00 | 0.94 |
| **Cefotaxime** | 0.42 | 0.68 | 0.21 | 0.62 | 0.82 | 0.40 | 0.71 | 0.90 | 0.45 | NA | NA | NA | 0.99 | 1.00 | 0.95 |
| **Cefuroxime** | 0.26 | 0.46 | 0.11 | 0.46 | 0.64 | 0.24 | 0.52 | 0.72 | 0.30 | 0.63 | 0.81 | 0.44 | NA | NA | NA |
| **Gentamicin** | 0.56 | 0.84 | 0.20 | 0.80 | 0.97 | 0.50 | 0.72 | 0.95 | 0.42 | 0.68 | 0.92 | 0.37 | 0.74 | 0.95 | 0.43 |
| **Amikacin** | 0.41 | 0.80 | 0.11 | 0.73 | 0.95 | 0.39 | 0.66 | 0.93 | 0.32 | 0.64 | 0.91 | 0.31 | 0.75 | 0.97 | 0.40 |
| **Coamoxiclav** | 0.21 | 0.41 | 0.09 | 0.48 | 0.67 | 0.30 | 0.36 | 0.56 | 0.19 | 0.46 | 0.66 | 0.30 | 0.66 | 0.82 | 0.49 |
| **Ciprofloxacin** | 0.14 | 0.43 | 0.01 | 0.40 | 0.67 | 0.14 | 0.39 | 0.68 | 0.13 | 0.44 | 0.71 | 0.17 | 0.49 | 0.79 | 0.13 |
| **Septrin** | 0.24 | 0.52 | 0.07 | 0.36 | 0.60 | 0.15 | 0.35 | 0.61 | 0.14 | 0.59 | 0.79 | 0.39 | 0.61 | 0.80 | 0.37 |

| | | Gentamicin | | | Amikacin | | | Coamoxiclav | | | Ciprofloxacin | | | Septrin. | |
|---|---|---|---|---|---|---|---|---|---|---|---|---|---|---|---|
| **Initial.Antibiotic** | **Mid** | **Upper** | **Lower** | **Mid** | **Upper** | **Lower** | **Mid** | **Upper** | **Lower** | **Mid** | **Upper** | **Lower** | **Mid** | **Upper** | **Lower** |
| **Meropenem** | 0.70 | 0.94 | 0.36 | 0.54 | 0.87 | 0.19 | 0.97 | 1.00 | 0.69 | 0.37 | 0.85 | 0.03 | 0.39 | 0.74 | 0.10 |
| **Pip.Taz** | 0.32 | 0.55 | 0.12 | 0.30 | 0.56 | 0.09 | 0.87 | 0.98 | 0.62 | 0.31 | 0.64 | 0.09 | 0.42 | 0.75 | 0.16 |
| **Ceftazidime** | 0.39 | 0.64 | 0.18 | 0.38 | 0.62 | 0.16 | 0.92 | 0.99 | 0.75 | 0.35 | 0.62 | 0.13 | 0.39 | 0.64 | 0.16 |
| **Cefotaxime** | 0.28 | 0.54 | 0.11 | 0.29 | 0.52 | 0.09 | 0.91 | 0.99 | 0.72 | 0.27 | 0.52 | 0.07 | 0.59 | 0.82 | 0.33 |
| **Cefuroxime** | 0.17 | 0.34 | 0.07 | 0.21 | 0.39 | 0.07 | 0.78 | 0.93 | 0.52 | 0.17 | 0.35 | 0.04 | 0.40 | 0.59 | 0.22 |
| **Gentamicin** | NA | NA | NA | 0.72 | 0.93 | 0.41 | 0.90 | 1.00 | 0.63 | 0.45 | 0.82 | 0.09 | 0.32 | 0.65 | 0.06 |
| **Amikacin** | 0.71 | 0.96 | 0.36 | NA | NA | NA | 0.69 | 0.97 | 0.25 | 0.68 | 0.96 | 0.18 | 0.03 | 0.22 | 0.00 |
| **Coamoxiclav** | 0.18 | 0.35 | 0.08 | 0.19 | 0.36 | 0.07 | NA | NA | NA | 0.29 | 0.62 | 0.12 | 0.44 | 0.64 | 0.26 |
| **Ciprofloxacin** | 0.22 | 0.50 | 0.05 | 0.19 | 0.53 | 0.01 | 0.71 | 0.92 | 0.38 | NA | NA | NA | 0.80 | 1.00 | 0.43 |
| **Septrin** | 0.11 | 0.26 | 0.02 | 0.02 | 0.12 | 0.00 | 0.86 | 0.97 | 0.66 | 0.40 | 0.71 | 0.18 | NA | NA | NA |

Haemato-oncology Patients

| | | Meropenem | | | Pip.Taz. | | | Ceftazidime | | | Cefotaxime. | | | Cefuroxime | |
|---|---|---|---|---|---|---|---|---|---|---|---|---|---|---|---|
| **Initial.Antibiotic** | **Mid** | **Upper** | **Lower** | **Mid** | **Upper** | **Lower** | **Mid** | **Upper** | **Lower** | **Mid** | **Upper** | **Lower** | **Mid** | **Upper** | **Lower** |
| **Meropenem** | NA | NA | NA | x | x | x | x | x | x | x | x | x | x | x | x |
| **Pip.Taz** | 0.01 | 0.07 | 0.00 | NA | NA | NA | 0.36 | 0.57 | 0.20 | 0.29 | 0.48 | 0.14 | 0.48 | 0.68 | 0.31 |
| **Ceftazidime** | 0.01 | 0.06 | 0.00 | 0.49 | 0.72 | 0.29 | NA | NA | NA | 0.79 | 0.94 | 0.56 | 0.98 | 1.00 | 0.88 |
| **Cefotaxime** | 0.06 | 0.20 | 0.00 | 0.26 | 0.44 | 0.13 | 0.50 | 0.69 | 0.34 | NA | NA | NA | 1.00 | 1.00 | 0.97 |
| **Cefuroxime** | 0.03 | 0.13 | 0.00 | 0.26 | 0.41 | 0.14 | 0.37 | 0.54 | 0.24 | 0.56 | 0.70 | 0.40 | NA | NA | NA |
| **Gentamicin** | 0.01 | 0.09 | 0.00 | 0.41 | 0.63 | 0.21 | 0.34 | 0.58 | 0.17 | 0.32 | 0.53 | 0.15 | 0.51 | 0.73 | 0.30 |
| **Amikacin** | 0.02 | 0.18 | 0.00 | 0.44 | 0.79 | 0.14 | 0.57 | 0.88 | 0.26 | 0.63 | 0.89 | 0.31 | 0.72 | 0.93 | 0.39 |
| **Coamoxiclav** | 0.02 | 0.07 | 0.00 | 0.23 | 0.32 | 0.14 | 0.14 | 0.22 | 0.08 | 0.25 | 0.35 | 0.16 | 0.41 | 0.51 | 0.30 |
| **Ciprofloxacin** | 0.02 | 0.12 | 0.00 | 0.30 | 0.51 | 0.14 | 0.38 | 0.56 | 0.22 | 0.38 | 0.55 | 0.22 | 0.58 | 0.76 | 0.40 |
| **Septrin** | 0.03 | 0.10 | 0.00 | 0.16 | 0.26 | 0.08 | 0.14 | 0.23 | 0.08 | 0.31 | 0.42 | 0.20 | 0.39 | 0.50 | 0.26 |

| | | Gentamicin | | | Amikacin | | | Coamoxiclav | | | Ciprofloxacin | | | Septrin. | |
|---|---|---|---|---|---|---|---|---|---|---|---|---|---|---|---|
| **Initial.Antibiotic** | **Mid** | **Upper** | **Lower** | **Mid** | **Upper** | **Lower** | **Mid** | **Upper** | **Lower** | **Mid** | **Upper** | **Lower** | **Mid** | **Upper** | **Lower** |
| **Meropenem** | x | x | x | x | x | x | x | x | x | x | x | x | x | x | x |
| **Pip.Taz** | 0.29 | 0.47 | 0.12 | 0.13 | 0.28 | 0.03 | 0.77 | 0.91 | 0.55 | 0.33 | 0.53 | 0.16 | 0.40 | 0.59 | 0.22 |
| **Ceftazidime** | 0.34 | 0.55 | 0.16 | 0.14 | 0.34 | 0.02 | 0.70 | 0.87 | 0.46 | 0.57 | 0.78 | 0.33 | 0.48 | 0.73 | 0.24 |
| **Cefotaxime** | 0.21 | 0.36 | 0.08 | 0.10 | 0.24 | 0.01 | 0.78 | 0.90 | 0.59 | 0.36 | 0.53 | 0.20 | 0.66 | 0.84 | 0.43 |
| **Cefuroxime** | 0.19 | 0.30 | 0.09 | 0.08 | 0.18 | 0.02 | 0.74 | 0.85 | 0.57 | 0.33 | 0.47 | 0.21 | 0.49 | 0.65 | 0.33 |
| **Gentamicin** | NA | NA | NA | 0.13 | 0.30 | 0.02 | 0.73 | 0.90 | 0.51 | 0.58 | 0.82 | 0.37 | 0.68 | 0.90 | 0.42 |
| **Amikacin** | 0.56 | 0.85 | 0.27 | NA | NA | NA | 0.66 | 0.93 | 0.24 | 0.63 | 0.91 | 0.23 | 0.38 | 0.75 | 0.08 |
| **Coamoxiclav** | 0.14 | 0.22 | 0.07 | 0.04 | 0.09 | 0.01 | NA | NA | NA | 0.19 | 0.27 | 0.11 | 0.57 | 0.71 | 0.43 |
| **Ciprofloxacin** | 0.42 | 0.61 | 0.26 | 0.11 | 0.26 | 0.01 | 0.58 | 0.78 | 0.35 | NA | NA | NA | 0.48 | 0.68 | 0.29 |
| **Septrin** | 0.18 | 0.27 | 0.10 | 0.05 | 0.11 | 0.01 | 0.74 | 0.85 | 0.62 | 0.23 | 0.34 | 0.14 | NA | NA | NA |

Urine Source

| | | Meropenem | | | Pip.Taz. | | | Ceftazidime | | | Cefotaxime. | | | Cefuroxime | |
|---|---|---|---|---|---|---|---|---|---|---|---|---|---|---|---|
| **Initial.Antibiotic** | **Mid** | **Upper** | **Lower** | **Mid** | **Upper** | **Lower** | **Mid** | **Upper** | **Lower** | **Mid** | **Upper** | **Lower** | **Mid** | **Upper** | **Lower** |
| **Meropenem** | NA | NA | NA | 0.05 | 0.51 | 0.00 | 0.11 | 0.72 | 0.00 | 0.93 | 1.00 | 0.35 | 0.93 | 1.00 | 0.36 |
| **Pip.Taz** | 0.05 | 0.16 | 0.00 | NA | NA | NA | 0.49 | 0.70 | 0.23 | 0.41 | 0.63 | 0.17 | 0.60 | 0.80 | 0.35 |
| **Ceftazidime** | 0.04 | 0.15 | 0.00 | 0.47 | 0.68 | 0.23 | NA | NA | NA | 0.61 | 0.80 | 0.39 | 0.92 | 0.99 | 0.77 |
| **Cefotaxime** | 0.11 | 0.25 | 0.02 | 0.38 | 0.59 | 0.17 | 0.64 | 0.82 | 0.44 | NA | NA | NA | 0.99 | 1.00 | 0.93 |
| **Cefuroxime** | 0.06 | 0.16 | 0.01 | 0.32 | 0.48 | 0.17 | 0.54 | 0.71 | 0.38 | 0.60 | 0.79 | 0.44 | NA | NA | NA |
| **Gentamicin** | 0.02 | 0.17 | 0.00 | 0.35 | 0.71 | 0.04 | 0.41 | 0.75 | 0.12 | 0.34 | 0.69 | 0.07 | 0.45 | 0.77 | 0.11 |
| **Amikacin** | 0.02 | 0.18 | 0.00 | 0.65 | 0.99 | 0.19 | 0.49 | 0.91 | 0.06 | 0.82 | 1.00 | 0.31 | 0.83 | 1.00 | 0.25 |
| **Coamoxiclav** | 0.04 | 0.09 | 0.01 | 0.28 | 0.39 | 0.18 | 0.29 | 0.40 | 0.19 | 0.29 | 0.41 | 0.19 | 0.46 | 0.57 | 0.33 |
| **Ciprofloxacin** | 0.02 | 0.09 | 0.00 | 0.38 | 0.61 | 0.18 | 0.44 | 0.66 | 0.24 | 0.60 | 0.93 | 0.32 | 0.71 | 0.94 | 0.47 |
| **Septrin** | 0.08 | 0.22 | 0.02 | 0.26 | 0.44 | 0.10 | 0.28 | 0.46 | 0.14 | 0.45 | 0.65 | 0.28 | 0.56 | 0.76 | 0.39 |

| | | Gentamicin | | | Amikacin | | | Coamoxiclav | | | Ciprofloxacin | | | Septrin. | |
|---|---|---|---|---|---|---|---|---|---|---|---|---|---|---|---|
| **Initial.Antibiotic** | **Mid** | **Upper** | **Lower** | **Mid** | **Upper** | **Lower** | **Mid** | **Upper** | **Lower** | **Mid** | **Upper** | **Lower** | **Mid** | **Upper** | **Lower** |
| **Meropenem** | 0.10 | 0.73 | 0.00 | 0.01 | 0.06 | 0.00 | 0.94 | 1.00 | 0.36 | 0.17 | 0.65 | 0.00 | 0.96 | 1.00 | 0.56 |
| **Pip.Taz** | 0.14 | 0.32 | 0.02 | 0.18 | 0.40 | 0.05 | 0.94 | 1.00 | 0.77 | 0.32 | 0.52 | 0.13 | 0.37 | 0.60 | 0.15 |
| **Ceftazidime** | 0.12 | 0.27 | 0.02 | 0.11 | 0.25 | 0.02 | 0.89 | 0.97 | 0.74 | 0.32 | 0.52 | 0.14 | 0.35 | 0.57 | 0.17 |
| **Cefotaxime** | 0.18 | 0.36 | 0.05 | 0.19 | 0.37 | 0.06 | 0.88 | 0.97 | 0.72 | 0.43 | 0.67 | 0.24 | 0.56 | 0.77 | 0.33 |
| **Cefuroxime** | 0.12 | 0.23 | 0.04 | 0.11 | 0.22 | 0.03 | 0.81 | 0.91 | 0.67 | 0.30 | 0.49 | 0.16 | 0.44 | 0.68 | 0.27 |
| **Gentamicin** | NA | NA | NA | 0.14 | 0.42 | 0.00 | 0.95 | 1.00 | 0.75 | 0.41 | 0.77 | 0.08 | 0.68 | 0.94 | 0.36 |
| **Amikacin** | 0.29 | 0.74 | 0.01 | NA | NA | NA | 0.85 | 1.00 | 0.33 | 0.81 | 1.00 | 0.28 | 0.73 | 1.00 | 0.21 |
| **Coamoxiclav** | 0.12 | 0.21 | 0.05 | 0.06 | 0.13 | 0.02 | NA | NA | NA | 0.22 | 0.44 | 0.11 | 0.34 | 0.46 | 0.21 |
| **Ciprofloxacin** | 0.25 | 0.50 | 0.07 | 0.25 | 0.47 | 0.09 | 0.88 | 0.98 | 0.73 | NA | NA | NA | 0.54 | 0.74 | 0.32 |
| **Septrin** | 0.17 | 0.32 | 0.06 | 0.07 | 0.18 | 0.01 | 0.81 | 0.92 | 0.64 | 0.32 | 0.49 | 0.17 | NA | NA | NA |

GI source

| | | Meropenem | | | Pip.Taz. | | | Ceftazidime | | | Cefotaxime. | | | Cefuroxime | |
|---|---|---|---|---|---|---|---|---|---|---|---|---|---|---|---|
| **Initial.Antibiotic** | **Mid** | **Upper** | **Lower** | **Mid** | **Upper** | **Lower** | **Mid** | **Upper** | **Lower** | **Mid** | **Upper** | **Lower** | **Mid** | **Upper** | **Lower** |
| **Meropenem** | NA | NA | NA | 0.90 | 1.00 | 0.59 | 0.88 | 1.00 | 0.57 | 0.98 | 1.00 | 0.78 | 0.98 | 1.00 | 0.78 |
| **Pip.Taz** | 0.34 | 0.58 | 0.14 | NA | NA | NA | 0.52 | 0.81 | 0.18 | 0.65 | 0.86 | 0.33 | 0.74 | 0.91 | 0.50 |
| **Ceftazidime** | 0.58 | 0.90 | 0.25 | 0.92 | 1.00 | 0.65 | NA | NA | NA | 0.92 | 1.00 | 0.69 | 0.90 | 1.00 | 0.68 |
| **Cefotaxime** | 0.37 | 0.67 | 0.15 | 0.67 | 0.89 | 0.40 | 0.57 | 0.82 | 0.34 | NA | NA | NA | 0.95 | 1.00 | 0.81 |
| **Cefuroxime** | 0.28 | 0.54 | 0.11 | 0.61 | 0.86 | 0.35 | 0.42 | 0.66 | 0.23 | 0.71 | 0.87 | 0.52 | NA | NA | NA |
| **Gentamicin** | 0.48 | 0.84 | 0.17 | 0.92 | 1.00 | 0.66 | 0.91 | 1.00 | 0.64 | 0.89 | 1.00 | 0.59 | 0.91 | 1.00 | 0.64 |
| **Amikacin** | 0.55 | 0.93 | 0.12 | 0.96 | 1.00 | 0.68 | 0.96 | 1.00 | 0.73 | 0.95 | 1.00 | 0.66 | 0.95 | 1.00 | 0.62 |
| **Coamoxiclav** | 0.16 | 0.31 | 0.06 | 0.49 | 0.68 | 0.30 | 0.28 | 0.44 | 0.15 | 0.44 | 0.61 | 0.26 | 0.54 | 0.73 | 0.32 |
| **Ciprofloxacin** | 0.44 | 0.86 | 0.06 | 0.86 | 1.00 | 0.42 | 0.83 | 1.00 | 0.41 | 0.91 | 1.00 | 0.53 | 0.94 | 1.00 | 0.60 |
| **Septrin** | 0.22 | 0.47 | 0.06 | 0.36 | 0.66 | 0.13 | 0.30 | 0.56 | 0.12 | 0.61 | 0.83 | 0.32 | 0.66 | 0.87 | 0.36 |

| | | Gentamicin | | | Amikacin | | | Coamoxiclav | | | Ciprofloxacin | | | Septrin. | |
|---|---|---|---|---|---|---|---|---|---|---|---|---|---|---|---|
| **Initial.Antibiotic** | **Mid** | **Upper** | **Lower** | **Mid** | **Upper** | **Lower** | **Mid** | **Upper** | **Lower** | **Mid** | **Upper** | **Lower** | **Mid** | **Upper** | **Lower** |
| **Pip.Taz** | 0.45 | 0.70 | 0.22 | 0.28 | 0.55 | 0.09 | 0.99 | 1.00 | 0.95 | 0.26 | 0.50 | 0.09 | 0.31 | 0.54 | 0.11 |
| **Ceftazidime** | 0.82 | 0.99 | 0.46 | 0.66 | 0.96 | 0.23 | 0.99 | 1.00 | 0.90 | 0.44 | 0.89 | 0.13 | 0.43 | 0.72 | 0.14 |
| **Cefotaxime** | 0.52 | 0.83 | 0.22 | 0.39 | 0.74 | 0.10 | 0.99 | 1.00 | 0.96 | 0.24 | 0.52 | 0.07 | 0.51 | 0.77 | 0.22 |
| **Cefuroxime** | 0.41 | 0.71 | 0.16 | 0.27 | 0.57 | 0.07 | 0.98 | 1.00 | 0.89 | 0.17 | 0.39 | 0.05 | 0.42 | 0.64 | 0.18 |
| **Gentamicin** | NA | NA | NA | 0.47 | 0.87 | 0.12 | 0.86 | 1.00 | 0.51 | 0.37 | 0.84 | 0.06 | 0.49 | 0.80 | 0.15 |
| **Amikacin** | 0.99 | 1.00 | 0.85 | NA | NA | NA | 0.97 | 1.00 | 0.72 | x | x | x | 0.23 | 0.69 | 0.01 |
| **Coamoxiclav** | 0.19 | 0.35 | 0.09 | 0.14 | 0.31 | 0.03 | NA | NA | NA | 0.09 | 0.20 | 0.03 | 0.39 | 0.55 | 0.22 |
| **Ciprofloxacin** | 0.73 | 0.99 | 0.27 | 0.30 | 0.92 | 0.00 | 0.92 | 1.00 | 0.54 | NA | NA | NA | 0.67 | 0.97 | 0.22 |
| **Septrin** | 0.21 | 0.45 | 0.06 | 0.04 | 0.20 | 0.00 | 0.93 | 0.99 | 0.79 | 0.13 | 0.34 | 0.03 | NA | NA | NA |

Patients over 80 years

| | | Meropenem | | | Pip.Taz. | | | Ceftazidime | | | Cefotaxime. | | | Cefuroxime | |
|---|---|---|---|---|---|---|---|---|---|---|---|---|---|---|---|
| **Initial.Antibioti** | **Mid** | **Upper** | **Lower** | **Mid** | **Upper** | **Lower** | **Mid** | **Upper** | **Lower** | **Mid** | **Upper** | **Lower** | **Mid** | **Upper** | **Lower** |
| **Meropenem** | NA | NA | NA | 0.90 | 1.00 | 0.59 | 0.88 | 1.00 | 0.57 | 0.98 | 1.00 | 0.78 | 0.98 | 1.00 | 0.78 |
| **Pip.Taz** | 0.34 | 0.58 | 0.14 | NA | NA | NA | 0.52 | 0.81 | 0.18 | 0.65 | 0.86 | 0.33 | 0.74 | 0.91 | 0.50 |
| **Ceftazidime** | 0.58 | 0.90 | 0.25 | 0.92 | 1.00 | 0.65 | NA | NA | NA | 0.92 | 1.00 | 0.69 | 0.90 | 1.00 | 0.68 |
| **Cefotaxime** | 0.37 | 0.67 | 0.15 | 0.67 | 0.89 | 0.40 | 0.57 | 0.82 | 0.34 | NA | NA | NA | 0.95 | 1.00 | 0.81 |
| **Cefuroxime** | 0.28 | 0.54 | 0.11 | 0.61 | 0.86 | 0.35 | 0.42 | 0.66 | 0.23 | 0.71 | 0.87 | 0.52 | NA | NA | NA |
| **Gentamicin** | 0.48 | 0.84 | 0.17 | 0.92 | 1.00 | 0.66 | 0.91 | 1.00 | 0.64 | 0.89 | 1.00 | 0.59 | 0.91 | 1.00 | 0.64 |
| **Amikacin** | 0.55 | 0.93 | 0.12 | 0.96 | 1.00 | 0.68 | 0.96 | 1.00 | 0.73 | 0.95 | 1.00 | 0.66 | 0.95 | 1.00 | 0.62 |
| **Coamoxiclav** | 0.16 | 0.31 | 0.06 | 0.49 | 0.68 | 0.30 | 0.28 | 0.44 | 0.15 | 0.44 | 0.61 | 0.26 | 0.54 | 0.73 | 0.32 |
| **Ciprofloxacin** | 0.44 | 0.86 | 0.06 | 0.86 | 1.00 | 0.42 | 0.83 | 1.00 | 0.41 | 0.91 | 1.00 | 0.53 | 0.94 | 1.00 | 0.60 |
| **Septrin** | 0.22 | 0.47 | 0.06 | 0.36 | 0.66 | 0.13 | 0.30 | 0.56 | 0.12 | 0.61 | 0.83 | 0.32 | 0.66 | 0.87 | 0.36 |

| | | Gentamicin | | | Amikacin | | | Coamoxiclav | | | Ciprofloxacin | | | Septrin. | |
|---|---|---|---|---|---|---|---|---|---|---|---|---|---|---|---|
| **Initial.Antibioti** | **Mid** | **Upper** | **Lower** | **Mid** | **Upper** | **Lower** | **Mid** | **Upper** | **Lower** | **Mid** | **Upper** | **Lower** | **Mid** | **Upper** | **Lower** |
| **Pip.Taz** | 0.45 | 0.70 | 0.22 | 0.28 | 0.55 | 0.09 | 0.99 | 1.00 | 0.95 | 0.26 | 0.50 | 0.09 | 0.31 | 0.54 | 0.11 |
| **Ceftazidime** | 0.82 | 0.99 | 0.46 | 0.66 | 0.96 | 0.23 | 0.99 | 1.00 | 0.90 | 0.44 | 0.89 | 0.13 | 0.43 | 0.72 | 0.14 |
| **Cefotaxime** | 0.52 | 0.83 | 0.22 | 0.39 | 0.74 | 0.10 | 0.99 | 1.00 | 0.96 | 0.24 | 0.52 | 0.07 | 0.51 | 0.77 | 0.22 |
| **Cefuroxime** | 0.41 | 0.71 | 0.16 | 0.27 | 0.57 | 0.07 | 0.98 | 1.00 | 0.89 | 0.17 | 0.39 | 0.05 | 0.42 | 0.64 | 0.18 |
| **Gentamicin** | NA | NA | NA | 0.47 | 0.87 | 0.12 | 0.86 | 1.00 | 0.51 | 0.37 | 0.84 | 0.06 | 0.49 | 0.80 | 0.15 |
| **Amikacin** | 0.99 | 1.00 | 0.85 | NA | NA | NA | 0.97 | 1.00 | 0.72 | x | x | x | 0.23 | 0.69 | 0.01 |
| **Coamoxiclav** | 0.19 | 0.35 | 0.09 | 0.14 | 0.31 | 0.03 | NA | NA | NA | 0.09 | 0.20 | 0.03 | 0.39 | 0.55 | 0.22 |
| **Ciprofloxacin** | 0.73 | 0.99 | 0.27 | 0.30 | 0.92 | 0.00 | 0.92 | 1.00 | 0.54 | NA | NA | NA | 0.67 | 0.97 | 0.22 |
| **Septrin** | 0.21 | 0.45 | 0.06 | 0.04 | 0.20 | 0.00 | 0.93 | 0.99 | 0.79 | 0.13 | 0.34 | 0.03 | NA | NA | NA |

Patients Under 18
