## Supplementary - ICU and Haematology-Oncology Patients for "Rational Empiric Antibiotic Escalation Applied to Specific Patient Groups" (from another version of this preprint: version 2, 2023-11-07)

**Supplementary Material**

**Antibiotic resistance rate over time**

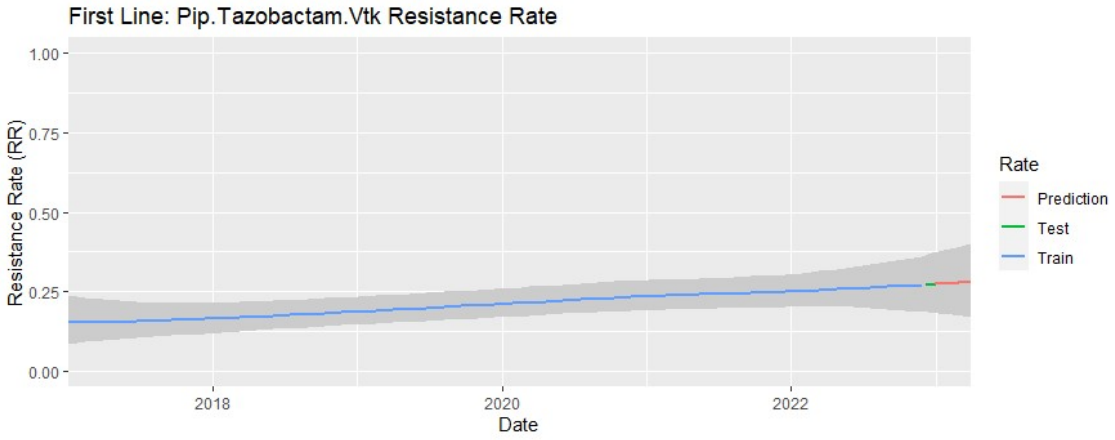


[1] "2017-01-01 Mid 0.151 (95%CI 0.085 to 0.239) Estimated Error 0.0383"

[1] "2022-12-06 Mid 0.271 (95%CI 0.184 to 0.365) Estimated Error 0.0456"

[1] "2022-12-29 Mid 0.273 (95%CI 0.181 to 0.372) Estimated Error 0.0478"

"2.9% posterior probability of decrease "

ICU patients

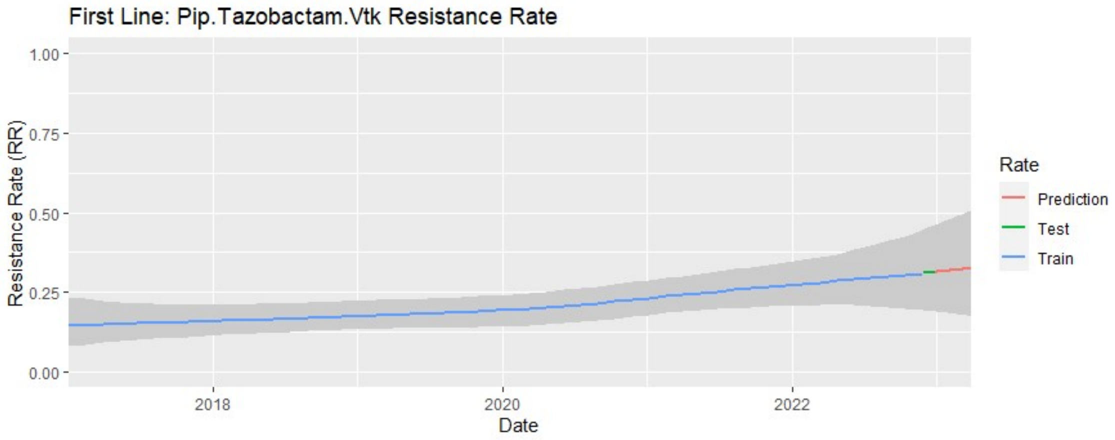


[1] "2017-01-02 Mid 0.145 (95%CI 0.081 to 0.235) Estimated Error 0.0394"

[1] "2022-11-30 Mid 0.310 (95%CI 0.192 to 0.447) Estimated Error 0.0644"

[1] "2022-12-28 Mid 0.313 (95%CI 0.188 to 0.459) Estimated Error 0.0683"

[1] "1.5% posterior probability of decrease "

Haematology/oncology patients

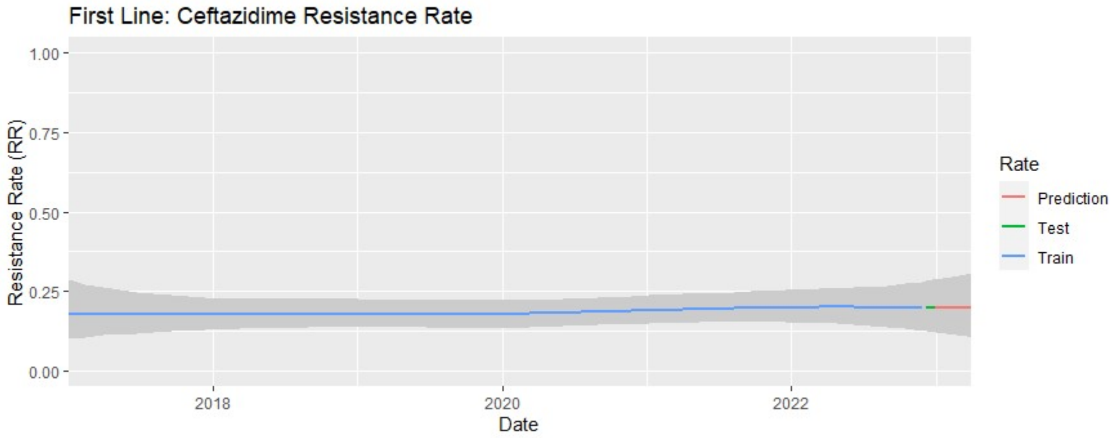


[1] "2017-01-01 Mid 0.179 (95%CI 0.101 to 0.285) Estimated Error 0.0462"

[1] "2022-12-06 Mid 0.199 (95%CI 0.123 to 0.282) Estimated Error 0.0403"

[1] "2022-12-29 Mid 0.199 (95%CI 0.119 to 0.286) Estimated Error 0.0421"

"35.6% posterior probability of decrease "

ICU patients

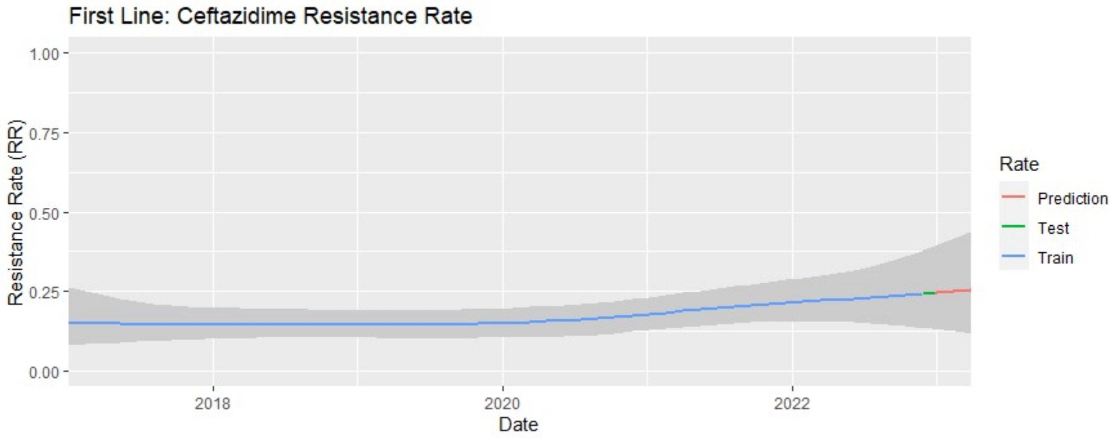


[1] "2017-01-02 Mid 0.152 (95%CI 0.078 to 0.264) Estimated Error 0.0479"

[1] "2022-11-30 Mid 0.242 (95%CI 0.134 to 0.379) Estimated Error 0.0615"

[1] "2022-12-28 Mid 0.244 (95%CI 0.130 to 0.391) Estimated Error 0.0655"

"11.9% posterior probability of decrease "

Haematology/oncology patients

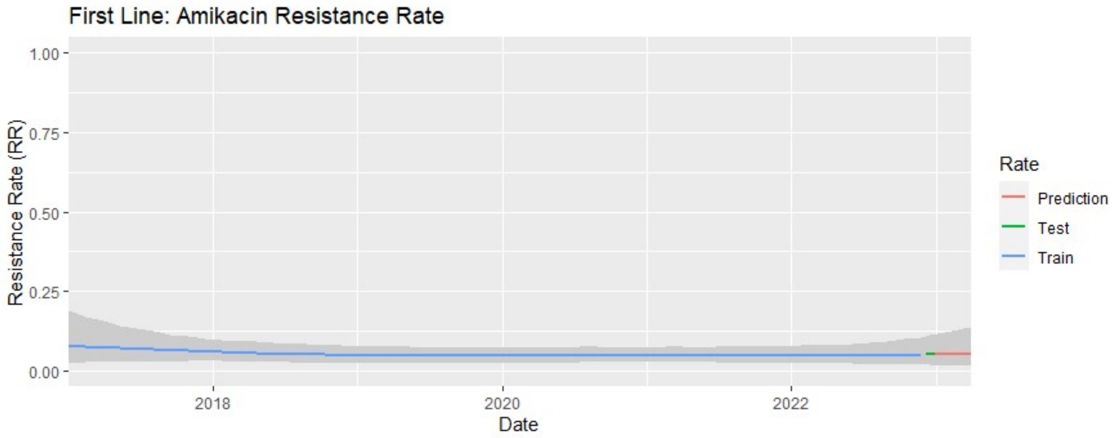


[1] "2017-01-01 Mid 0.080 (95%CI 0.024 to 0.189) Estimated Error 0.0422"

[1] "2022-12-06 Mid 0.050 (95%CI 0.017 to 0.108) Estimated Error 0.0237"

[1] "2022-12-29 Mid 0.051 (95%CI 0.016 to 0.113) Estimated Error 0.0251"

"73.2% posterior probability of decrease "

ICU patients

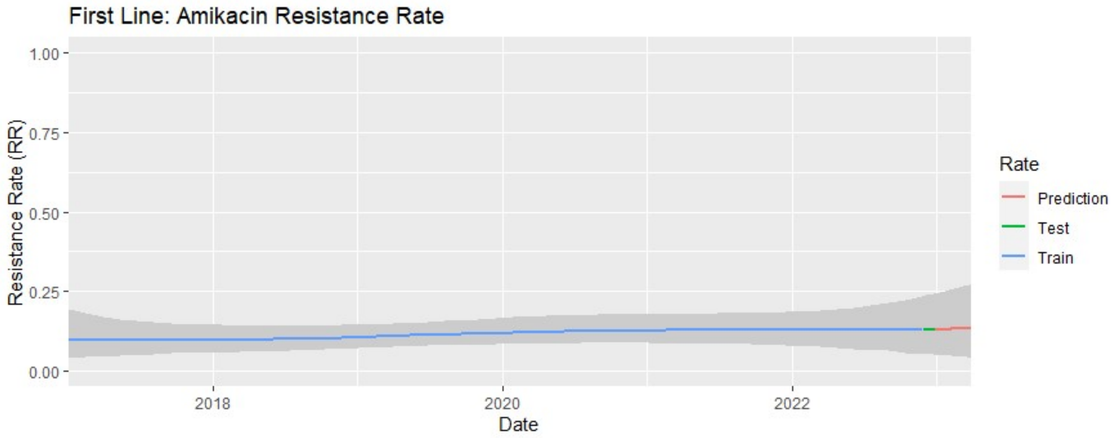


[1] "2017-01-02 Mid 0.099 (95%CI 0.039 to 0.194) Estimated Error 0.0389"

[1] "2022-11-30 Mid 0.129 (95%CI 0.052 to 0.234) Estimated Error 0.0459"

[1] "2022-12-28 Mid 0.130 (95%CI 0.049 to 0.241) Estimated Error 0.0484"

"30.0% posterior probability of decrease "

Haematology/oncology patients

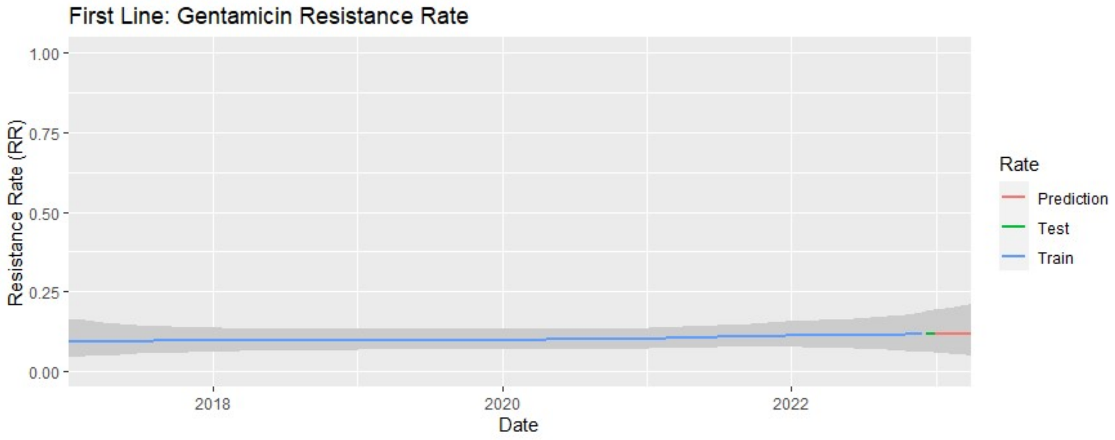


[1] "2017-01-01 Mid 0.094 (95%CI 0.042 to 0.166) Estimated Error 0.0316"

[1] "2022-12-06 Mid 0.115 (95%CI 0.059 to 0.188) Estimated Error 0.0323"

[1] "2022-12-29 Mid 0.115 (95%CI 0.057 to 0.192) Estimated Error 0.0338"

"31.6% posterior probability of decrease "

ICU patients

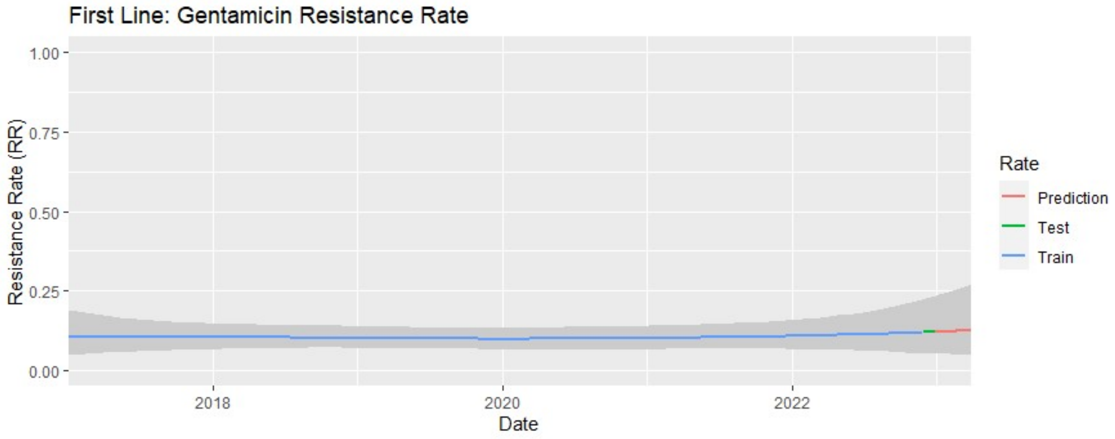


[1] "2017-01-02 Mid 0.106 (95%CI 0.049 to 0.188) Estimated Error 0.0355"

[1] "2022-11-30 Mid 0.119 (95%CI 0.053 to 0.222) Estimated Error 0.0424"

[1] "2022-12-28 Mid 0.120 (95%CI 0.052 to 0.232) Estimated Error 0.0453"

"41.4% posterior probability of decrease "

Haematology/oncology patients

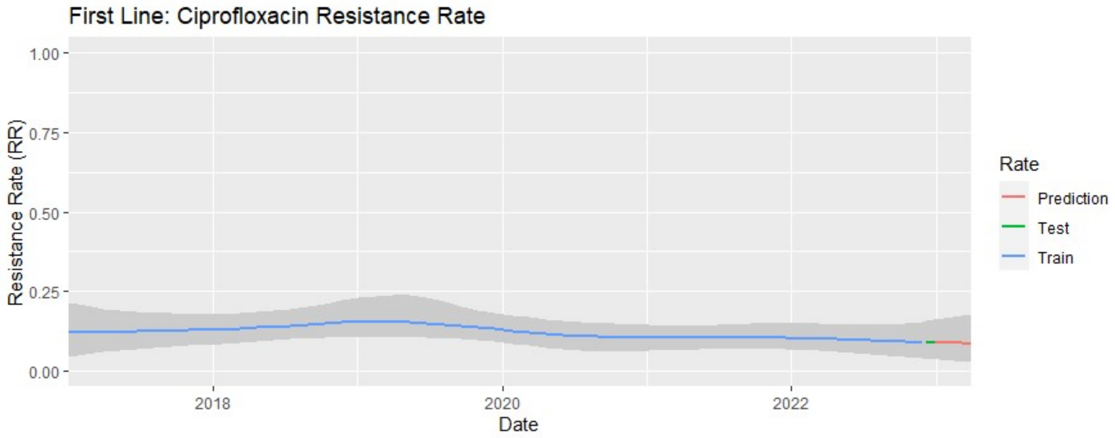


[1] "2017-01-01 Mid 0.121 (95%CI 0.045 to 0.215) Estimated Error 0.0434"

[1] "2022-12-06 Mid 0.089 (95%CI 0.037 to 0.156) Estimated Error 0.0306"

[1] "2022-12-29 Mid 0.088 (95%CI 0.035 to 0.160) Estimated Error 0.0321"

"73.6% posterior probability of decrease "

ICU patients

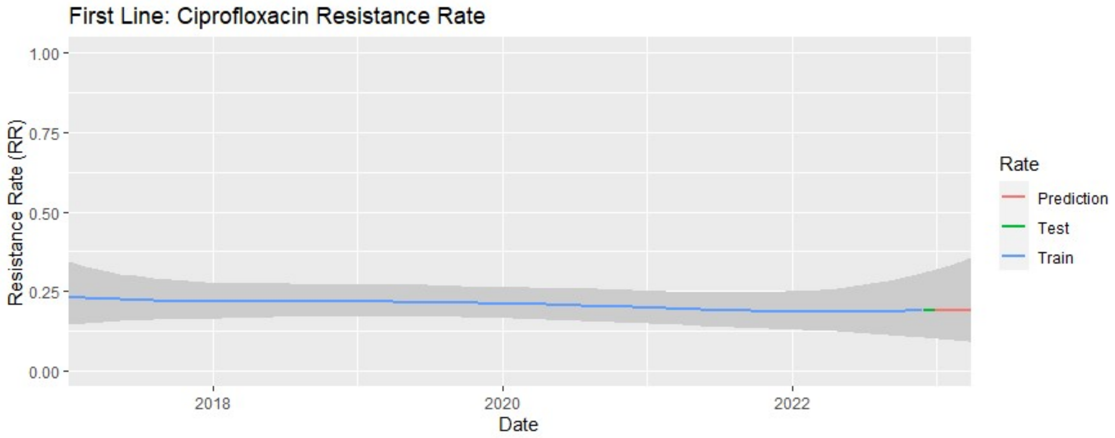


[1] "2017-01-02 Mid 0.234 (95%CI 0.145 to 0.346) Estimated Error 0.0508"

[1] "2022-11-30 Mid 0.188 (95%CI 0.103 to 0.307) Estimated Error 0.0522"

[1] "2022-12-28 Mid 0.189 (95%CI 0.099 to 0.316) Estimated Error 0.0556"

[ "74.6% posterior probability of decrease "

Haematology/oncology patients

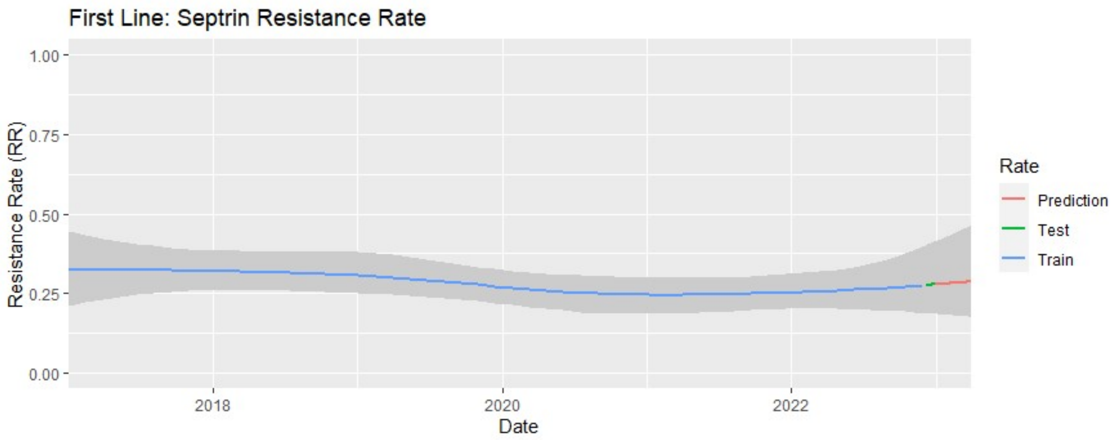


[1] "2017-01-01 Mid 0.324 (95%CI 0.209 to 0.446) Estimated Error 0.0599"

[1] "2022-12-06 Mid 0.276 (95%CI 0.186 to 0.400) Estimated Error 0.0543"

[1] "2022-12-29 Mid 0.278 (95%CI 0.184 to 0.412) Estimated Error 0.0580"

"73.8% posterior probability of decrease "

ICU patients

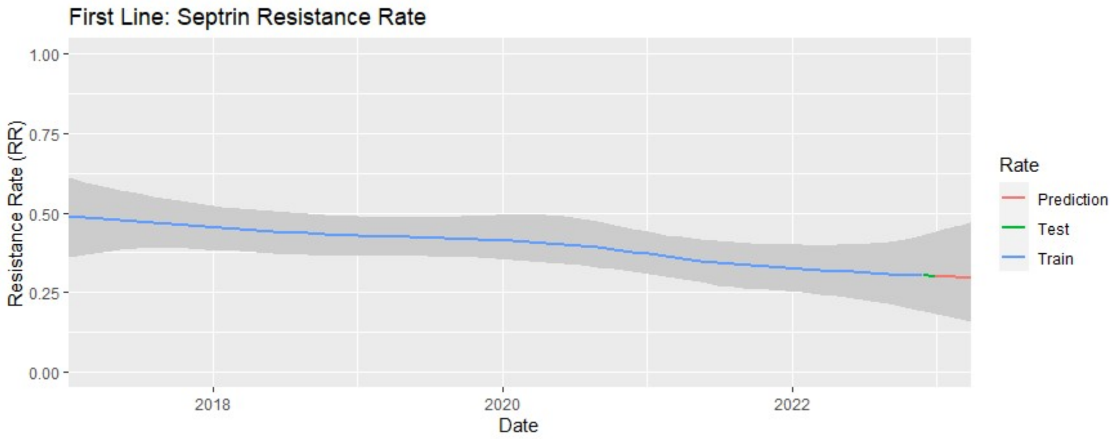


[1] "2017-01-02 Mid 0.487 (95%CI 0.359 to 0.610) Estimated Error 0.0626"

[1] "2022-11-30 Mid 0.302 (95%CI 0.188 to 0.430) Estimated Error 0.0603"

[1] "2022-12-28 Mid 0.300 (95%CI 0.181 to 0.438) Estimated Error 0.0638"

"97.9% posterior probability of decrease "

Haematology/oncology patients

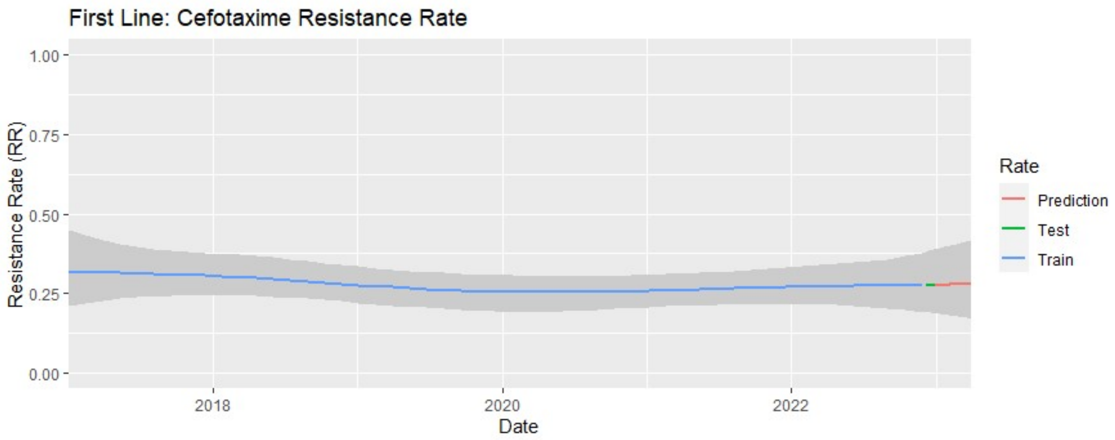


[1] "2017-01-01 Mid 0.319 (95%CI 0.210 to 0.448) Estimated Error 0.0592"

[1] "2022-12-06 Mid 0.276 (95%CI 0.190 to 0.381) Estimated Error 0.0485"

[1] "2022-12-29 Mid 0.276 (95%CI 0.186 to 0.387) Estimated Error 0.0509"

"72.2% posterior probability of decrease "

ICU patients

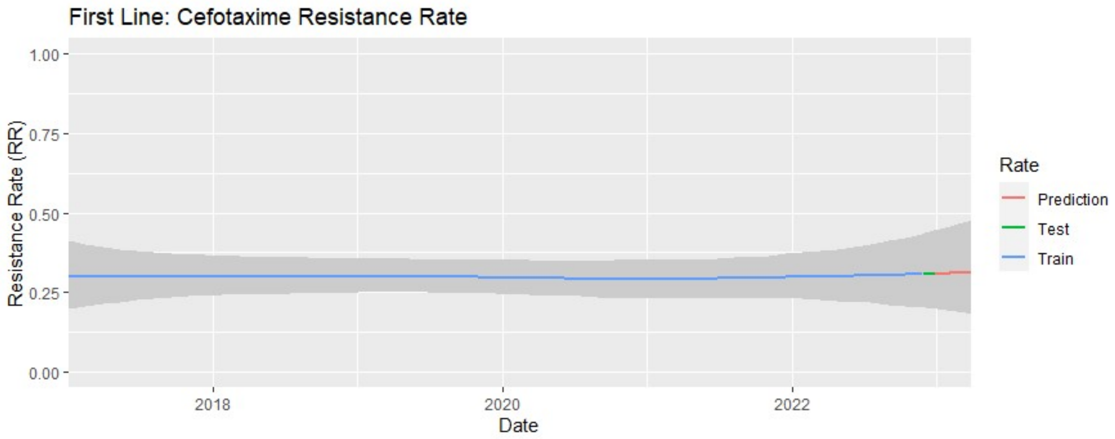


[1] "2017-01-02 Mid 0.298 (95%CI 0.196 to 0.411) Estimated Error 0.0538"

[1] "2022-11-30 Mid 0.307 (95%CI 0.200 to 0.435) Estimated Error 0.0597"

[1] "2022-12-28 Mid 0.308 (95%CI 0.196 to 0.444) Estimated Error 0.0628"

"46.3% posterior probability of decrease "

Haematology/oncology patients

**Antibiotic resistance rate over time assuming pip/taz resistance**

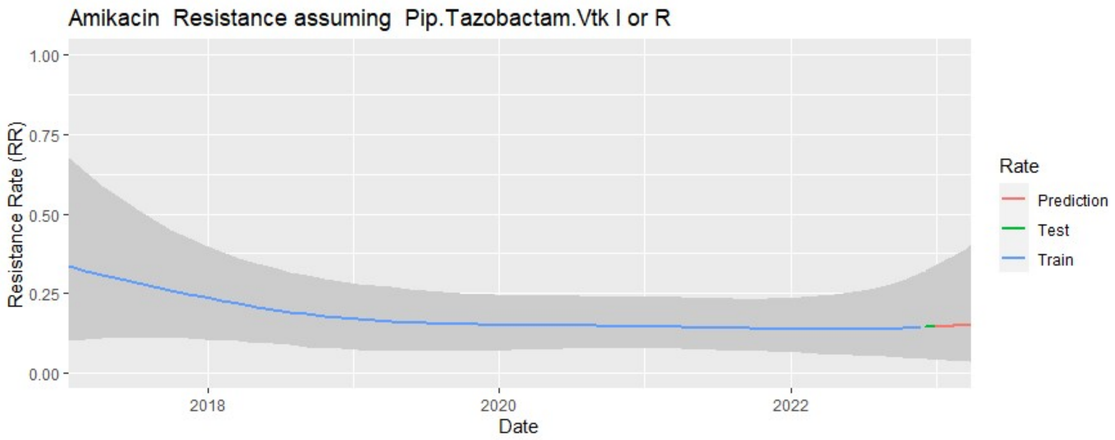


1] "2017-01-16 Mid 0.334 (95%CI 0.101 to 0.678) Estimated Error 0.1503"

[1] "2022-12-06 Mid 0.143 (95%CI 0.042 to 0.321) Estimated Error 0.0717"

[1] "2022-12-29 Mid 0.144 (95%CI 0.040 to 0.335) Estimated Error 0.0758"

"88.6% posterior probability of decrease "

ICU patients

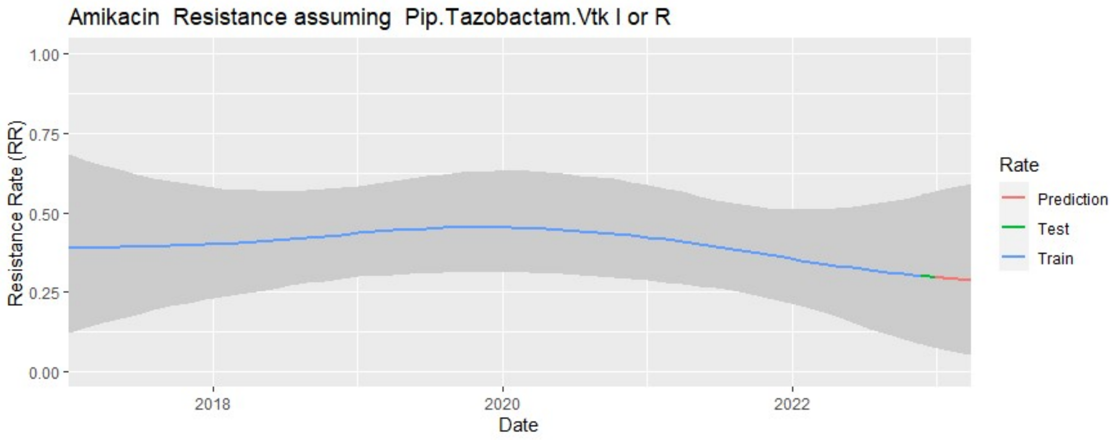


[1] "2017-01-02 Mid 0.389 (95%CI 0.121 to 0.685) Estimated Error 0.1475"

[1] "2022-11-23 Mid 0.300 (95%CI 0.084 to 0.556) Estimated Error 0.1216"

[1] "2022-12-28 Mid 0.296 (95%CI 0.072 to 0.565) Estimated Error 0.1276"

"68.1% posterior probability of decrease "

Haematology/oncology patients

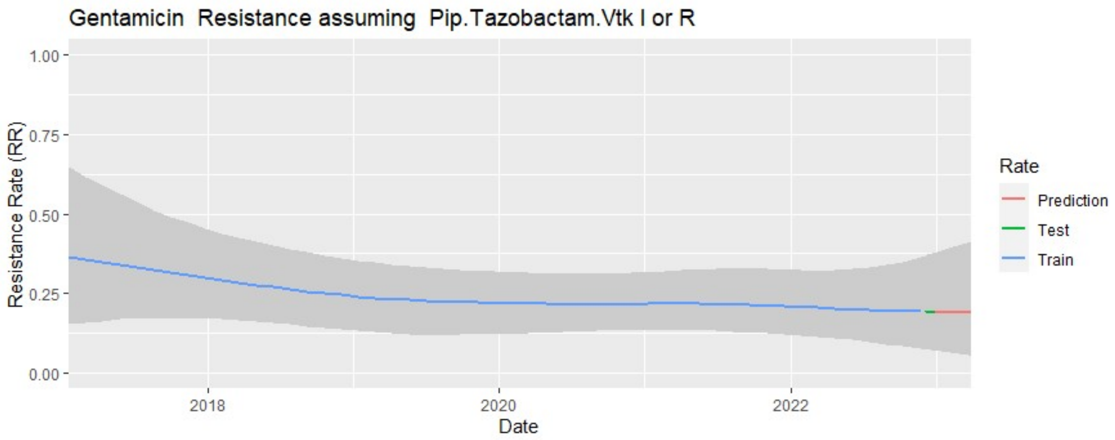


[1] "2017-01-16 Mid 0.366 (95%CI 0.153 to 0.649) Estimated Error 0.1258"

[1] "2022-12-06 Mid 0.192 (95%CI 0.072 to 0.368) Estimated Error 0.0757"

[1] "2022-12-29 Mid 0.191 (95%CI 0.068 to 0.377) Estimated Error 0.0787"

"88.5% posterior probability of decrease "

ICU patients

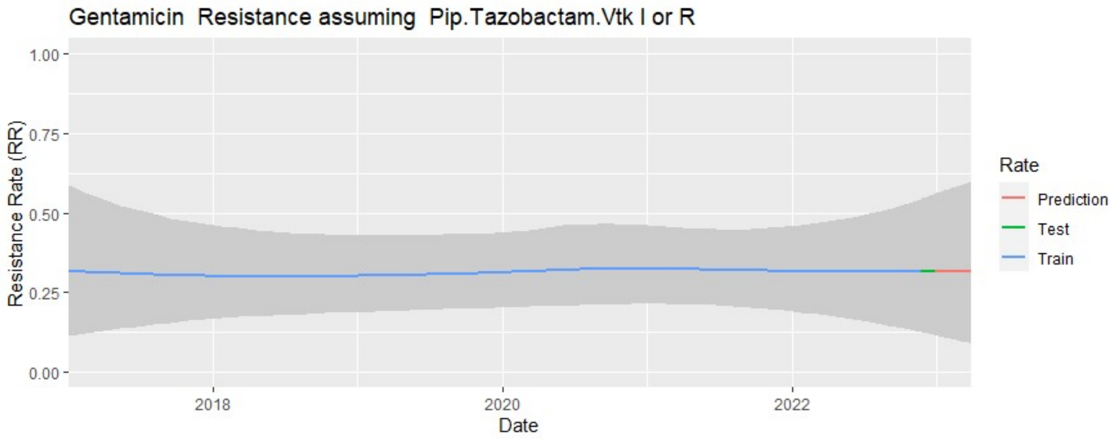


[1] "2017-01-02 Mid 0.318 (95%CI 0.113 to 0.588) Estimated Error 0.1225"

[1] "2022-11-23 Mid 0.314 (95%CI 0.123 to 0.544) Estimated Error 0.1098"

[1] "2022-12-28 Mid 0.315 (95%CI 0.114 to 0.559) Estimated Error 0.1161"

"49.6% posterior probability of decrease "

Haematology/oncology patients

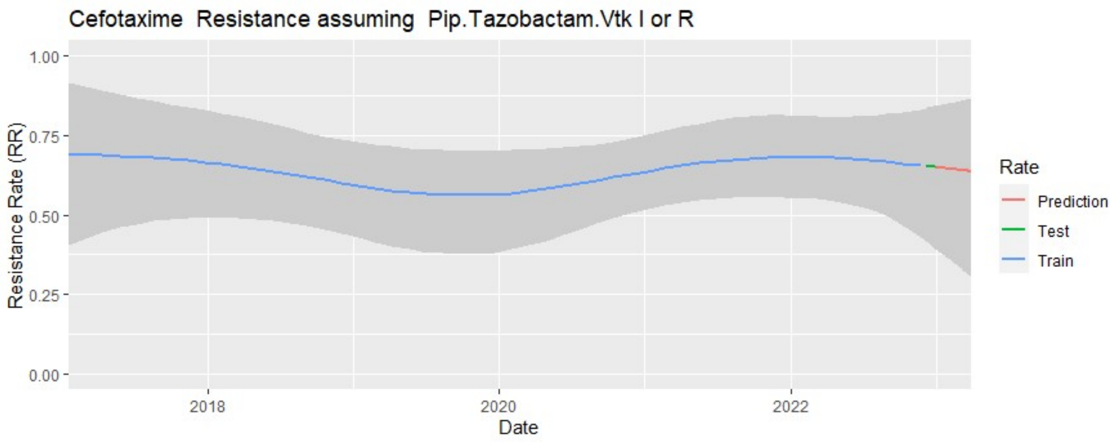


[1] "2017-01-16 Mid 0.690 (95%CI 0.404 to 0.915) Estimated Error 0.1333"

[1] "2022-12-06 Mid 0.654 (95%CI 0.417 to 0.836) Estimated Error 0.1062"

[1] "2022-12-29 Mid 0.650 (95%CI 0.393 to 0.842) Estimated Error 0.1130"

"58.0% posterior probability of decrease "

ICU patients

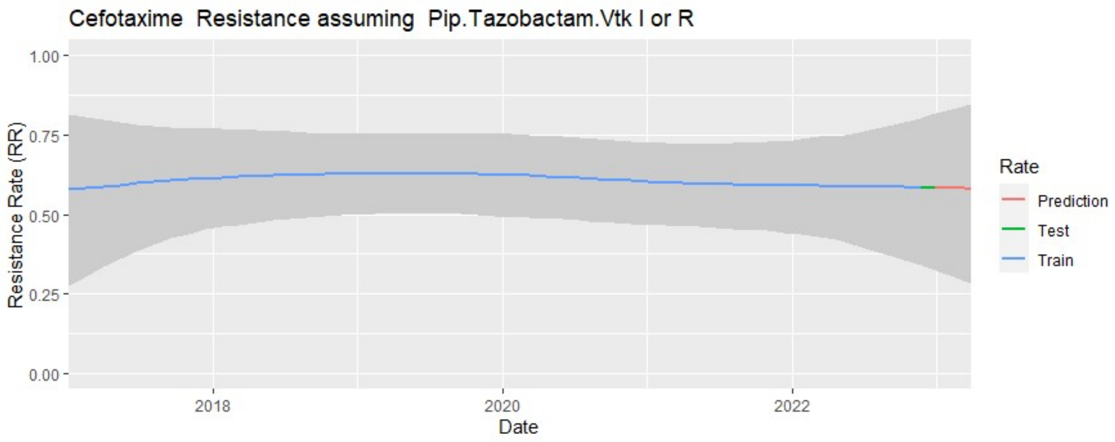


[1] "2017-01-02 Mid 0.579 (95%CI 0.275 to 0.815) Estimated Error 0.1387"

[1] "2022-11-23 Mid 0.584 (95%CI 0.339 to 0.804) Estimated Error 0.1186"

[1] "2022-12-28 Mid 0.584 (95%CI 0.323 to 0.816) Estimated Error 0.1254"

"50.4% posterior probability of decrease "

Haematology/oncology patients

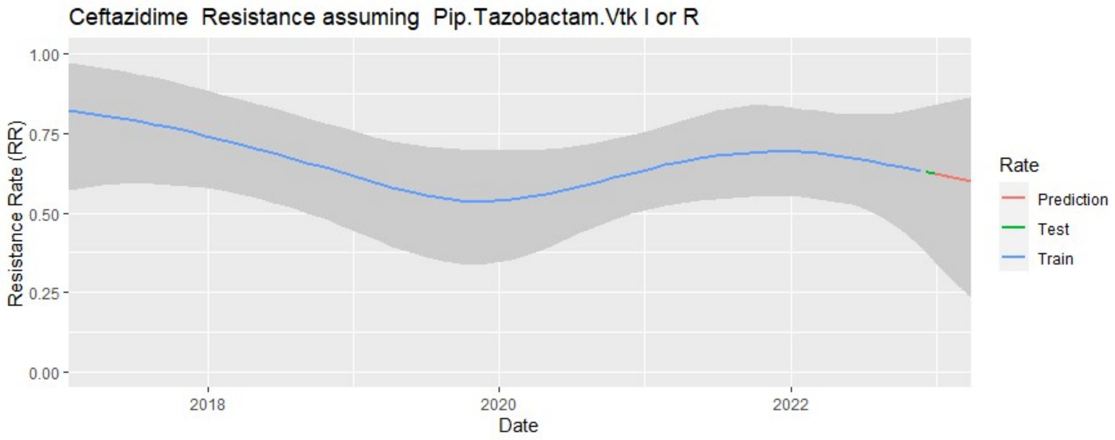


[1] "2017-01-16 Mid 0.822 (95%CI 0.572 to 0.974) Estimated Error 0.1083"

[1] "2022-12-06 Mid 0.628 (95%CI 0.375 to 0.833) Estimated Error 0.1175"

[1] "2022-12-29 Mid 0.622 (95%CI 0.345 to 0.839) Estimated Error 0.1264"

"88.8% posterior probability of decrease "

ICU patients

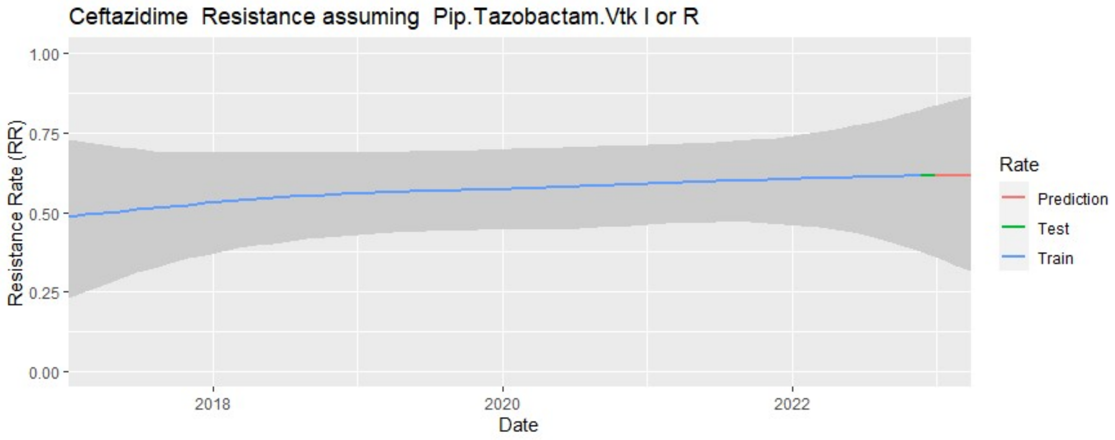


"2017-01-02 Mid 0.487 (95%CI 0.231 to 0.730) Estimated Error 0.1277"

[1] "2022-11-23 Mid 0.615 (95%CI 0.374 to 0.823) Estimated Error 0.1140"

[1] "2022-12-28 Mid 0.616 (95%CI 0.359 to 0.835) Estimated Error 0.1206"

"23.6% posterior probability of decrease "

Haematology/oncology patients

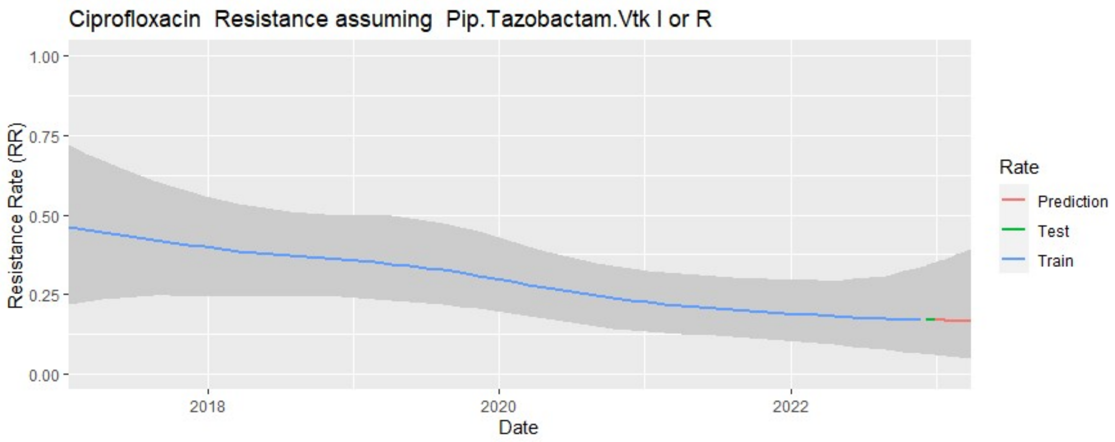


[1] "2017-01-16 Mid 0.463 (95%CI 0.217 to 0.721) Estimated Error 0.1283"

[1] "2022-12-06 Mid 0.169 (95%CI 0.061 to 0.341) Estimated Error 0.0715"

[1] "2022-12-29 Mid 0.168 (95%CI 0.058 to 0.349) Estimated Error 0.0745"

"97.5% posterior probability of decrease "

ICU patients

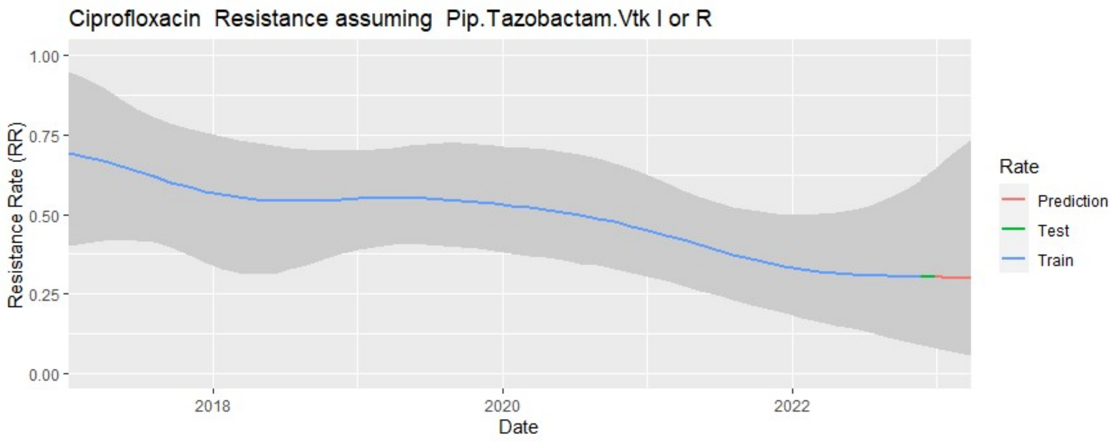


[1] "2017-01-02 Mid 0.694 (95%CI 0.399 to 0.950) Estimated Error 0.1421"

[1] "2022-11-23 Mid 0.302 (95%CI 0.087 to 0.610) Estimated Error 0.1342"

[1] "2022-12-28 Mid 0.302 (95%CI 0.077 to 0.642) Estimated Error 0.1443"

"96.9% posterior probability of decrease "

Haematology/oncology patients

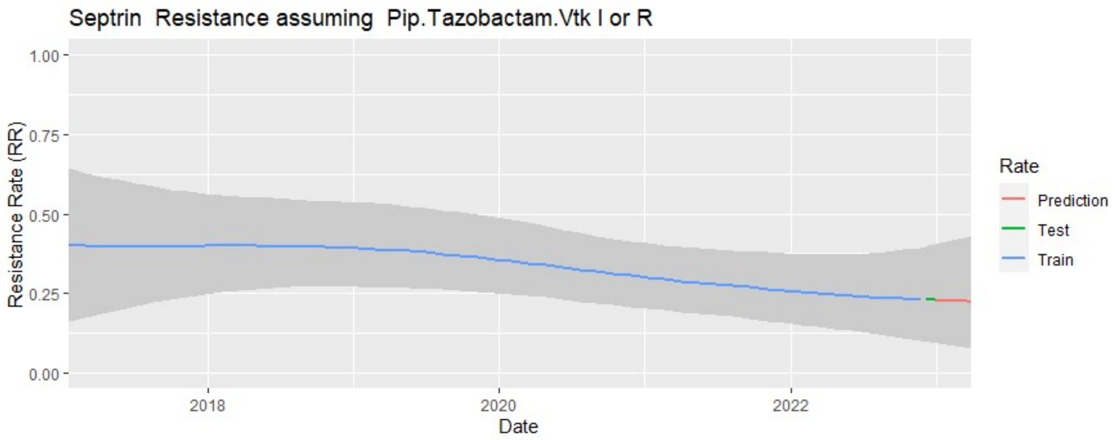


[1] "2017-01-16 Mid 0.399 (95%CI 0.160 to 0.643) Estimated Error 0.1261"

[1] "2022-12-06 Mid 0.229 (95%CI 0.096 to 0.397) Estimated Error 0.0776"

[1] "2022-12-29 Mid 0.228 (95%CI 0.092 to 0.403) Estimated Error 0.0802"

[1] "85.9% posterior probability of decrease "

ICU patients

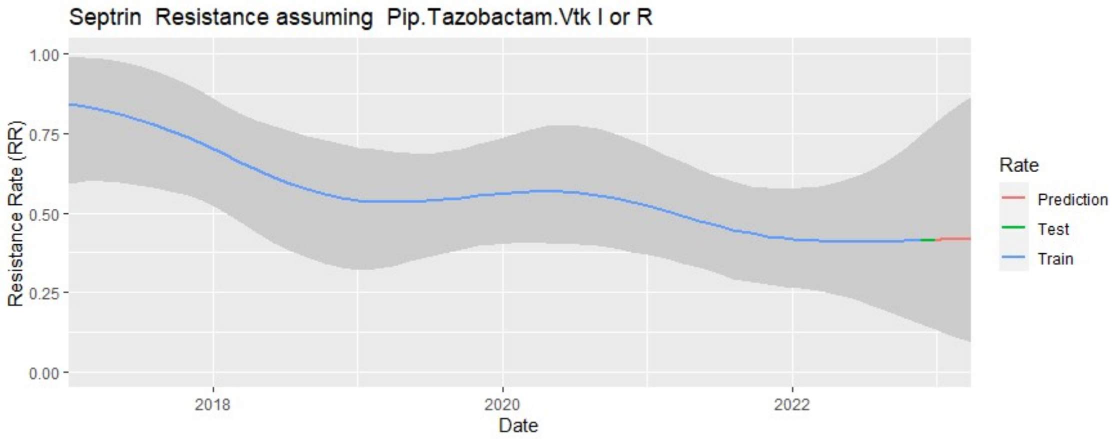


[1] "2017-01-02 Mid 0.845 (95%CI 0.592 to 0.994) Estimated Error 0.1113"

[1] "2022-11-23 Mid 0.413 (95%CI 0.149 to 0.745) Estimated Error 0.1543"

[1] "2022-12-28 Mid 0.414 (95%CI 0.132 to 0.781) Estimated Error 0.1677"

"99.0% posterior probability of decrease "

Haematology/oncology patients

**Antibiotic resistance rate over time assuming ceftazidime resistance**

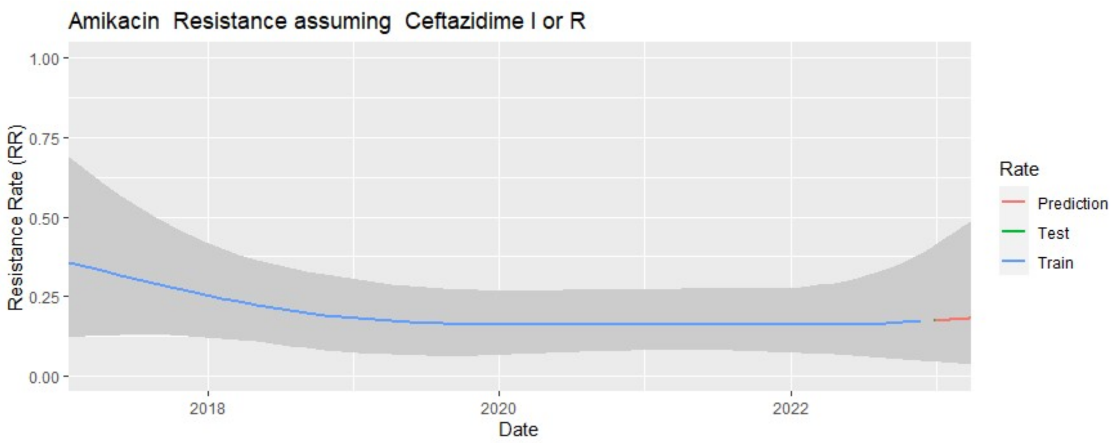


[1] "2017-01-16 Mid 0.355 (95%CI 0.121 to 0.689) Estimated Error 0.1485"

[1] "2022-12-26 Mid 0.172 (95%CI 0.043 to 0.409) Estimated Error 0.0940"

[1] "2022-12-29 Mid 0.172 (95%CI 0.043 to 0.411) Estimated Error 0.0946"

"85.9% posterior probability of decrease "

ICU patients

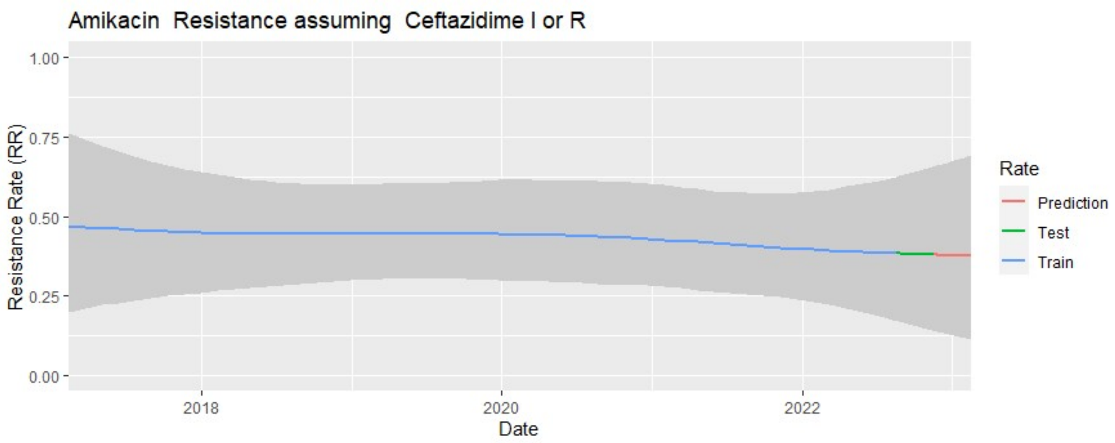


[1] "2017-02-11 Mid 0.470 (95%CI 0.198 to 0.761) Estimated Error 0.1445"

[1] "2022-08-17 Mid 0.383 (95%CI 0.170 to 0.624) Estimated Error 0.1177"

[1] "2022-11-16 Mid 0.379 (95%CI 0.138 to 0.656) Estimated Error 0.1340"

"66.9% posterior probability of decrease "

Haematology/oncology patients

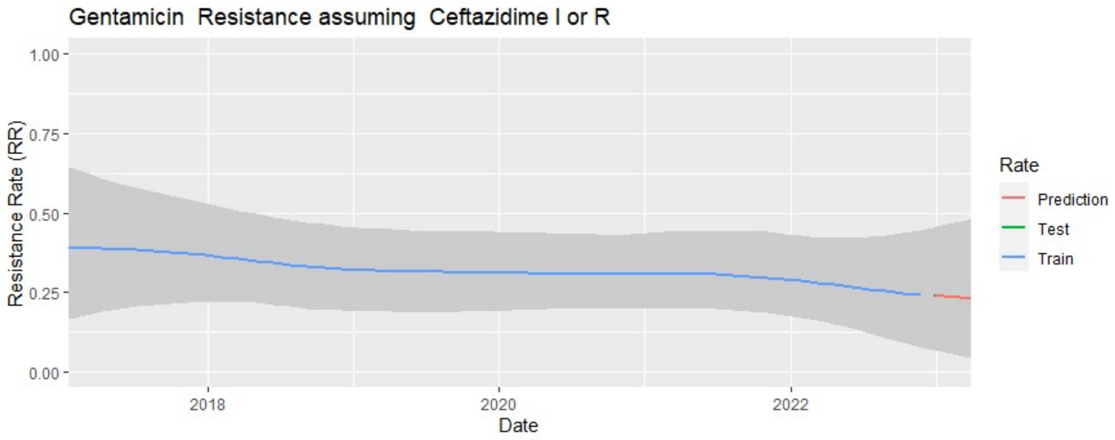


[1] "2017-01-16 Mid 0.392 (95%CI 0.165 to 0.646) Estimated Error 0.1236"

[1] "2022-12-26 Mid 0.239 (95%CI 0.067 to 0.454) Estimated Error 0.0989"

[1] "2022-12-29 Mid 0.239 (95%CI 0.066 to 0.455) Estimated Error 0.0993"

"82.3% posterior probability of decrease "

ICU patients

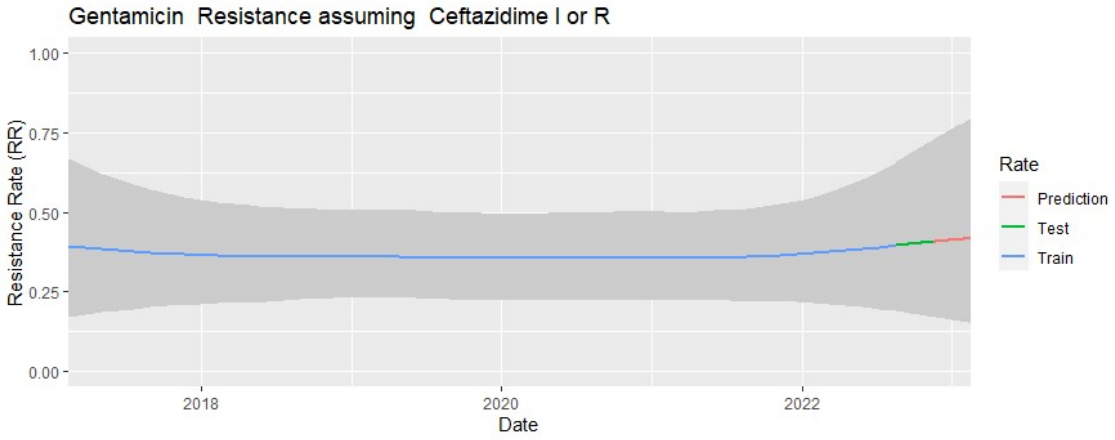


[1] "2017-02-11 Mid 0.393 (95%CI 0.169 to 0.667) Estimated Error 0.1281"

[1] "2022-08-17 Mid 0.395 (95%CI 0.188 to 0.658) Estimated Error 0.1204"

[1] "2022-11-16 Mid 0.407 (95%CI 0.168 to 0.728) Estimated Error 0.1424"

"49.9% posterior probability of decrease "

Haematology/oncology patients

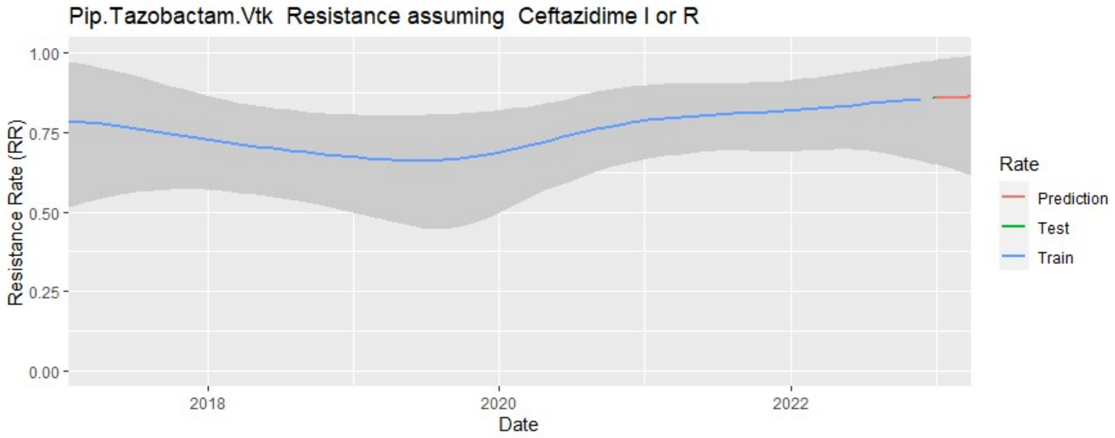


[1] "2017-01-16 Mid 0.787 (95%CI 0.513 to 0.973) Estimated Error 0.1211"

[1] "2022-12-26 Mid 0.857 (95%CI 0.650 to 0.978) Estimated Error 0.0859"

[1] "2022-12-29 Mid 0.858 (95%CI 0.649 to 0.978) Estimated Error 0.0864"

"31.0% posterior probability of decrease "

ICU patients

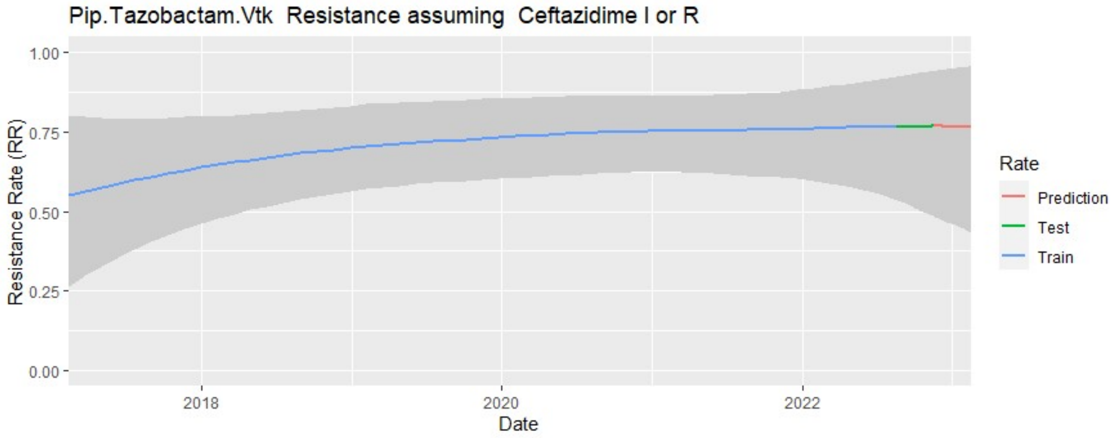


[1] "2017-02-11 Mid 0.552 (95%CI 0.263 to 0.801) Estimated Error 0.1389"

[1] "2022-08-17 Mid 0.768 (95%CI 0.535 to 0.923) Estimated Error 0.1008"

[1] "2022-11-16 Mid 0.768 (95%CI 0.484 to 0.941) Estimated Error 0.1173"

"11.0% posterior probability of decrease "

Haematology/oncology patients

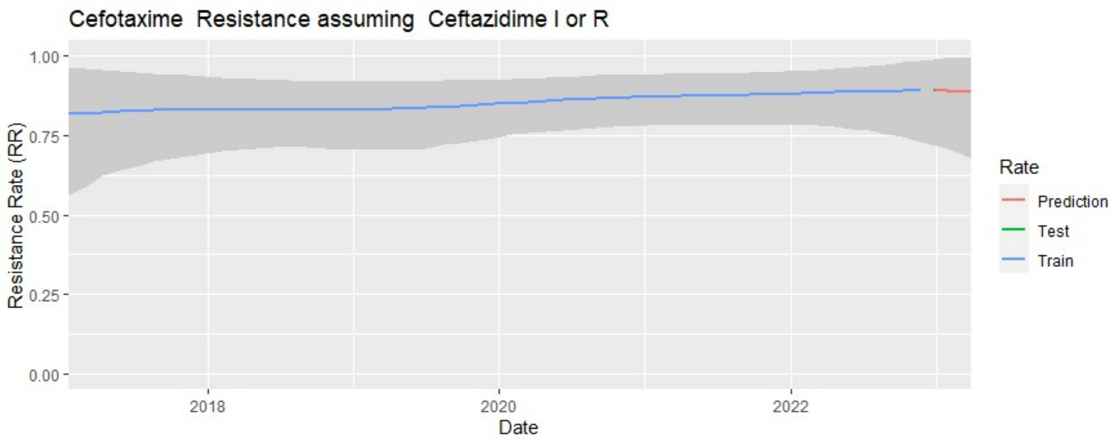


[1] "2017-01-16 Mid 0.817 (95%CI 0.562 to 0.965) Estimated Error 0.1031"

[1] "2022-12-26 Mid 0.890 (95%CI 0.718 to 0.990) Estimated Error 0.0692"

[1] "2022-12-29 Mid 0.890 (95%CI 0.717 to 0.990) Estimated Error 0.0696"

"28.9% posterior probability of decrease "

ICU patients

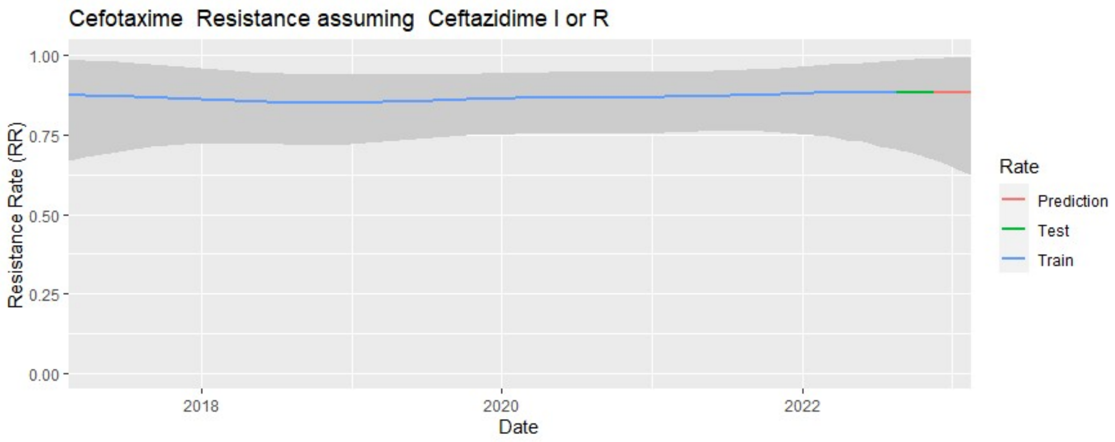


[1] "2017-02-11 Mid 0.875 (95%CI 0.670 to 0.989) Estimated Error 0.0844"

[1] "2022-08-17 Mid 0.885 (95%CI 0.704 to 0.985) Estimated Error 0.0736"

[1] "2022-11-16 Mid 0.884 (95%CI 0.671 to 0.990) Estimated Error 0.0851"

"47.4% posterior probability of decrease "

Haematology/oncology patients

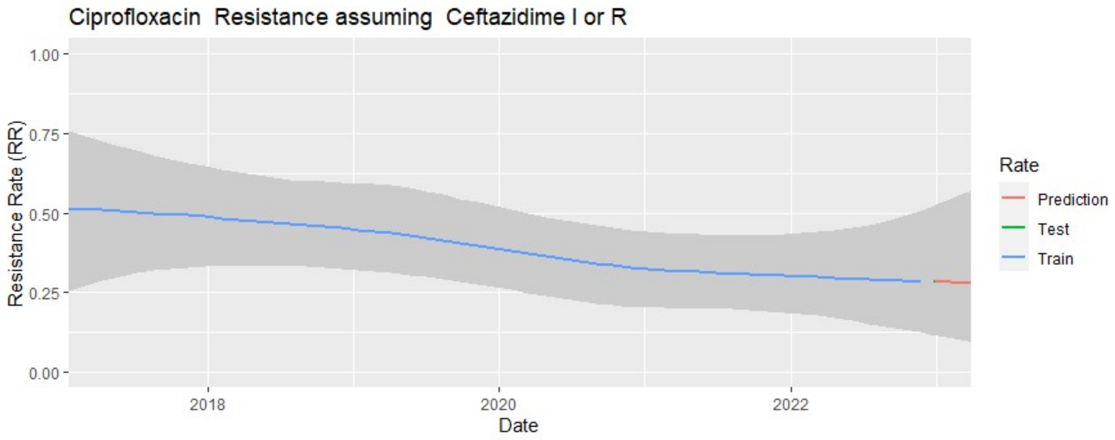


[1] "2017-01-16 Mid 0.515 (95%CI 0.254 to 0.757) Estimated Error 0.1281"

[1] "2022-12-26 Mid 0.282 (95%CI 0.114 to 0.522) Estimated Error 0.1025"

[1] "2022-12-29 Mid 0.282 (95%CI 0.113 to 0.523) Estimated Error 0.1030"

"90.8% posterior probability of decrease "
ICU patients

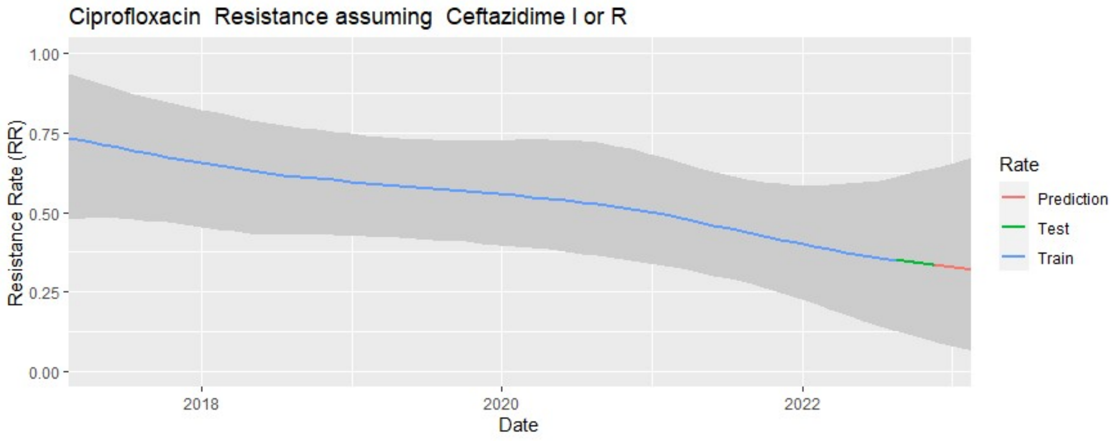


[1] "2017-02-11 Mid 0.733 (95%CI 0.478 to 0.935) Estimated Error 0.1188"

[1] "2022-08-17 Mid 0.350 (95%CI 0.125 to 0.611) Estimated Error 0.1243"

[1] "2022-11-16 Mid 0.333 (95%CI 0.092 to 0.637) Estimated Error 0.1399"

"97.4% posterior probability of decrease "Haematology/oncology patients

Haematology/oncology patients

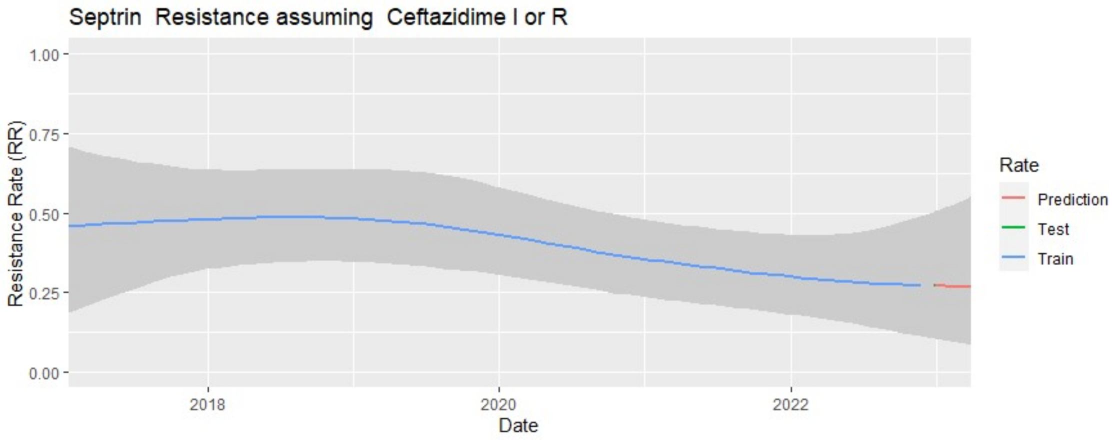


[1] "2017-01-16 Mid 0.459 (95%CI 0.186 to 0.707) Estimated Error 0.1352"

[1] "2022-12-26 Mid 0.270 (95%CI 0.103 to 0.502) Estimated Error 0.1019"

[1] "2022-12-29 Mid 0.270 (95%CI 0.103 to 0.504) Estimated Error 0.1024"

"85.7% posterior probability of decrease "

ICU patients

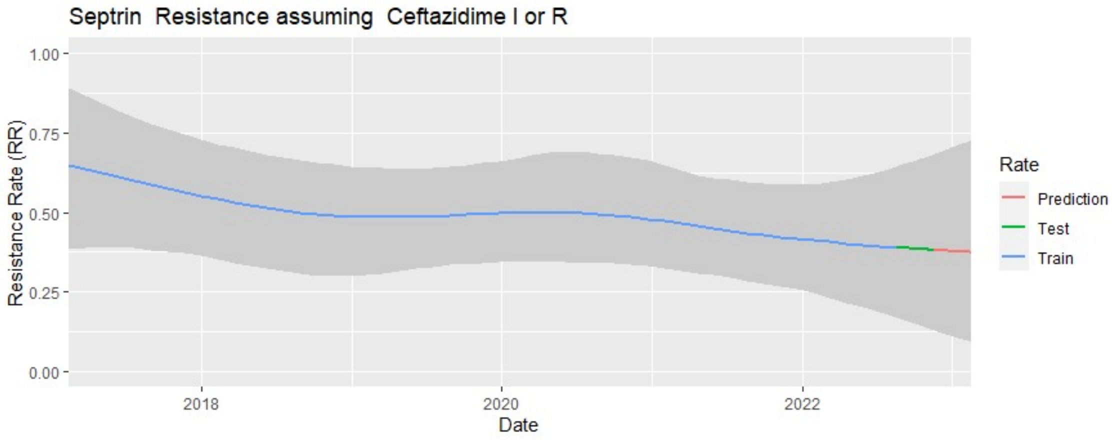


[1] "2017-02-11 Mid 0.648 (95%CI 0.383 to 0.892) Estimated Error 0.1331"

[1] "2022-08-17 Mid 0.390 (95%CI 0.168 to 0.645) Estimated Error 0.1213"

[1] "2022-11-16 Mid 0.381 (95%CI 0.127 to 0.681) Estimated Error 0.1411"

"90.4% posterior probability of decrease "

Haematology/oncology patients
