## Supplementary - Urine or GI source Patients for "Rational Empiric Antibiotic Escalation Applied to Specific Patient Groups" (from another version of this preprint: version 2, 2023-11-07)

**Supplementary Material**

**Antibiotic resistance rate over time**

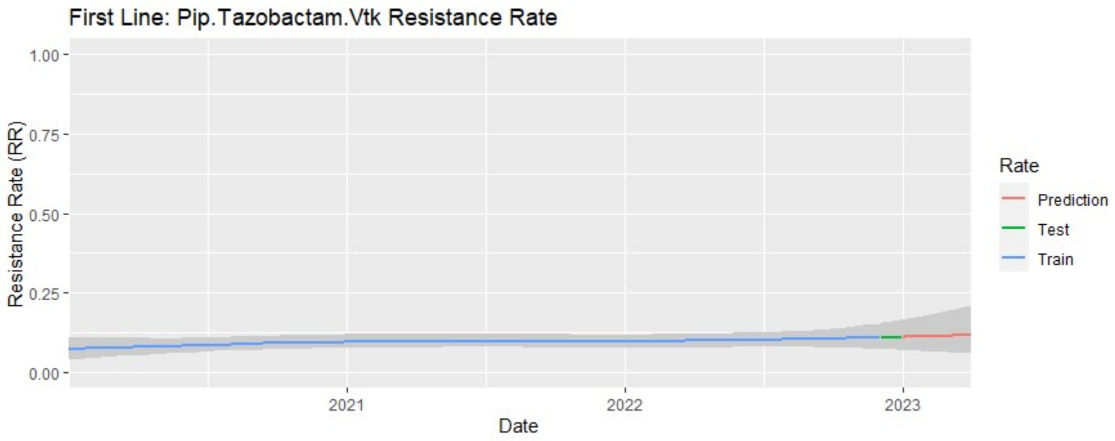


[1] "2020-01-01 Mid 0.073 (95%CI 0.039 to 0.110) Estimated Error 0.0179"

[1] "2022-12-03 Mid 0.109 (95%CI 0.072 to 0.154) Estimated Error 0.0208"

[1] "2022-12-30 Mid 0.110 (95%CI 0.069 to 0.164) Estimated Error 0.0239"

"9.1% posterior probability of decrease "

Urine/renal Source

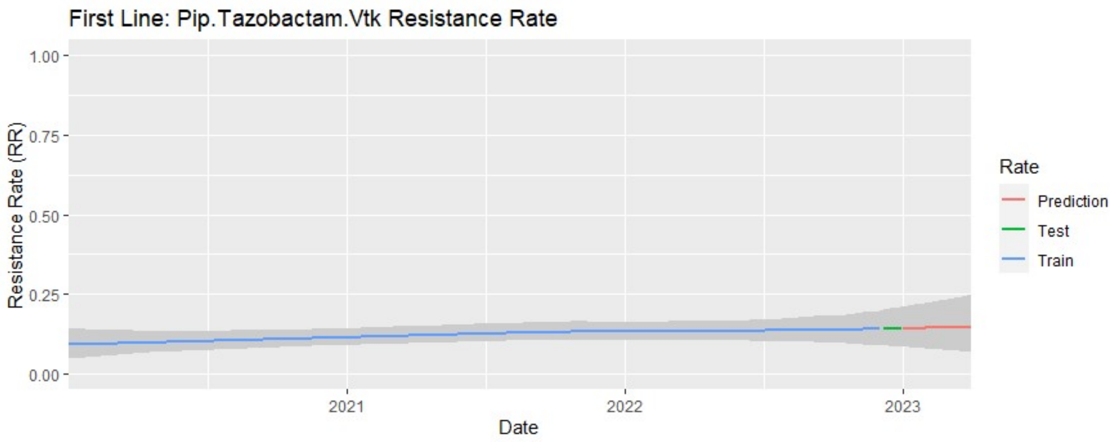


[1] "2020-01-02 Mid 0.092 (95%CI 0.049 to 0.143) Estimated Error 0.0238"

[1] "2022-12-06 Mid 0.140 (95%CI 0.087 to 0.201) Estimated Error 0.0288"

[1] "2022-12-31 Mid 0.141 (95%CI 0.083 to 0.209) Estimated Error 0.0320"

"9.6% posterior probability of decrease "

GI Source

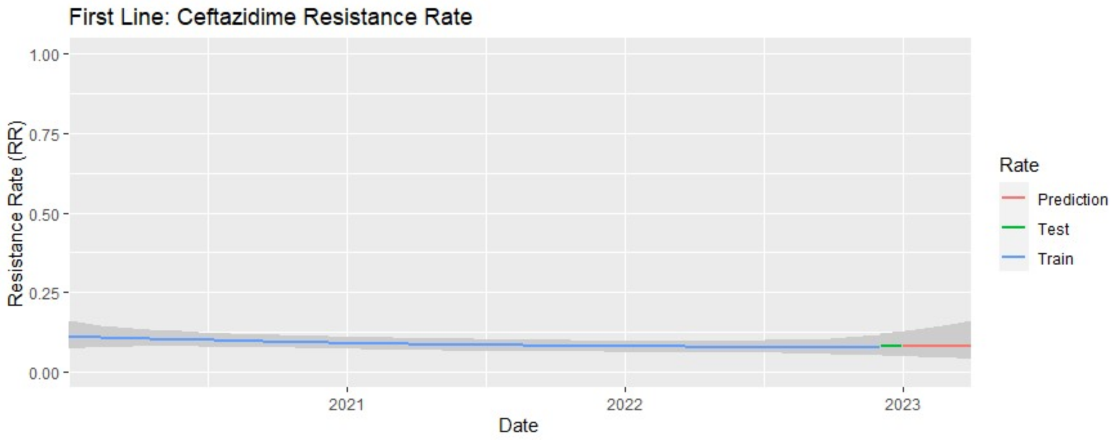


[1] "2020-01-01 Mid 0.111 (95%CI 0.073 to 0.160) Estimated Error 0.0221"

[1] "2022-12-03 Mid 0.078 (95%CI 0.049 to 0.119) Estimated Error 0.0172"

[1] "2022-12-30 Mid 0.078 (95%CI 0.047 to 0.126) Estimated Error 0.0196"

"88.5% posterior probability of decrease "

Urine/renal Source

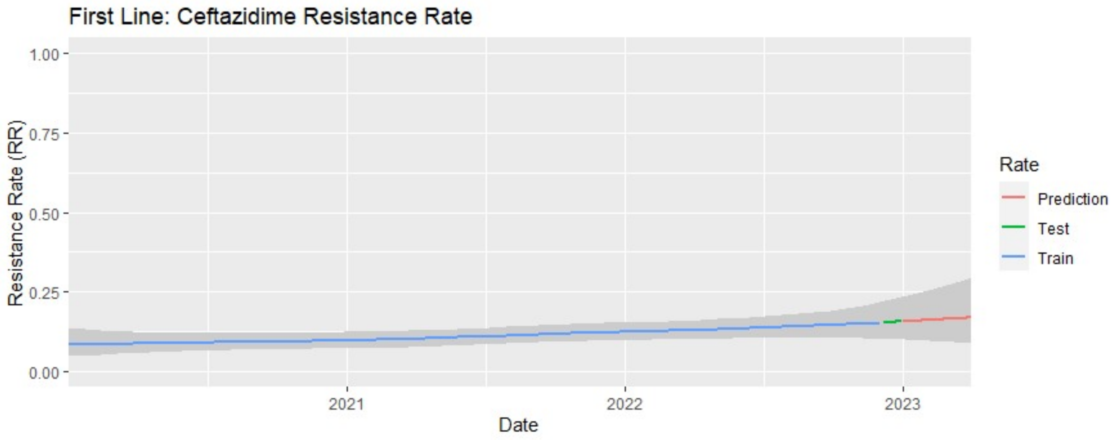


[1] "2020-01-02 Mid 0.083 (95%CI 0.046 to 0.135) Estimated Error 0.0220"

[1] "2022-12-06 Mid 0.153 (95%CI 0.101 to 0.219) Estimated Error 0.0299"

[1] "2022-12-31 Mid 0.156 (95%CI 0.099 to 0.232) Estimated Error 0.0338"

"3.1% posterior probability of decrease "

GI Source

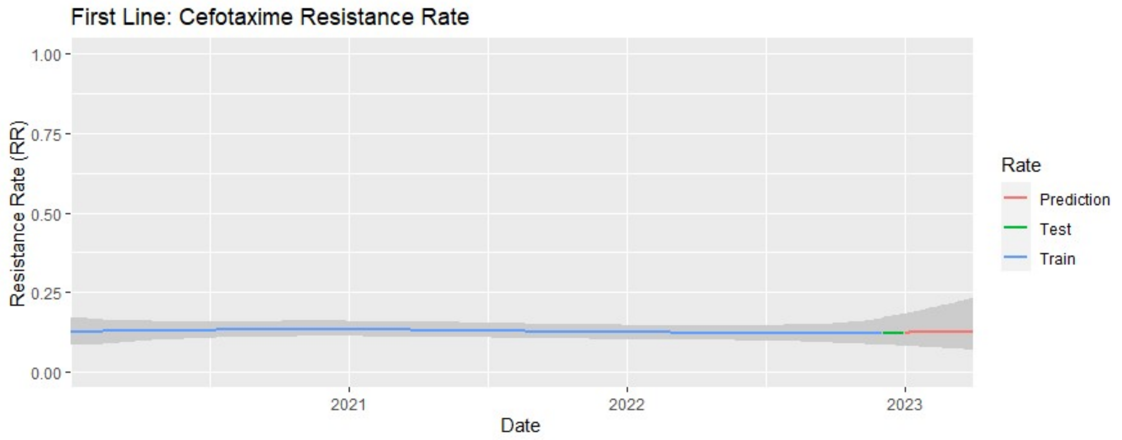


[1] "2020-01-01 Mid 0.126 (95%CI 0.082 to 0.172) Estimated Error 0.0223"

[1] "2022-12-03 Mid 0.122 (95%CI 0.084 to 0.171) Estimated Error 0.0216"

[1] "2022-12-30 Mid 0.122 (95%CI 0.080 to 0.181) Estimated Error 0.0250"

"56.9% posterior probability of decrease "

Urine/renal Source

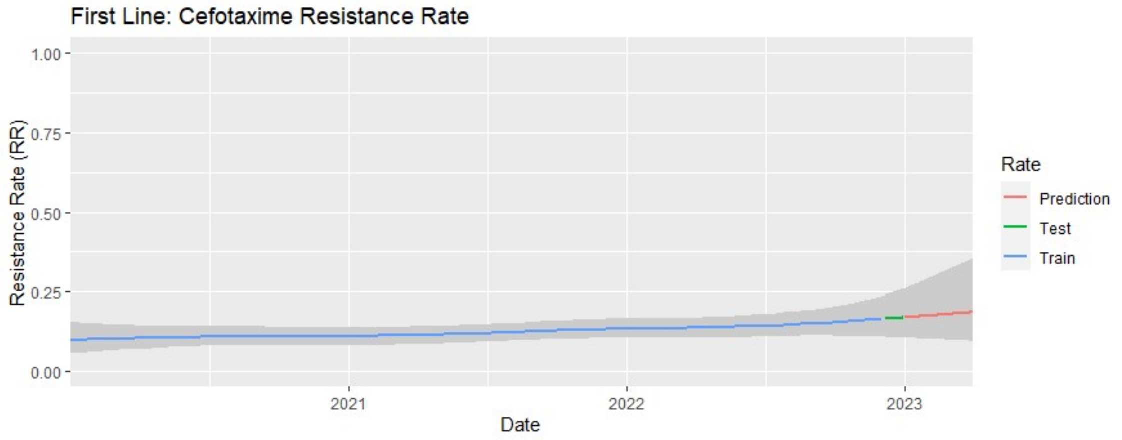


[1] "2020-01-02 Mid 0.098 (95%CI 0.054 to 0.155) Estimated Error 0.0254"

[1] "2022-12-06 Mid 0.164 (95%CI 0.106 to 0.240) Estimated Error 0.0333"

[1] "2022-12-31 Mid 0.168 (95%CI 0.104 to 0.259) Estimated Error 0.0387"

"5.4% posterior probability of decrease "GI Source

GI Source

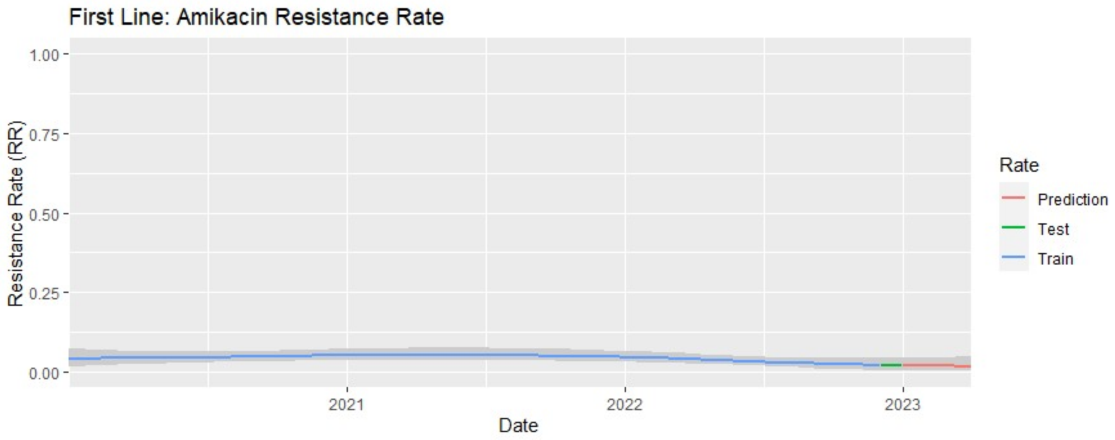


[1] "2020-01-01 Mid 0.041 (95%CI 0.016 to 0.077) Estimated Error 0.0158"

[1] "2022-12-03 Mid 0.020 (95%CI 0.004 to 0.043) Estimated Error 0.0101"

[1] "2022-12-30 Mid 0.019 (95%CI 0.003 to 0.043) Estimated Error 0.0107"

"89.9% posterior probability of decrease "

Urine/renal Source

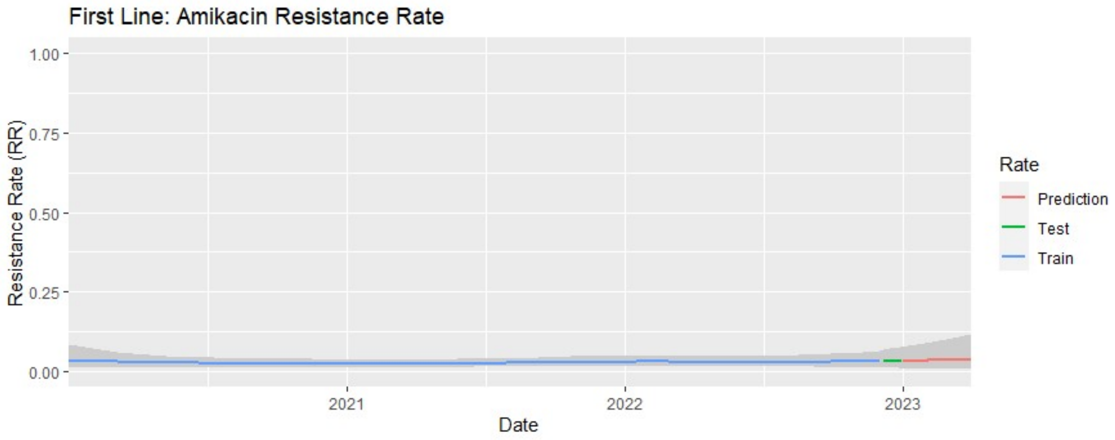


[1] "2020-01-02 Mid 0.034 (95%CI 0.011 to 0.084) Estimated Error 0.0187"

[1] "2022-12-06 Mid 0.031 (95%CI 0.009 to 0.067) Estimated Error 0.0150"

[1] "2022-12-31 Mid 0.032 (95%CI 0.009 to 0.075) Estimated Error 0.0174"

"53.5% posterior probability of decrease "

GI Source

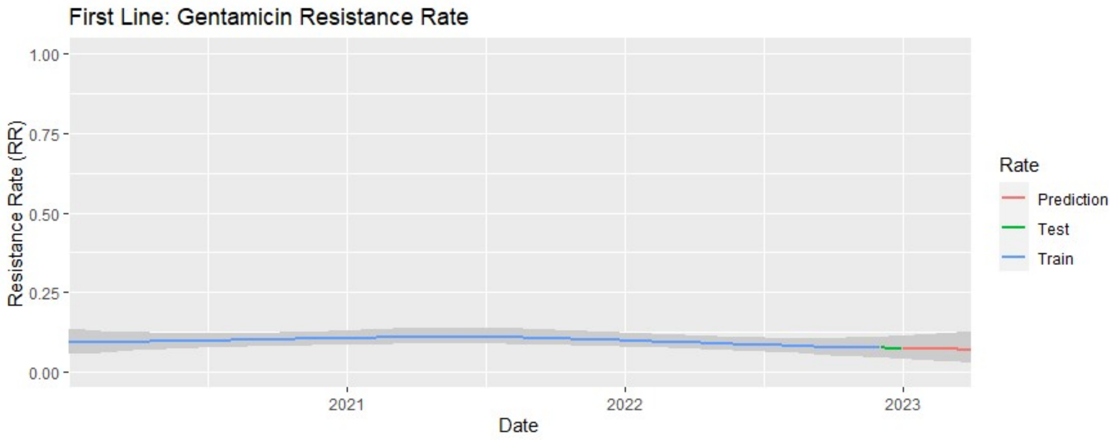


[1] "2020-01-01 Mid 0.093 (95%CI 0.054 to 0.137) Estimated Error 0.0211"

[1] "2022-12-03 Mid 0.074 (95%CI 0.043 to 0.109) Estimated Error 0.0173"

[1] "2022-12-30 Mid 0.073 (95%CI 0.039 to 0.112) Estimated Error 0.0191"

"76.0% posterior probability of decrease "

Urine/renal Source

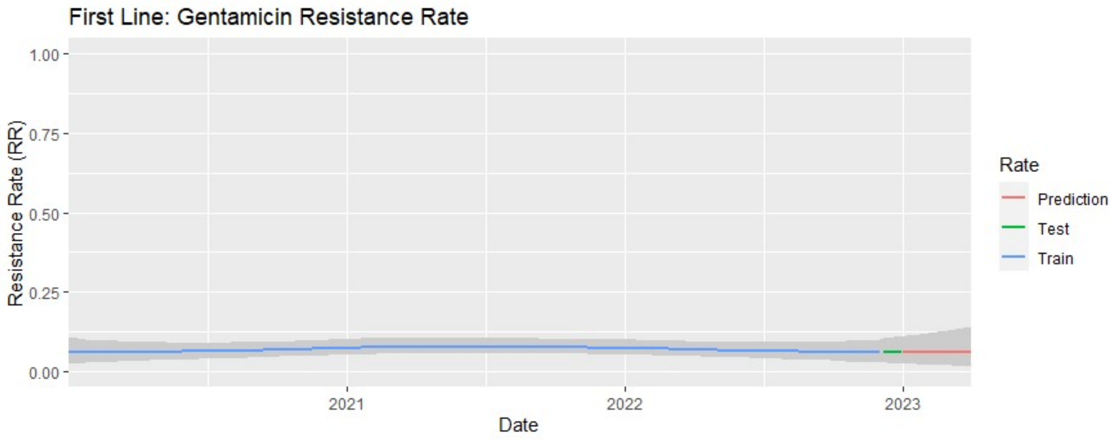


[1] "2020-01-02 Mid 0.059 (95%CI 0.025 to 0.106) Estimated Error 0.0210"

[1] "2022-12-06 Mid 0.059 (95%CI 0.026 to 0.103) Estimated Error 0.0199"

[1] "2022-12-31 Mid 0.059 (95%CI 0.023 to 0.109) Estimated Error 0.0220"

"49.3% posterior probability of decrease "

GI Source

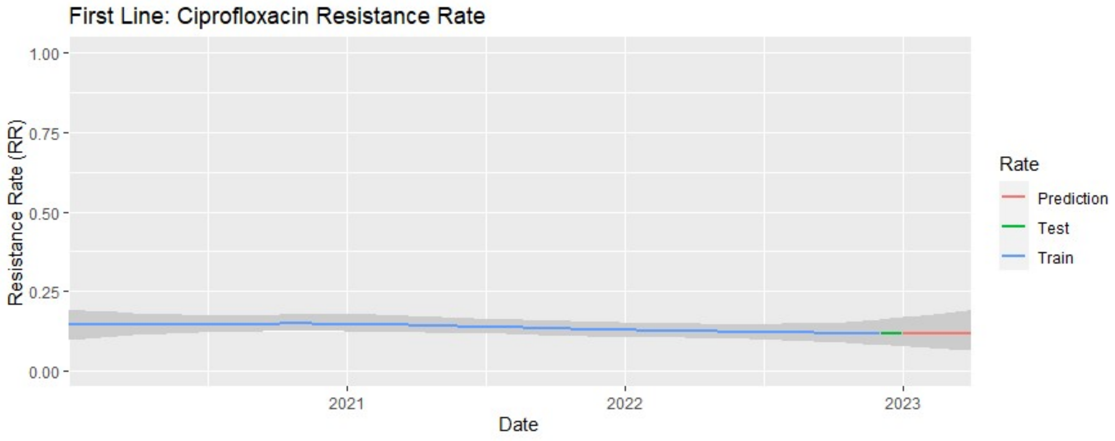


[1] "2020-01-01 Mid 0.145 (95%CI 0.097 to 0.194) Estimated Error 0.0244"

[1] "2022-12-03 Mid 0.116 (95%CI 0.080 to 0.161) Estimated Error 0.0203"

[1] "2022-12-30 Mid 0.116 (95%CI 0.076 to 0.167) Estimated Error 0.0228"

"81.8% posterior probability of decrease "

Urine/renal Source

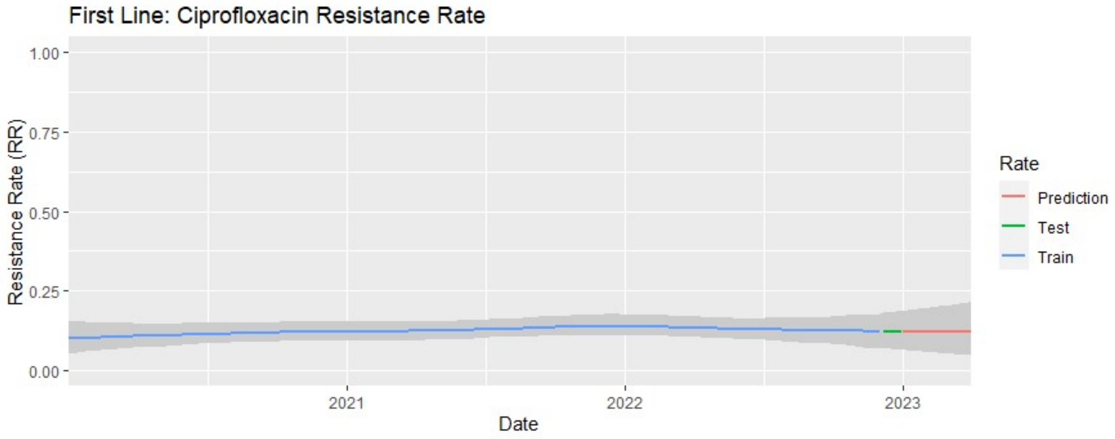


[1] "2020-01-02 Mid 0.101 (95%CI 0.053 to 0.157) Estimated Error 0.0265"

[1] "2022-12-06 Mid 0.122 (95%CI 0.068 to 0.180) Estimated Error 0.0284"

[1] "2022-12-31 Mid 0.122 (95%CI 0.063 to 0.185) Estimated Error 0.0312"

"27.5% posterior probability of decrease "

GI Source

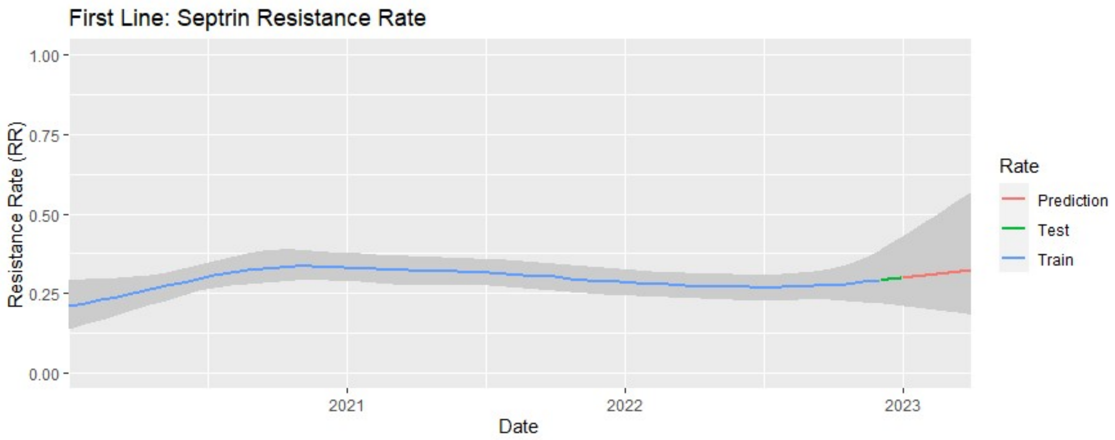


[1] "2020-01-01 Mid 0.210 (95%CI 0.135 to 0.293) Estimated Error 0.0407"

[1] "2022-12-03 Mid 0.291 (95%CI 0.217 to 0.389) Estimated Error 0.0432"

[1] "2022-12-30 Mid 0.298 (95%CI 0.209 to 0.426) Estimated Error 0.0539"

"9.4% posterior probability of decrease "

Urine/renal Source

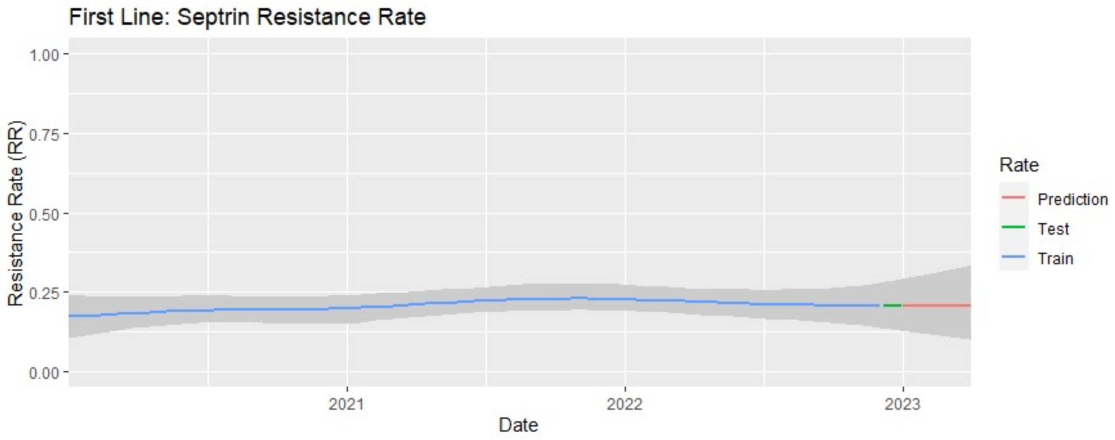


[1] "2020-01-02 Mid 0.172 (95%CI 0.103 to 0.243) Estimated Error 0.0353"

[1] "2022-12-06 Mid 0.206 (95%CI 0.134 to 0.280) Estimated Error 0.0366"

[1] "2022-12-31 Mid 0.205 (95%CI 0.126 to 0.290) Estimated Error 0.0409"

"23.7% posterior probability of decrease "

GI Source

**Antibiotic resistance rate over time assuming pip/taz resistance**

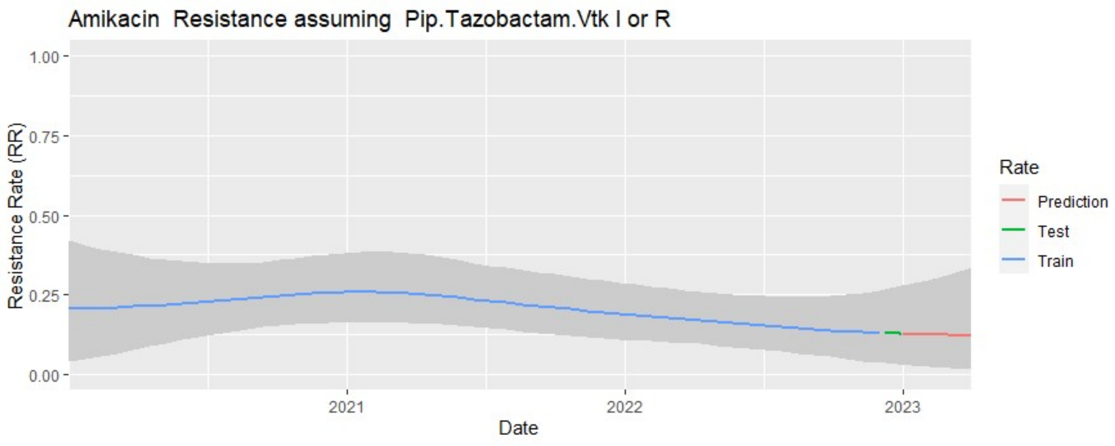


[1] "2020-01-01 Mid 0.205 (95%CI 0.040 to 0.422) Estimated Error 0.1009"

[1] "2022-12-07 Mid 0.128 (95%CI 0.034 to 0.267) Estimated Error 0.0607"

[1] "2022-12-30 Mid 0.126 (95%CI 0.028 to 0.277) Estimated Error 0.0648"

"74.1% posterior probability of decrease "

Urine/renal Source

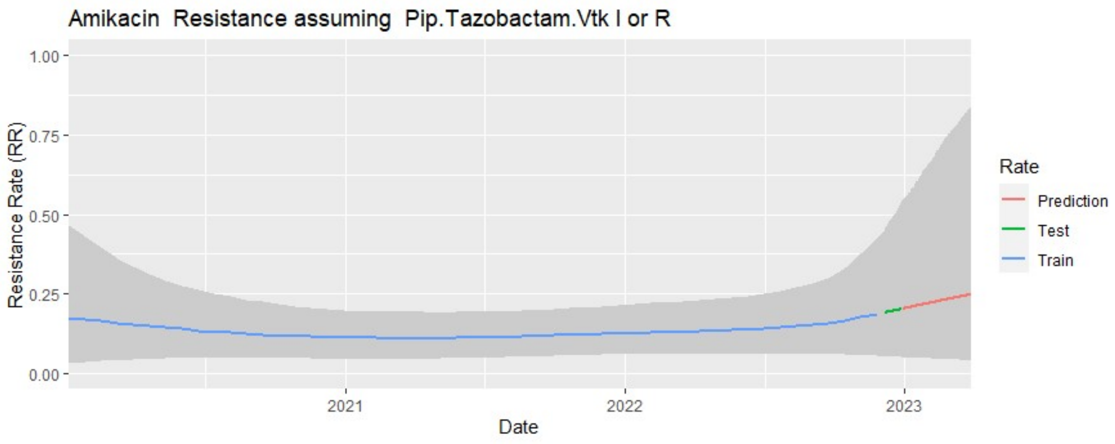


[1] "2020-01-06 Mid 0.175 (95%CI 0.033 to 0.465) Estimated Error 0.1147"

[1] "2022-12-06 Mid 0.191 (95%CI 0.052 to 0.453) Estimated Error 0.1050"

[1] "2022-12-28 Mid 0.202 (95%CI 0.049 to 0.541) Estimated Error 0.1232"

"43.9% posterior probability of decrease "

GI Source

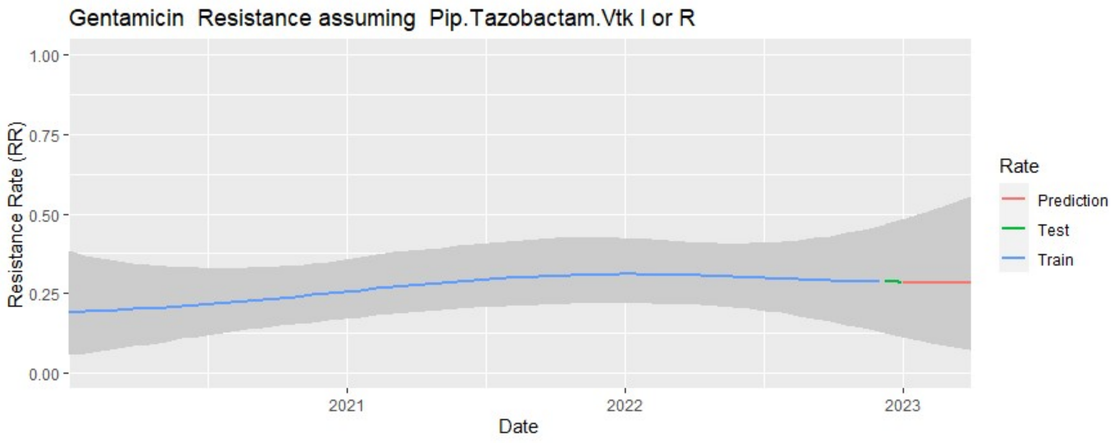


[1] "2020-01-01 Mid 0.191 (95%CI 0.055 to 0.383) Estimated Error 0.0860"

[1] "2022-12-07 Mid 0.286 (95%CI 0.123 to 0.467) Estimated Error 0.0886"

[1] "2022-12-30 Mid 0.285 (95%CI 0.110 to 0.481) Estimated Error 0.0958"

"21.3% posterior probability of decrease "

Urine/renal Source

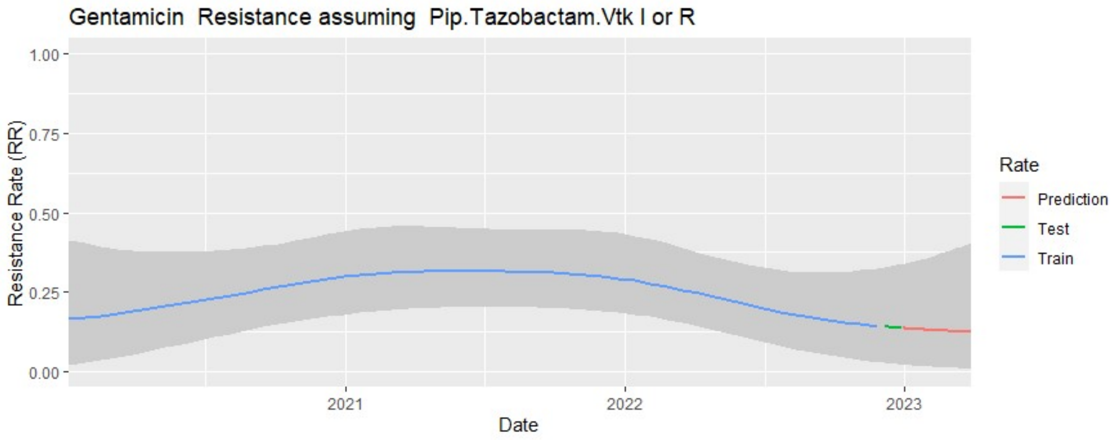


[1] "2020-01-06 Mid 0.164 (95%CI 0.020 to 0.415) Estimated Error 0.1054"

[1] "2022-12-06 Mid 0.140 (95%CI 0.025 to 0.326) Estimated Error 0.0803"

[1] "2022-12-28 Mid 0.135 (95%CI 0.020 to 0.336) Estimated Error 0.0849"

"56.4% posterior probability of decrease "

GI Source

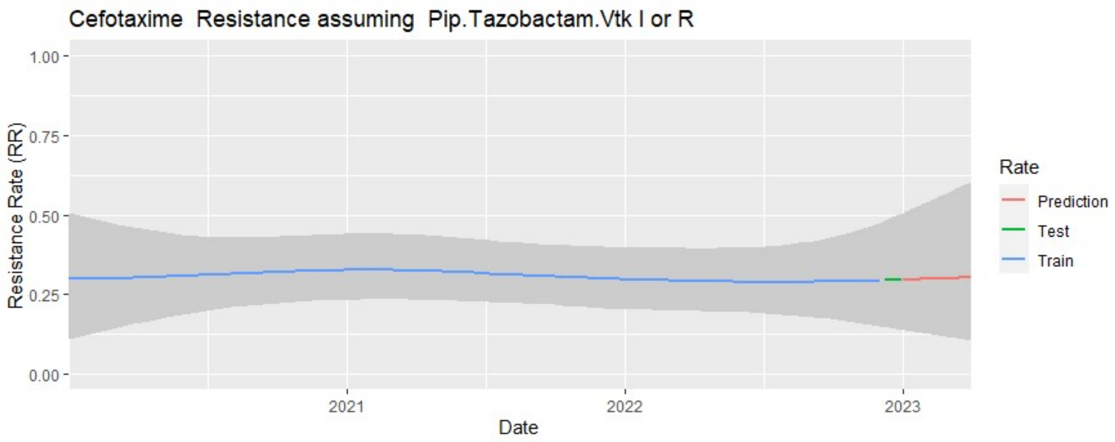


[1] "2020-01-01 Mid 0.298 (95%CI 0.110 to 0.505) Estimated Error 0.1007"

[1] "2022-12-07 Mid 0.294 (95%CI 0.145 to 0.481) Estimated Error 0.0856"

[1] "2022-12-30 Mid 0.295 (95%CI 0.136 to 0.503) Estimated Error 0.0933"

"51.5% posterior probability of decrease "

Urine/renal Source

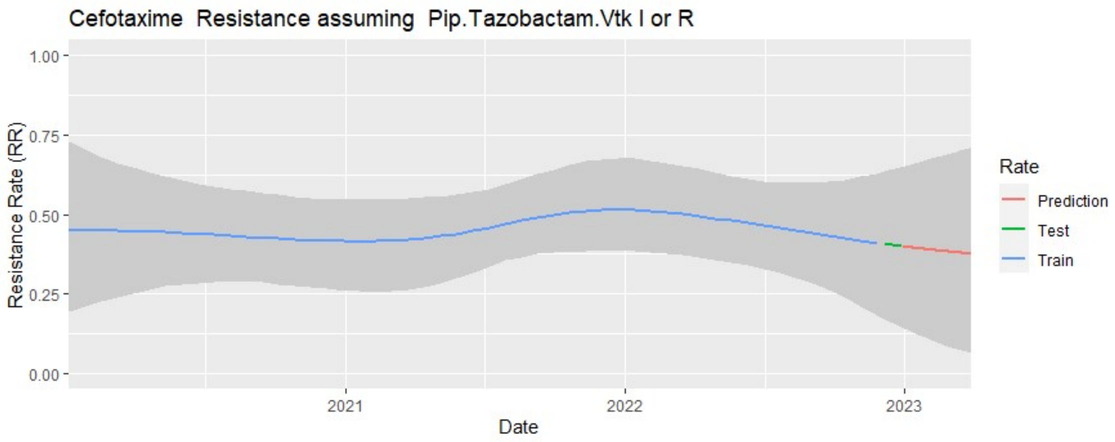


[1] "2020-01-06 Mid 0.450 (95%CI 0.193 to 0.730) Estimated Error 0.1376"

[1] "2022-12-06 Mid 0.406 (95%CI 0.169 to 0.635) Estimated Error 0.1188"

[1] "2022-12-28 Mid 0.399 (95%CI 0.143 to 0.648) Estimated Error 0.1297"

"57.5% posterior probability of decrease "

GI Source

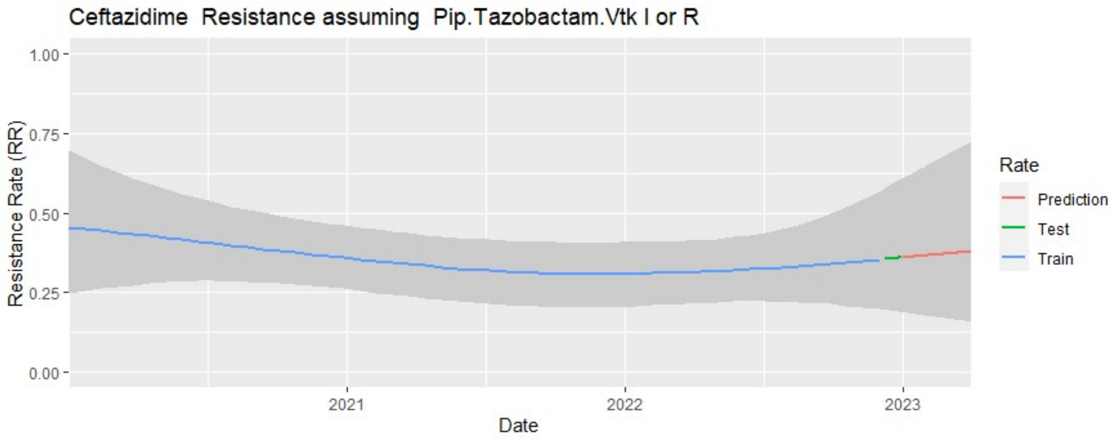


[1] "2020-01-01 Mid 0.454 (95%CI 0.245 to 0.697) Estimated Error 0.1157"

[1] "2022-12-07 Mid 0.354 (95%CI 0.194 to 0.577) Estimated Error 0.0981"

[1] "2022-12-30 Mid 0.359 (95%CI 0.187 to 0.606) Estimated Error 0.1075"

[1] "75.2% posterior probability of decrease "

Urine/renal Source

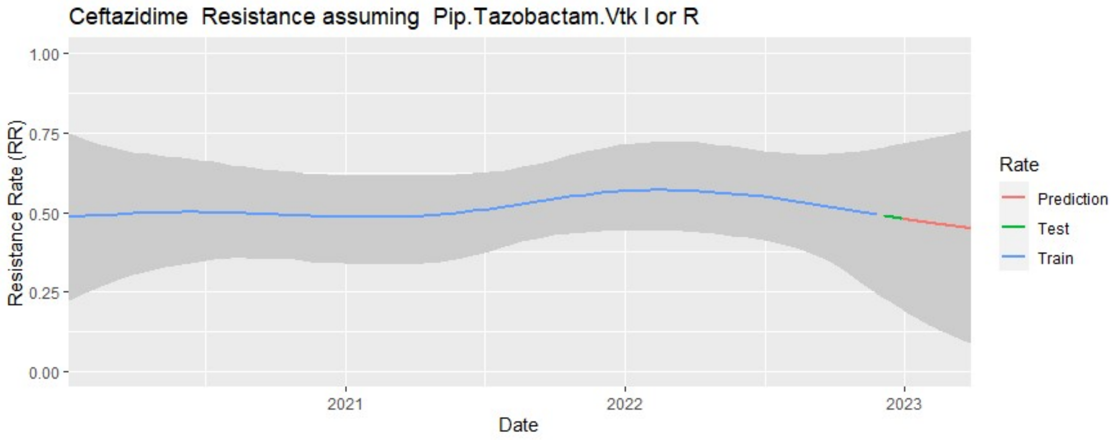


[1] "2020-01-06 Mid 0.486 (95%CI 0.220 to 0.750) Estimated Error 0.1335"

[1] "2022-12-06 Mid 0.488 (95%CI 0.228 to 0.706) Estimated Error 0.1218"

[1] "2022-12-28 Mid 0.479 (95%CI 0.195 to 0.715) Estimated Error 0.1338"

"48.4% posterior probability of decrease "

GI Source

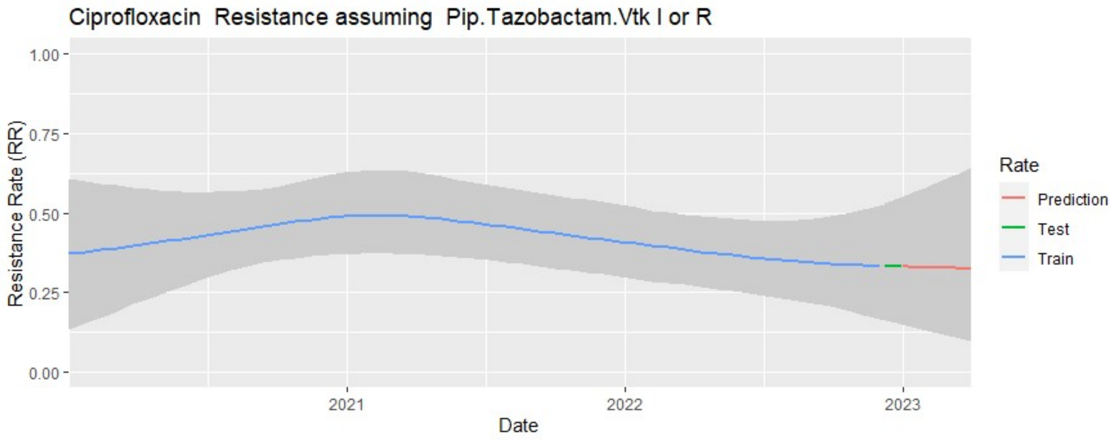


[1] "2020-01-01 Mid 0.370 (95%CI 0.133 to 0.608) Estimated Error 0.1245"

[1] "2022-12-07 Mid 0.332 (95%CI 0.160 to 0.528) Estimated Error 0.0945"

[1] "2022-12-30 Mid 0.330 (95%CI 0.147 to 0.548) Estimated Error 0.1029"

"59.4% posterior probability of decrease "

Urine/renal Source

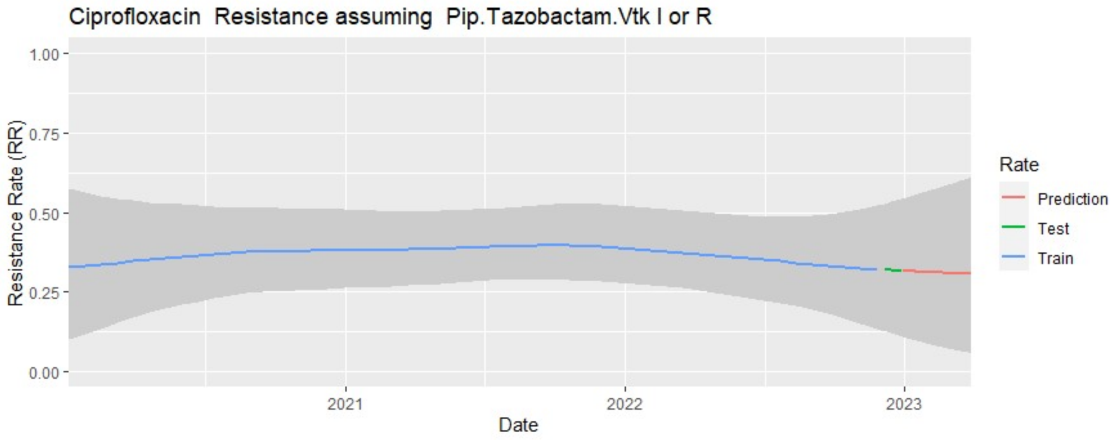


[1] "2020-01-06 Mid 0.326 (95%CI 0.099 to 0.574) Estimated Error 0.1231"

[1] "2022-12-06 Mid 0.319 (95%CI 0.125 to 0.527) Estimated Error 0.1033"

[1] "2022-12-28 Mid 0.316 (95%CI 0.108 to 0.542) Estimated Error 0.1107"

"51.0% posterior probability of decrease "

GI Source

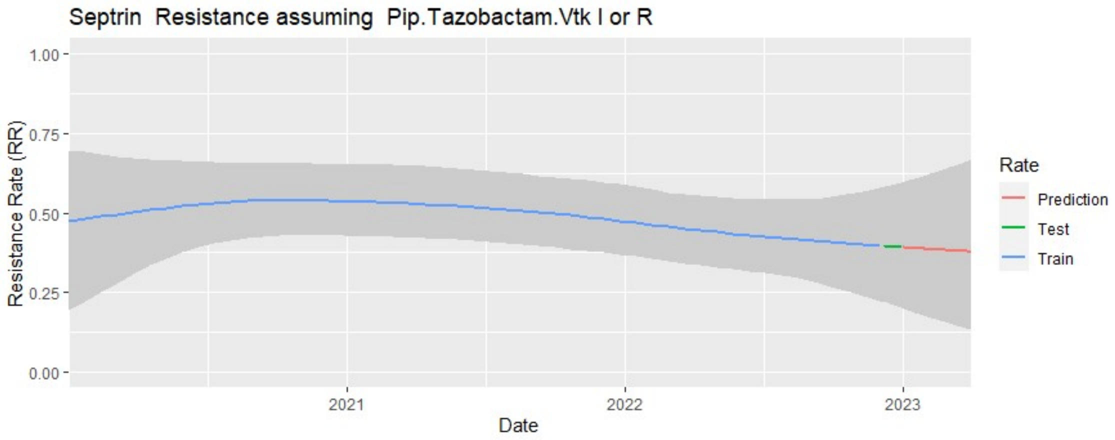


[1] "2020-01-01 Mid 0.475 (95%CI 0.193 to 0.699) Estimated Error 0.1309"

[1] "2022-12-07 Mid 0.395 (95%CI 0.219 to 0.581) Estimated Error 0.0929"

[1] "2022-12-30 Mid 0.391 (95%CI 0.199 to 0.594) Estimated Error 0.1009"

"70.2% posterior probability of decrease "

Urine/renal Source

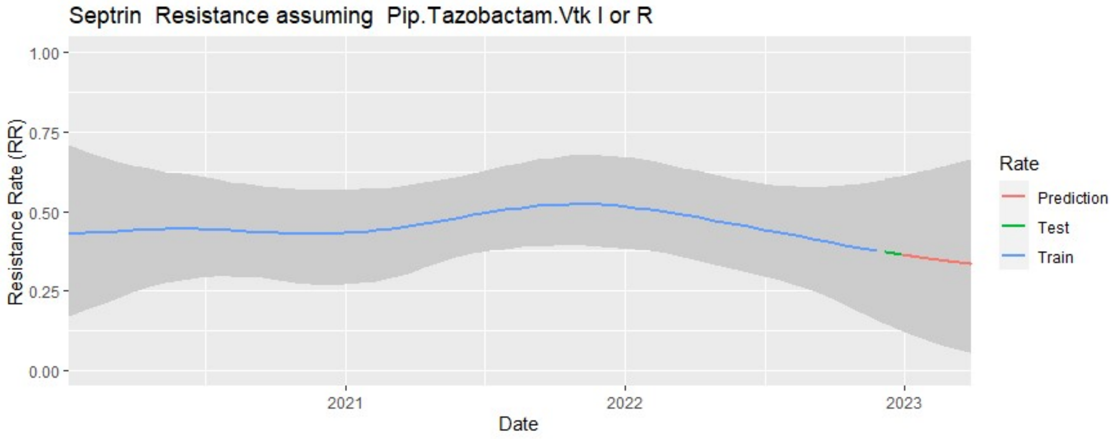


[1] "2020-01-06 Mid 0.428 (95%CI 0.169 to 0.707) Estimated Error 0.1382"

[1] "2022-12-06 Mid 0.371 (95%CI 0.145 to 0.602) Estimated Error 0.1183"

[1] "2022-12-28 Mid 0.362 (95%CI 0.122 to 0.611) Estimated Error 0.1280"

"61.5% posterior probability of decrease "

GI Source

**Antibiotic resistance rate over time assuming ceftazidime resistance**

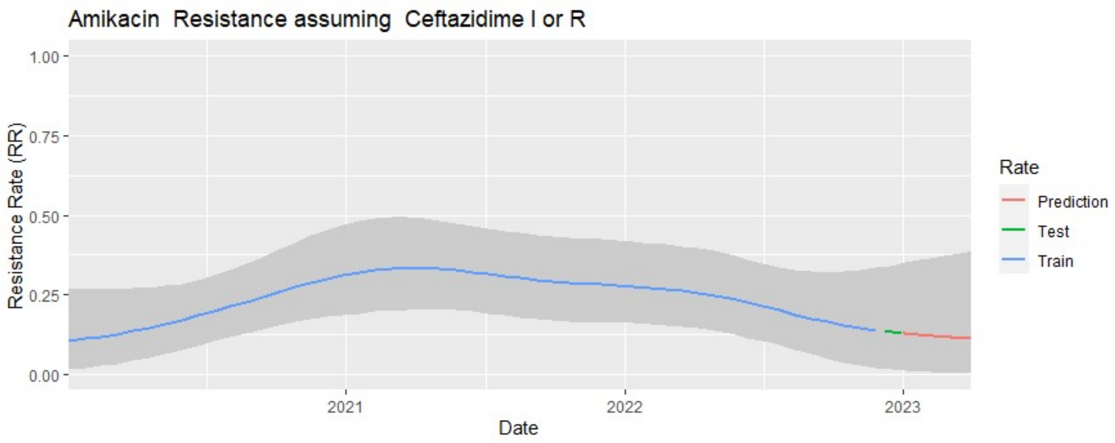


[1] "2020-01-03 Mid 0.105 (95%CI 0.014 to 0.271) Estimated Error 0.0679"

[1] "2022-12-07 Mid 0.134 (95%CI 0.015 to 0.336) Estimated Error 0.0849"

[1] "2022-12-30 Mid 0.127 (95%CI 0.010 to 0.348) Estimated Error 0.0893"

**"38.5% posterior probability of decrease "**

Urine/renal Source

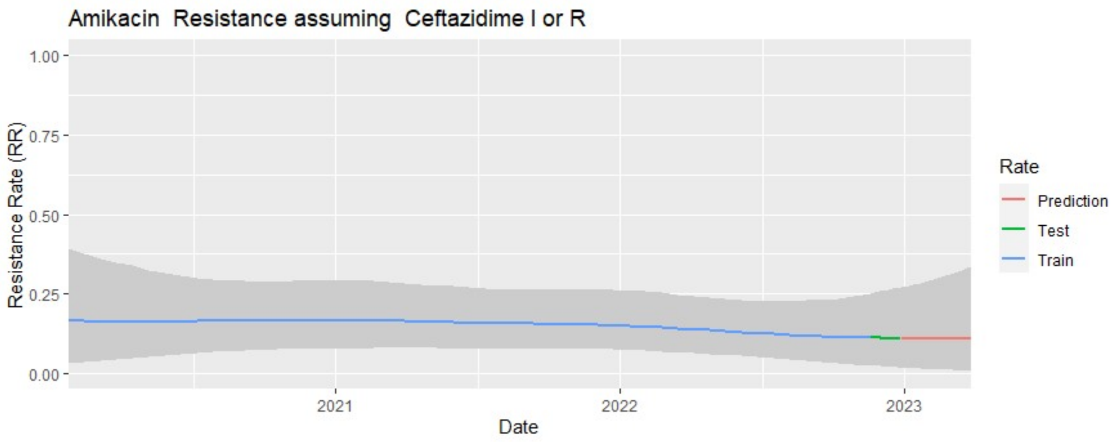


[1] "2020-01-24 Mid 0.164 (95%CI 0.029 to 0.394) Estimated Error 0.0957"

[1] "2022-11-17 Mid 0.111 (95%CI 0.023 to 0.250) Estimated Error 0.0594"

[1] "2022-12-26 Mid 0.109 (95%CI 0.016 to 0.268) Estimated Error 0.0666"

"66.6% posterior probability of decrease "

GI Source

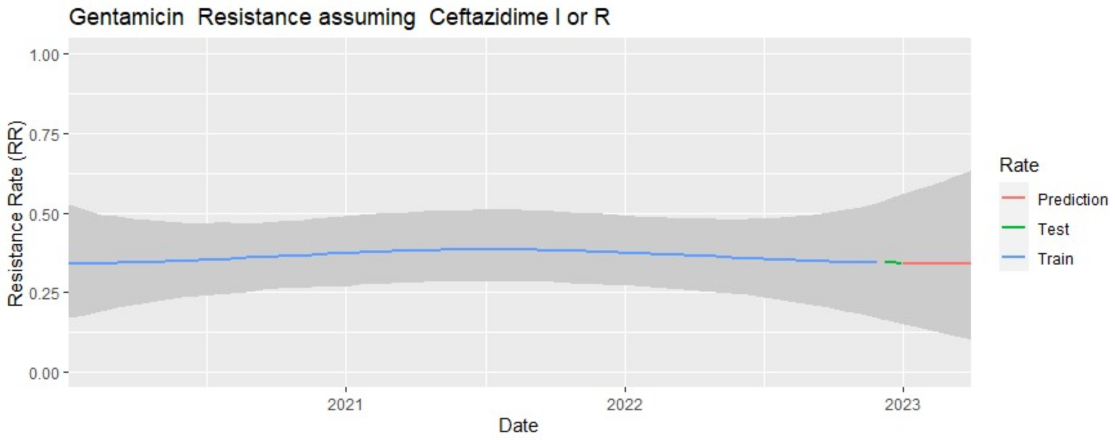


[1] "2020-01-03 Mid 0.339 (95%CI 0.168 to 0.526) Estimated Error 0.0917"

[1] "2022-12-07 Mid 0.342 (95%CI 0.163 to 0.539) Estimated Error 0.0965"

[1] "2022-12-30 Mid 0.341 (95%CI 0.149 to 0.557) Estimated Error 0.1037"

"48.5% posterior probability of decrease "

Urine/renal Source

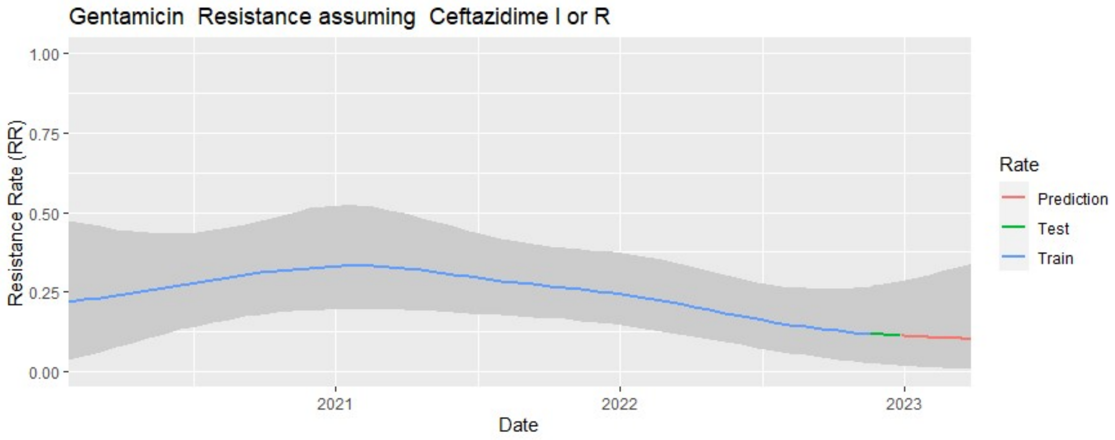


[1] "2020-01-24 Mid 0.218 (95%CI 0.036 to 0.476) Estimated Error 0.1182"

[1] "2022-11-17 Mid 0.118 (95%CI 0.024 to 0.268) Estimated Error 0.0647"

[1] "2022-12-26 Mid 0.111 (95%CI 0.016 to 0.282) Estimated Error 0.0716"

"78.1% posterior probability of decrease "

GI Source

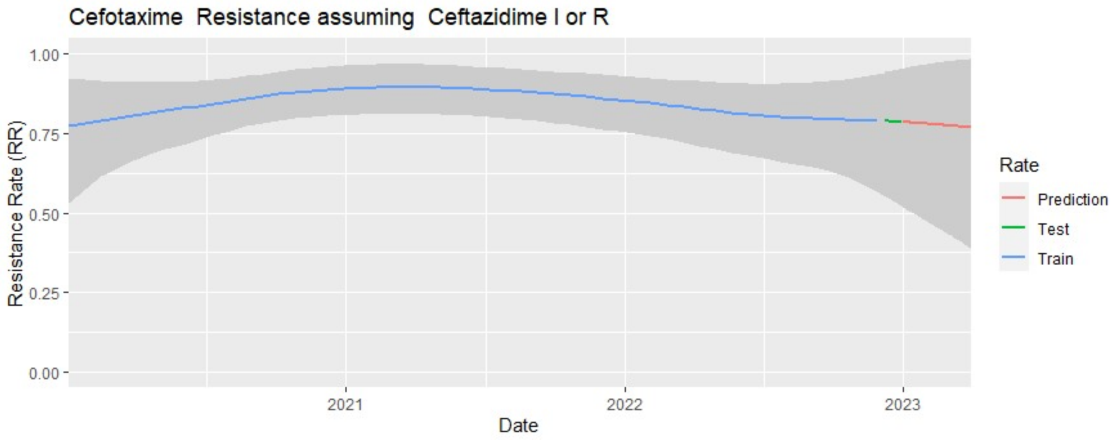


[1] "2020-01-03 Mid 0.774 (95%CI 0.531 to 0.925) Estimated Error 0.1028"

[1] "2022-12-07 Mid 0.789 (95%CI 0.552 to 0.942) Estimated Error 0.1009"

[1] "2022-12-30 Mid 0.786 (95%CI 0.520 to 0.953) Estimated Error 0.1120"

"46.4% posterior probability of decrease "

Urine/renal Source

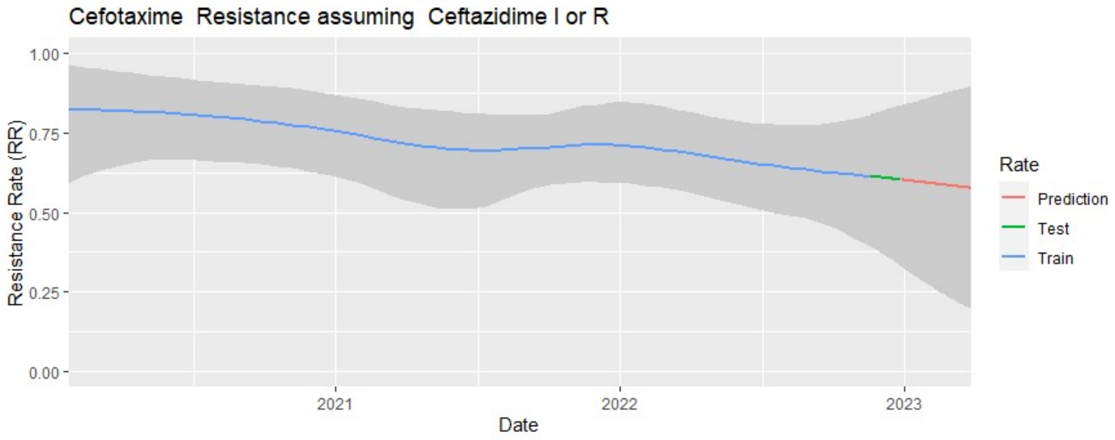


[1] "2020-01-24 Mid 0.824 (95%CI 0.592 to 0.965) Estimated Error 0.0958"

[1] "2022-11-17 Mid 0.614 (95%CI 0.393 to 0.808) Estimated Error 0.1047"

[1] "2022-12-26 Mid 0.603 (95%CI 0.332 to 0.836) Estimated Error 0.1261"

"92.7% posterior probability of decrease "

GI Source

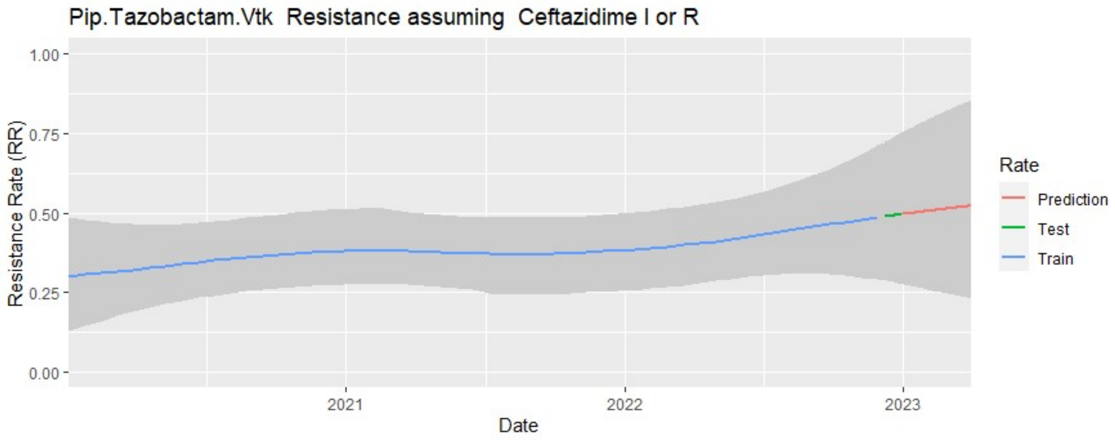


[1] "2020-01-03 Mid 0.301 (95%CI 0.128 to 0.488) Estimated Error 0.0911"

[1] "2022-12-07 Mid 0.488 (95%CI 0.286 to 0.722) Estimated Error 0.1116"

[1] "2022-12-30 Mid 0.495 (95%CI 0.275 to 0.753) Estimated Error 0.1213"

"10.3% posterior probability of decrease "

Urine/renal Source

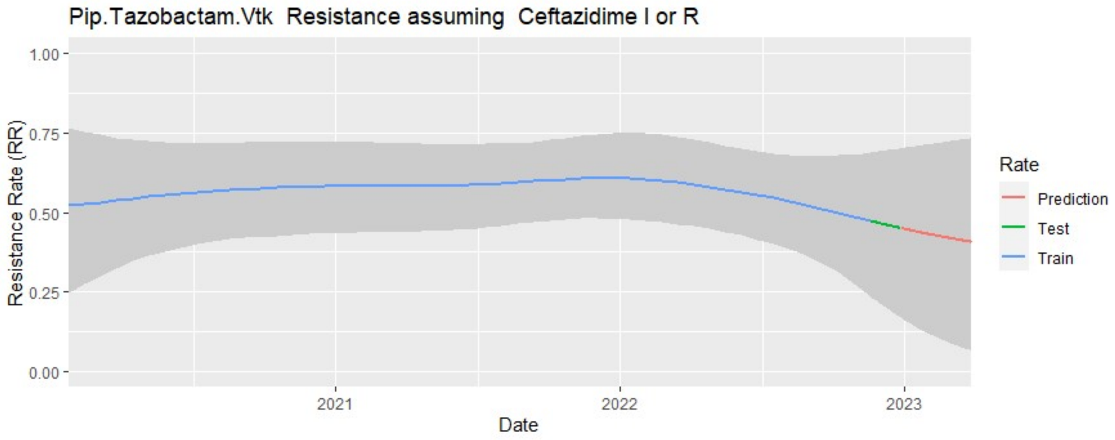


[1] "2020-01-24 Mid 0.521 (95%CI 0.245 to 0.764) Estimated Error 0.1344"

[1] "2022-11-17 Mid 0.472 (95%CI 0.236 to 0.688) Estimated Error 0.1166"

[1] "2022-12-26 Mid 0.450 (95%CI 0.169 to 0.700) Estimated Error 0.1379"

"60.9% posterior probability of decrease "

GI Source

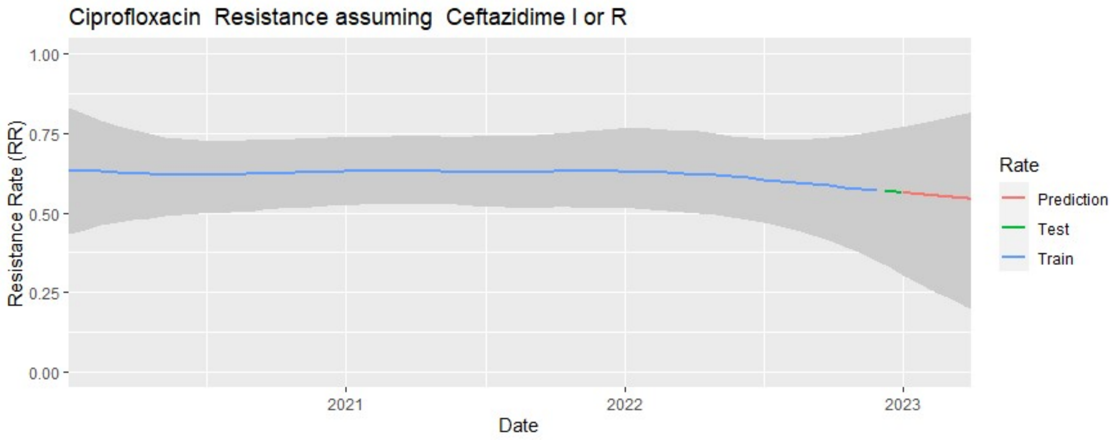


[1] "2020-01-03 Mid 0.635 (95%CI 0.433 to 0.830) Estimated Error 0.0991"

[1] "2022-12-07 Mid 0.569 (95%CI 0.336 to 0.761) Estimated Error 0.1083"

[1] "2022-12-30 Mid 0.564 (95%CI 0.305 to 0.769) Estimated Error 0.1181"

"65.9% posterior probability of decrease "

Urine/renal Source

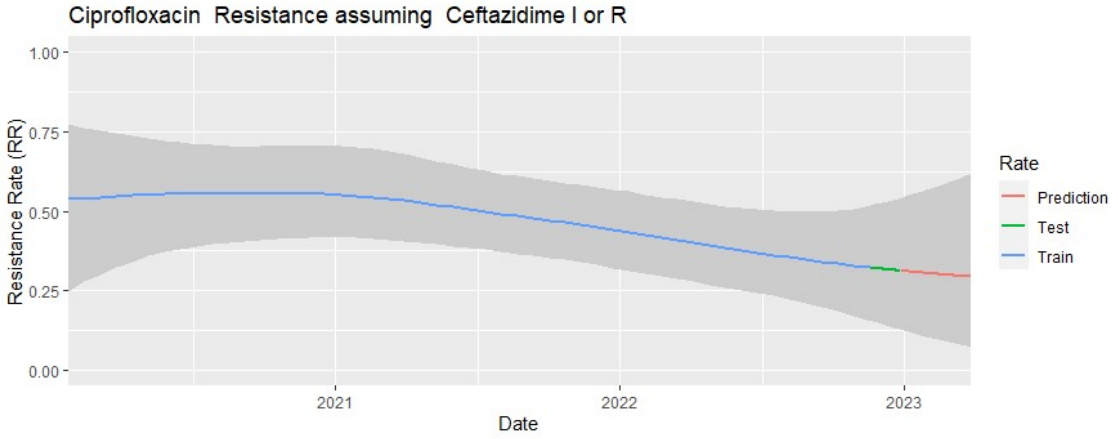


[1] "2020-01-24 Mid 0.537 (95%CI 0.247 to 0.774) Estimated Error 0.1355"

[1] "2022-11-17 Mid 0.322 (95%CI 0.154 to 0.518) Estimated Error 0.0925"

[1] "2022-12-26 Mid 0.312 (95%CI 0.126 to 0.539) Estimated Error 0.1054"

"90.3% posterior probability of decrease "

GI Source

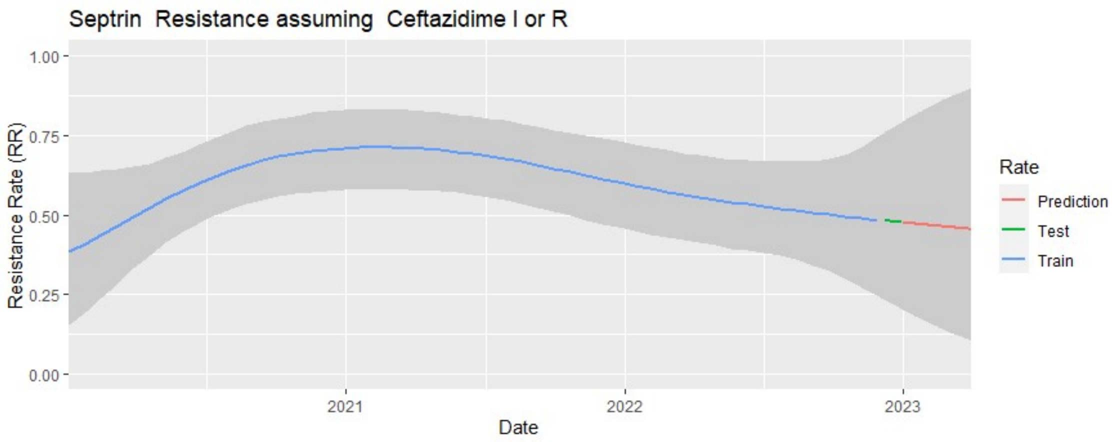


[1] "2020-01-03 Mid 0.383 (95%CI 0.152 to 0.633) Estimated Error 0.1267"

[1] "2022-12-07 Mid 0.482 (95%CI 0.232 to 0.759) Estimated Error 0.1332"

[1] "2022-12-30 Mid 0.476 (95%CI 0.202 to 0.791) Estimated Error 0.1488"

"31.1% posterior probability of decrease "

Urine/renal Source

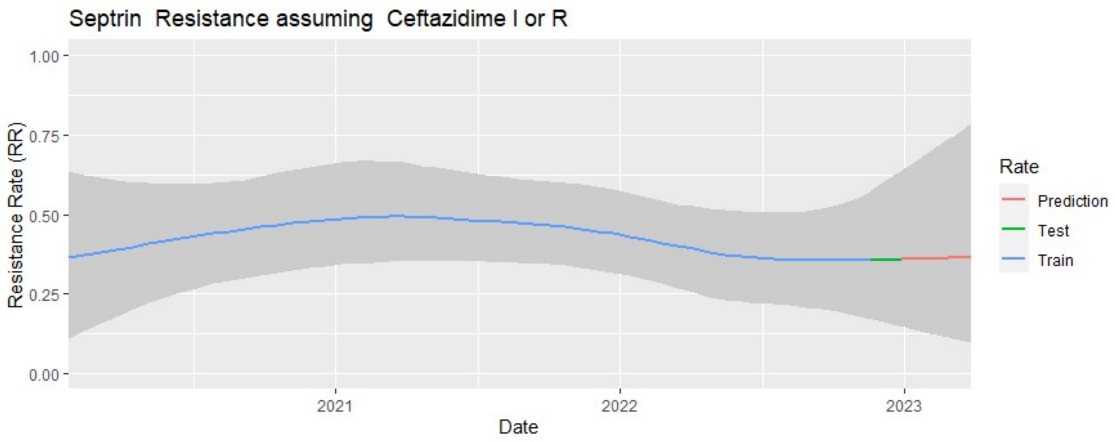


[1] "2020-01-24 Mid 0.365 (95%CI 0.109 to 0.635) Estimated Error 0.1392"

[1] "2022-11-17 Mid 0.356 (95%CI 0.169 to 0.573) Estimated Error 0.1039"

[1] "2022-12-26 Mid 0.358 (95%CI 0.146 to 0.633) Estimated Error 0.1243"

"52.6% posterior probability of decrease "

GI Source
